# An Online Application to Explain Community Immunity with Personalized Avatars: A Randomized Controlled Trial

**DOI:** 10.1101/2024.10.18.24314709

**Authors:** Hina Hakim, Julie A Bettinger, Christine T. Chambers, S. Michelle Driedger, Eve Dubé, Teresa Gavaruzzi, Anik Giguere, Noah M. Ivers, Anne-Sophie Julien, Shannon E. MacDonald, Magniol Noubi, Rita Orji, Elizabeth Parent, Beate Sander, Aaron M. Scherer, Kumanan Wilson, Daniel Reinharz, Holly O. Witteman

## Abstract

**Background:** To evaluate the effects of a web-based, personalized avatar intervention conveying the concept of community immunity (herd immunity) on risk perception (perceptions of the risk of infection spreading (to self, family, community, and vulnerable individuals)) and other cognitive and emotional responses across 4 vaccine-preventable disease contexts: measles, pertussis, influenza, and an unnamed “vaccine-preventable disease.”

**Methods:** Through a robust user-centered design process, we developed a web application, “*herdimm*,” showing how community immunity works. In our application, people personalize a virtual community by creating avatars (themselves, 2 vulnerable people in their community, and 6 other people around them; e.g., family members or co-workers.) *Herdimm* integrates these avatars in a 2-minute narrated animation showing visually how infections spread with and without the protection of community immunity. The present study was a 2×4 factorial randomized controlled trial to assess *herdimm*’s effects. We recruited 3883 adults via Qualtrics living in Canada who could complete an online study in English or French. We pre-registered our study, including depositing our questionnaire and pre-scripted statistical code on Open Science Framework (https://osf.io/hkysb/). The trial ran from March 1 to July 1, 2021. We compared the web application to no intervention (i.e. control) on primary outcome risk perception, divided into *objective risk perception* (accuracy of risk perception) and *subjective risk perception* (subjective sense of risk), and on secondary outcomes–emotions (worry, anticipated guilt), knowledge, and vaccination intentions–using analysis of variance for continuous outcomes and logistic regression for dichotomous outcomes. We conducted planned moderation analyses using participants’ scores on a validated scale of individualism and collectivism as moderators.

**Results:** Overall, *herdimm* had desirable effects on all outcomes. People randomized to *herdimm* were more likely to score high on objective risk perception (58.0%, 95% confidence interval 56.0%-59.9%) compared to those assigned to the control condition (38.2%, 95% confidence interval 35.5%-40.9%). *Herdimm* increased subjective risk perception from a mean of 5.30 on a scale from 1 to 7 among those assigned to the control to 5.54 among those assigned to *herdimm*. The application also increased emotions (worry, anticipated guilt) (F(1,3875)=13.13, p<0.001), knowledge (F(1,3875)=36.37, p<0.001) and vaccination intentions (Chi-squared(1)=9.4136, p=0.002). While objective risk perception did not differ by disease (Chi-squared(3)=6.94, p=0.074), other outcomes did (subjective risk perception F(3,3875) = 5.6430, p<0.001; emotions F(3,3875)=78.54, p<0.001; knowledge (F(3,3875)=5.20, p=0.001); vaccination intentions Chi-squared (3)=15.02, p=0.002). Moderation models showed that many findings were moderated by participants’ individualism and collectivism scores. Overall, whereas outcomes tended not to vary by individualism and collectivism among participants in the control condition, the positive effects of *herdimm* were larger among participants with more collectivist orientations and effects were sometimes negative among participants with more individualist orientations.

**Conclusions:** Conveying the concept of community immunity through a web application using personalized avatars increases objective and subjective risk perception and positively influences intentions to receive vaccines, particularly among people who have more collectivist worldviews. Including prosocial messages about the collective benefits of vaccination in public health campaigns may increase positive effects among people who are more collectivist while possibly backfiring among those who are more individualistic.

## Introduction

Many vaccines protect against disease by not only preventing infection in those receiving the vaccine but also preventing the infection from being transmitted from one person to another [1,2]. Such transmission prevention can occur because the vaccines confer sterilizing immunity, because they lower viral load and therefore partially prevent transmission, and/or because, by lowering infection risk, they decrease the number of people within a population who are infected and thus lower the chances of interacting with an infectious individual [3].

The concept of community immunity, also known as herd immunity, refers to the indirect protection of unvaccinated or undervaccinated people by increasing the number of people around them who have immunity. Such an increase in immunity can break the chain of transmission by decreasing the overall probability of contact with an infectious agent, thereby preventing the spread of infectious agents within susceptible populations [1].

Despite the success of vaccines and immunization programs as public health measures, some people perceive such measures as unsafe or even unnecessary [4,5]. The reasons behind such perceptions include concerns about vaccine safety, a mistrust in healthcare agencies or pharmaceutical companies, unfamiliarity with vaccine-preventable diseases, and inaccurate mental models that ignore community immunity [6–9]. Providing accurate education about community immunity may be useful for increasing willingness to vaccinate, generating benefits both to individuals and communities, although the success of these efforts may depend on the extent the population of interest is more collectivist or more individualist [10,11].

The COVID-19 pandemic provided a salient example of the challenge of ensuring public understanding of individual and collective benefits of vaccines. On top of already-existing public health communication challenges regarding established vaccines, there was uncertainty about community immunity thresholds due to fluctuating estimates of the disease’s reproductive number [12], new variants reducing vaccine effectiveness [13], the lack of sterilizing immunity offered by available vaccines, and evolving hybrid immunity due to widespread infection [14]. The Covid-19 pandemic therefore highlighted both the importance and the difficulty of communication strategies emphasizing individuals’ roles in reducing transmission and protecting vulnerable people within communities [13].

In a previous systematic review, we identified graphic visualization as a potentially promising approach for achieving effective communication about community immunity [15]. Visualization is a powerful communication mechanism that uses pre-attentive processing to communicate large amounts of information rapidly in understandable and compelling ways [16]. While some visualizations about community immunity existed, few had been evaluated for their effects on vaccination intentions and uptake nor on vaccine uptake’s precursors, namely knowledge, attitudes, beliefs and emotions. Furthermore, none had been evaluated in multiple disease contexts [15]. Because disease transmission dynamics and community immunity parameters (for example, vaccine effectiveness) differ across vaccine-preventable diseases, ideally, an effective means of conveying the concept of community immunity should be usable across multiple diseases. Finally, no existing visualizations used personalized avatars. Studies of the Proteus Effect have shown that people identify with a personalized avatar to the point that they change their behaviour according to their avatar’s characteristics [17–19], suggesting that personalized avatars in a community immunity visualization may reinforce people’s sense of actually being part of the community. This sense of community would ideally make the visualization more salient and personally relevant, thus influencing risk perception, a key antecedent of vaccine uptake. [20,21]

Risk perception refers to the perceived severity and likelihood of an adverse event. Risk perception may draw on an individual’s comprehension of the objective likelihood of events, and also on an individual’s subjective feelings about the severity and likelihood of events. This latter subjective dimension of risk perception is referred to as the ‘risk as feelings’ framework which aligns with observations of people’s reactions to danger as instinctive and intuitive [23,24]. When people receive information about health risks, they may translate it intuitively into feelings of anxiety, worry, or distress, or a more vague feeling of ‘badness’ associated with the risk [25,26]. Risk perception influences vaccine decisions and behavior by shaping how individuals perceive and weigh the risks of vaccination against the risks of not getting vaccinated [27,28]. Therefore, when seeking to understand the impact of an educational intervention about community immunity, its effects on risk perception are of particular interest.

### Objective

Our study objective was to evaluate a previously-developed web application, hereafter referred to as *herdimm*. The main purpose of *herdimm* was to clearly convey the concept of community immunity to people from various backgrounds, including varying levels of education. We developed it with attention to its potential effects on people’s feelings relevant to their role in community immunity [29]. In the web application, people personalize a virtual community by creating avatars representing themselves, 2 vulnerable people in their community, and 6 other people around them; for example, family members or co-workers. The web application integrates these avatars in a 2-minute narrated animation explaining how community immunity works [30,31].

In this study, we aimed to evaluate the effects of *herdimm* on risk perception (that is, perceptions of the risk of spreading infection to oneself, one’s family, one’s community, and vulnerable individuals within one’s community), and on known antecedents of decisions to get vaccinated across four vaccine-preventable disease contexts. Our research questions were therefore:

1. Does *herdimm* influence risk perception (meaning vaccine-preventable disease risk to oneself, family, community and vulnerable people) compared to a control (no intervention)?
2. Does *herdimm* influence other outcomes (vaccination intentions, emotion, knowledge, and trust in information) compared to a control (no intervention)?

## Methods

### Study design

We conducted a 2 (visualization: *herdimm*, no education about community immunity) x 4 (disease: measles, pertussis, flu, unnamed “vaccine-preventable disease” hereby referred to as “generic”) factorial randomized controlled trial among adults in Canada using Qualtrics, an online survey platform. The study arms differed by the two independent variables (visualization and disease). In other words, participants in different arms saw either no intervention (control) or the herdimm intervention (*herdimm*) for one of four possible vaccine-preventable diseases. For efficient data collection, the present 2×4 trial (8 study arms) conducted in both English and French was complemented by a head-to-head comparison of existing available interventions about community immunity, most of which were available only in English and none of which offered different versions for different diseases. This complementary study added 5 arms to the larger study and will be reported separately.

### Interventions

The main intervention was the application named *herdimm*, developed in our previous study [29]; web development code available on GitHub (https://github.com/Witteman-Lab/herdimm). A version of *herdimm* (measles/generic, English) is shown here on YouTube for illustration: [30]. Links to sample visualizations of all possible *herdimm* versions are available in Appendix 1.

In the current study, we assessed the effects of *herdimm* across four possible diseases (measles, pertussis, flu/influenza, and generic). We tested the intervention across multiple diseases because disease transmission varies (including the number of new infections caused by the first infected person in a population) and community immunity parameters differ across vaccine-preventable diseases. Among other differences, for some diseases such as measles, a high vaccination rate can achieve a sufficient level of community immunity. For other diseases such as influenza, vaccines must be complemented by other protective measures to optimize protection of all members of a community. Therefore, it was important to assess whether any potential effects would be robust and would generalize across infectious diseases with different transmission characteristics. For the “generic” condition, we used the visualization for measles, but did not specify the disease and merely referred to, “a vaccine-preventable disease,” in all questions. For example, participants assigned to the measles condition were asked to indicate their level of agreement or disagreement with the statement, “I am worried about getting measles,” while the equivalent item for those assigned to the generic disease was, “I am worried about getting a vaccine preventable disease.” We developed *herdimm* in both official languages of Canada (English and French), and the trial was conducted in both languages.

### Randomization and allocation

We used computerized randomization within Qualtrics [32] to automatically assign participants to study arms randomly. Each study arm was accessible in both languages and we pre-specified our sampling parameters such that 75% of the sample for each study arm would be people who elected to participate in English and 25% in French, based on the percentage of spoken languages within the Canadian population [33]. This was a single-blinded study because we could not mask participants to the fact that they have been randomized to a computer application including a visualization. However, participants did not know the purpose of the study arm to which they were assigned, and investigators were blinded to the study arm during the data collection and in the pre-scripted statistical code.

### Ethics and trial registration

This study was approved by the ethics committee of Laval University. We pre-registered the trial at ClinicalTrials.gov (NCT04787913). We pre-planned all statistical analyses by developing our statistical code prior to beginning the trial using simulated data with the support of a statistician (ASJ). This pre-scripted statistical code was also peer-reviewed by an external statistician. We pre-registered all study materials (protocol, study questionnaire, data dictionary, and pre-scripted statistical code) on Open Science Framework on February 26, 2021 (https://osf.io/hkysb/). We report the trial following the CONSORT guidelines [34]. The trial began on March 1, 2021 and ended on July 1, 2021.

### Study participants

We conducted the online randomized controlled trial in Qualtrics among adults living in Canada. Participants were recruited using established survey panels subcontracted in Qualtrics. Participants were eligible if they were 18 years and above, residing in Canada, able to use a computer, could read and understand at least one of French or English, and were able to provide consent. Those who did not meet this criteria were excluded from the study. We used sampling quotas in an effort to represent key characteristics of the Canadian population in the most recent national census [35]. Specifically, we required that the overall sample population for the study consisted of 50% people who identified as female, 49% as male, and 1% as other (e.g., non-binary, two spirit). Regarding age, 30.5% of the study population were required to be within the age group 18-34 years old, 34.4% within 35-49 years old, and 35.2% within the group aged 50 years and older. For education level, we specified that 25-35% of the study population must report their highest education level as elementary school (completed or not) or high school diploma, 10-15% as apprenticeship or trade certificate or diploma, 20-25% as college or polytechnical school certificate or diploma, and 30-40% as university degree or above. Finally, we required a minimum of 22% of the study population to report their racial identity as other than white, and 25% of the study population to be French speakers. A small incentive (typically $1-1.50, depending on panel) was offered to the participants to complete the survey.

### Outcomes

Our primary outcome was risk perception. We chose this as our primary outcome as people’s decisions regarding vaccination depend on their perception of risks associated with diseases and vaccines, including vaccine safety and potential side effects. Such risk perceptions may be formed long before they are presented with a decision to vaccinate or not, suggesting that interventions that shape such perceptions may be important upstream influences on subsequent vaccine acceptance. High risk perception of vaccines can deter vaccination, resulting in lower vaccination rates, but building trust in vaccines, science and effective communication can alleviate risk perception and increase vaccination intentions [36]. We defined risk perception for this study as the participants’ sense of risk posed by a vaccine-preventable disease to an individual, their family, their community, and vulnerable people in their community. Secondary outcomes were measures of emotions (worry, anticipated guilt), knowledge about community immunity, and vaccination intentions.

### Measures

We measured our primary outcome using questions developed and pre-tested to assess risk perception (6 items, Figure 1 and Table 1). Based on this pre-testing, we were concerned that the first item might be measuring a different construct than the other five items. Our pre-scripted analyses (written prior to starting data collection, with this potential difference in mind) suggested this was indeed the case. Specifically, the scatter plot and Bland-Altman test (see Figures S1 S2 in Appendix 2) showed that the first item did not measure the same construct as the second through sixth items. We therefore divided the six items into *objective risk perception* (Figure 1; item 1, which assessed how people perceived risk compared to actual risk) and *subjective risk perception* (Table1; items 2 to 6, which assessed people’s subjective feelings of risk.) The Cronbach alpha for subjective risk perception was 0.76, which is acceptable for combining 5 items into a single measure.

**Figure 1.**
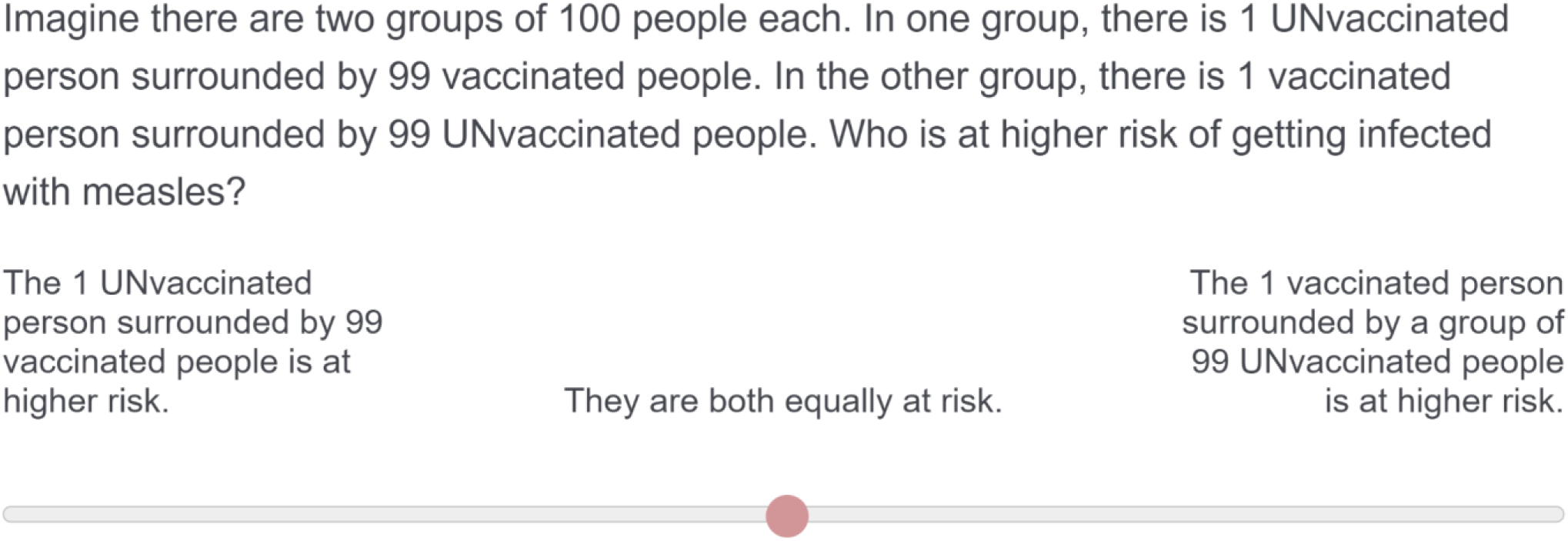
Objective risk perception (condition: measles)

**Table 1.**
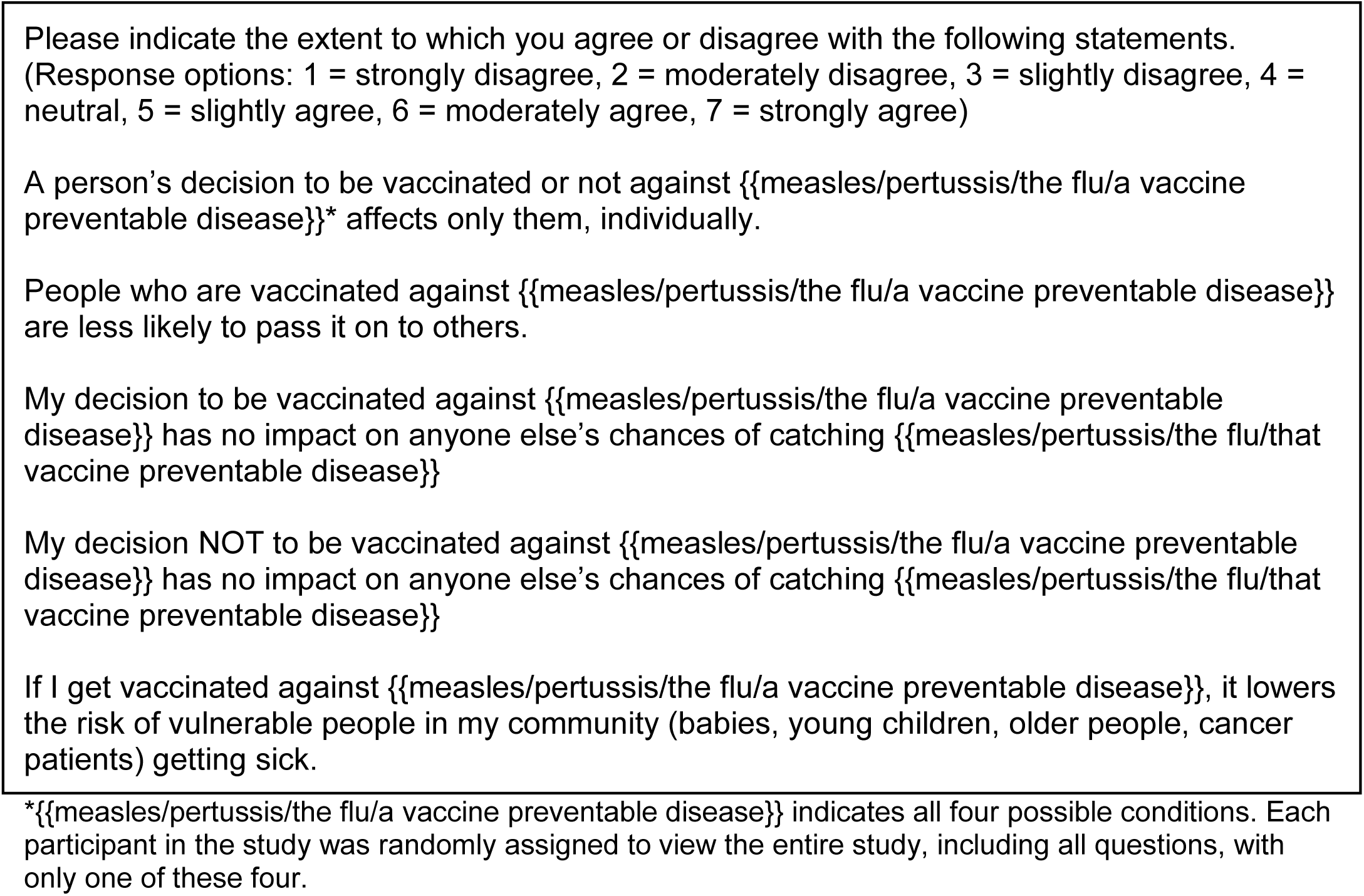
Subjective risk perception.

We measured emotions by asking participants to indicate their level of agreement using a 7-point Likert-type scale from strongly agree to strongly disagree with 5 items including, for example, “I am worried about people in my life (family, friends) getting {{measles/pertussis/flu/a vaccine preventable disease}},” and “I would feel guilty if someone in my life (a family member, a friend) got {{measles/pertussis/flu/a vaccine preventable disease}} from me”. We measured knowledge about community immunity by asking participants to respond to 15 items with response options that were either multiple choice or true/false/I don’t know. Items included, for example, ”A vaccine preventable disease can spread from one person to another person,” and, “Unvaccinated people in a population can be protected from infections when enough people in their community are vaccinated.” We measured vaccination intentions by asking people to indicate their likelihood of getting a free vaccine against the disease in question on a 100-point slider labeled “extremely unlikely, I would definitely NOT be vaccinated” at one end and “extremely likely, I would definitely BE vaccinated” at the other end. Specifically, the wording of the question was, “If you were eligible to receive a free vaccine against measles/pertussis/flu/a vaccine preventable disease, how likely would you be to get vaccinated?” Appendix 3 provides the complete questionnaire.

### Covariates and Moderating Variables

#### Individualism and collectivism

We used validated measures of individualism and collectivism from the cultural orientation scale [37] as moderating variables. This scale measures four dimensions of individualism and collectivism: horizontal individualism, vertical individualism, horizontal collectivism, and vertical collectivism. Horizontal Individualism is about, “seeing the self as fully autonomous, and believing that equality between individuals is the ideal.” Vertical Individualism is about, “seeing the self as fully autonomous, but recognizing that inequality will exist among individuals [and] accepting this inequality.” Horizontal Collectivism is about, “seeing the self as part of a collective but perceiving all the members of that collective as equal”, while Vertical Collectivism is about, “seeing the self as a part of a collective and being willing to accept hierarchy and inequality within that collective.” [38] All questions are answered on a 9-point Likert-type scale, where 1 indicates, “never or definitely no,” and 9 indicates, “always or definitely yes.” We included this scale to assess whether a person’s orientation towards individualism or collectivism might change the effects that *herdimm* might have on their risk perception, emotions, knowledge, and vaccination intentions.

#### Demographics

We used socio-demographic variables as covariates: born in Canada, age, gender, ethnicity, education level, income, disability, and preferred language. (All question wordings in English and French are available in Appendix 3.)

### Sample size

To determine the sample size for a 2*4 factorial randomized controlled trial using a two-way analysis of variance, assuming a small effect size (Cohen’s f=0.10), power of 80% (beta = 0.80), 5% type 1 error (alpha = 0.05), we determined that we required n=302 participants per group. We increased this to n=320 per group to allow for a secondary analysis (study reported separately) in which we would be comparing five groups in a one-way analysis of variance using only the English-speaking participants. The sample size of *herdimm* arms was then doubled to enable an embedded study (secondary to this work, not reported in this manuscript) of male versus female voice for the narration. This enabled more precise estimates of the effects of the herdimm intervention. We estimated the sample size by using G*power, version 3.1.9.2 [39] for a continuous outcome.

### Statistical analysis

To determine the effects of *herdimm* compared to control, we performed a two-way analyses of variance using the two independent variables (visualization, disease) and their interaction (visualization*disease) for continuous outcomes (subjective risk perception, knowledge, emotions) and logistic regression for dichotomous outcomes (objective risk perception, vaccination intentions.)

As planned in our pre-scripted statistical analyses written prior to collecting data, we transformed outcomes from continuous to dichotomous whenever the distribution did not respect the assumptions of the models described below; for example, if there were ceiling effects. This was the case for two variables: objective risk perception and vaccination intentions. For objective risk perception, we had set a minimum value of 80 out of 100 as a cutoff indicating high risk perception because our pre-testing had suggested that we might observe ceiling effects on this variable. For vaccination intentions, we had pre-scripted a median threshold in the event of model assumption violations. We therefore dichotomized vaccination intentions as being at/above the median or below the median.

We analyzed 3 models for each outcome. We chose our primary model to examine overall effects, and subsequent models to examine the robustness of results when adjusted for covariates or in the presence of moderators. Accordingly, we first determined the effects of factors (visualization and disease) without any covariates on each outcome (Model 1). For Model 2, we first examined our planned covariates and collapsed categories containing less than 5% of participants (that is ethnicity, language, education levels, and disability.) We used Model 2 to determine the effects of factors (visualization and disease) with adjustment for covariates (socio-demographic characteristics, individualism and collectivism.) Next, we determined the moderating effects of individualism and collectivism with adjustment for socio-demographic covariates (Model 3). Our overall 3-model approach is summarized in table 2 below.

**Table 2.** Models used for analysis.

| Model | Analytical goal |
| --- | --- |
| Model 1 (primary model) | Check for effects of factors without any covariates |
| Model 2 | Check for effects of factors with adjustment for covariates (individualism and collectivism, born in Canada, age, gender, ethnicity, education level, income, disability, and preferred language) |
| Model 3 | Check for moderating effects of individualism & collectivism with adjustment for other covariates (born in Canada, age, gender, |
|  | ethnicity, education level, income, disability, and preferred language) |

For simplicity here, we report only Model 1 results when these results remained consistent after adjusting for covariates in Model 2. When inclusion of covariates affected the results, we also report the adjusted (Model 2) results. We report results from Models 3 when these showed statistically-significant interactions. We report group-based means with 95% confidence intervals for continuous variables (i.e., subjective risk perception, knowledge, emotions) and probabilities of being at or above the median with 95% confidence intervals for the dichotomous variables (i.e., objective risk perception, vaccination intentions.)

To address missing outcome data, we also repeated all the primary models after using data imputation for any missing outcome values. To do this, fifteen multiple imputation datasets were generated. The primary author (HH) and a statistician (ASJ) compared results, examining whether results from analyses of the imputed datasets replicated or contradicted analyses from the original dataset.

We performed all analyses using R version 4.1.0 [40]. We used the package *psych* (version 1.9.12.31) [41] for descriptive analyses, the packages *stats*, *car* (version 3.0-8) [42] and *emmeans* [43] for the analyses of variance and logistic regressions, and the package *mice* (version 3.11.0) [44] for multiple imputation. We developed statistical code with the support of a statistician (ASJ), using simulated data prior to collecting study data. An external statistician peer reviewed the R code prior to conducting the study. We preregistered the statistical code in Open Science Framework on February 26th, 2021 (https://osf.io/hkysb/).

### Data collection methods

The trial ran from March 1 to July 1, 2021. After providing consent, each participant was randomized into a single study arm. All participants completed the same post-intervention questionnaire containing questions about risk perception, emotion, knowledge, vaccination intentions (see Appendix 3 for the complete questionnaire.)

### Data management

In order to be considered as contributing valid data, participants randomly allocated to intervention arms were required to spend a predetermined amount of time viewing and/or interacting with the intervention. These time requirements were set based on the minimum time possible for each intervention: control = no minimum restrictions; *herdimm* = minimum 206 seconds. To determine the minimum length of time, we calculated the shortest possible interaction time with the narrated animation, assuming very high internet speed and near-instantaneous transitions between the survey and the web application. Participants whose timestamps on the survey page were less than the predetermined minimum were excluded from the dataset. All the information we collected was kept confidential and used for research purposes only. Data was stored on Qualtrics servers located in Canada and subject to Canadian data privacy laws. Anonymized data are available at Université Laval’s Dataverse, Boréalis: https://doi.org/10.5683/SP3/41MWKO.

## Results

Of 18509 participants who started the study, 3883 participants were eligible for the analysis. Most excluded participants (n=10724) were excluded because their timestamps showed that they did not view the complete *herdimm* intervention. Figure 2 shows the CONSORT flow diagram.

**Figure 2.**
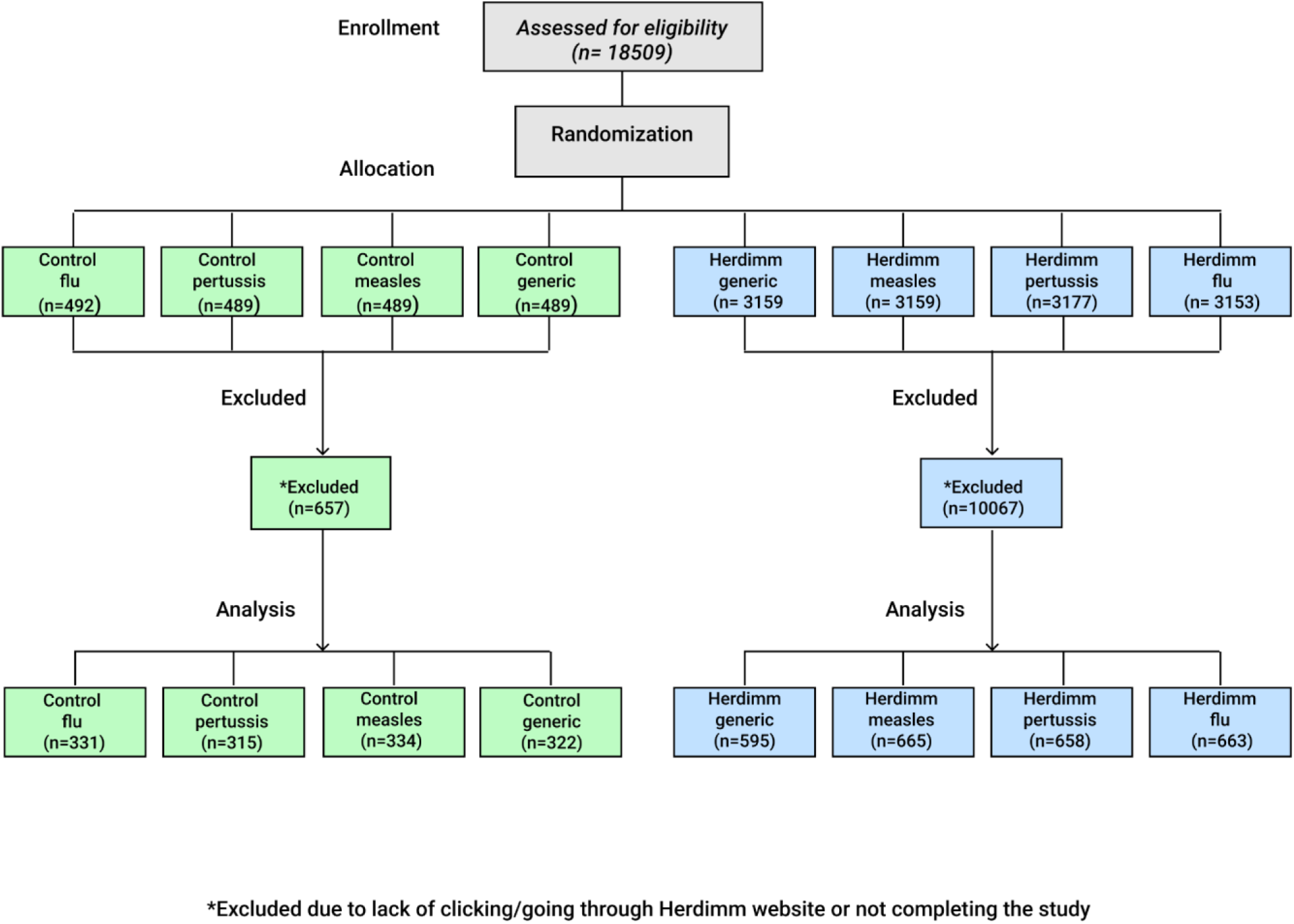
CONSORT flow diagram. *Note: Non-applicable elements of the CONSORT template were removed. There was no time interval between intervention allocation and follow-up*.

We collapsed covariate categories containing less than 5% of the total sample within that covariate. For example, when the people identifying as a particular ethnicity comprised less than 5% of the total sample, we collapsed that category with the closest other category within that covariate to facilitate analysis. Specifically, the covariate *ethnicity* (in which participants could select multiple options) was planned to have grouped categories Asian, Black, Indigenous, Maghrebian/Middle Eastern, white, and others (e.g., Latin American, self-reported “other”). However, because the Black, Indigenous, and Maghrebian/Middle Eastern groups each represented <5% of the total sample, categories were reduced to Asian, white, and others. Along the same lines, to account for low counts in some of the seven *education levels*, we recoded this covariate as *college degree and above* and *no college degree* (elementary school, high school diploma, or apprenticeship or trade certificate or diploma.) Similarly, due to low counts of people identifying as having a disability that specifically affects their ability to use technology like computers, we merged this covariate with the larger group of people identifying as having any disability. In the new, merged covariate *disability*, anyone indicating a disability of any kind (technical or other) was coded as having a disability. When no other category was available or when merging with another category would be inappropriate, as was the case for *gender identity*, we coded those data as missing rather than assigning those participants to a larger gender group. Similarly, the covariate *language* was planned to include French, English and other languages, but due to low counts for other languages, only the French and English categories remained.

Of 3883 participants, 50% identified as female, 49% as male. Eighty-one percent were comfortable communicating in English with their healthcare professional, and 24% in French. Eighty-two percent of the participants identified themselves as white, a category including white North American and white European. Ten percent identified themselves as Asian, a category including East, Central, South, Southeast Asian. Twenty-three percent participants reported a disability, while 75% reported no disability. Sixty-one percent of the participants reported higher education (college degree or higher) and 39% lower education (no college degree). Eighty-five percent of participants indicated their place of birth as in Canada. Table 3 shows details of participants’ characteristics across the study. Results per arm are shown in Appendix 4. Full results of all analyses using all models are available in Appendix 5.

**Table 3.** Participants’ characteristics (N=3883)

| Characteristic |  | Study population* |
| --- | --- | --- |
| **Age: median (IQR)** |  | 42 (32-57) |
| **Gender identity: n (%)** |  |  |
|  | Male | 1914 (49%) |
|  | Female | 1934 (50%) |
|  | Not available** | 35 (1%) |
| **Language(s) spoken: n (%)** |  |  |
| English | Yes | 3144 (81%) |
|  | No | 734 (19%) |
|  | Not available | 5 (0) |
| French | Yes | 937 (24%) |
|  | No | 2946 (76%) |
|  | Not available | - |
| **Ethnicity: n (%)** |  |  |
| White | Yes | 3172 (82%) |
|  | No | 645 (17%) |
|  | Not available | 66 (2%) |
| Asian | Yes | 385 (10%) |
|  | No | 3432 (88%) |
|  | Not available | 66 (2%) |
| Other | Yes | 269 (7%) |
|  | No | 3548 (91%) |
|  | Not available | 66 (2%) |
| **Disability: n (%)** | Yes | 883 (23%) |
|  | No | 2924 (75%) |
|  | Not available | 76 (2%) |
| **Education levels: n (%)** |  |  |
|  | College degree and above | 2361 (61%) |
|  | No college degree<br>(elementary school, high school, apprenticeship or trade certificate) | 1497 (39%) |
|  | Not available | 25 (1%) |
| **Born in Canada: n (%)** | Yes | 3303 (85%) |
|  | No | 536 (14%) |
|  | Not available | 44 (1%) |
| **Individualism and collectivism: mean (SD)** | Vertical individualism (e.g., "It is important that I do my job better than others.") | 5.12 (1.65) |
|  | Horizontal individualism (e.g., "I rely on myself most of the time, I rarely rely on others.") | 7.22 (1.16) |
|  | Vertical collectivism (e.g., "It is important to me that I respect the decisions made | 6.83 (1.39) |
|  | by my groups.”) |  |
|  | Horizontal collectivism (e.g., “I feel good when I cooperate with others.”) | 6.85 (1.33) |

\*Percentages may add up to +/-100% because of rounding.
\*\*Data could be not available due to participants choosing another option (e.g., gender-nonconforming), choosing, “I prefer not to answer,” or leaving the question unanswered.

### Primary outcome (risk perception)

#### Objective risk perception

Compared to participants in the control condition, those in the *herdimm* condition had significantly higher odds of providing a correct objective risk perception response (Chi-squared (1) = 134.54, p < 0.001). More specifically, people who were assigned to *herdimm* were more likely to understand that 1 unvaccinated person surrounded by 99 vaccinated people is more protected from infection than 1 vaccinated person surrounded by 99 unvaccinated people (58.0%, 95% confidence interval 56.0%-59.9%) compared to those assigned to the control condition (38.2%, 95% confidence interval 35.5%-40.9%). There was no main effect of disease (measles, pertussis, flu, generic vaccine-preventable disease) (Chi-squared (3) = 6.94, p = 0.074), and no statistically-significant interaction between intervention and disease (Chi-squared (3) = 1.19, p = 0.755). These results did not change when including covariates (Model 2) or when testing for moderation effects (Model 3).

#### Subjective risk perception

Participants assigned to the *herdimm* condition had higher subjective risk perception compared to those assigned to the control condition (F(1,3875) = 28.7856, p < 0.001). Subjective risk perception also differed whether the disease was presented as measles, pertussis, flu, or a generic “vaccine preventable disease” (F(3,3875) = 5.6430, p < 0.001). We also observed an interaction between intervention and disease on subjective risk perception. More specifically, subjective risk perception was higher among people who were assigned to *herdimm* compared to those assigned to the control. Differences estimates for each disease showed that this effect was likely driven by the larger difference among people assigned to the flu condition (difference estimate = 0.456, standard error = 0.0869, t(3875) = -5.243, p < 0.0001; all difference estimates available in Appendix 5). Table 4 shows means and 95% confidence intervals for subjective risk perception for each combination of intervention and disease.

**Table 4.** Two way analysis of variance of subjective risk perception.

|  | Intervention |  |  |
| --- | --- | --- | --- |
| Disease | Control: mean (95% confidence interval) | Herdimm: mean (95% confidence interval) |  |
| Generic | 5.31 (5.17-5.45) | 5.46 (5.36-5.56) | $F(3,3875) = 5.6430$, $p < 0.001$ |
| Measles | 5.44 (5.30-5.58) | 5.61 (5.51-5.71) |  |
| Pertussis | 5.39 (5.25-5.54) | 5.56 (5.46-5.66) |  |
| Flu | 5.06 (4.92-5.20) | 5.52 (5.42-5.62) |  |
|  | F(1,3875) = 28.7856, p < 0.001 |  | F(3,3875) = 2.8324, p = 0.037 |

**Abbreviations:** *p*, *p*-value; *F*, *F*-statistic

Our second model showed that intervention and disease interaction continued to demonstrate the same effects on subjective risk perception after adjustment for covariates, and is driven by the effects of the *herdimm* intervention among participants assigned to measles, pertussis and flu. The third model suggested that the increase in subjective risk perception for *herdimm* versus control was somewhat driven by the responses of people who scored low on vertical individualism (i.e., who disagreed with statements like, “It is important that I do my job better than others,”) (see Figure 3) and low on horizontal collectivism (i.e., people who disagreed with statements like, “I feel good when I cooperate with others.”) The third model further suggested that the relationship between participants’ horizontal individualism scores (i.e., their agreement with statements like, “I rely on myself most of the time, I rarely rely on others,”) and subjective risk perception was negative among those randomized to the flu condition while it was positive among those randomized to other diseases.

**Figure 3.**
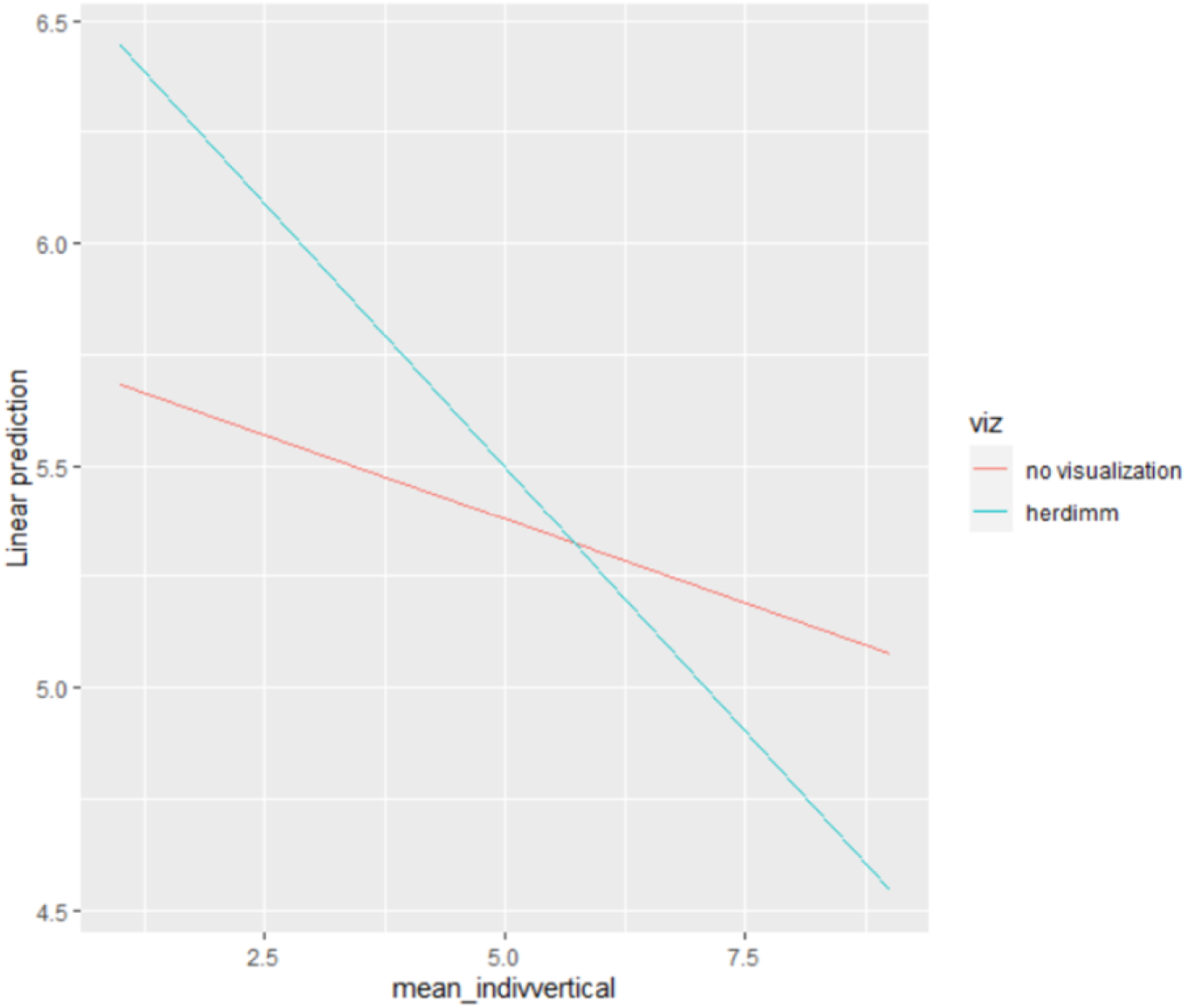
Subjective risk perception for *herdimm* versus control by participants’ vertical individualism.

### Secondary outcomes

For our second research question we assessed the effects of the *herdimm* intervention on other outcomes (namely, emotions, knowledge and vaccination intentions) compared to a control of no intervention.

#### Emotions

The primary model showed a statistically significant main effect of the intervention (*herdimm* or control) (F(1,3875) = 13.13, p < 0.001), main effect of disease (measles, pertussis, flu, generic vaccine-preventable disease) F(3,3875) = 78.54, p < 0.001), and no statistically significant interaction between intervention and disease on emotions F(3,3875) = 1.49, p = 0.216. More specifically, people who were assigned to *herdimm* reported stronger emotional responses (mean 5.07, 95% confidence interval 5.02-5.13) compared to those assigned to control (mean 4.90, 95% confidence interval 4.83-4.98). In other words, they were more likely to indicate higher levels of agreement with statements like, “I am worried about people in my life (family, friends) getting a vaccine-preventable disease,” and “I would feel guilty if someone in my life (a family member, a friend) got a vaccine-preventable disease from me.” People assigned to the generic ‘vaccine-preventable disease’ condition (mean 5.48, 95% confidence interval 5.39-5.57) had a higher mean score on emotions than those assigned to any specific diseases. People assigned to the measles (mean 4.59, 95% confidence interval 4.50-4.68) and pertussis conditions (mean 4.70, 95% confidence interval 4.6-4.79) had lower mean emotions than those assigned to flu (mean 5.18, 95% confidence interval 5.09-5.27).

The second model showed that inclusion of planned covariates, which required excluding 419 participants due to missing covariates, changed the results. The effect of the intervention became not significant (F(1,3440) = 2.93, p = 0.087) and the effect of disease remained significant (F(3,3440) = 75.47, p < 0.001), while the interaction between intervention and disease remained not significant. The third model suggested that the higher mean on emotions for *herdimm* was driven by the responses of people who scored low on horizontal collectivism (i.e., people who disagreed with statements like, “I feel good when I cooperate with others”) in the generic disease and who scored high on horizontal collectivism (i.e., people who agreed with the above statement) for measles and pertussis.

#### Knowledge

The primary model showed a statistically significant main effect of the interventions (F(1,3875) = 36.37, p < 0.001), main effect of disease (measles, pertussis, flu, generic vaccine-preventable disease) (F(3,3875) = 5.20, p = 0.001), and no statistically significant interaction between intervention and disease on knowledge (F(3,3875) = 2.54, p = 0.055). More specifically, people assigned to *herdimm* had higher mean knowledge (mean 9.12, 95% confidence interval 9.02-9.22) compared to those assigned to the control (mean 8.58, 95% confidence interval 8.44-8.73). People who were assigned to the generic vaccine-preventable disease had higher mean knowledge than those assigned to measles (difference estimate = 0.410, standard error = 0.127, t(3875) = 3.239, p = 0.007) and pertussis (difference estimate = 0.337, standard error = 0.128, t(3875) = 2.631, p = 0.042). Those assigned to disease flu also had higher mean knowledge than those assigned to measles (difference estimate = 0.357, standard error = 0.125, t(3875) = 2.861, p = 0.022).

The second model showed that, after inclusion of planned covariates (thus excluding 419 participants with missing covariate data), the effect of the interaction between intervention and disease became significant as well (F(3,3440) = 3.2962, p = 0.0196). After adjusting for covariates, people assigned to *herdimm* had higher mean knowledge than those assigned the control when they were also assigned to measles (difference estimate = 0.844, standard error = 0.182, t(3440) = 4.627, p < 0.001), pertussis (difference estimate = 0.508, standard error = 0.187, t(3440) = 2.725, p = 0.007) or flu (difference estimate = 0.680, standard error = 0.182, t(3440) = 3.743, p < 0.001) but not when they were assigned to generic vaccine-preventable disease (difference estimate = 0.076, standard error = 0.184, t(3440) = 0.413, p = 0.680).

The third model suggested that the higher mean knowledge for *herdimm* was driven by the responses of people who scored low on vertical individualism (i.e., who disagreed with statements like, “It is important that I do my job better than others.”) To explore whether people who scored differently on individualism and collectivism subscales might have paid more or less attention to the information in *herdimm*, we conducted additional exploratory posthoc analyses on time spent away from the Qualtrics survey, when participants were supposed to be viewing *herdimm*. Overall, differences appeared to be minimal (Appendix 6)

#### Vaccination intentions

The primary logistic regression model showed a statistically significant main effect of the intervention (*herdimm* or control) (Chi-squared (1) = 9.41, p = 0.002), main effect of disease (measles, pertussis, flu, generic vaccine-preventable disease) (Chi-squared (3) = 15.02, p = 0.002), and no statistically significant interaction between intervention and disease on vaccination intentions (Chi-squared (3) = 3.72, p = 0.293). More specifically, people assigned to *herdimm* were more likely to score high on vaccination intentions (52.1%, 95% confidence interval 50.1%-54.0%) compared to those assigned to the control (46.9%, 95% confidence interval 44.1%-49.6%). People who were assigned to the flu (45.8%, 95% confidence interval 42.5% - 49.1%) were less likely to score high on vaccination intentions compared to those assigned to a generic ‘vaccine-preventable disease’ (52.7%, 95% confidence interval 49.3%-56.1%) and measles (52.9%, 95% confidence interval 49.5%-56.1%). Those assigned to measles were also more likely to score high on vaccination intentions compared to those assigned to pertussis (46.6%, 95% confidence interval 43.2%-49.9%).

The second model showed that inclusion of planned covariates (and its according exclusion of 419 participants who did not provide sociodemographic information) changed the results. The effect of the intervention became not significant (Chi-squared (1) = 2.69, p=0.10). The covariates with statistically significant effects at the 5% level on vaccination intentions were age (Chi-squared (1) = 56.646, p<0.001), collective horizontalism (Chi-squared (1) = 46.271, p<0.001), income (Chi-squared (3) = 42.401, p<0.001), disability (Chi-squared (1) = 19.7276, p<0.001), vertical individualism (Chi-squared (1) = 6.422, p= 0.011), horizontal individualism (Chi-squared (1) = 4.261, p= 0.04), and education level (Chi-squared (1) = 4.141, p= 0.042). The third model suggested that the relationship between participants’ vertical individualism scores (i.e., their agreement with statements like, “I rely on myself most of the time, I rarely rely on others,”) and vaccination intentions was positive among those randomized to the flu condition while it was negative among those randomized to other diseases. In addition to moderation model results available above and in Appendix 5, Appendix 6 presents graphs of all outcome data, including vaccination intentions, by Collectivism and Individualism subscales.

### Impact of missing outcome data

Comparing results from analyses with multiple imputation on missing outcome data to the analyses without, we observed no notable differences. Some outcomes (subjective risk perception, knowledge, and emotion) had no missing values, and there were no changes from original results for the primary model. The lack of missing data might be because these variables are a function of several items, and therefore, even if some items were missing, a mean or sum could still be calculated. For the other outcomes measured by a single item (objective risk perception, vaccination intention) the rate of missing data was approximately 1%. After imputing these missing data points, there were no changes to the results of any analyses.

## Discussion

This study offers 4 key findings. First, compared to the control condition of no education, the *herdimm* web app improved objective risk perception, increased subjective risk perception, improved knowledge about how herd immunity or community immunity works, increased emotions such as concern for vulnerable people, and increased intentions to receive vaccines. People often perceive risk based on their impressions of overall disease prevalence and severity [45,46]. By presenting epidemiological evidence about how vaccines protect whole communities in a way that allowed people across backgrounds, education levels, and disability statuses to understand it, the *herdimm* intervention led to improved understanding of levels of risk (objective risk perception) and increased subjective risk perception and associated emotions, indicating higher levels of concern for others in one’s community and vulnerable people within it. This finding is consistent with the previous research in which risk perception and emotions influenced vaccine decisions [47–49] and similar web-based interventions improved vaccine knowledge, attitudes, and intentions to receive vaccines [10,50–52]. Our study therefore adds to previous research suggesting that communicating population-level benefits of vaccination may help encourage vaccine uptake, at least in some populations.

Second, our study showed that people with more collectivist orientations are more responsive to messages demonstrating collective benefits of widespread vaccination than people with more individualist orientations. Previous work by Betsch and colleagues analyzed the effects of a similar intervention across countries demonstrating more individualist and collectivist orientations at the population level. They found that their intervention had larger effects in encouraging vaccination intentions in countries with higher overall individualist scores (United States, Germany and the Netherlands) than in collectivist countries with higher overall collectivist scores (South Korea, Vietnam and Hong Kong) because baseline vaccine uptake was already high in the second group, and there was therefore less room for change [10]. Our study built on this finding by incorporating a validated scale of individualism and collectivism [37] to assess whether a person’s orientation towards individualism or collectivism moderated how they responded to the intervention. Our study was conducted in Canada, a multicultural, Western country that was not represented in Betsch and colleagues’ study but would typically be grouped with the United States, Germany and the Netherlands, rather than with South Korea, Vietnam and Hong Kong. Overall, our findings suggested that people with more collectivist orientations were more responsive to the community immunity idea than those with more individualist orientations. In other words, by conducting within-country rather than between-country analyses, we found that some people are more collectivist while others are more individualist, and these individual differences may influence how people respond to public health appeals to protect others. Other research by Amin and colleagues has similarly shown that individuals’ orientations toward purity and liberty as moral foundations are associated with vaccine hesitancy, while the moral foundations of harm and fairness that are more commonly emphasized in messaging around vaccines were not associated with vaccine hesitancy or uptake [53]. Taken together, such findings emphasize the importance of potentially tailoring approaches to vaccine messaging according to individuals’ broader values, worldviews, and orientations towards self and others [54].

Third, although most findings were consistent across the four disease conditions we used in our study, we did observe signals of potential disease-specific findings that offer potential implications or avenues for future work. One such signal occurred on the outcome of subjective risk perception, which was higher overall among people who were assigned to the *herdimm* intervention compared to those assigned to the control condition, but particularly so among people assigned to the influenza condition due to lower baseline subjective risk perception among people who completed the study in the context of influenza. Specifically, people assigned to the control condition in the context of influenza indicated lower subjective risk perception, meaning they were less likely to agree with statements like, “If I get vaccinated against the flu, it lowers the risk of vulnerable people in my community (babies, young children, older people, cancer patients) getting sick,” compared to those asked to indicate their level of agreement with those same statements but with terms measles, pertussis, or a vaccine-preventable disease in place of, “the flu.” This is consistent with lower uptake of influenza vaccines in many jurisdictions compared to uptake of vaccines against measles, pertussis, and many other diseases. Among those assigned to the *herdimm* intervention, however, subjective risk perception was similar across diseases. This may imply that helping people understand how community immunity works may be particularly useful in the context of influenza, a condition for which vaccine uptake is often lower, especially among those at lower risk of severe influenza [55].

Fourth and finally, our study also offers some methodological lessons for future evaluations of similar digital health interventions that may be useful to other research groups. Participants in our study assigned to the *herdimm* condition had to answer sociodemographic questions in Qualtrics, click on a link to visit a university-based website hosting the *herdimm* intervention, then return to their Qualtrics questionnaire to answer questions making up the outcome measures. We observed very high dropout rates due to people either abandoning the study at the point of visiting the university-based website, or completing the outcome questions with insufficient time in between to have actually visited the *herdimm* website. A related study our group conducted in which we adapted the *herdimm* intervention to show why social distancing was being recommended to help reduce COVID-19 transmission showed that the largest proportion of people who abandoned the university-based website in that study did so at the step of creating 8 additional avatars to represent the people around the individual [56]. For future research, we therefore added the option for people to auto-generate additional avatars. Additionally, rather than simply instructing participants to visit an external site and return to the questionnaire to complete the study, in future studies in which we need to ask participants to visit an external site, we plan to use individualized links containing the anonymized Qualtrics respondent IDs as parameters within the link, then provide an individualized link back to their Qualtrics questionnaire. Although this is a more complex procedure, it will ensure better intervention fidelity as well as allow the collection of data about people’s behaviour at the website (for example, time spent on each step, choice of avatars, etc.) for analysis. Additionally, our planned analyses for this study included plans for multiple imputation to account for missing outcome data. However, we did not pre-plan similar imputation for missing sociodemographic data, meaning that our adjusted analyses removed about 11% of participants while also adding variables, and therefore had lower statistical power. Future studies should include plans for addressing missing data among covariates to avoid such diminished statistical power.

### Strengths and Limitations

Our study had five key limitations. First, some outcomes were evaluated with items developed specifically for this study due to the lack of validated scales in the context of communication about community immunity. While we pre-tested our measures and examined standard psychometric indicators of validity such as Cronbach alpha, full validation studies were outside the scope of this work. Second, as noted above, we observed high dropout rates in *herdimm* arms, likely because people had to visit the *herdimm* website in the middle of the questionnaire and then come back to finish the study. This means that the effects of the *herdimm* intervention may be either overestimated or underestimated. To explore this issue, we examined sociodemographic differences between people who did and did not complete the study. The largest difference we observed was among participants who self-identified as white, who were more populous among the group who completed the study (80% among completes versus 62% among incompletes.) Identifying as white was also associated with higher knowledge. This means that the observed effects of *herdimm* on knowledge may be overestimated because, in the final dataset, there were more people with a characteristic associated with higher knowledge in the intervention arms. Alternatively, effects may be underestimated if the higher knowledge scores are attributable to pre-existing knowledge. Third, some identity elements (e.g., some racial and ethnic groups) had to be merged due to low numbers. As a result, we do not know the influence of specific groups, e.g., Black Caribbean, as a covariate on our outcomes. This may be a limitation if such covariates are influential on our outcomes. However, given that the racial and ethnic groups that we did retain had little influence as covariates, it is unlikely that collapsing groups would have had a substantial influence. Fourth, the distribution of data on a number of continuous or quasi-continuous outcomes required establishing a cutoff to perform logistic regressions. Although we prepared our statistical code before collecting data and preregistered our analyses, we had little prior data to guide our choices for such cutoffs, meaning that the cutoffs were necessarily somewhat arbitrary. Fifth and finally, our study took place during early COVID-19 vaccination campaigns in Canada. We did not add COVID-19 as a named vaccine-preventable disease in our study for a number of reasons, including that it was not yet known whether the early COVID-19 vaccines would be able to produce robust community immunity. It is possible, and perhaps likely, that participants assigned to the generic, “vaccine-preventable disease,” condition may have been thinking of COVID-19 as they responded to the study questions, which may have influenced their responses in one direction or another. However, for the vast majority of our outcomes, we did not observe significant differences between people assigned to the generic disease and those assigned to measles, pertussis, and flu. This suggests that if participants were thinking of COVID-19, their responses were nonetheless similar to the responses of people thinking of other diseases.

Our study also had three key strengths. First, for methodological robustness, we preregistered the entire study, including all study materials and statistical code developed with simulated data. Such preregistration may have been particularly important for this study, given that we anticipated the need to divide risk perception into objective and subjective components and use logistic regressions for a number of continuous outcomes. Second, we evaluated the *herdimm* intervention across four disease conditions–three named diseases and an unnamed, “vaccine-preventable disease.” Much research on vaccine decision making is conducted on only one vaccine-preventable disease at a time or without ever naming a particular disease, thus limiting its generalizability. People may respond differently to a situation involving an abstract, unnamed disease than they do to a known, named disease, and yet we cannot assume that responses to messaging about one specific disease will be applicable to messaging about another specific disease. Identifying approaches that work across diseases is important for achieving broader public health goals. Studies such as ours therefore offer findings that are more robust than they would have been had we used only a single disease. Third, the *herdimm* intervention offers insights about the potential applicability of personalization and tailoring to vaccine messaging. By having people create their own avatar, the avatars of people around them, and then using those avatars in an animation, the *herdimm* intervention aimed to help people literally see how their vaccination decisions could impact others around them. This message was more impactful among people with more collectivist orientations, underlining the idea that although public health messages are often universal, delivering them differently to different people may increase their impact.

## Conclusions

Visually conveying the concept of community immunity using personalized avatars improves objective and subjective risk perception, emotions, knowledge and increases intentions to receive vaccines. Our work also contributes evidence of how community immunity messages may influence vaccine acceptance differently among people whose personal orientations are more collectivist versus those who are more individualist, which offers insights about how we might target messages to different people. Put another way, conveying the concept of community immunity in understandable ways may contribute to public health efforts to achieve adequate vaccine coverage. However, this needs to be done with care, as not everyone will respond to messages about protecting others in the same way.

## Data Availability

All anonymized data are available at Université Laval's Dataverse, Boréalis: https://doi.org/10.5683/SP3/41MWKO. All study materials were pre-registered and are available on Open Science Framework (https://osf.io/hkysb/). Web development code is available on GitHub: https://github.com/Witteman-Lab/herdimm.

## DECLARATIONS

### Ethics approval and consent to participate

This project is approved by the “Comité d’éthique de la recherche en sciences de la santé” ethics committee of Laval University (Approval No. 2017-137 Phase II / 03-09-2019). All participants included in the study consented to participate in the study.

### Consent for publication

#### Availability of data and materials

All study materials were pre-registered and are available on Open Science Framework (https://osf.io/hkysb/). Web development code is available on GitHub: https://github.com/Witteman-Lab/herdimm. Anonymized data are available at Université Laval’s Dataverse, Boréalis: https://doi.org/10.5683/SP3/41MWKO.

#### Conflict of interest

## None

## Acknowledgments

The authors gratefully acknowledge the contributions of Mehdi Aouami and Rosalie Baker for designing the first version of our prototype, and Ali Ben Charif for constructive comments on earlier versions of this paper.

## Authors’ Contributions

## Funding

This study was funded by the Canadian Institutes of Health Research (grant number FDN-148426, 2016-2021, PI: Witteman). The CIHR had no role in determining the study design, the plans for data collection or analysis, the decision to publish, nor the preparation of this manuscript. HOW is funded by a Tier 2 Canada Research Chair in Human-Centred Digital Health. Dr. Sander is funded by a Tier 1 Canada Research Chair in Economics of Infectious Diseases (CRC-2022-00362). Dr. Scherer was supported by the National Institute on Aging of the National Institutes of Health under Award Number K01AG065440. The content is solely the responsibility of the authors and does not necessarily represent the official views of NIH.

## Abbreviation

OSF: Open Science Framewor
(IQR): Interquartile range
CI: Confidence interval
Df: Degree of freedom
p: p-value
F: F-statistic
LR Chisq: Likelihood ratio tests
ANOVA: Analysis of variance

## Appendices

## Appendix 1: Links or video of herdimm intervention

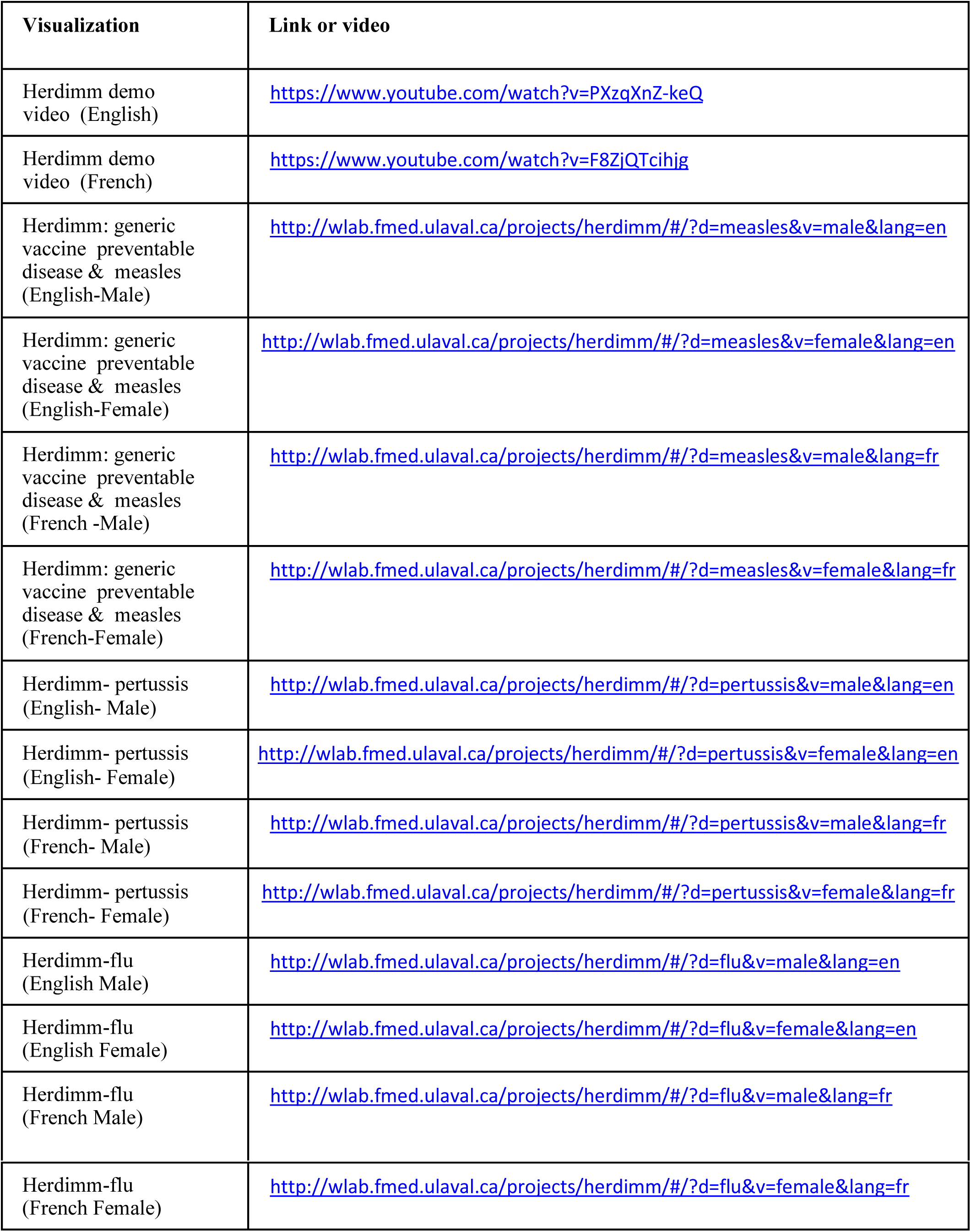

## Appendix 2: Scatterplot and bland-altman for risk perception

**S1:**
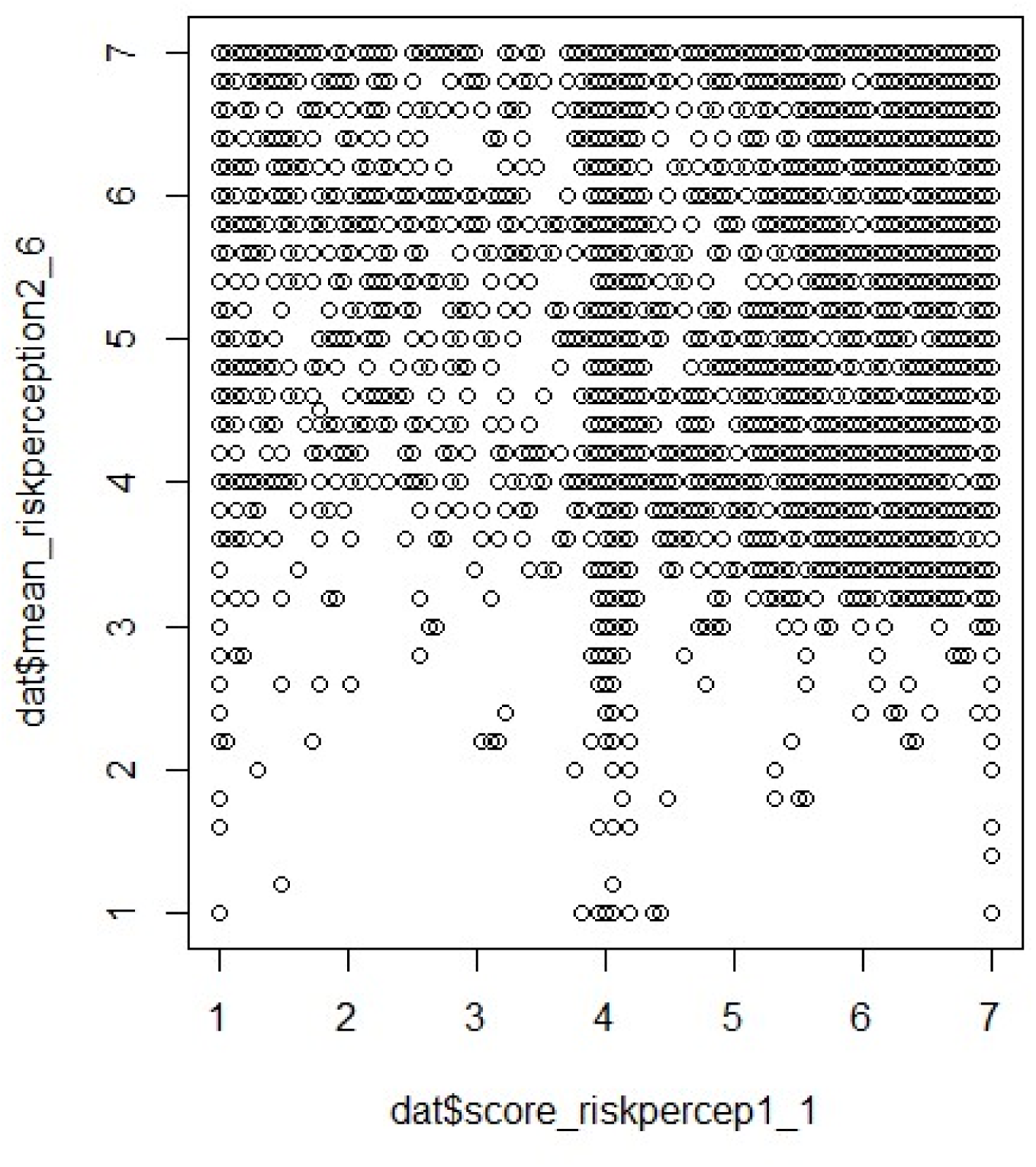
Scatter plot to see if the item risk percep_1 (objective risk perception) and risk percept_2 to 6 (subjective risk perception) are all measuring the same thing

**S2:**
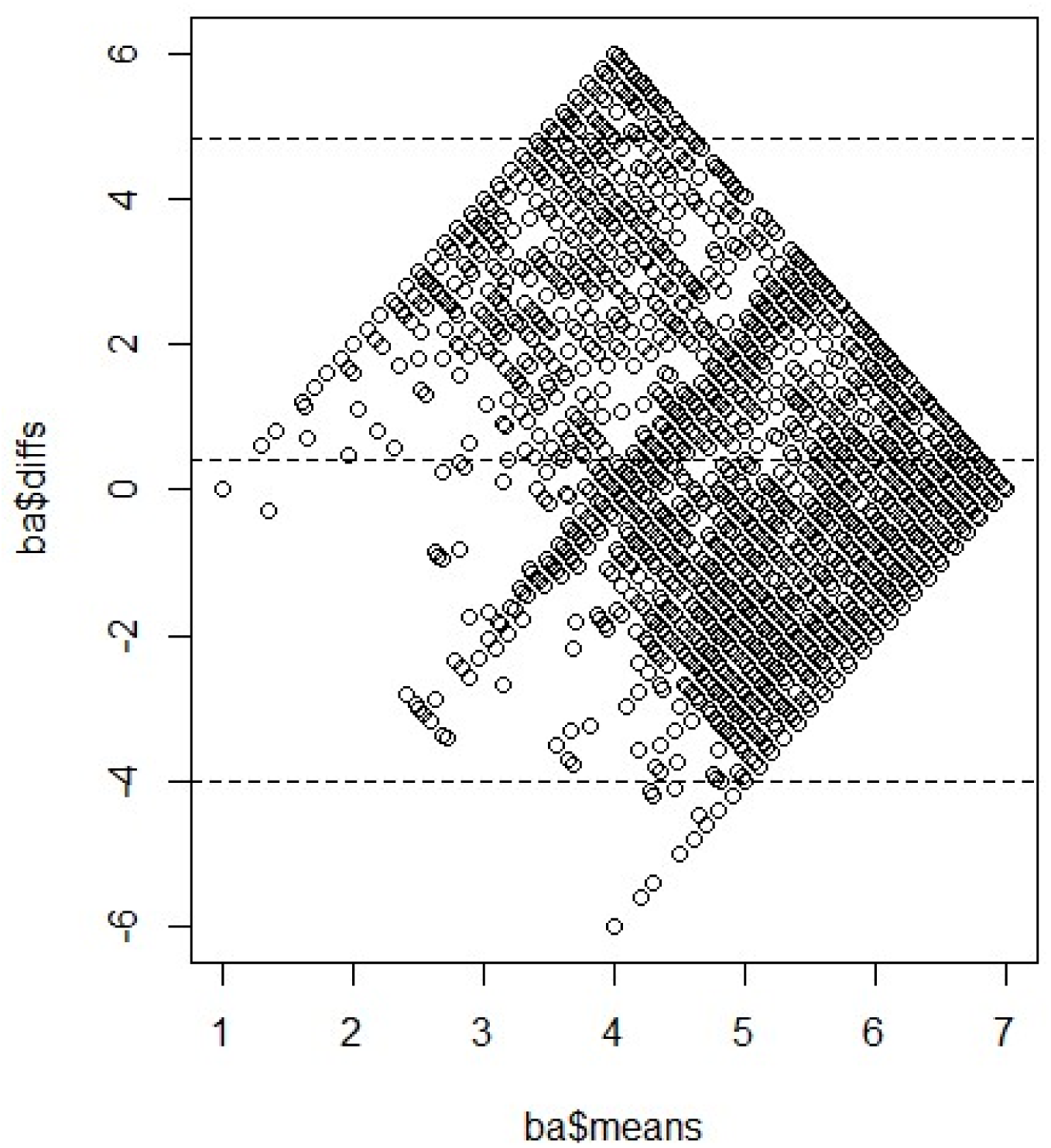
Bland altman to see if the item risk percep_1 and risk percept_2 to 6 are all measuring the same thing

## Appendix 3: *Herdimm* survey quesstionnaire

### contag_phase3b

Intro **Project Description**

#### Project Description

##### Introduction

Thank you for participating in this study. This project is a collaboration with University Laval, Public Health Ontario (PHO) and The National Institute of Public Health of Quebec (INSPQ).

In this study, we will first give you some information to do with contagious diseases and then ask you to answer some questions.

The survey will take about 15 to 20 minutes. Your name will not be recorded anywhere in this survey. Researchers will not be able to associate your name with your answers. If you choose to leave the study, you can stop at any time. You may choose not to answer any questions you don’t want to answer. If, for some reason, you cannot complete the survey, you may **restart** the survey at a later time by clicking on the survey link in your email invitation. You will have to **restart** the survey from the beginning.

If you become worried about your health while taking this survey, please talk to a healthcare provider.

##### About the researchers

This study is a PhD project conducted by Ms. Hina Hakim, PhD student, supervised by Holly O. Witteman, PhD, professor and researcher in the Faculty of Medicine at Laval University. Ms.

Hakim is co-supervised by Dr. Daniel Reinharz, professor and researcher in the Faculty of Medicine at Laval University.

##### Purpose of the study

The purpose of this study is to evaluate the effects of information to do with contagious diseases on risk perception, knowledge, emotions, attitudes, beliefs, and behavioural intentions.

##### What your participation entails

Participating in this study will involve following:

- Answering survey questions.
- You may be asked to view or listen to information, which will require either the ability to read or hear.
- You may be asked to briefly visit another website in the middle the survey.

##### Risks and benefits

The risks associated with participating in the study, specifically:

- Information and questions, we ask may make you feel uncomfortable or anxious. We suggest you to consult your healthcare professional, as the case may be;
- You may feel some discomfort having to focus on a computer screen.
- You may feel some fatigue from concentrating throughout the survey.

There is no personal benefit to participate in this study, however, the information we gather will help us to evaluate the effects of the information, which may help in improving the way health information is shared, and population health.

Your help means a lot to us. We thank you again for taking the time to complete this survey.

##### Confidentiality and data protection

All information we collect will be confidential and used only for research purposes. In other words:

- Participant credentials will not be associated with the results of the study and no individual information will be presented in reports, publications or presentations.
- All data will be presented in aggregate form without individual identifiers.
- Data will be stored on Qualtrics servers located in Canada. When we work with data, the only people who will have access to our Qualtrics account will be the investigators and our team members who have complete and relevant ethics training. When the data is stored on our computers, each of our computers will be protected by a password. The data will be stored on a secure server, to which access is maintained and reserved for the members of the team (members of the team affiliated with Université Laval).

After the end of the study:

- Anonymized data (answers to questions in the survey including socio-demographic information) will be deposited in a public repository (Dataverse de l’Université Laval (Laval University)) which will allow data sharing with the scientific community. No information that would allow anyone to identify a person will be deposited in this public repository.
- Any other electronic data will be destroyed in June 2027.

##### Contact people

If you have any questions about the research study, or if you experience a problem as a result of participating in the study, please contact:

Holly Witteman

Laval University

Tel: (418) 656-2131 ext: 403981

If you have concerns about your rights as a participant in this study or any complaints, you can communicate with the University Laval Ombudsman at:

Pavillon Alphonse-Desjardins

2325, rue de l’Université, Local 3320

Québec, QC G1V 0A6

Tel: 418-656-3081

Toll free: 1-866-323-2271

*This project has been approved by Laval University Research Ethics Board: Approval No. 2017-137 Phase II A-6/ 10-09-2020*

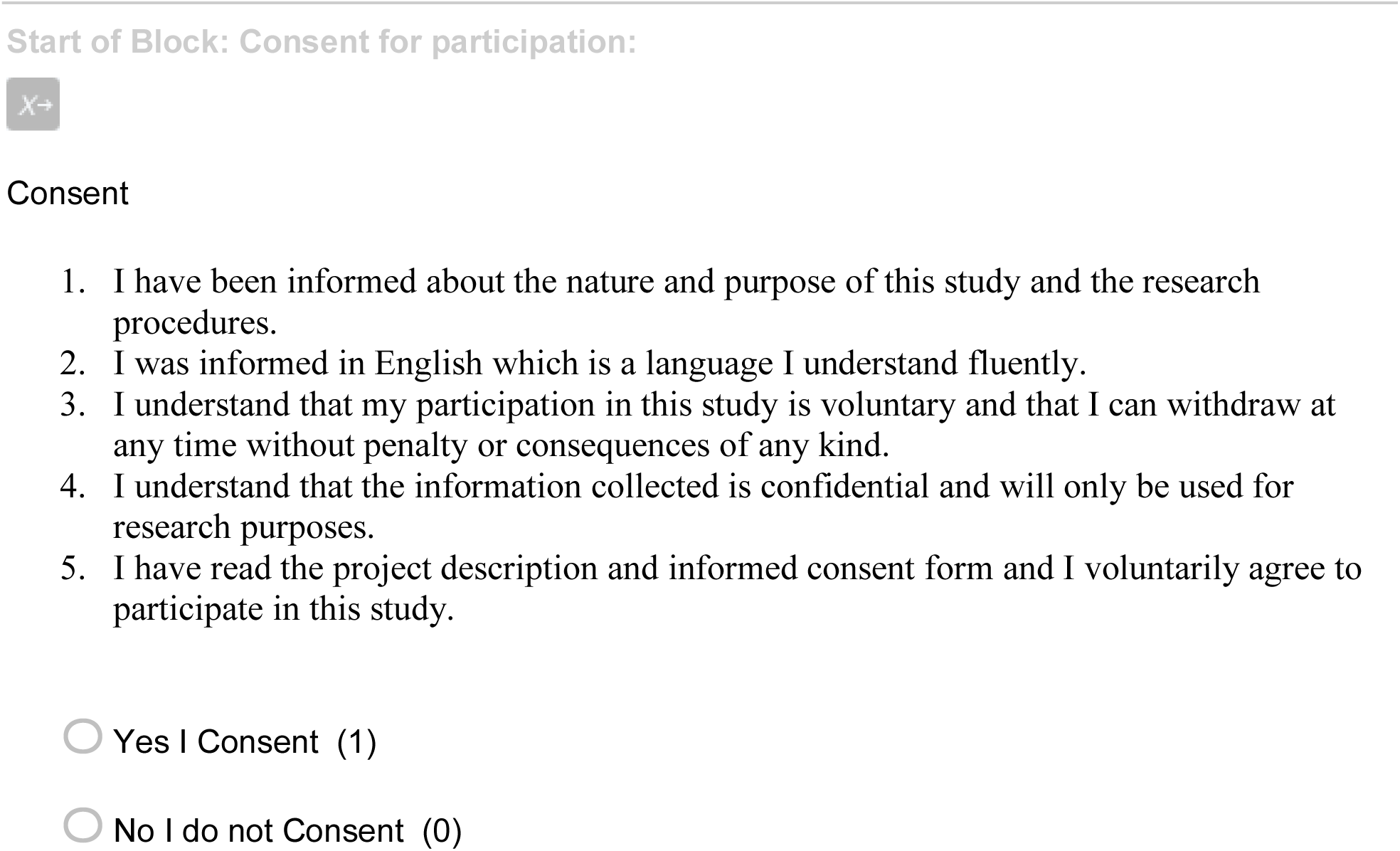
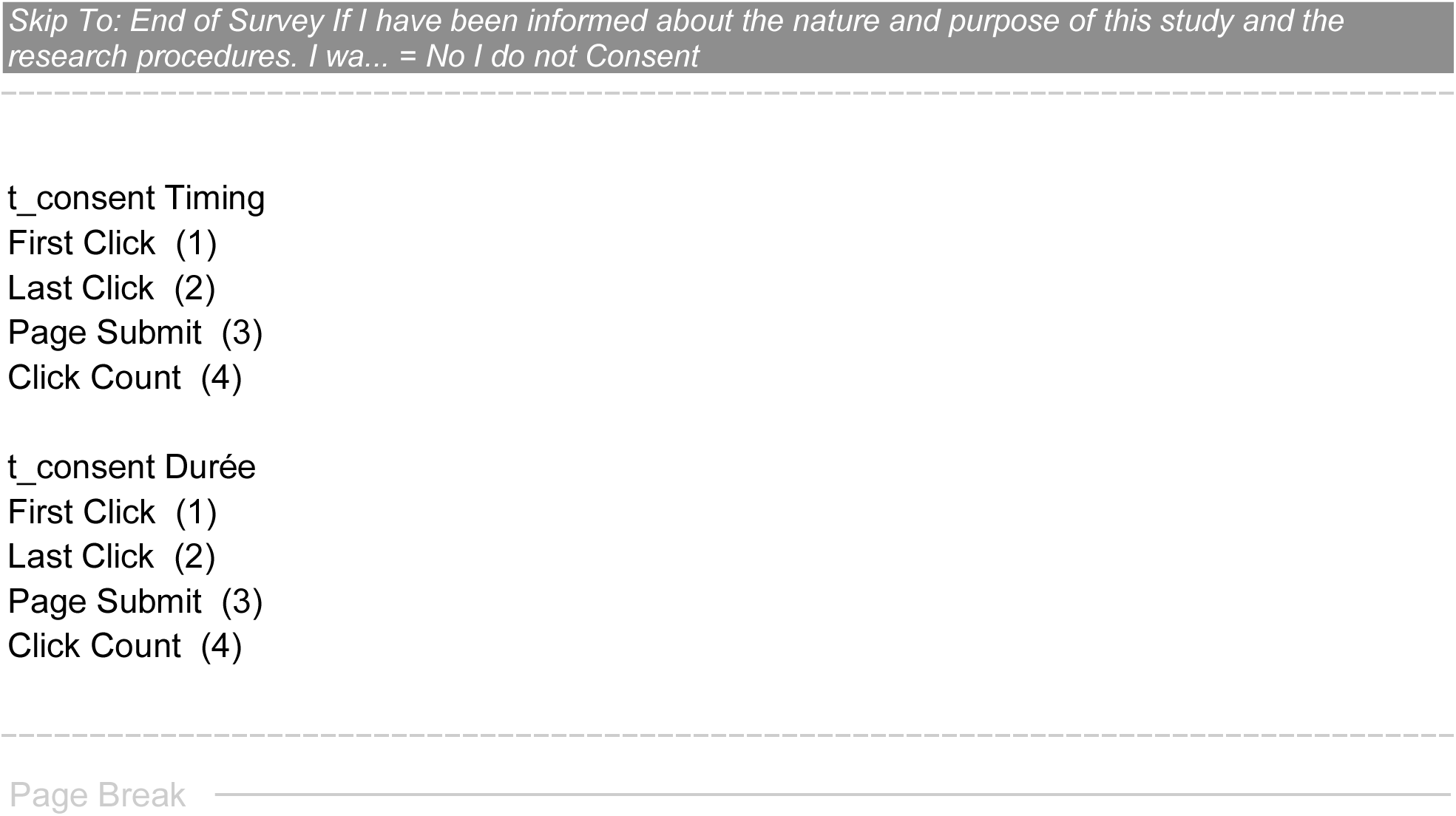
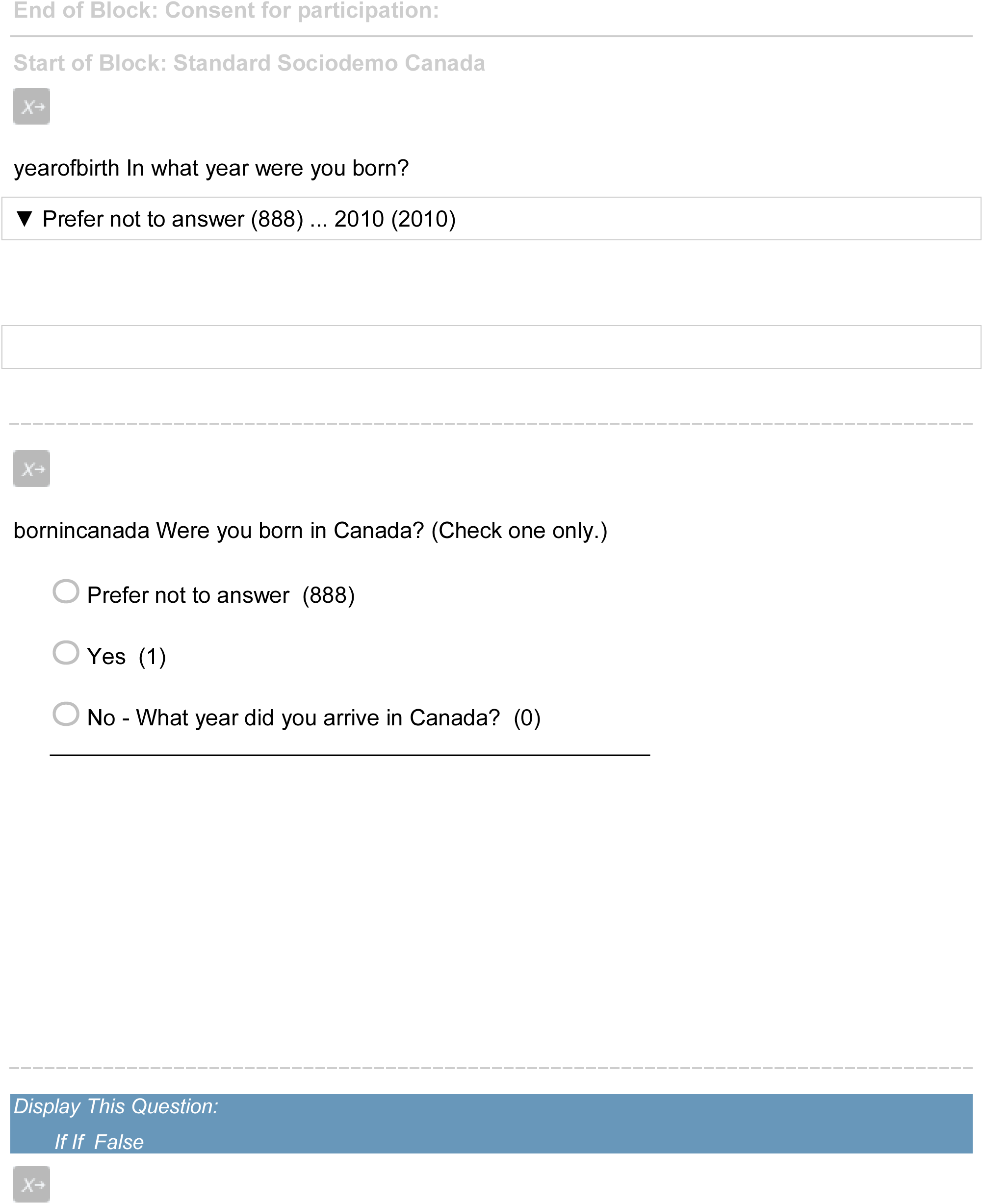
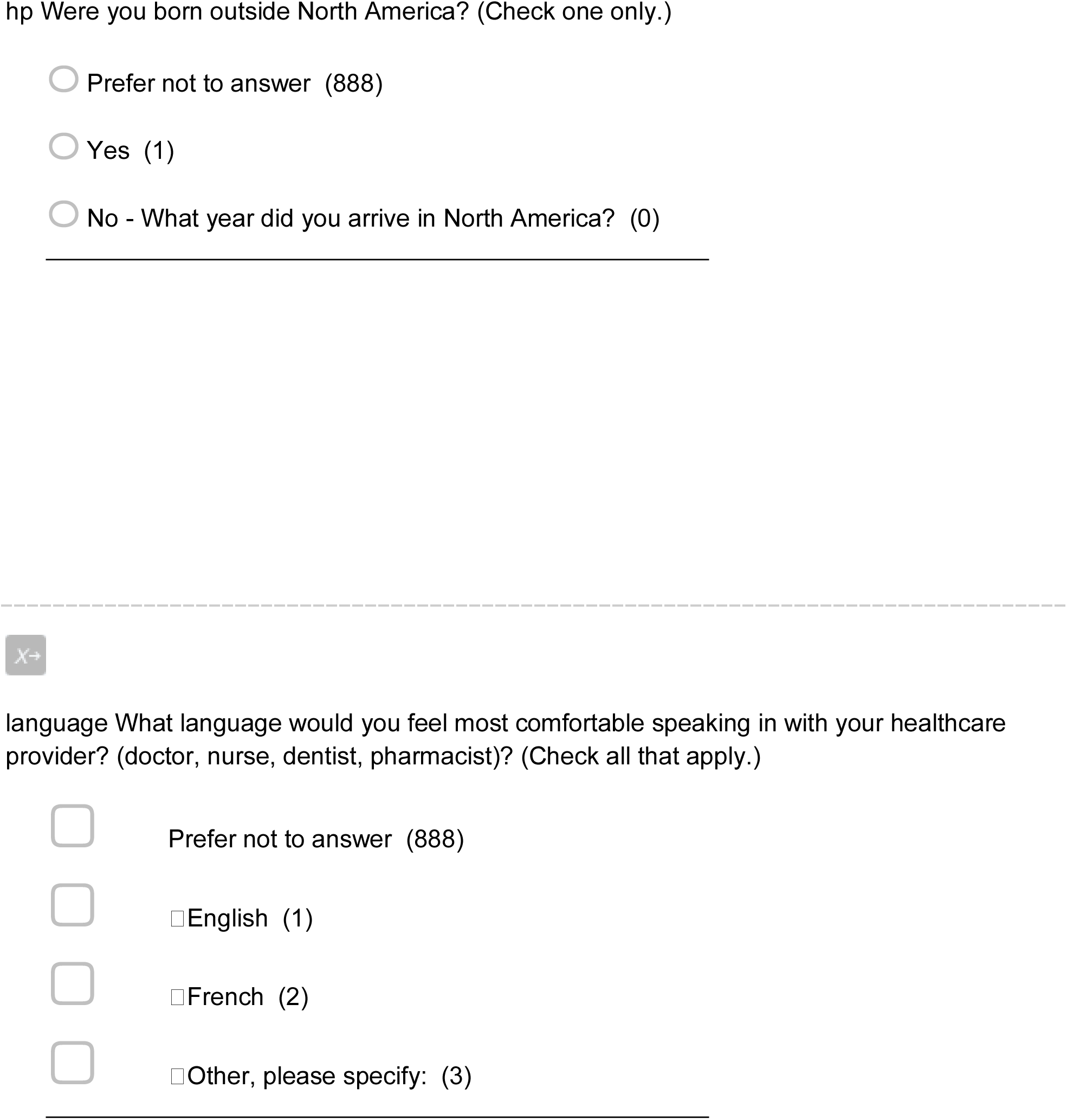
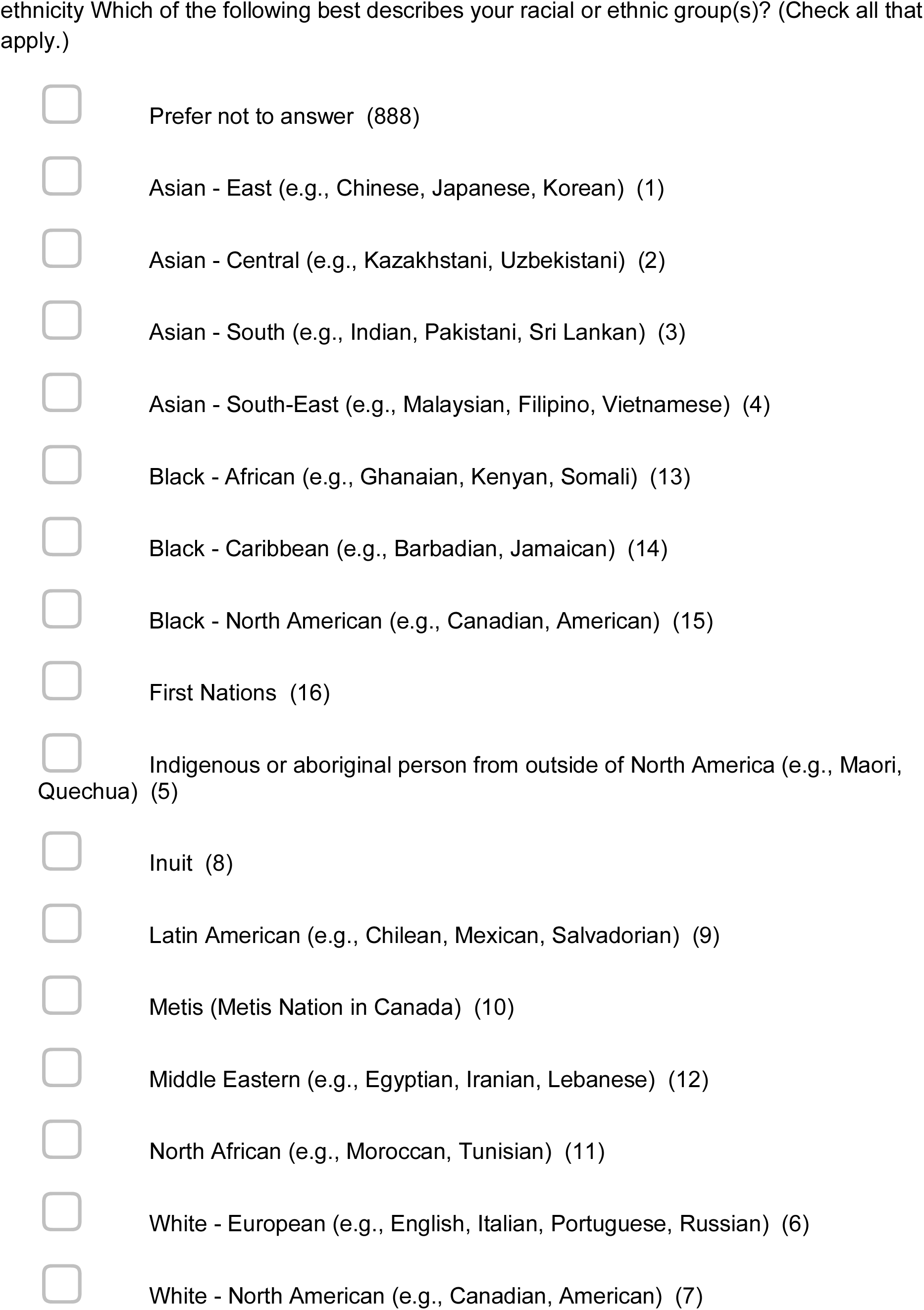
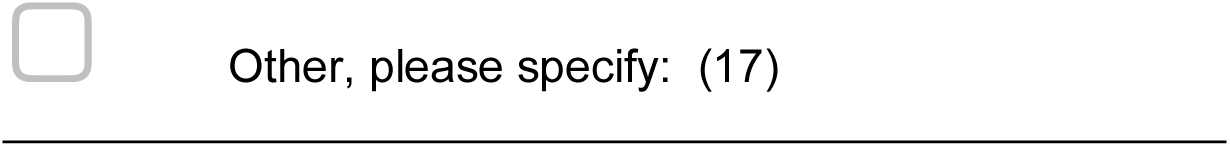
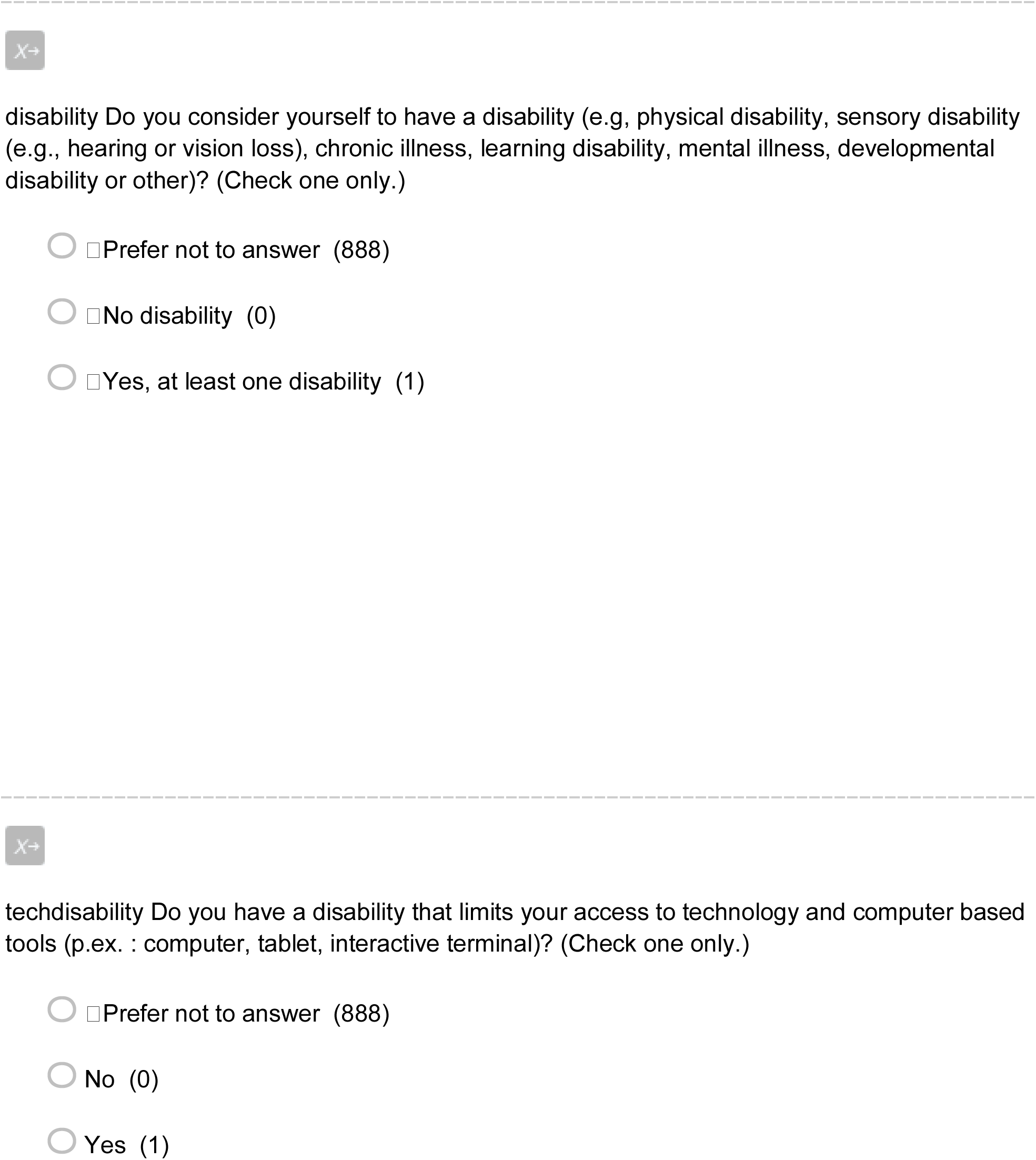
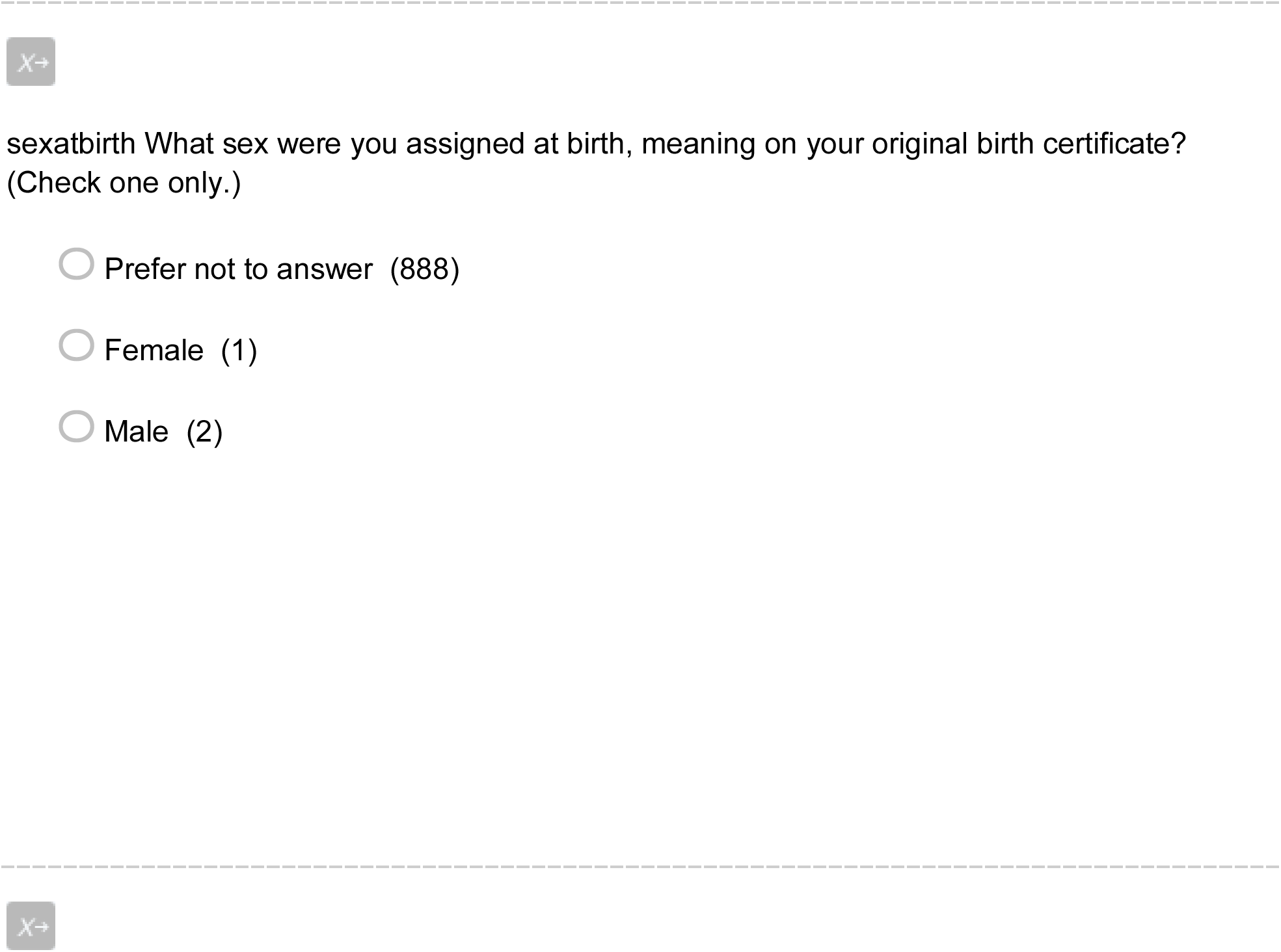
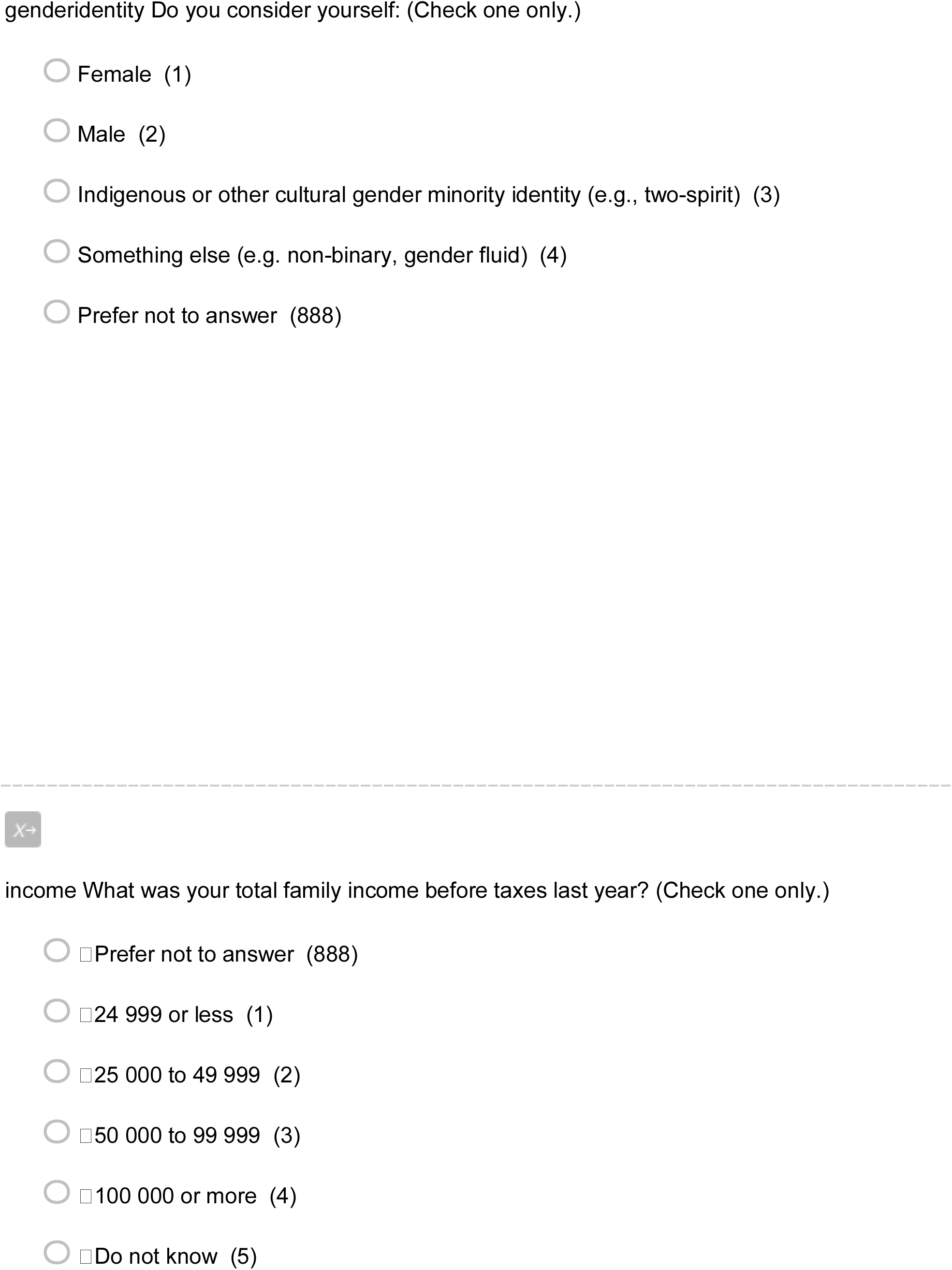
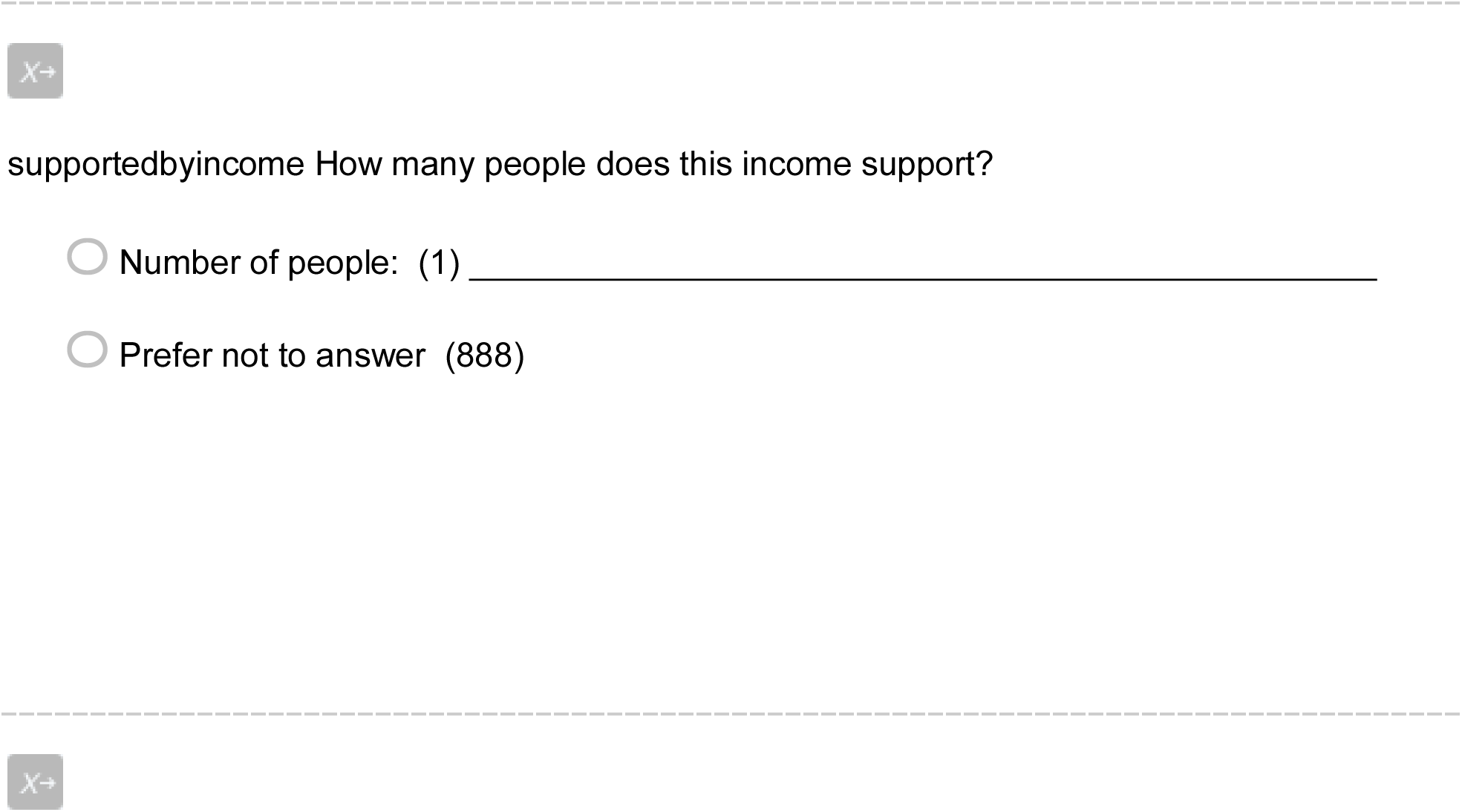
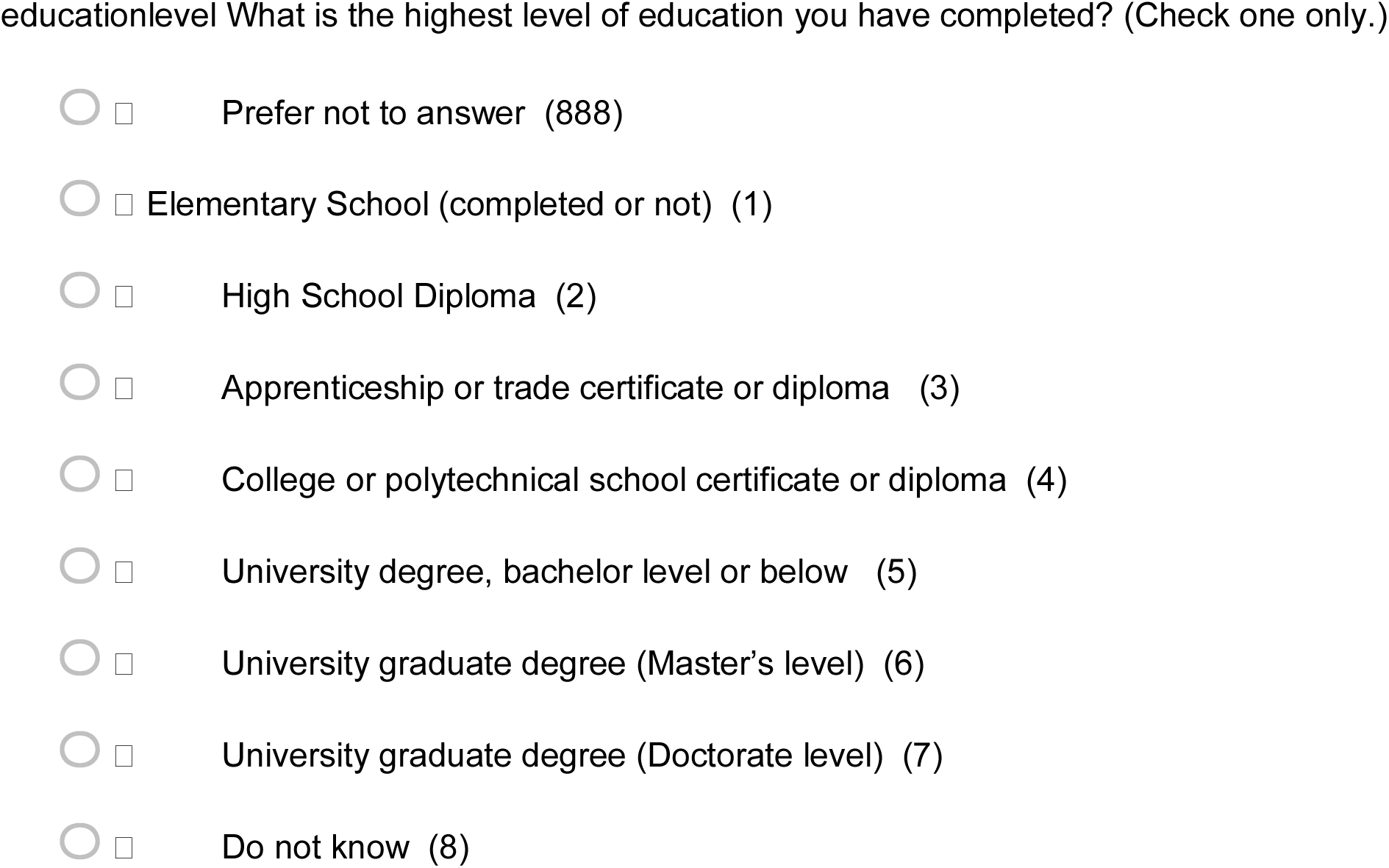
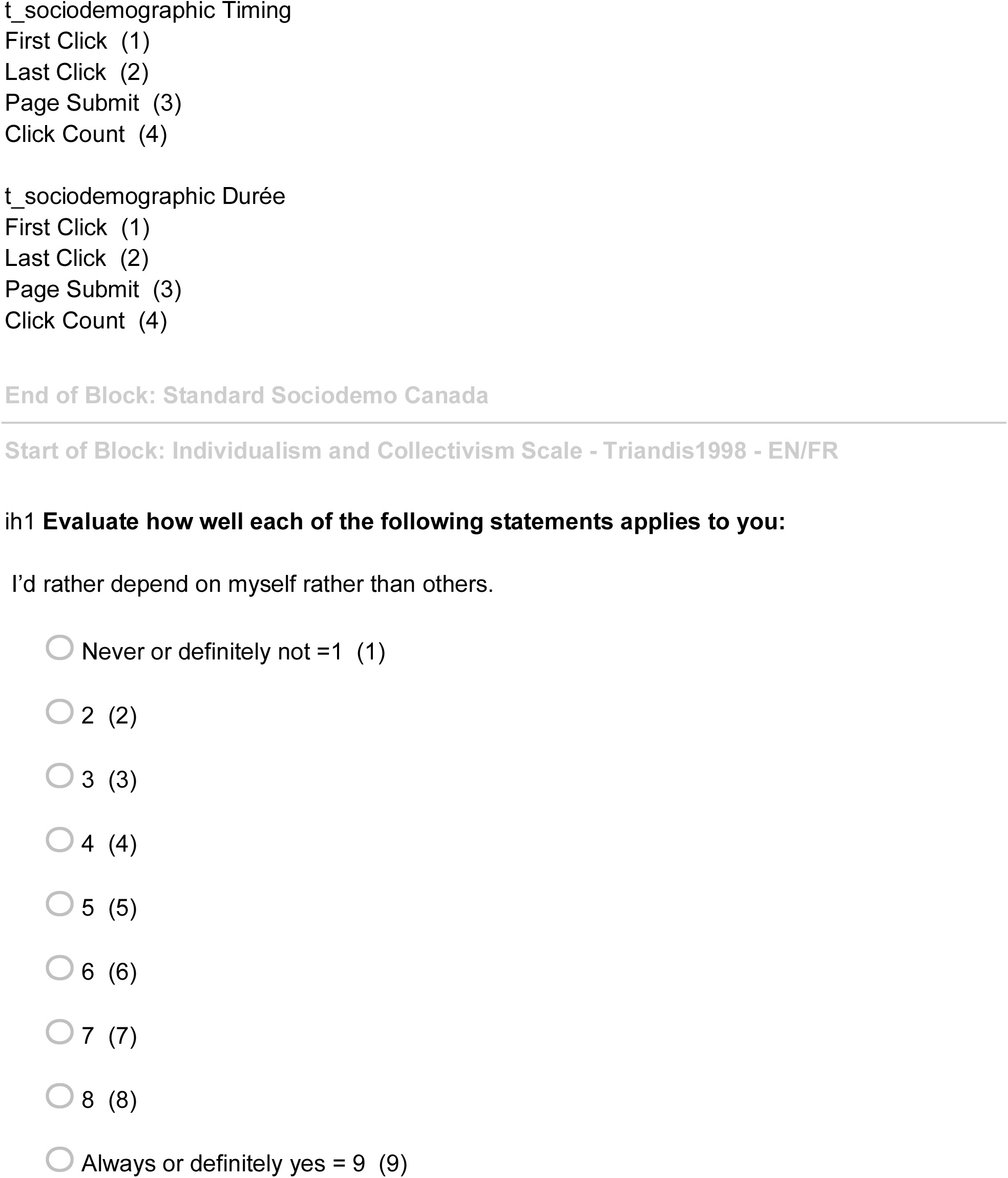
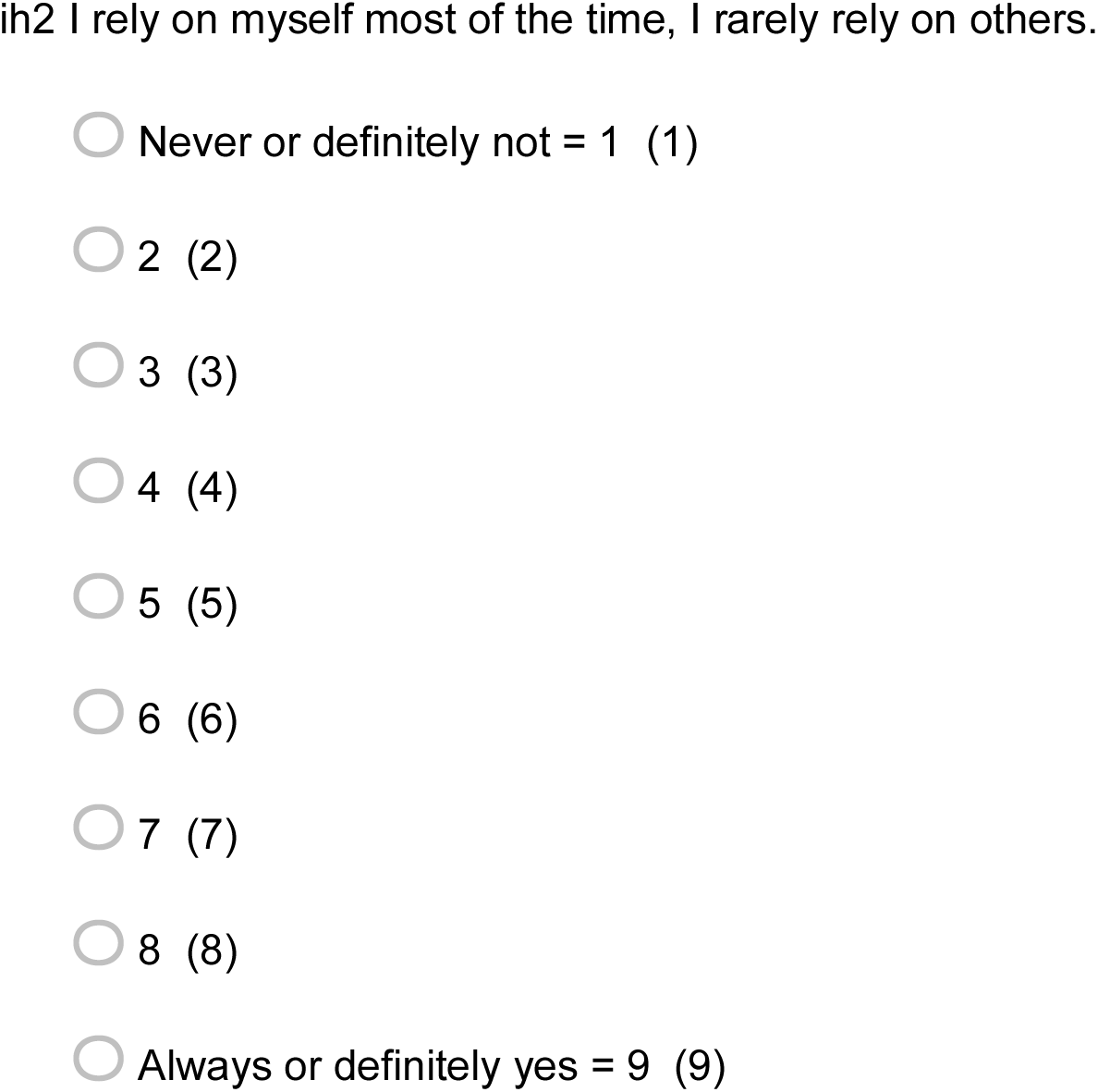
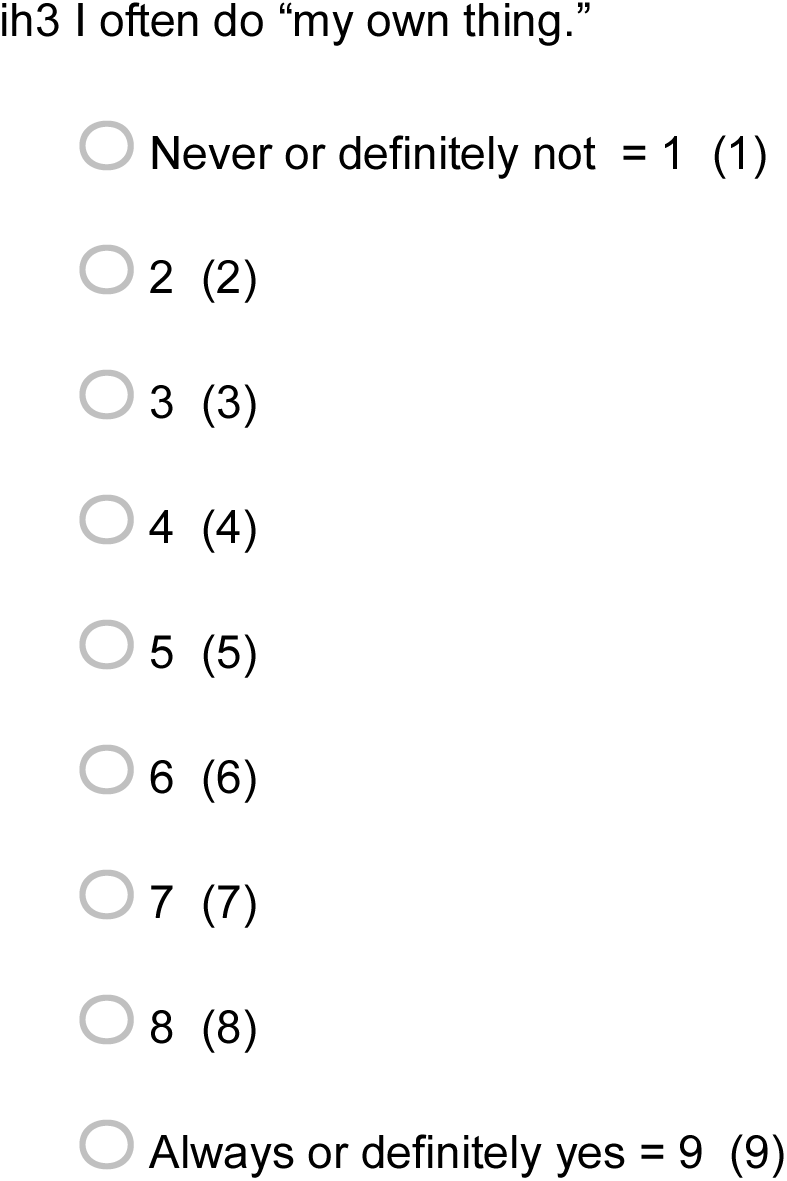
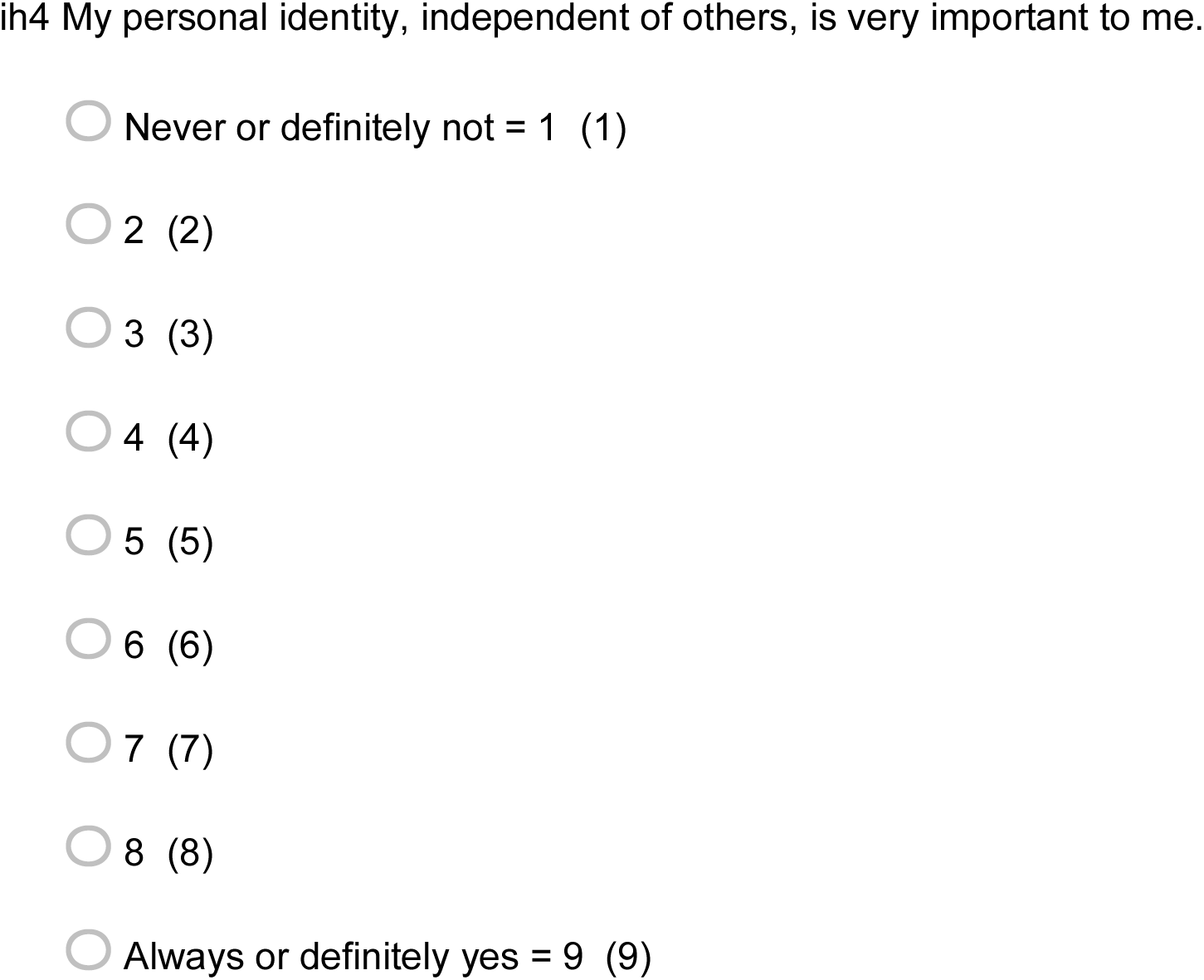
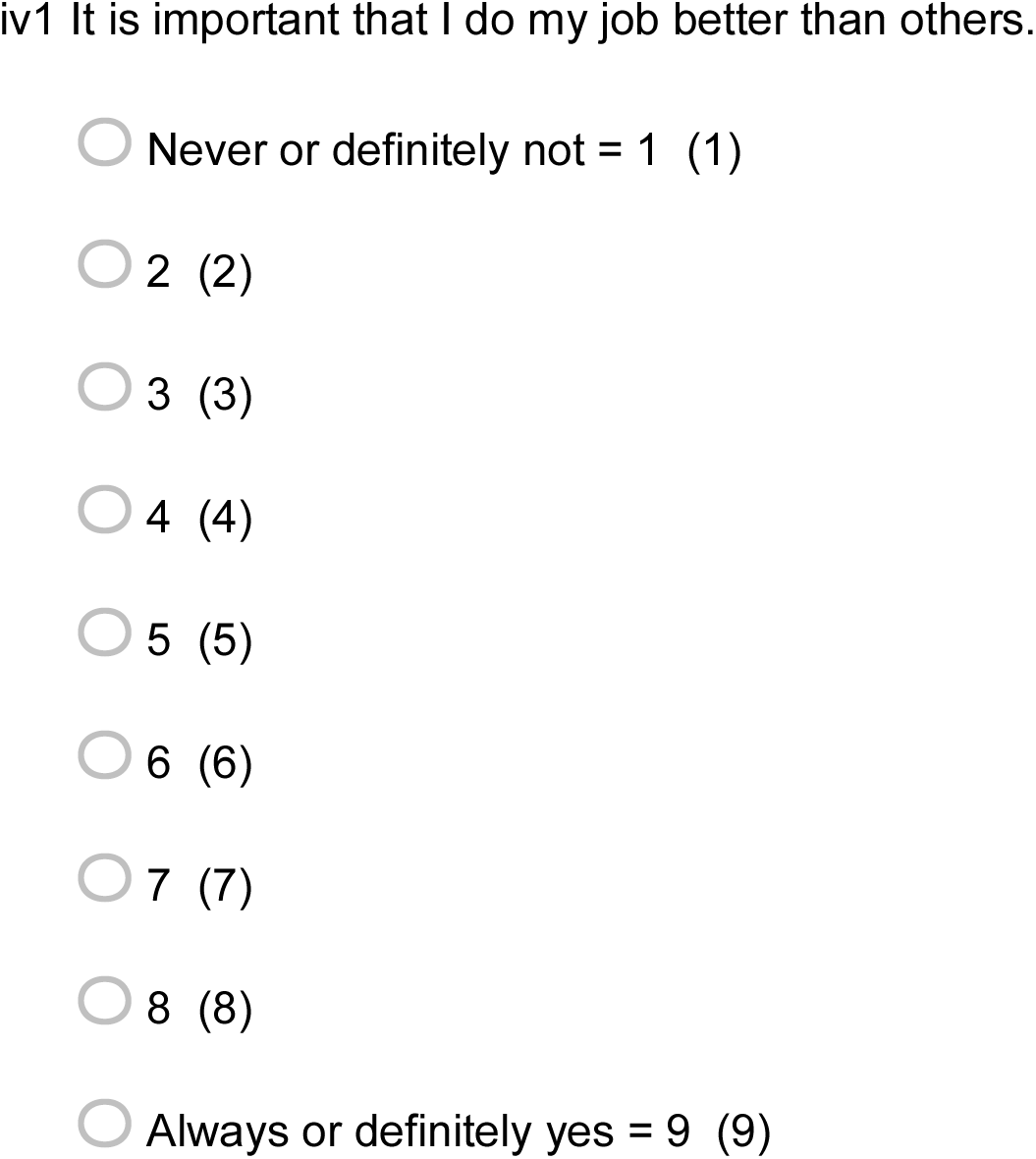
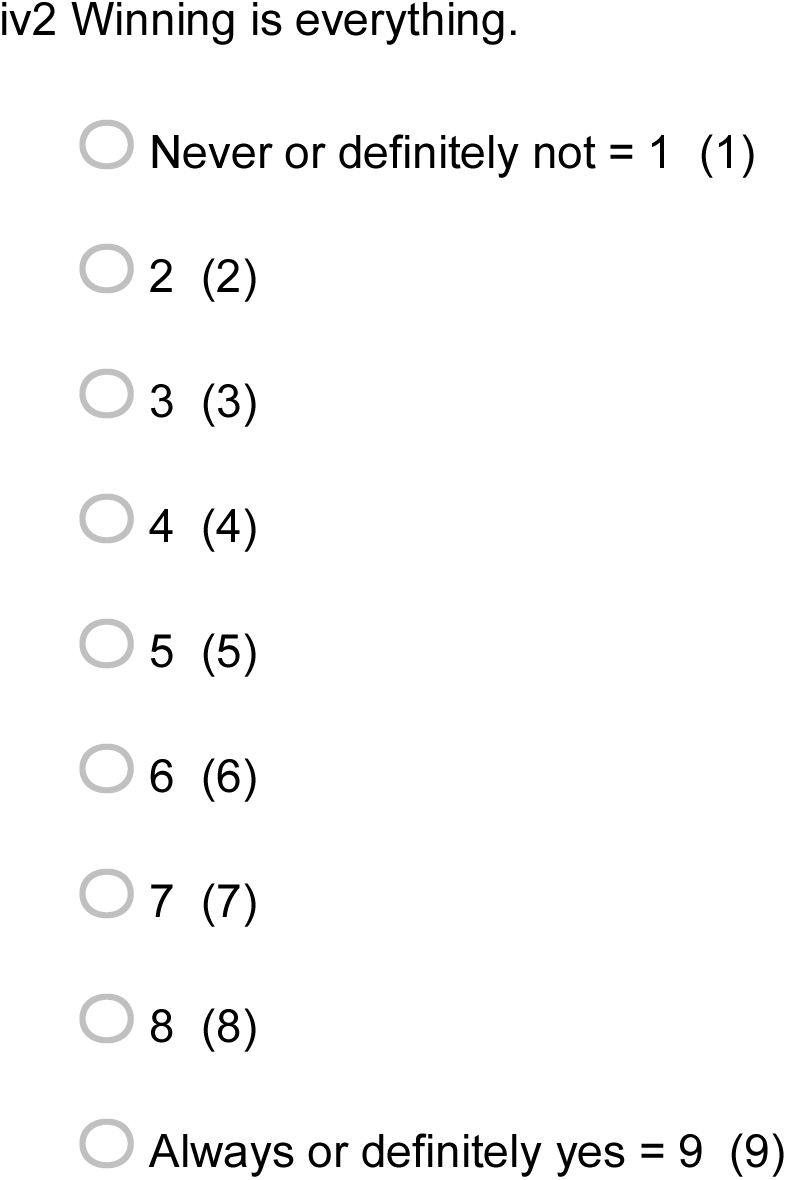
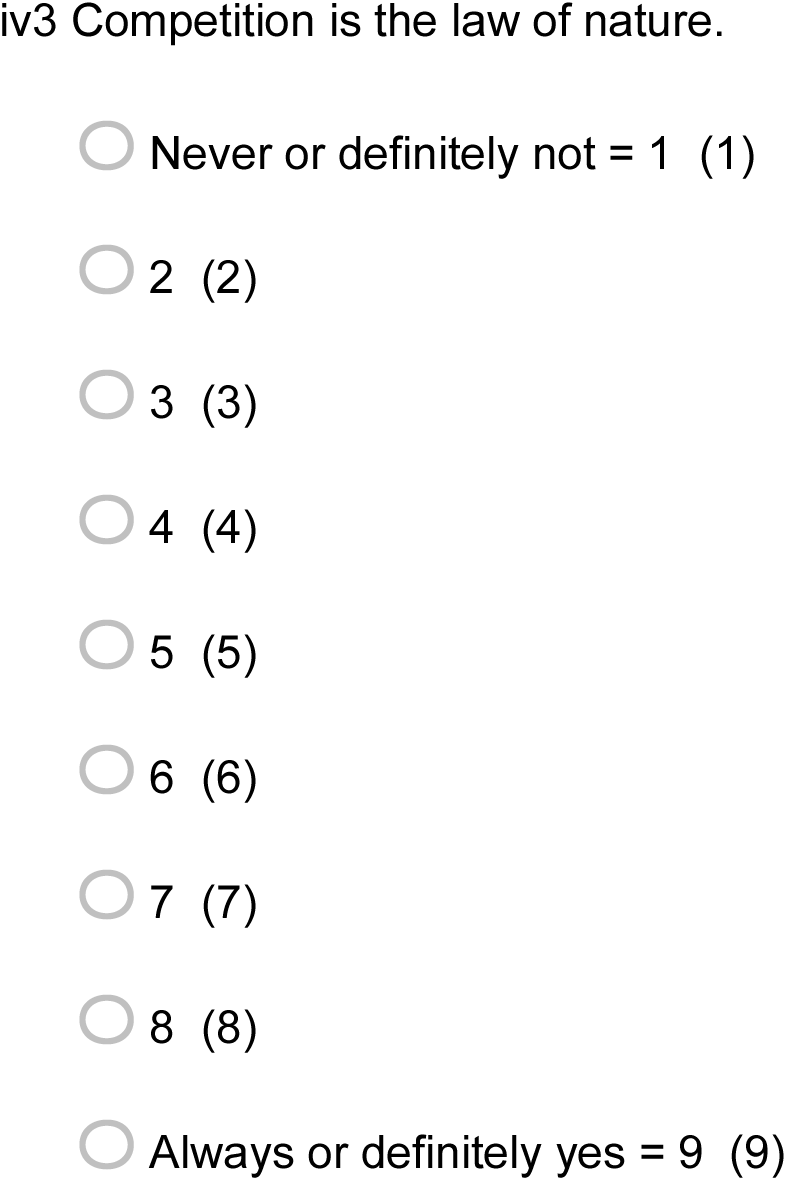
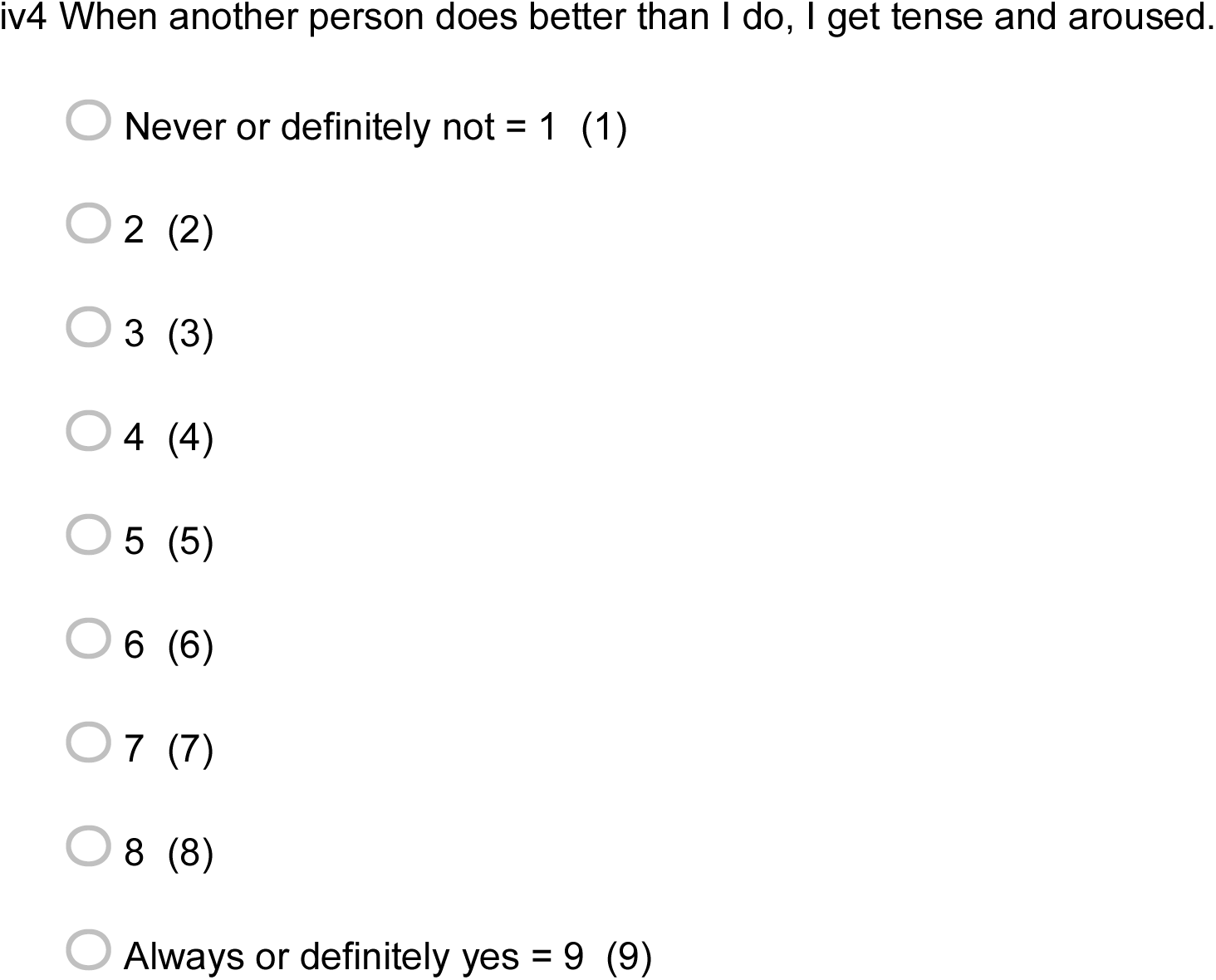
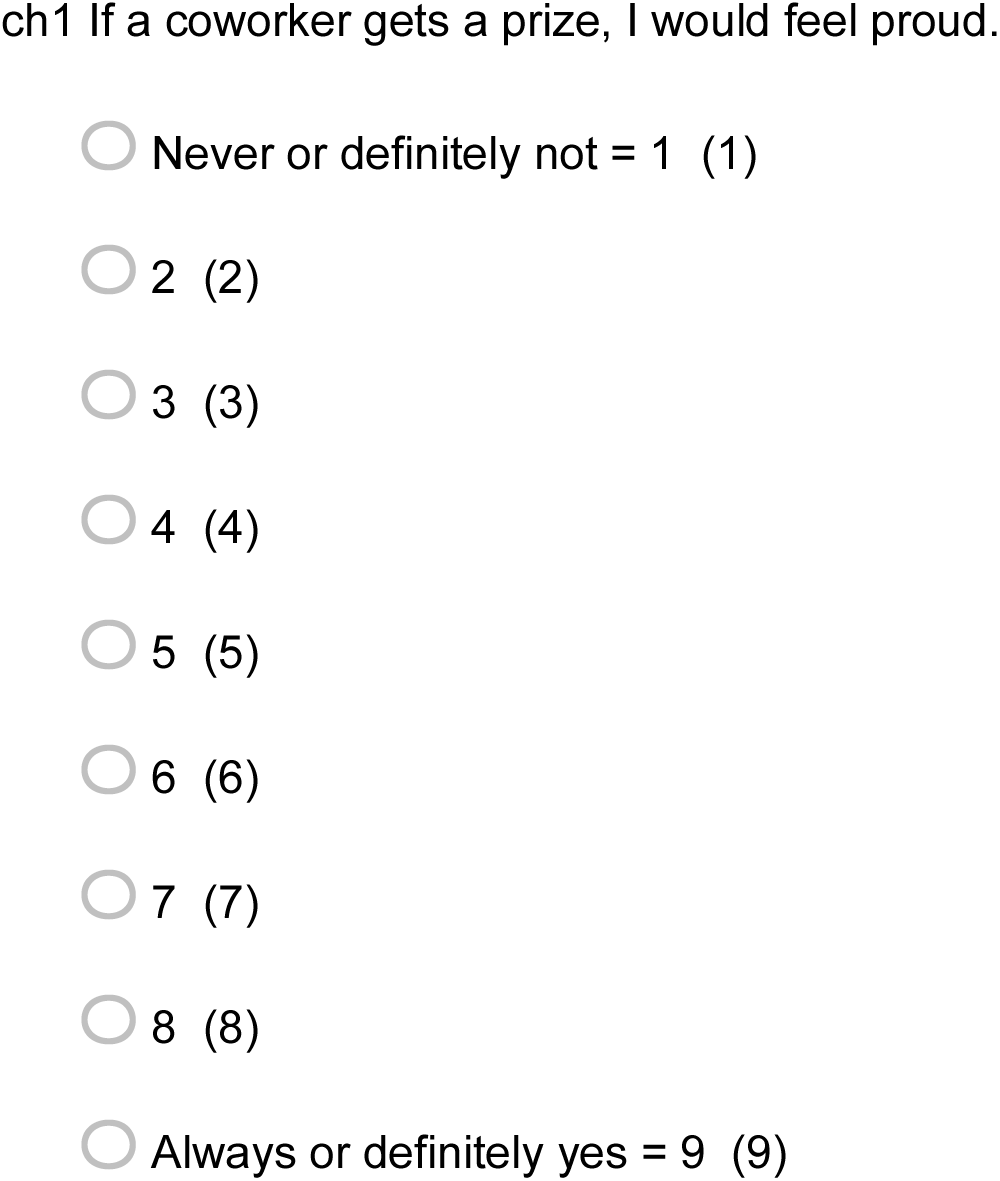
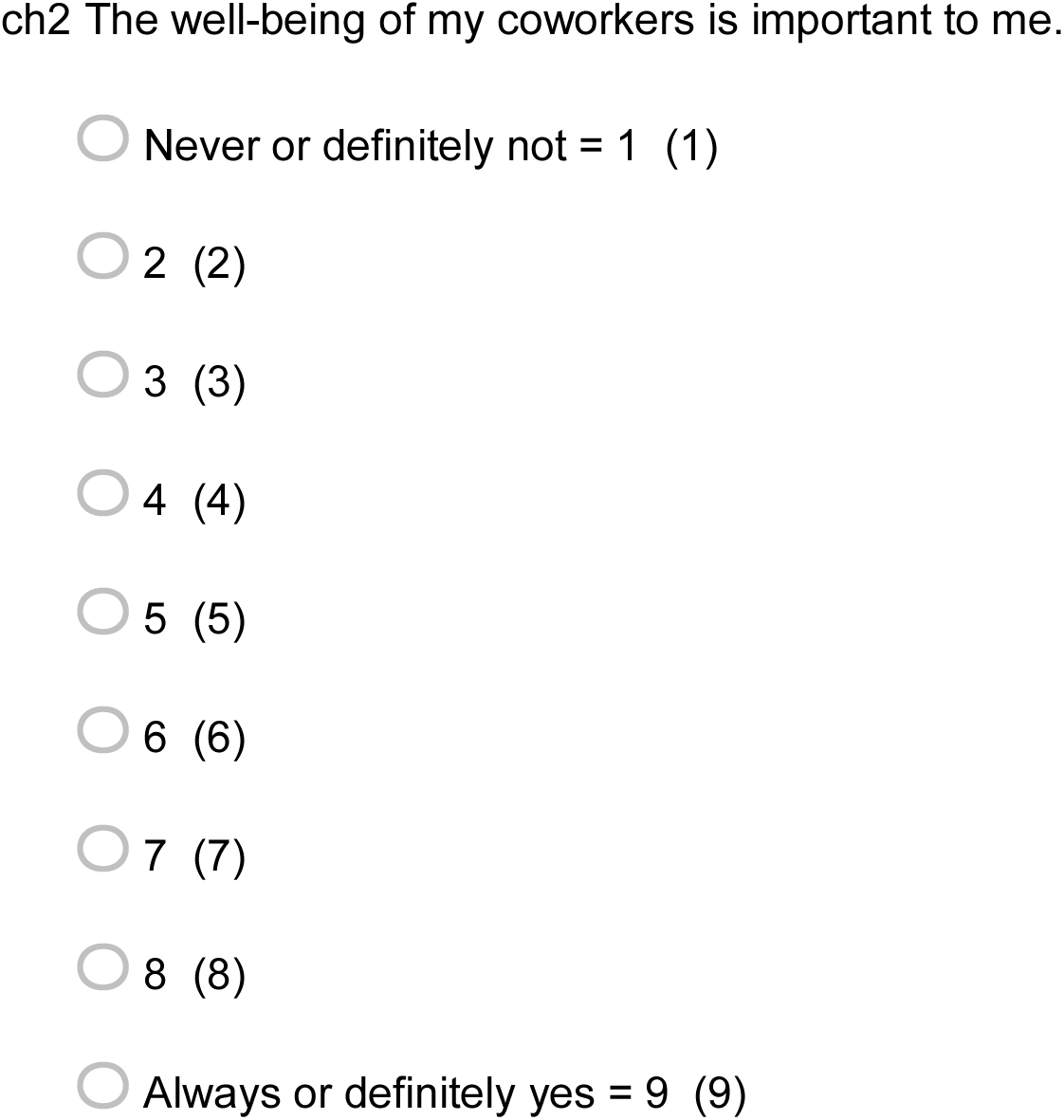
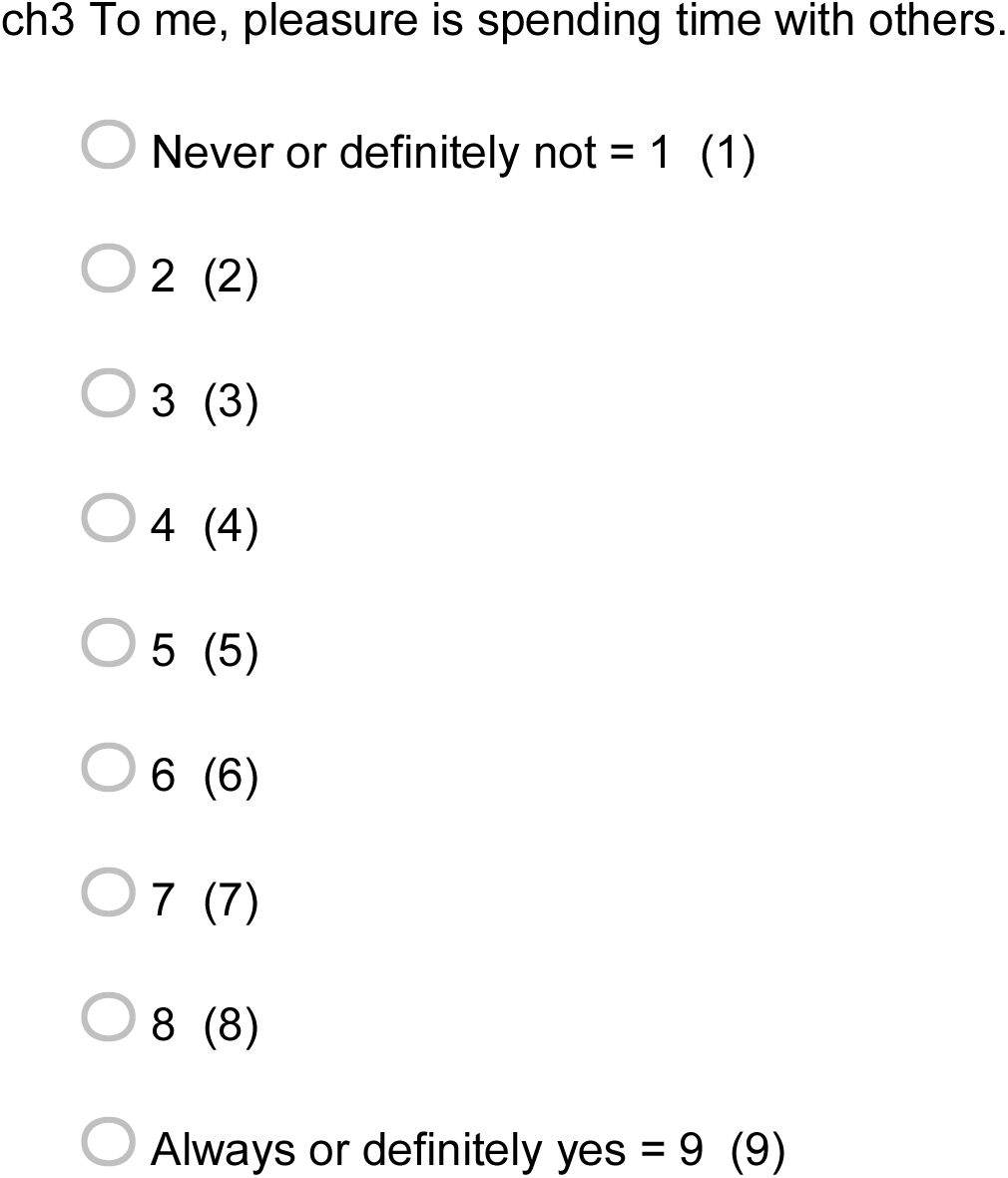
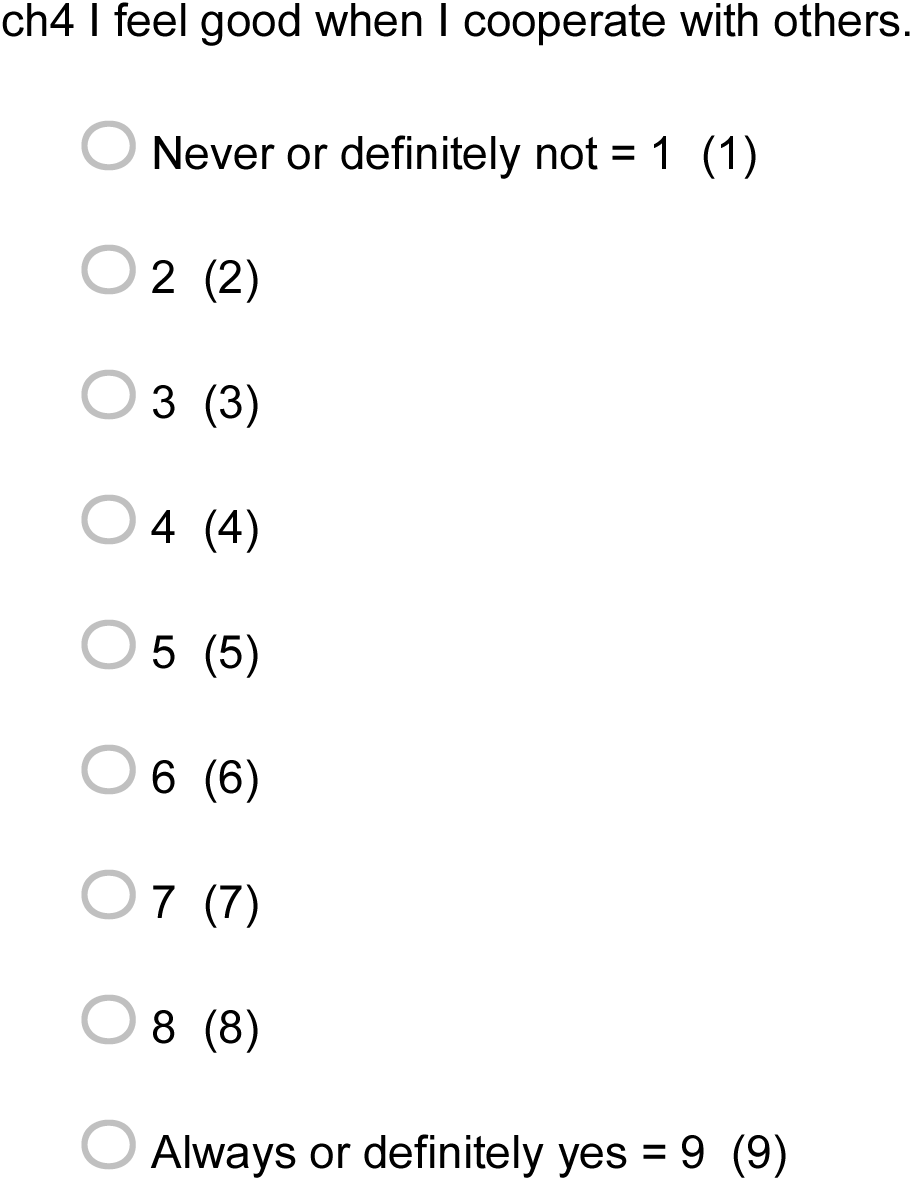
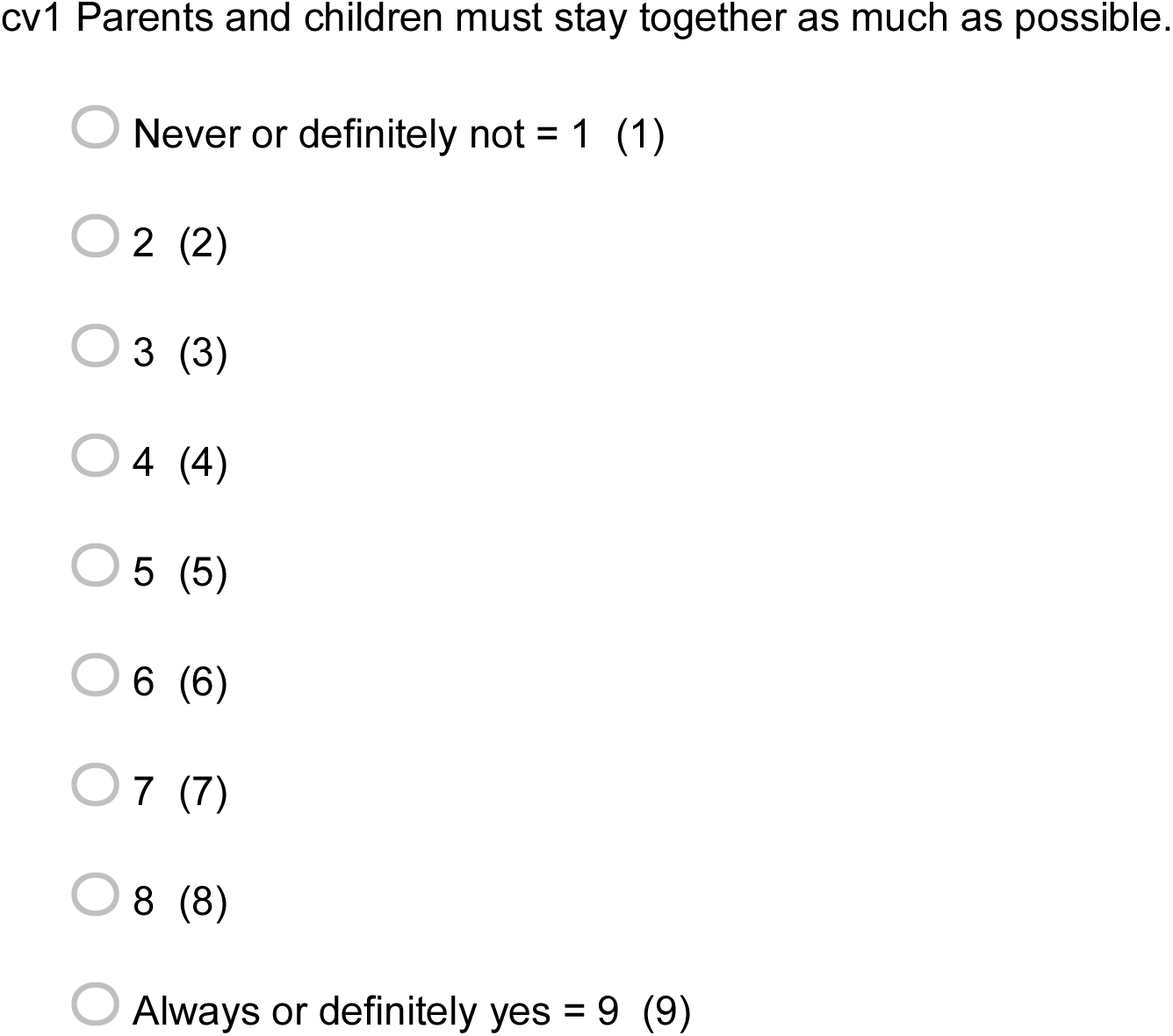
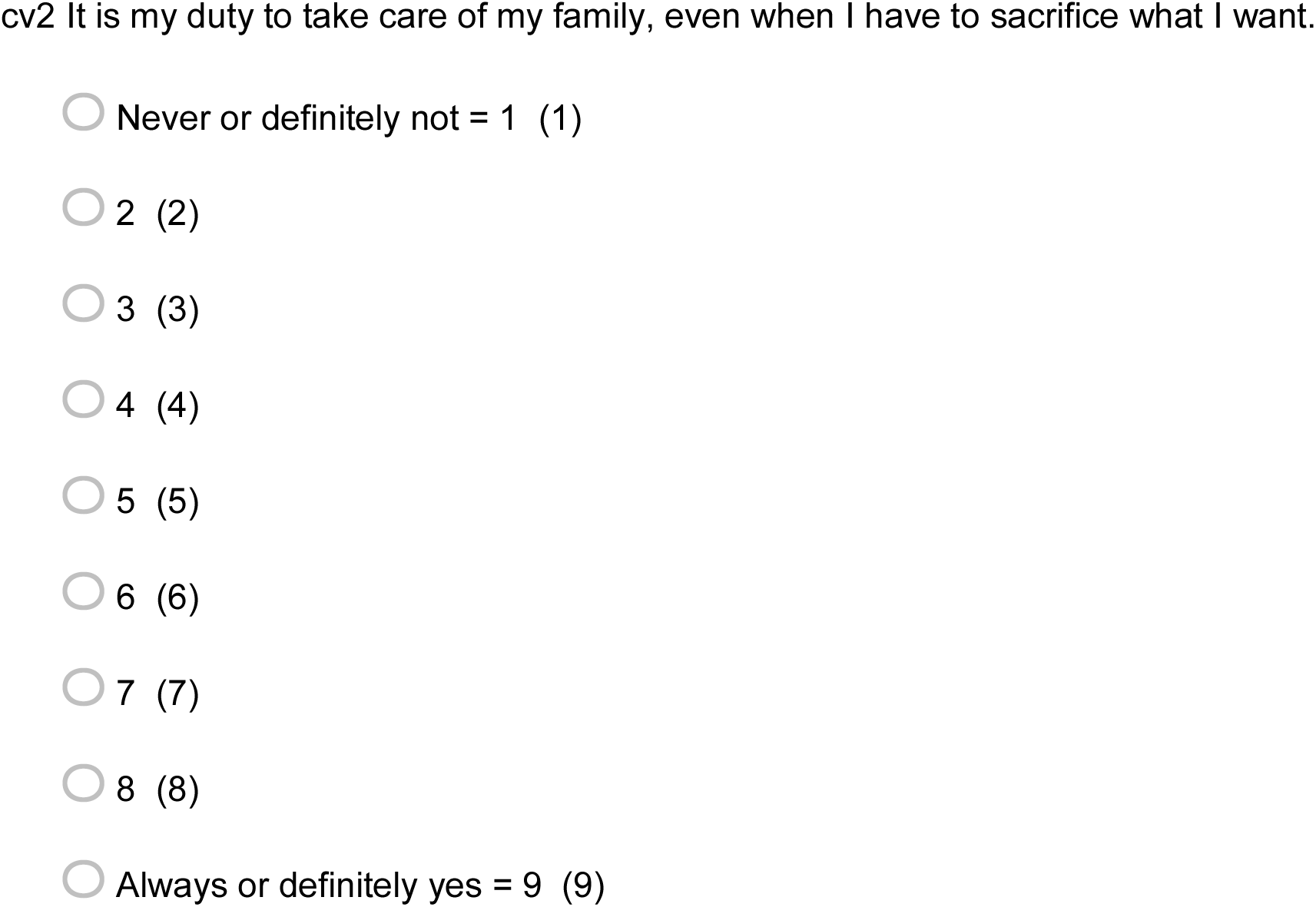
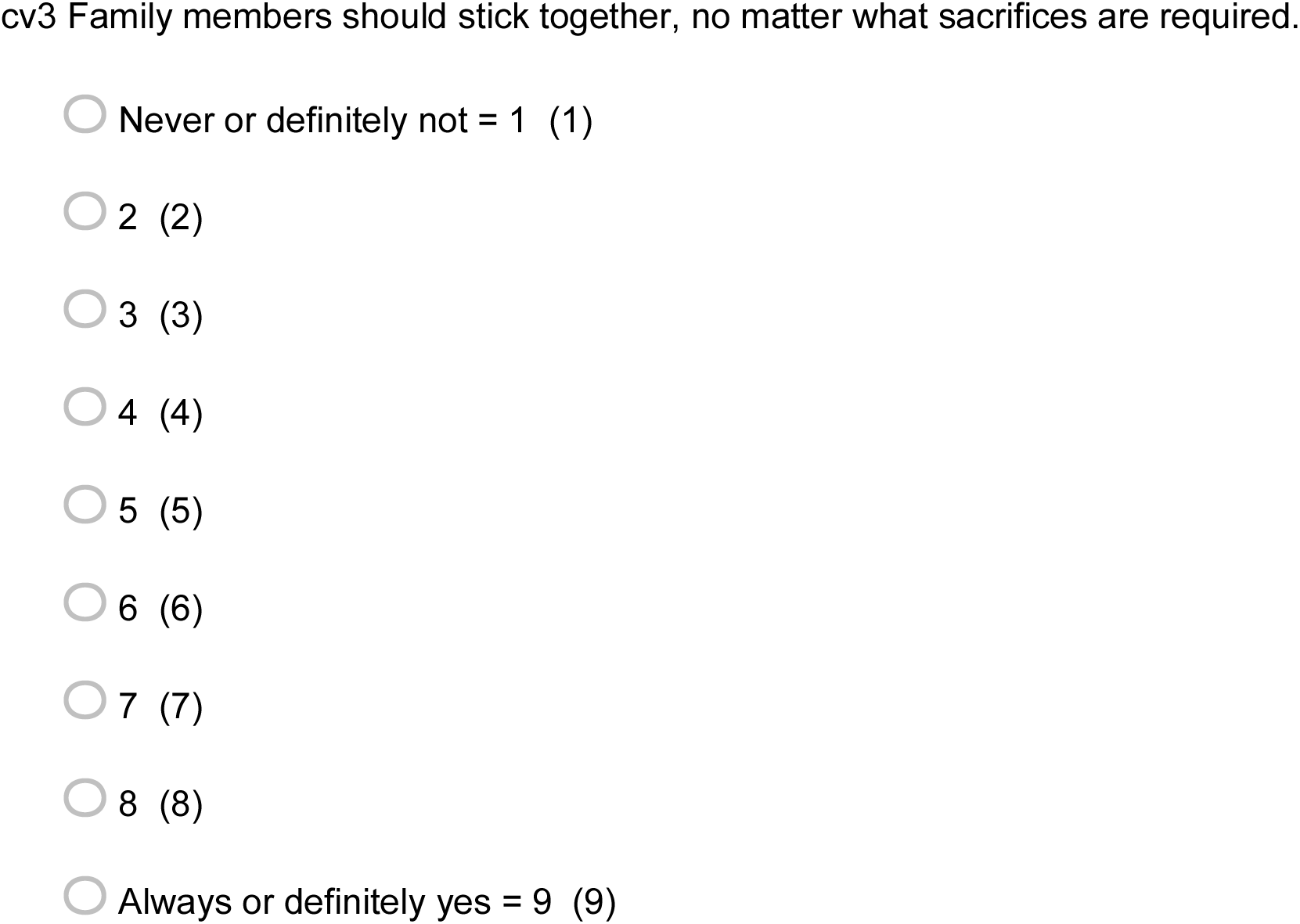
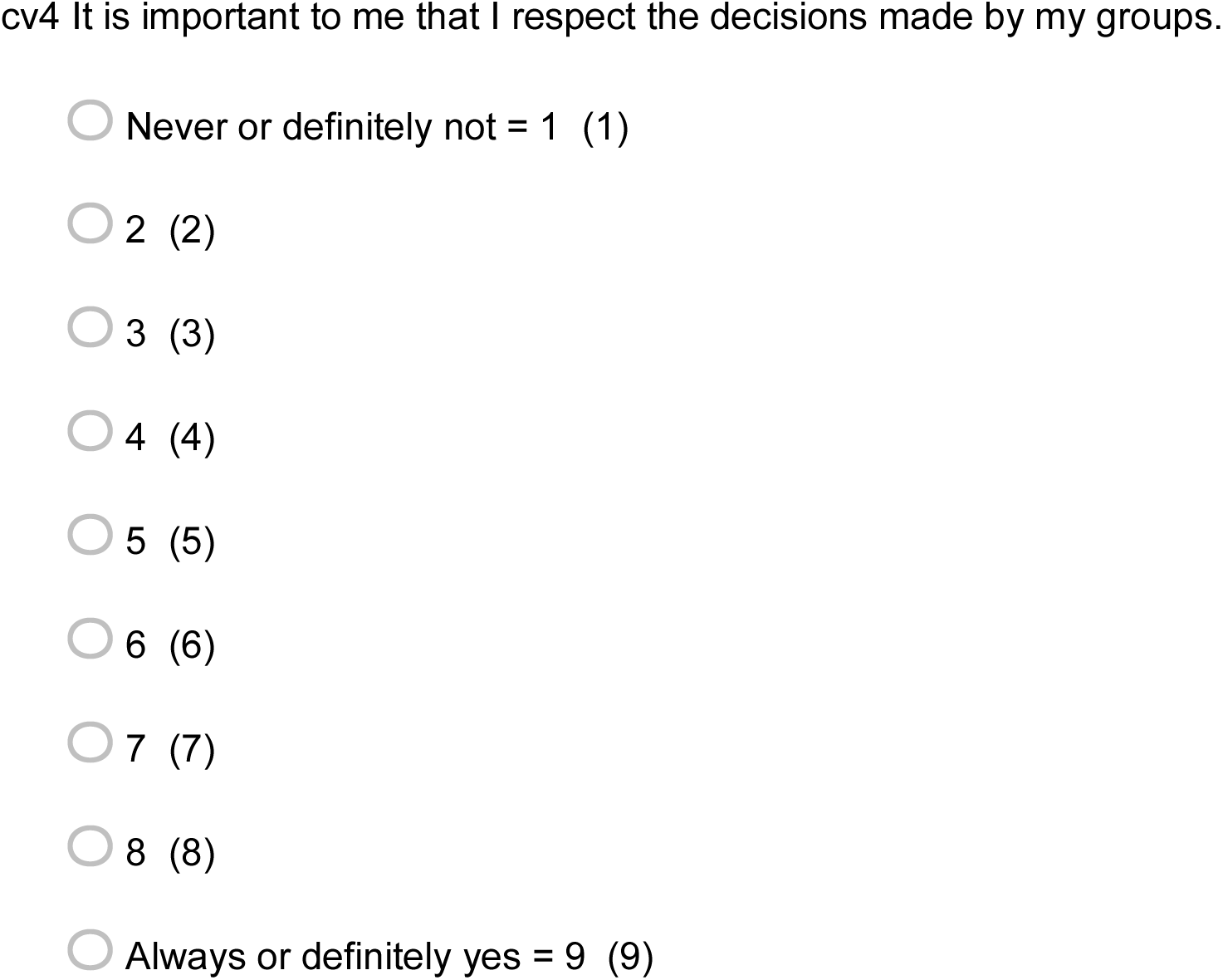
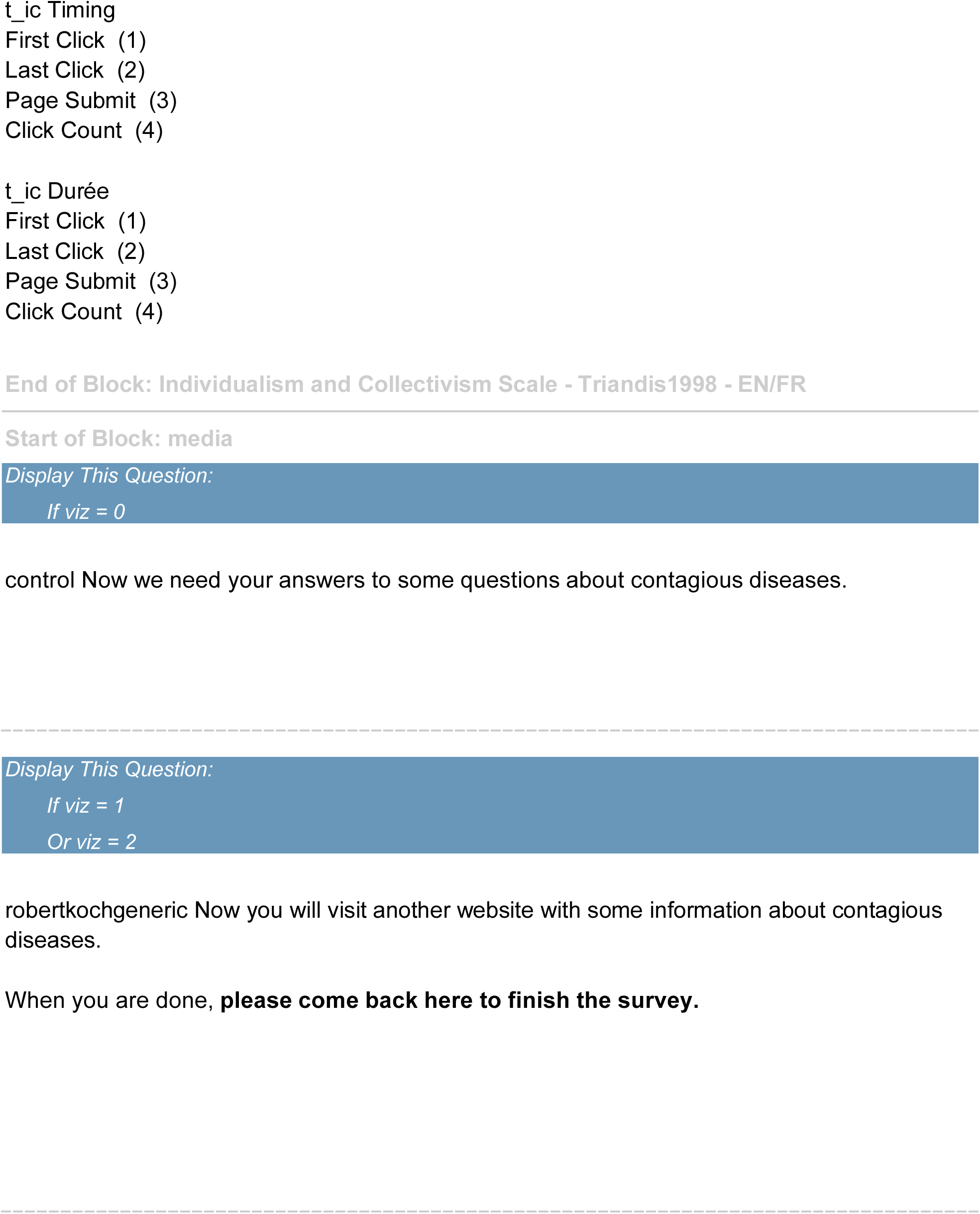
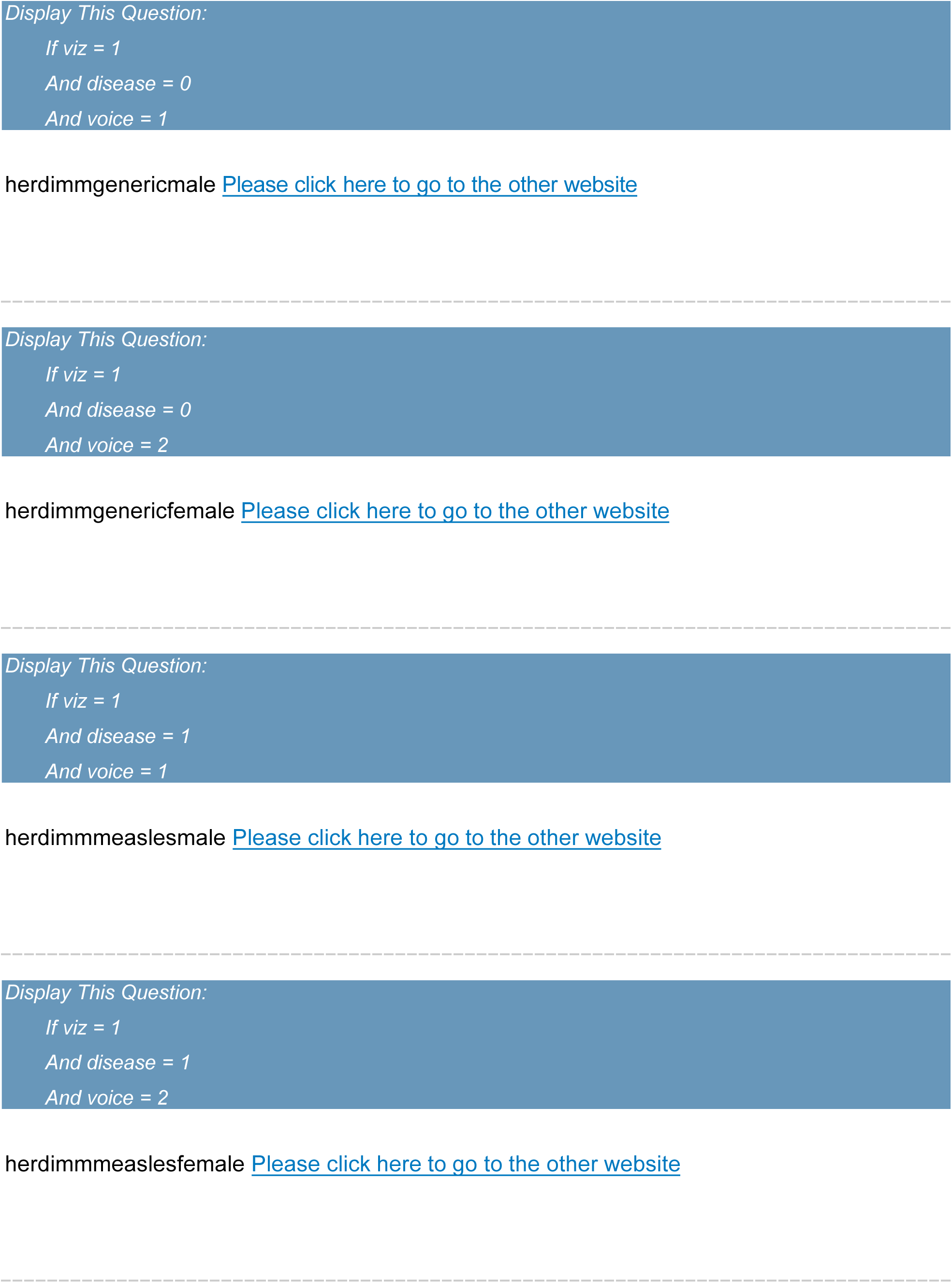
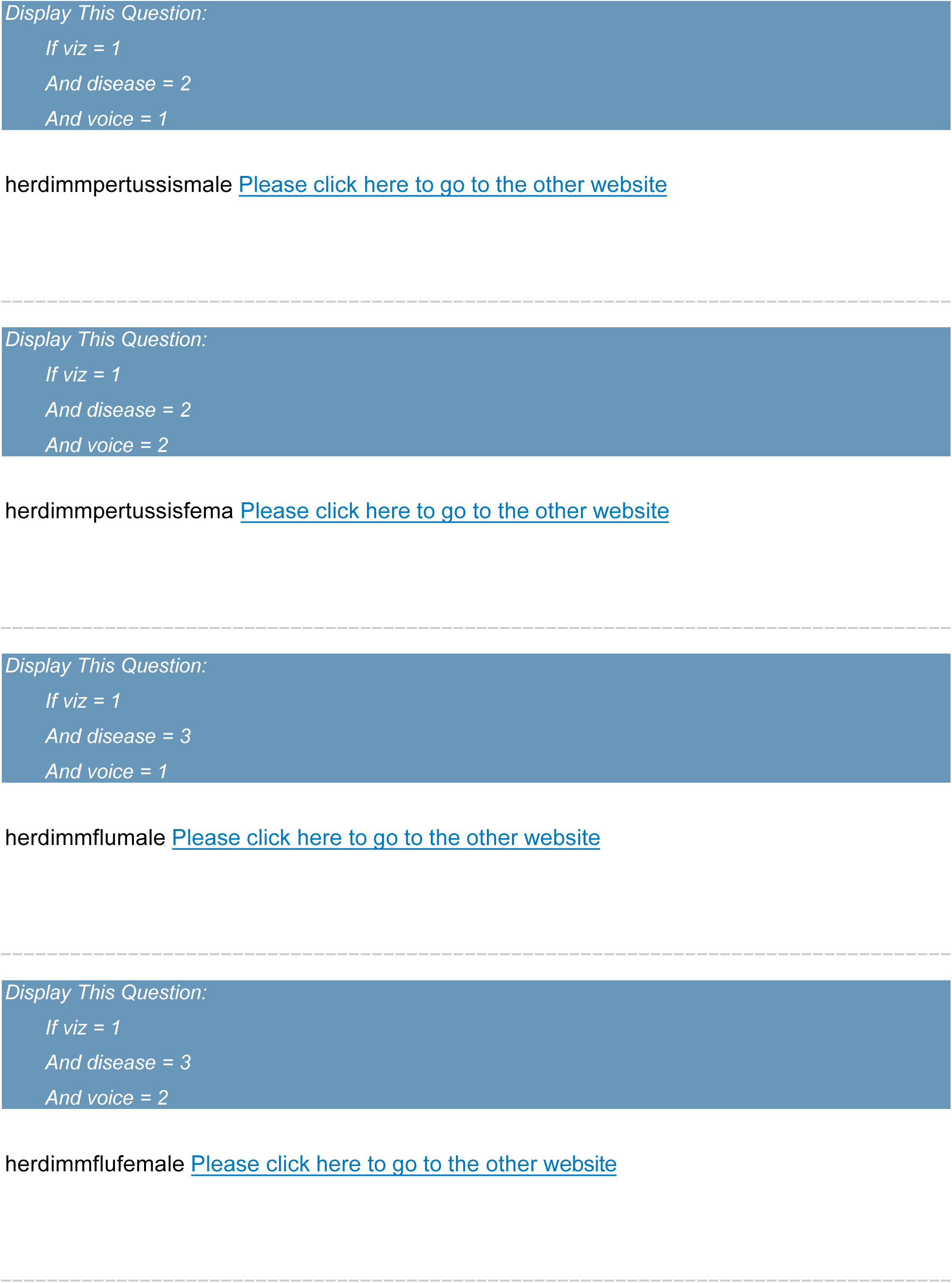
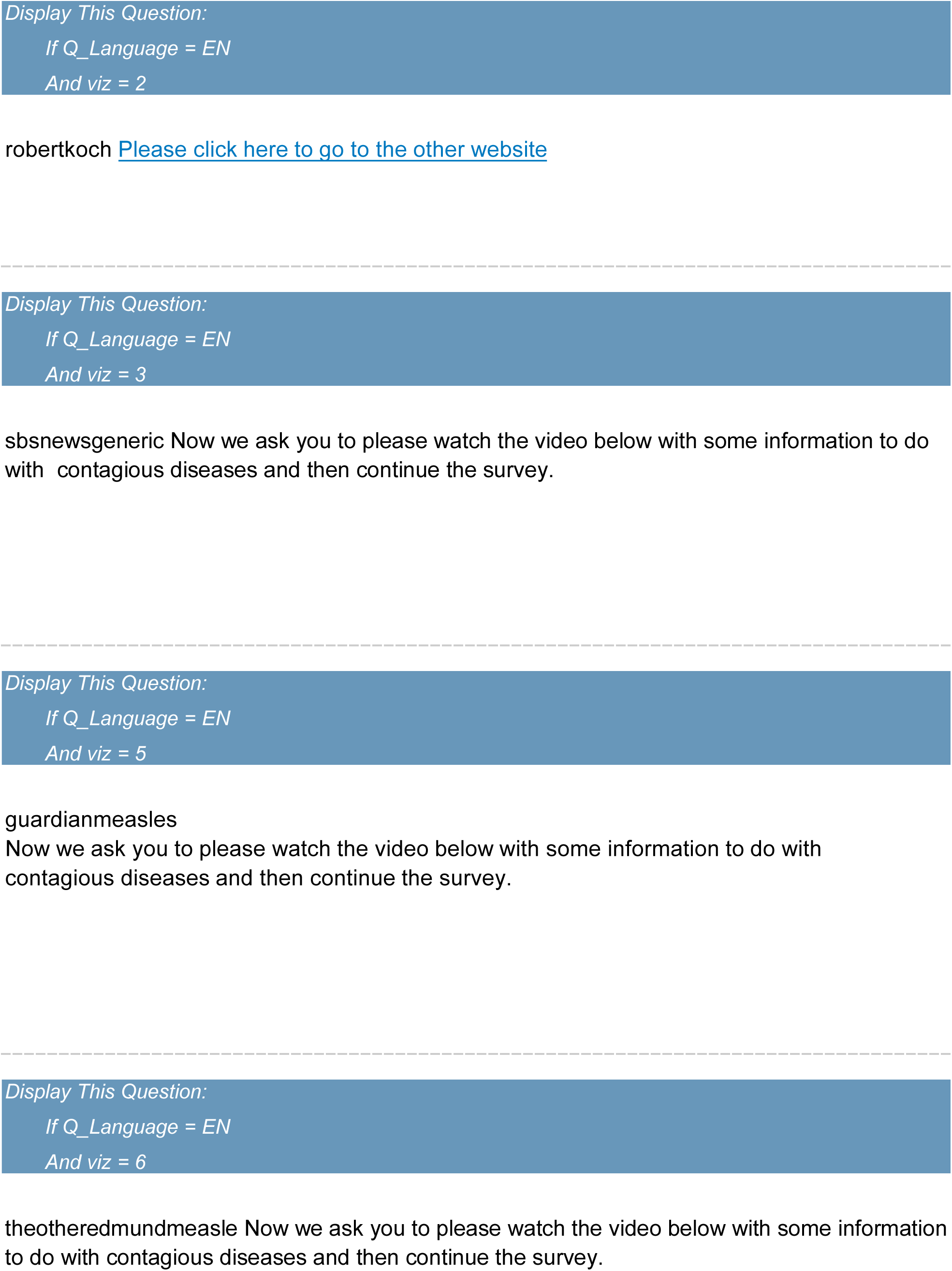
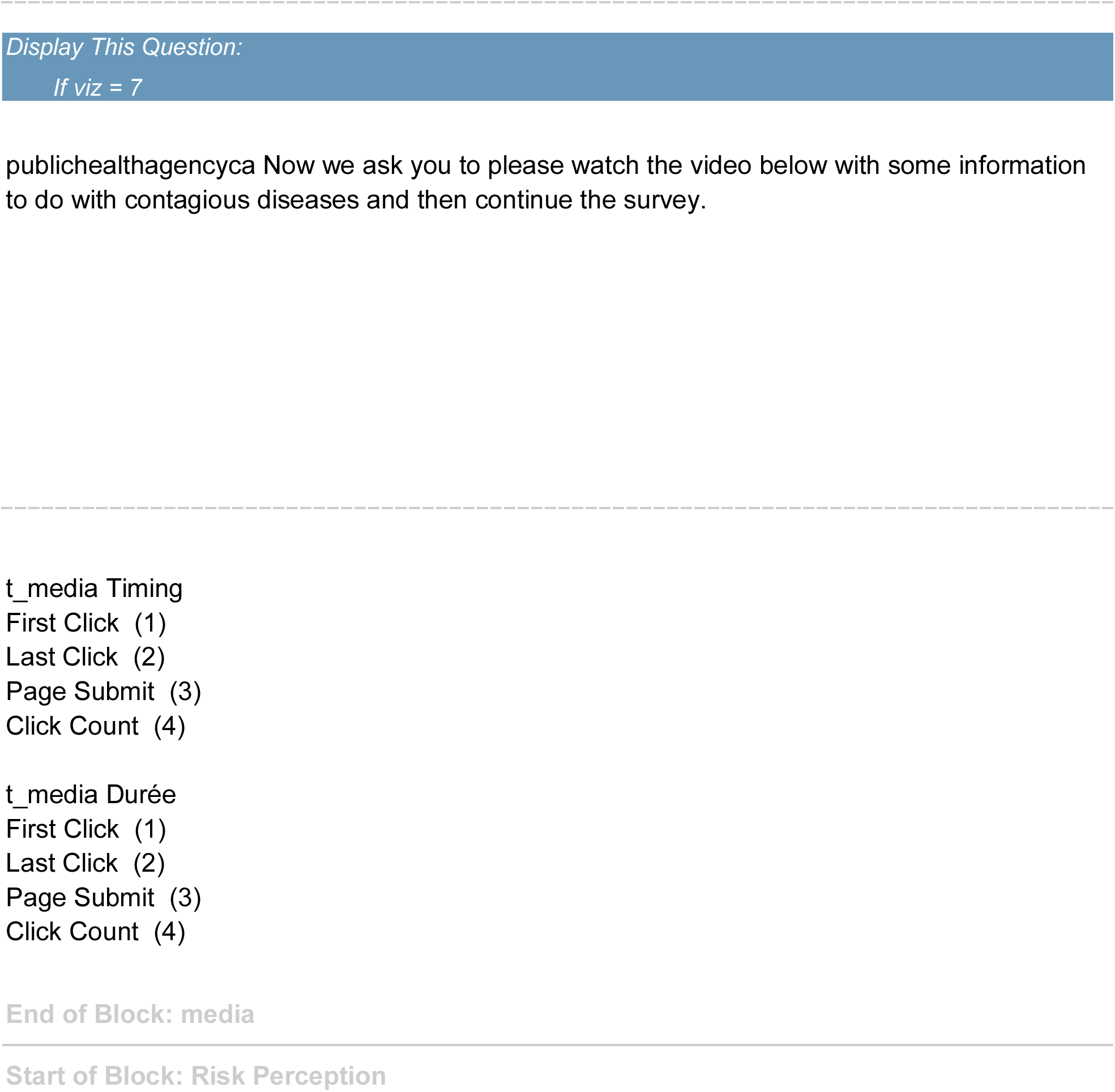
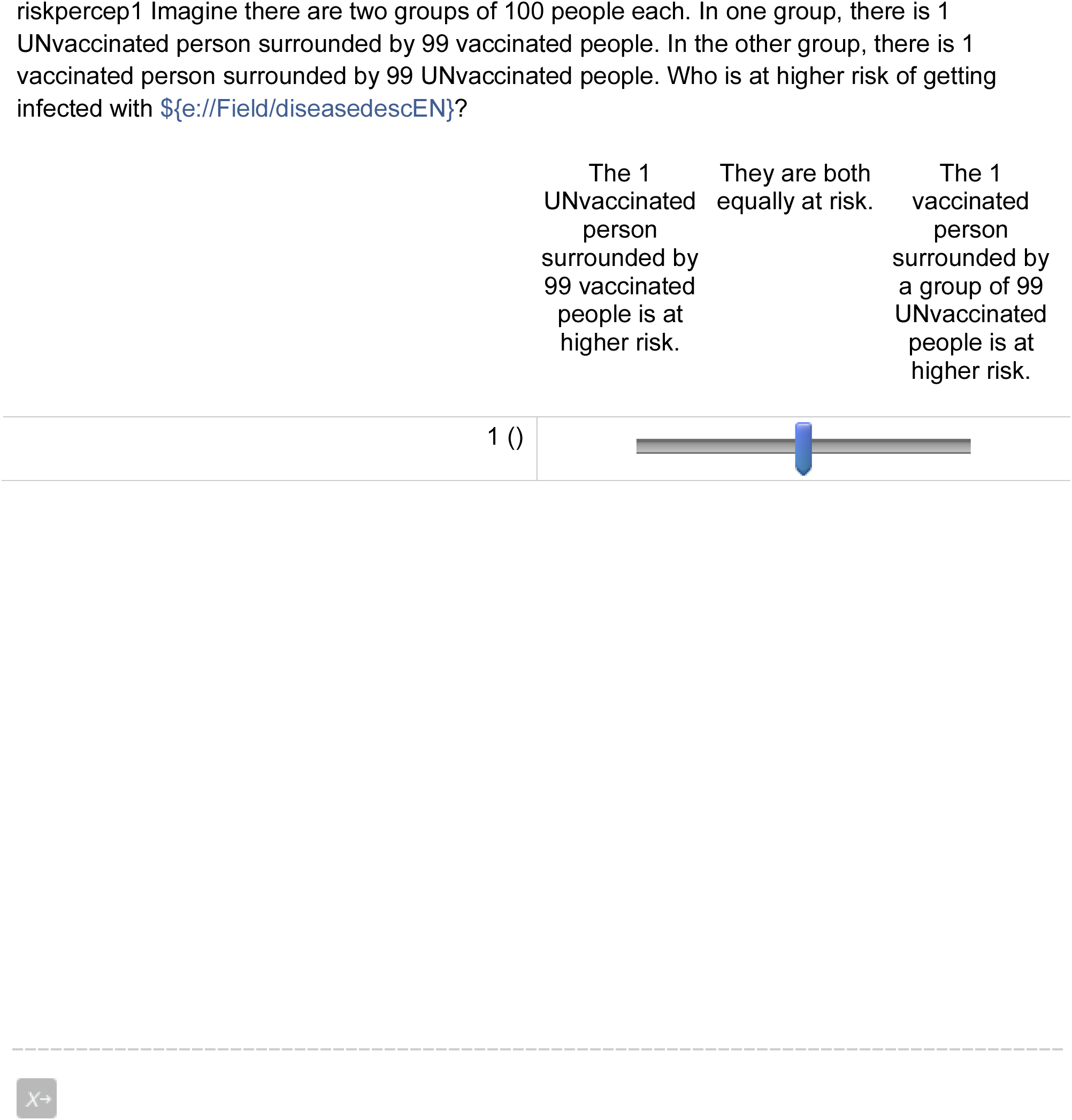
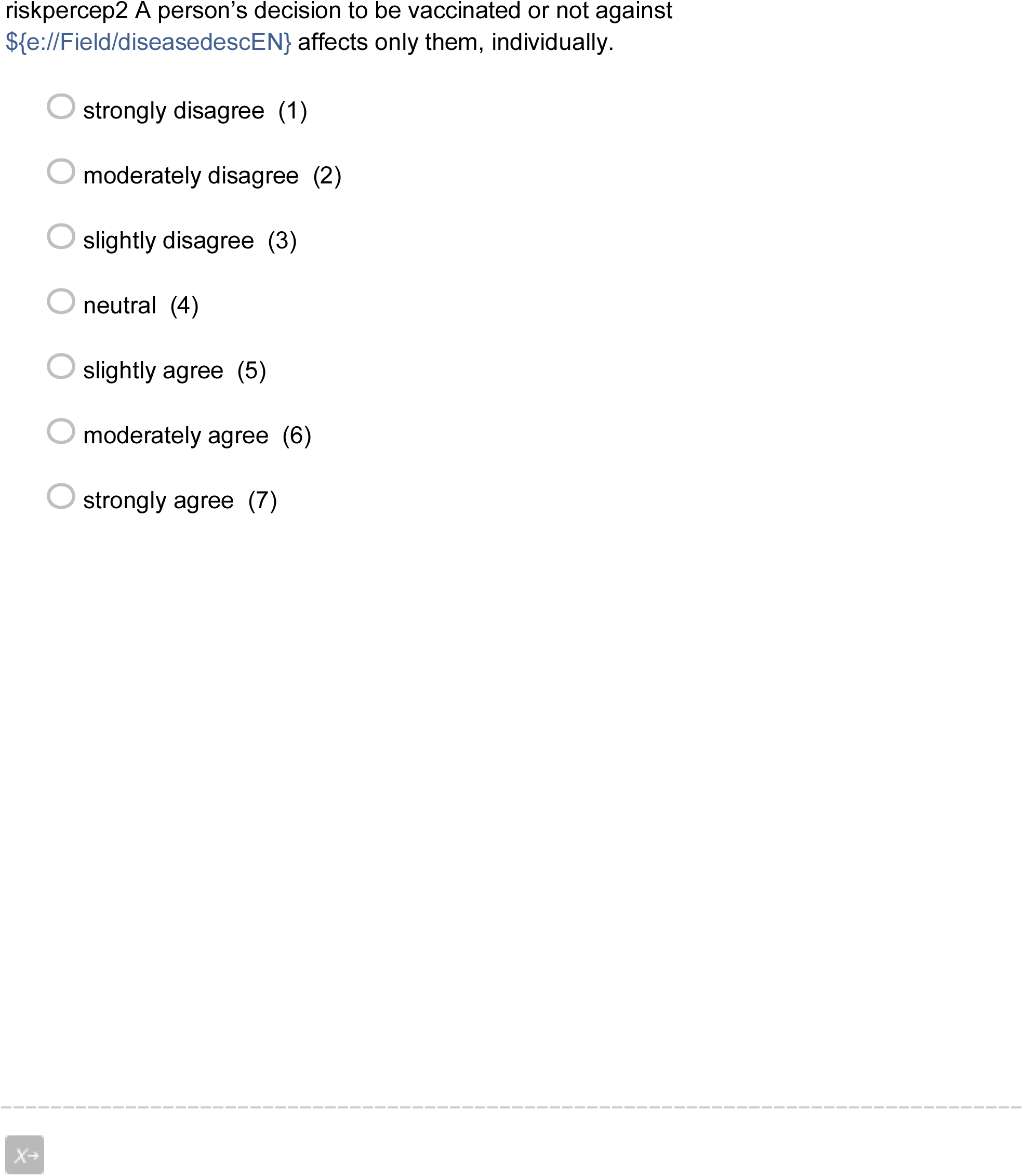
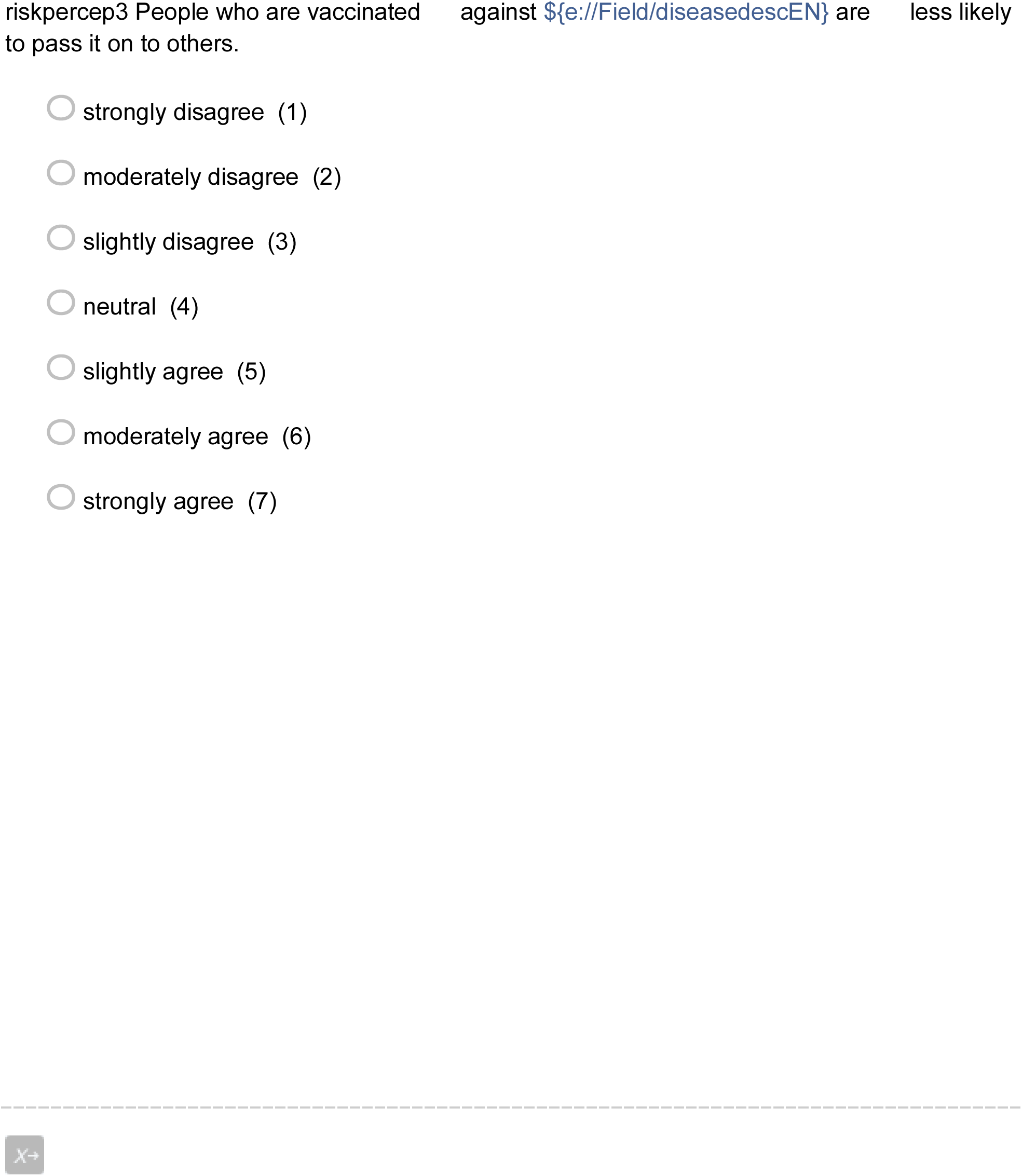
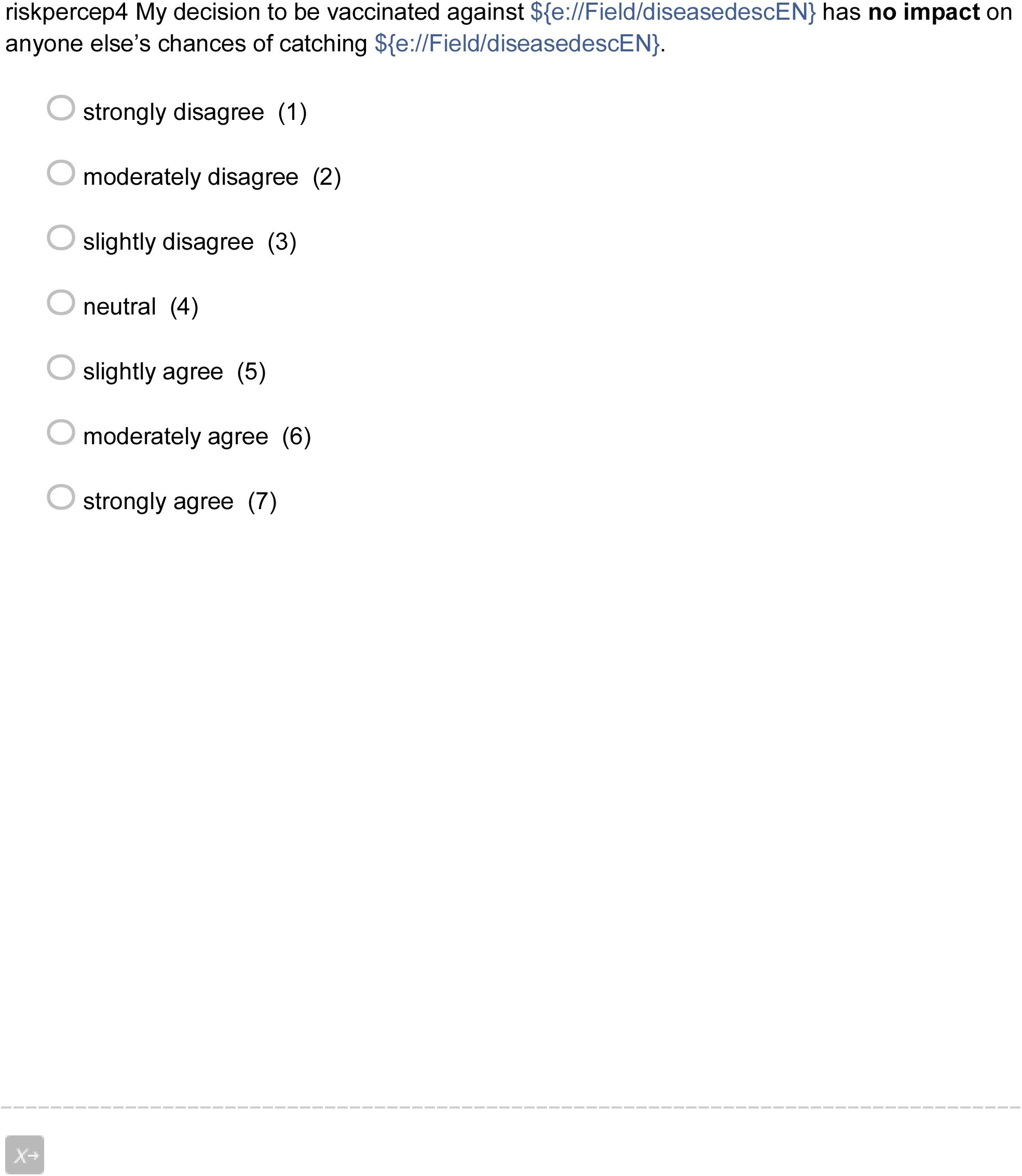
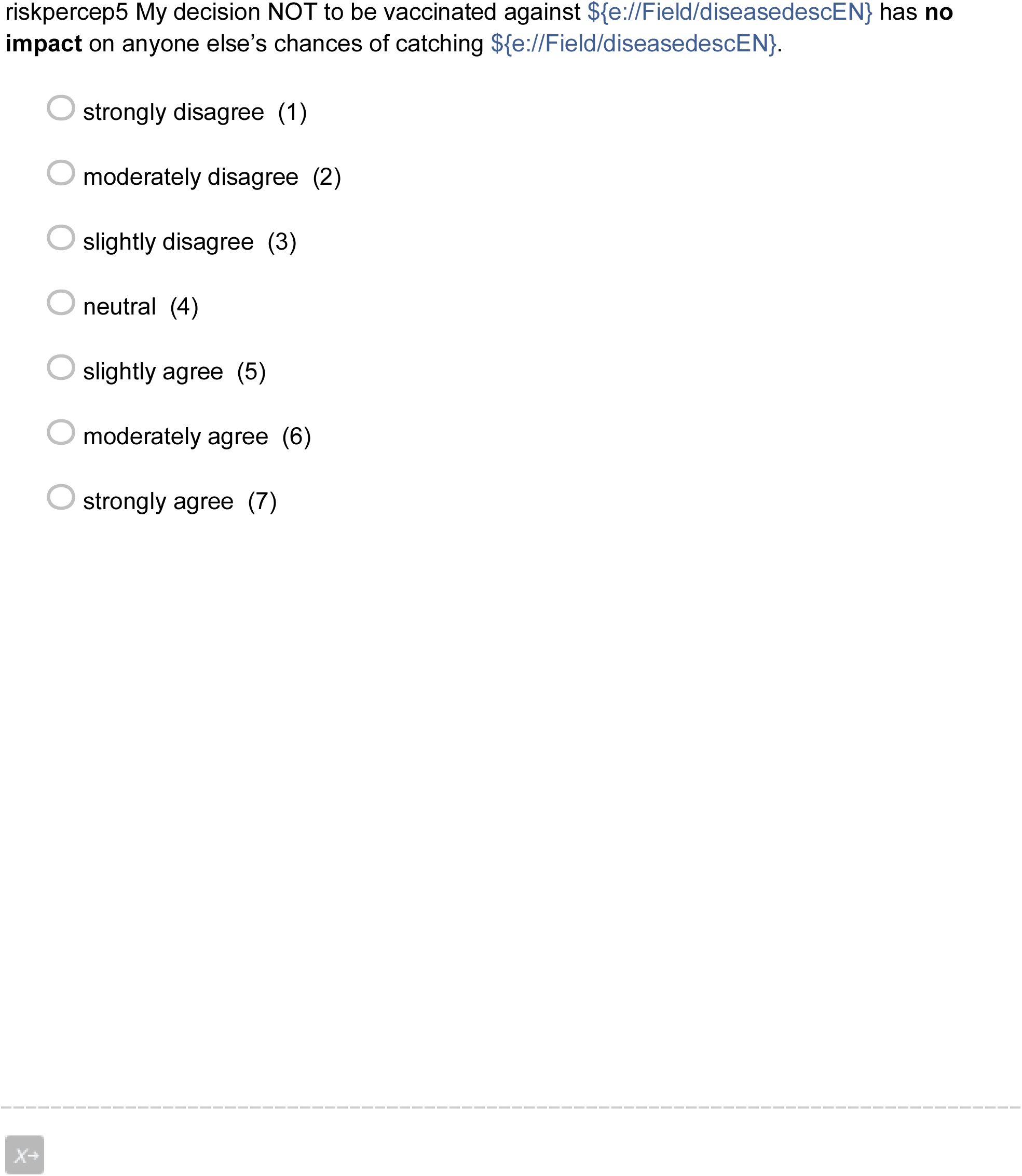
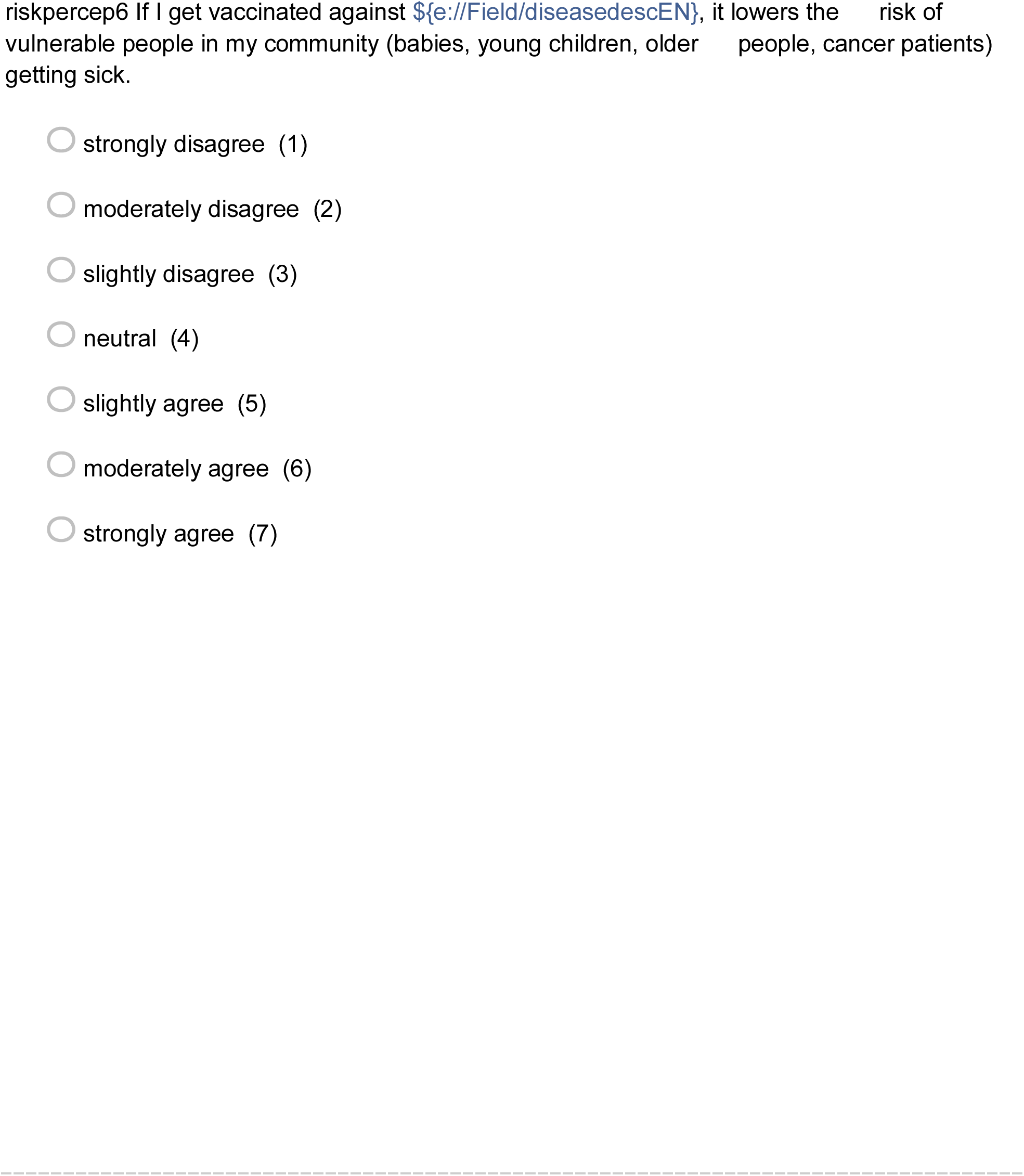
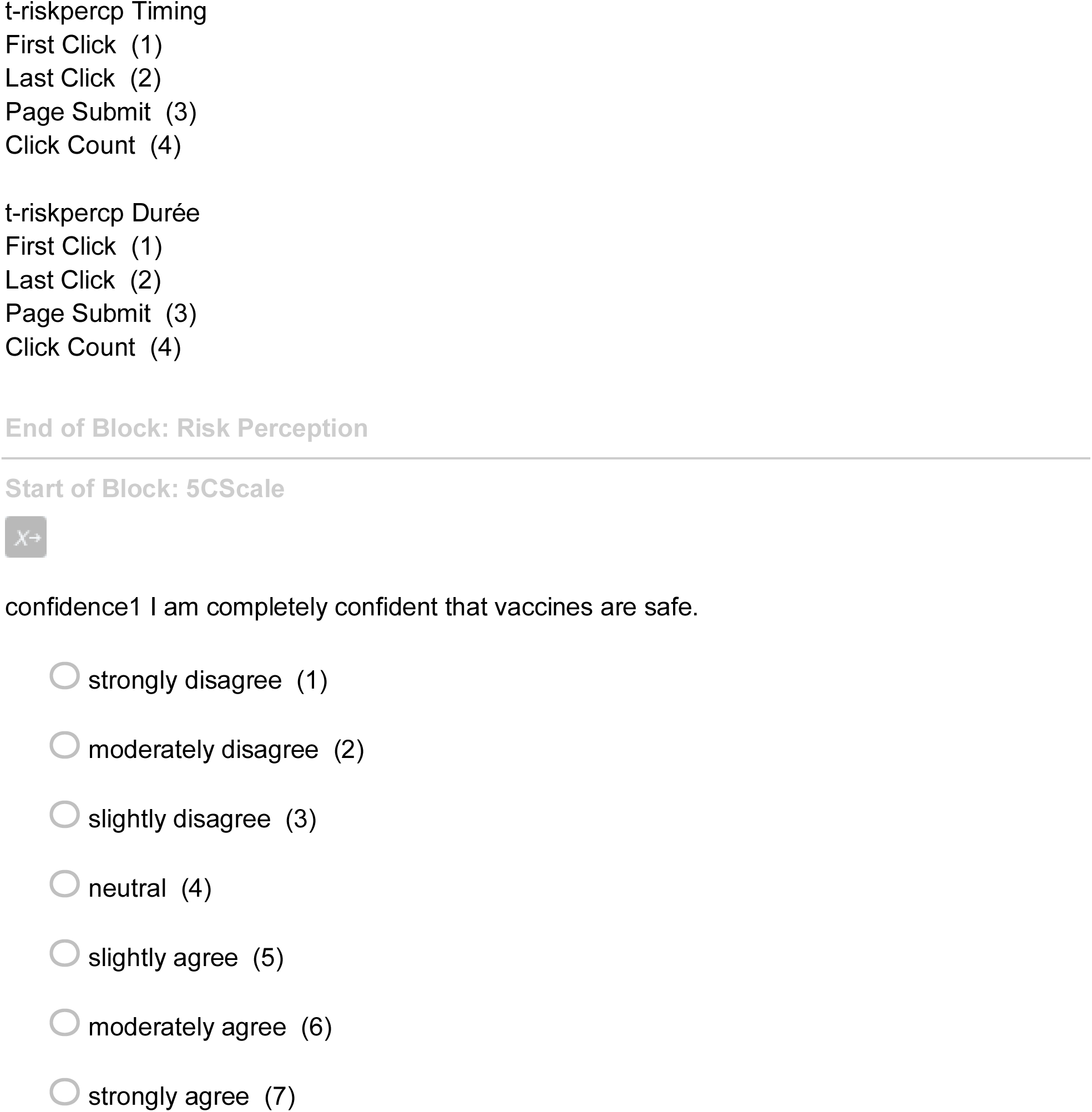
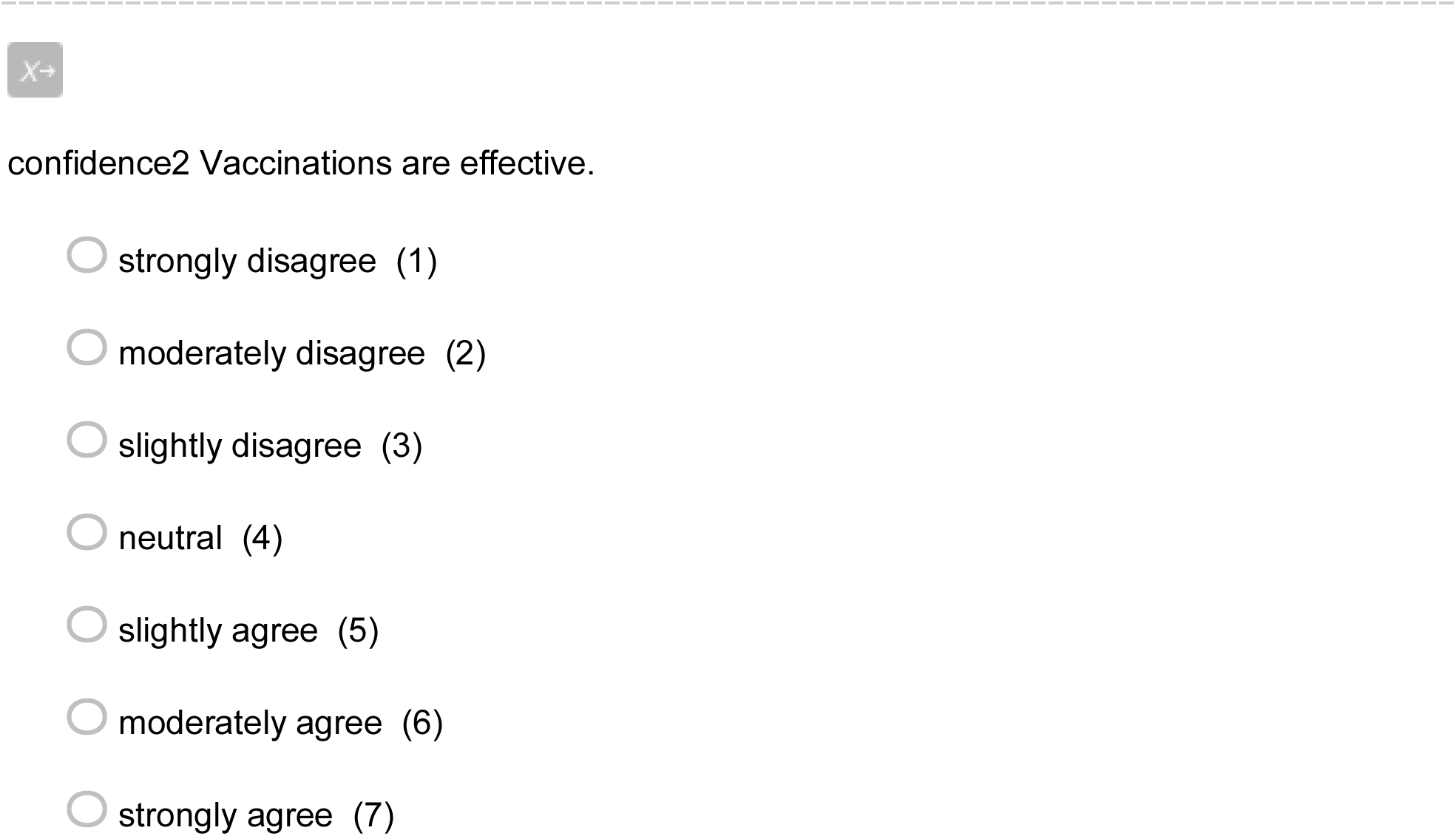
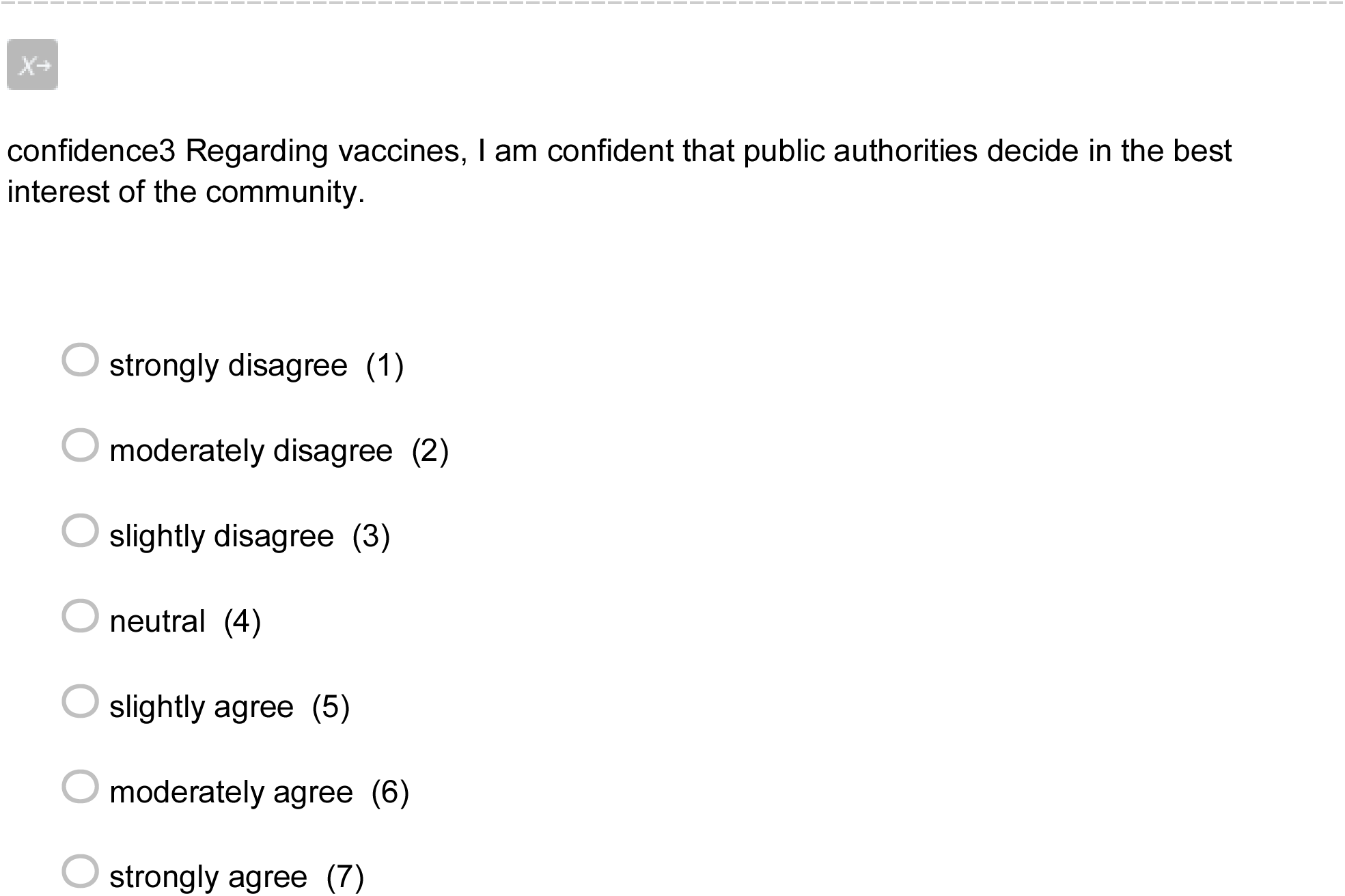
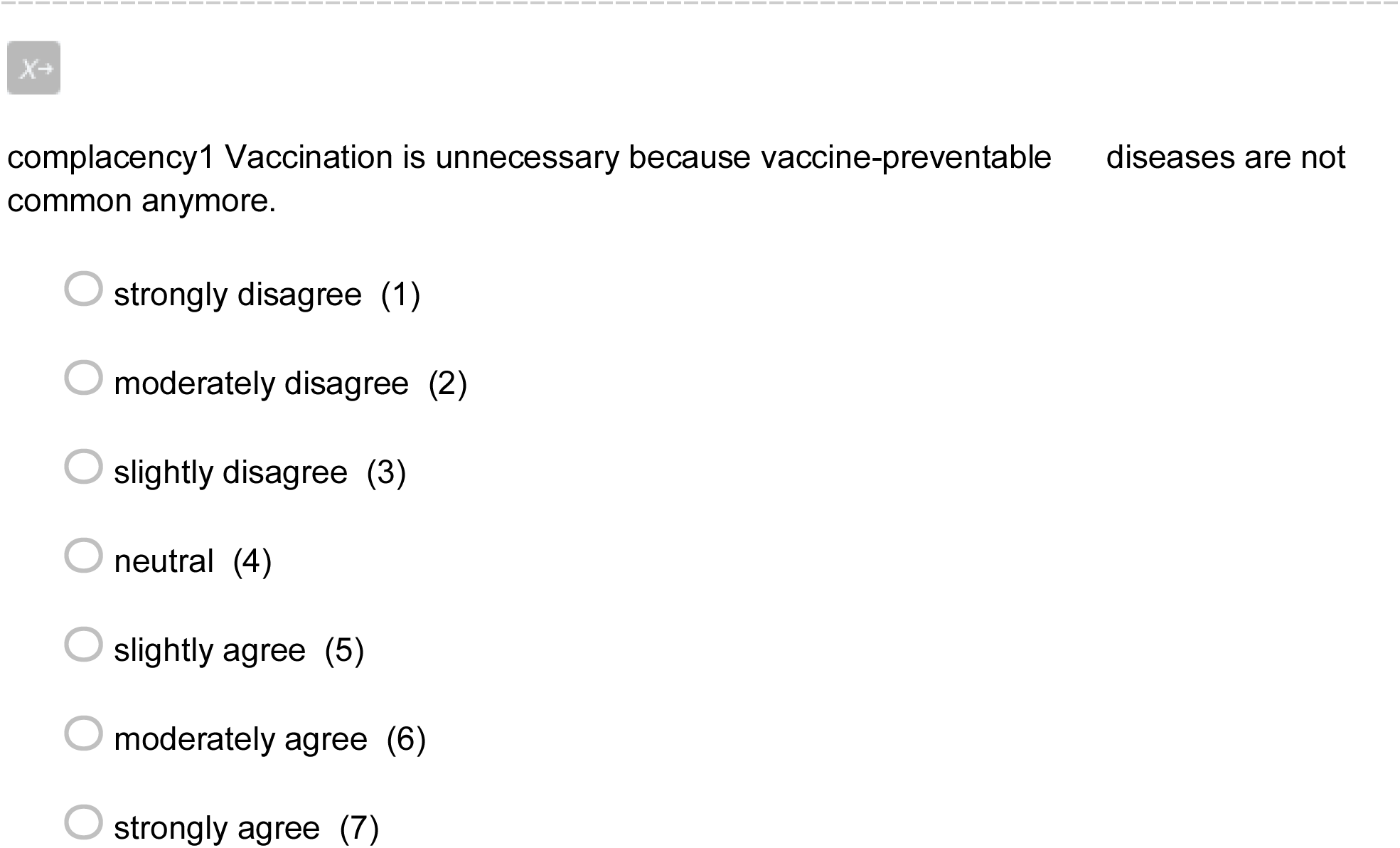
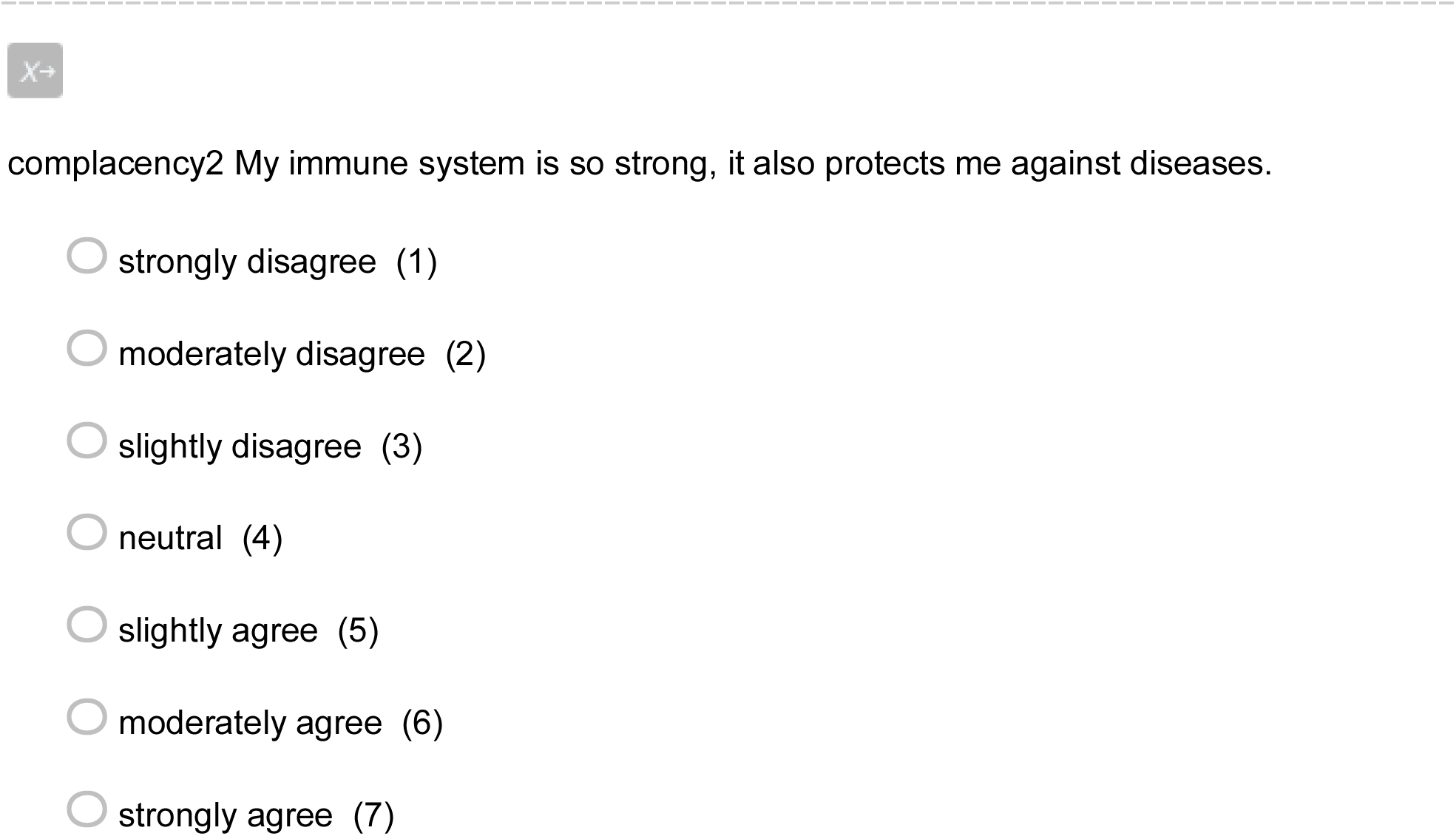
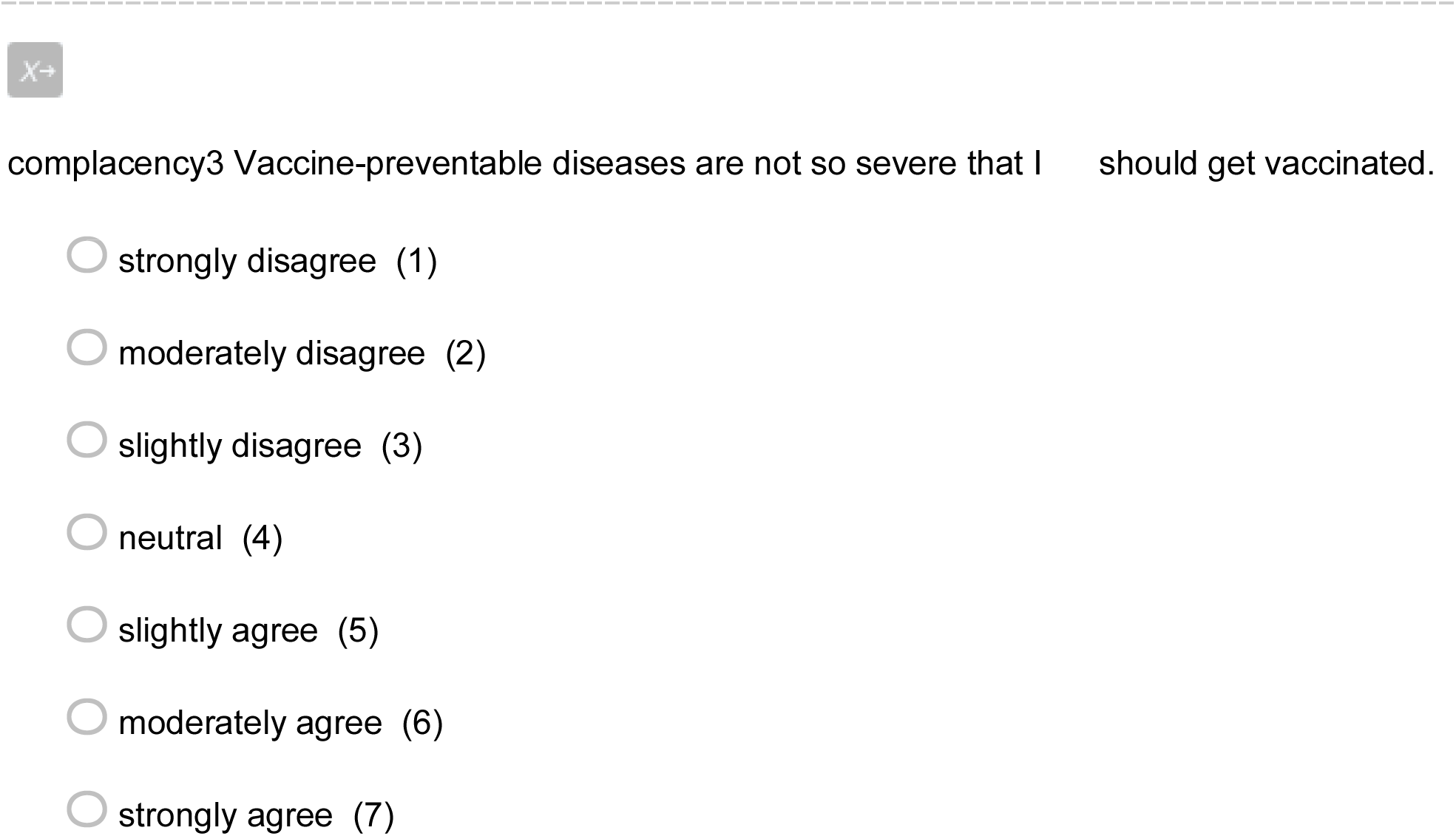
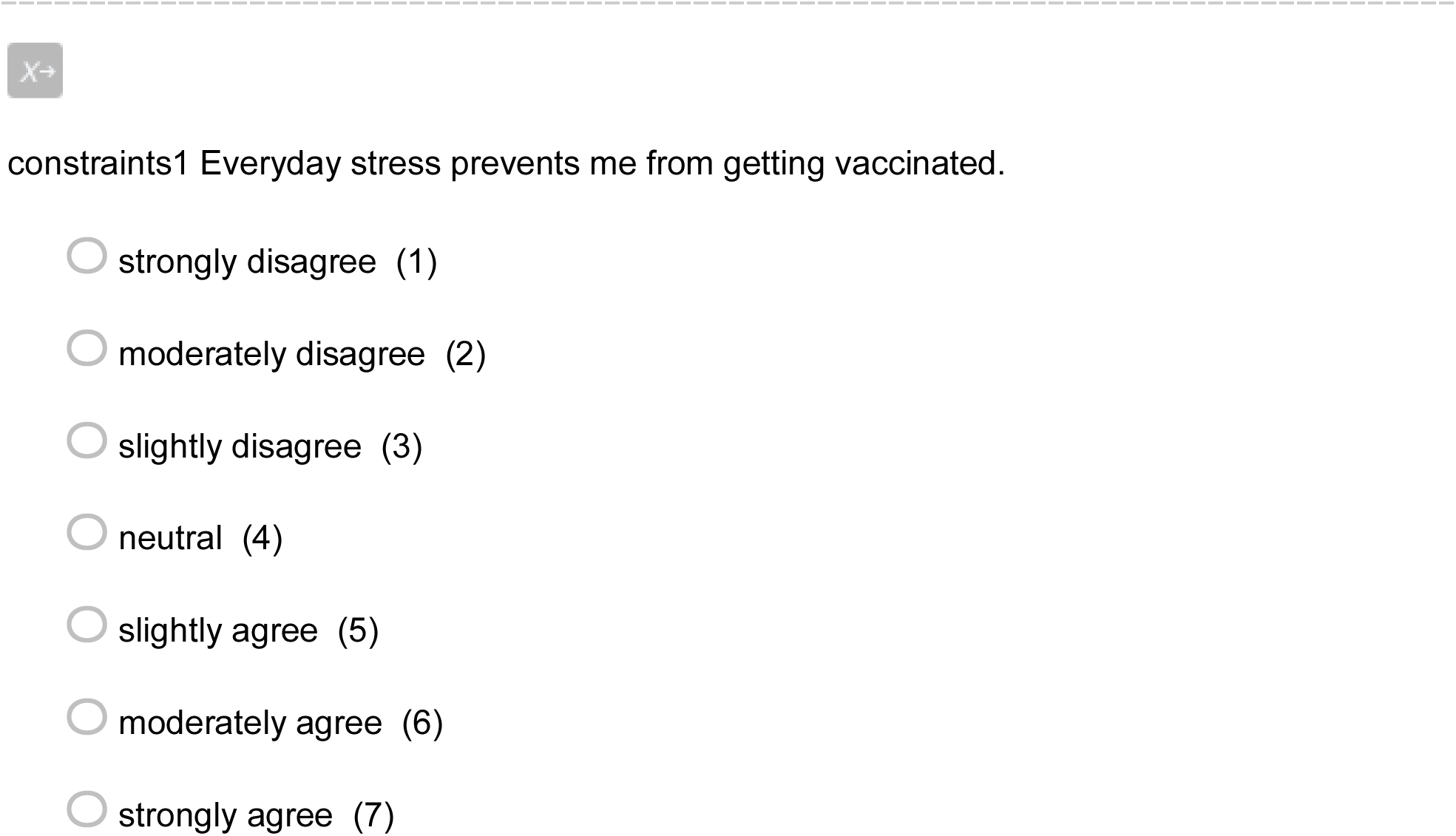
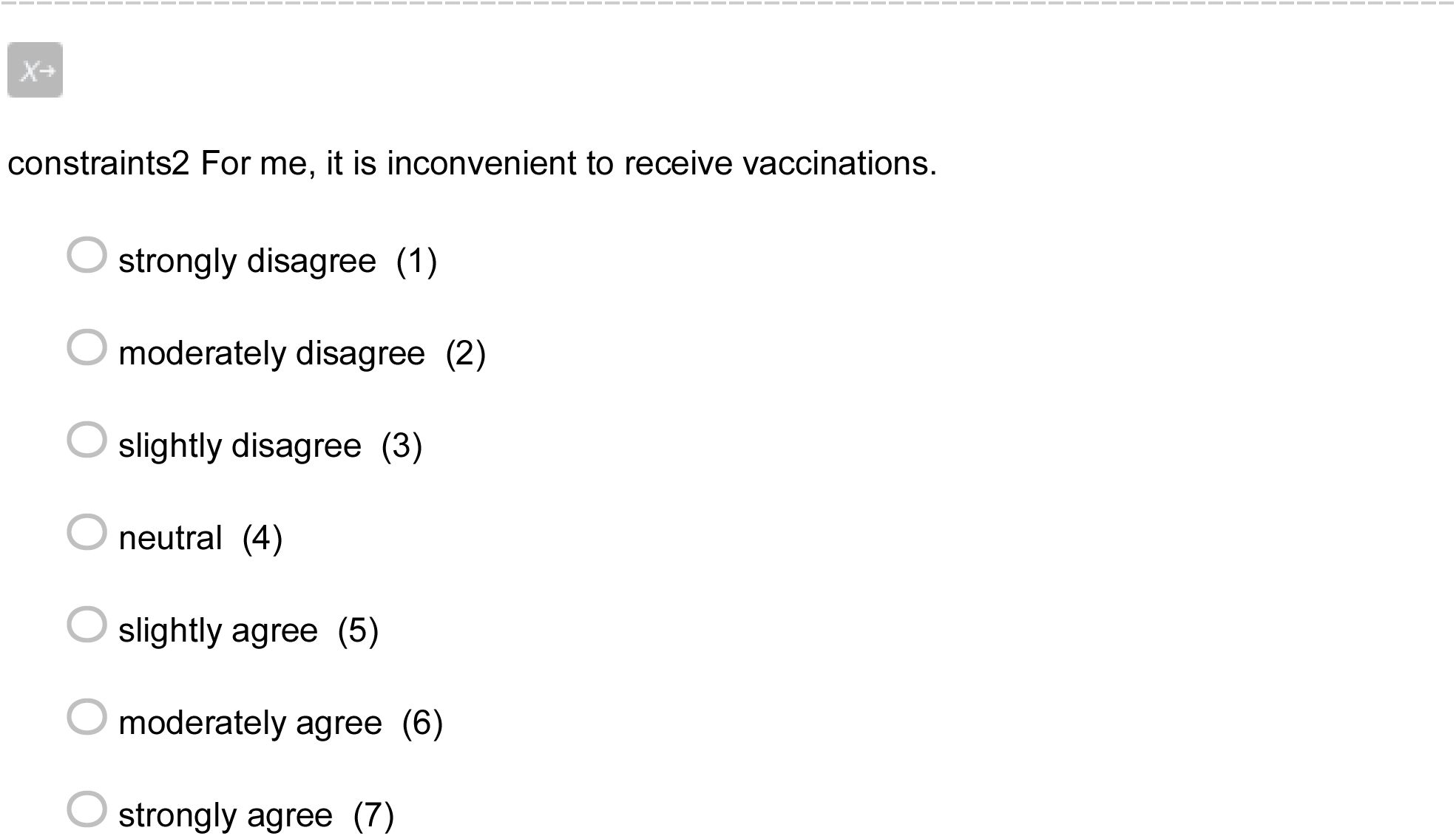
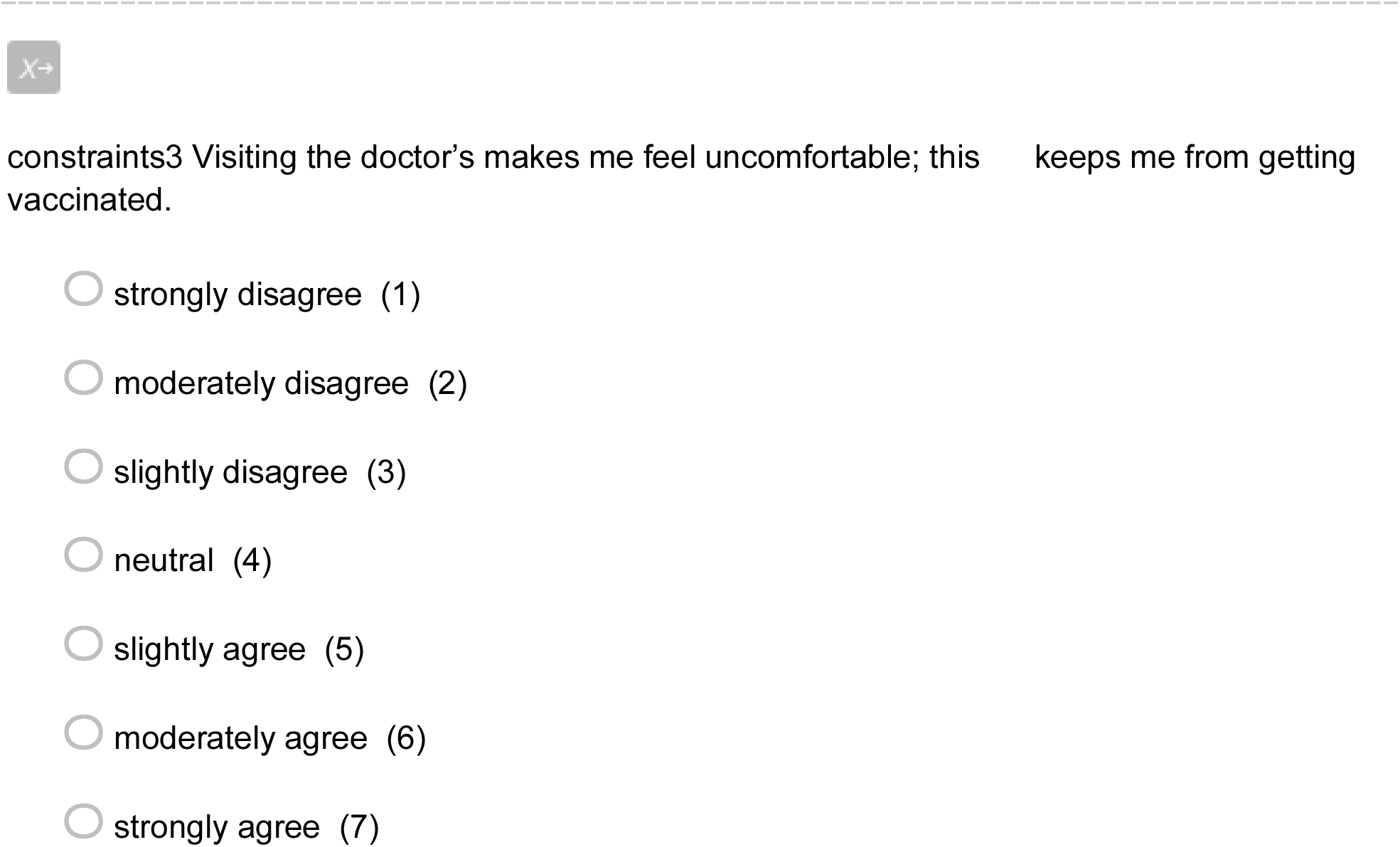
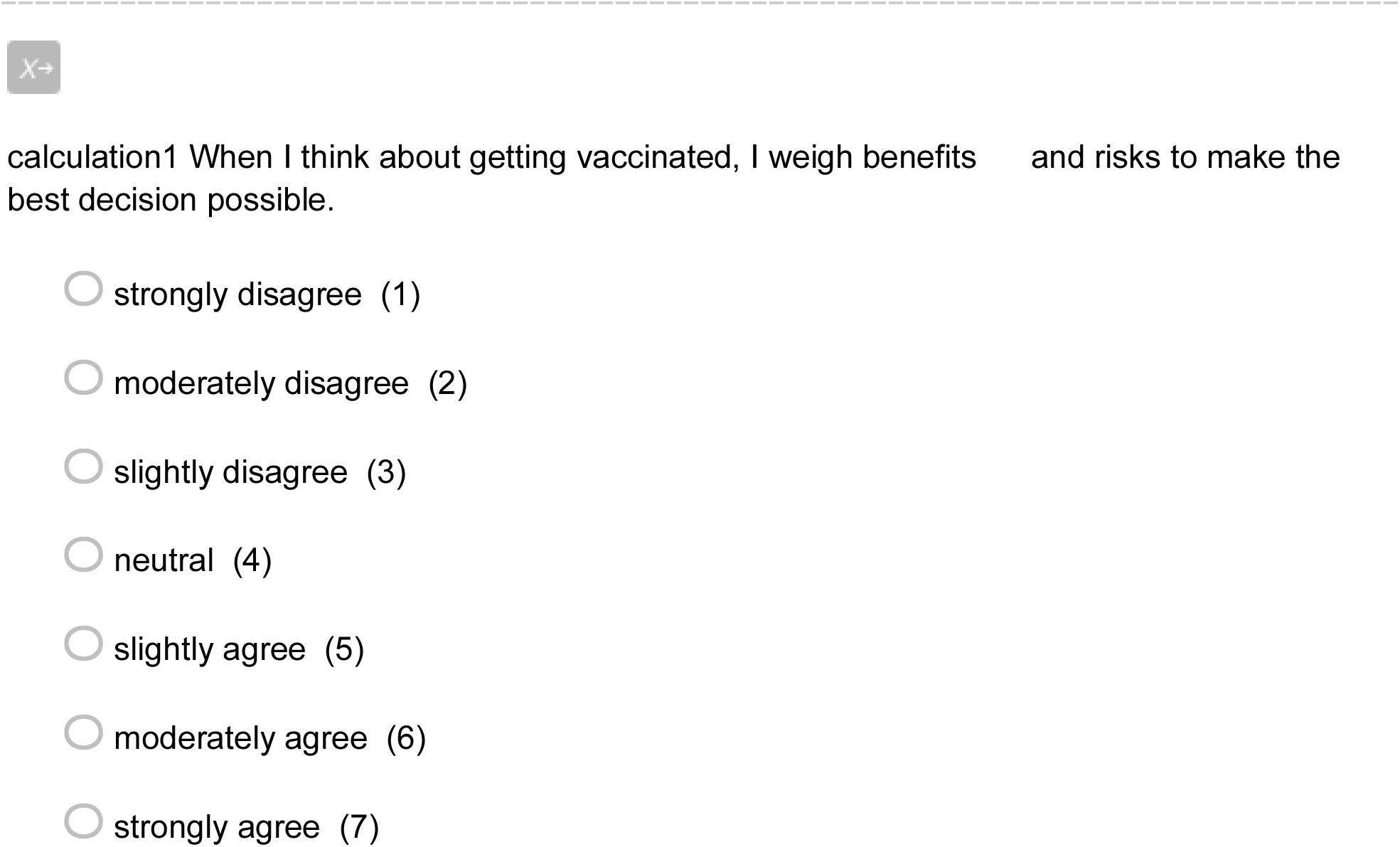
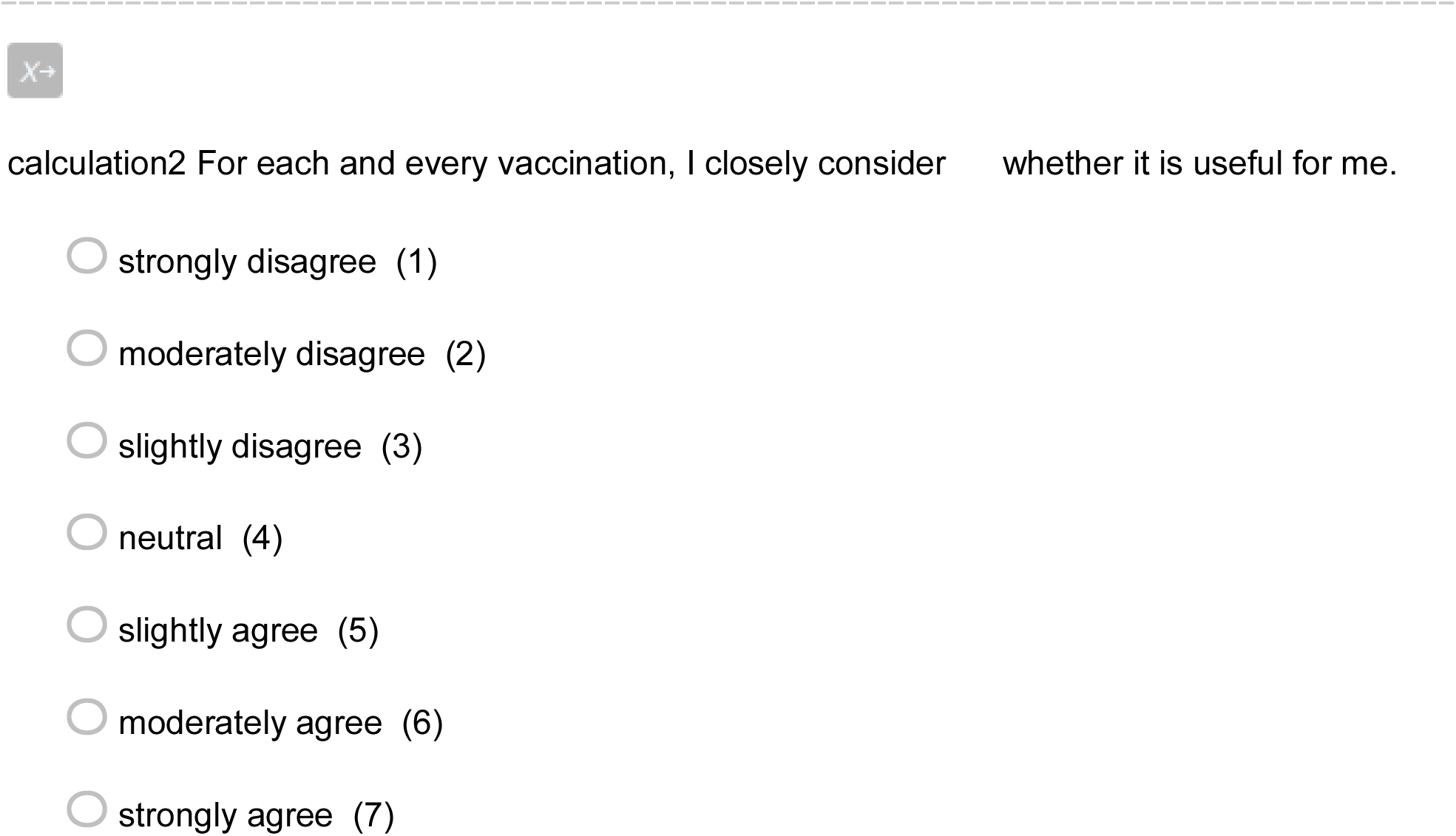
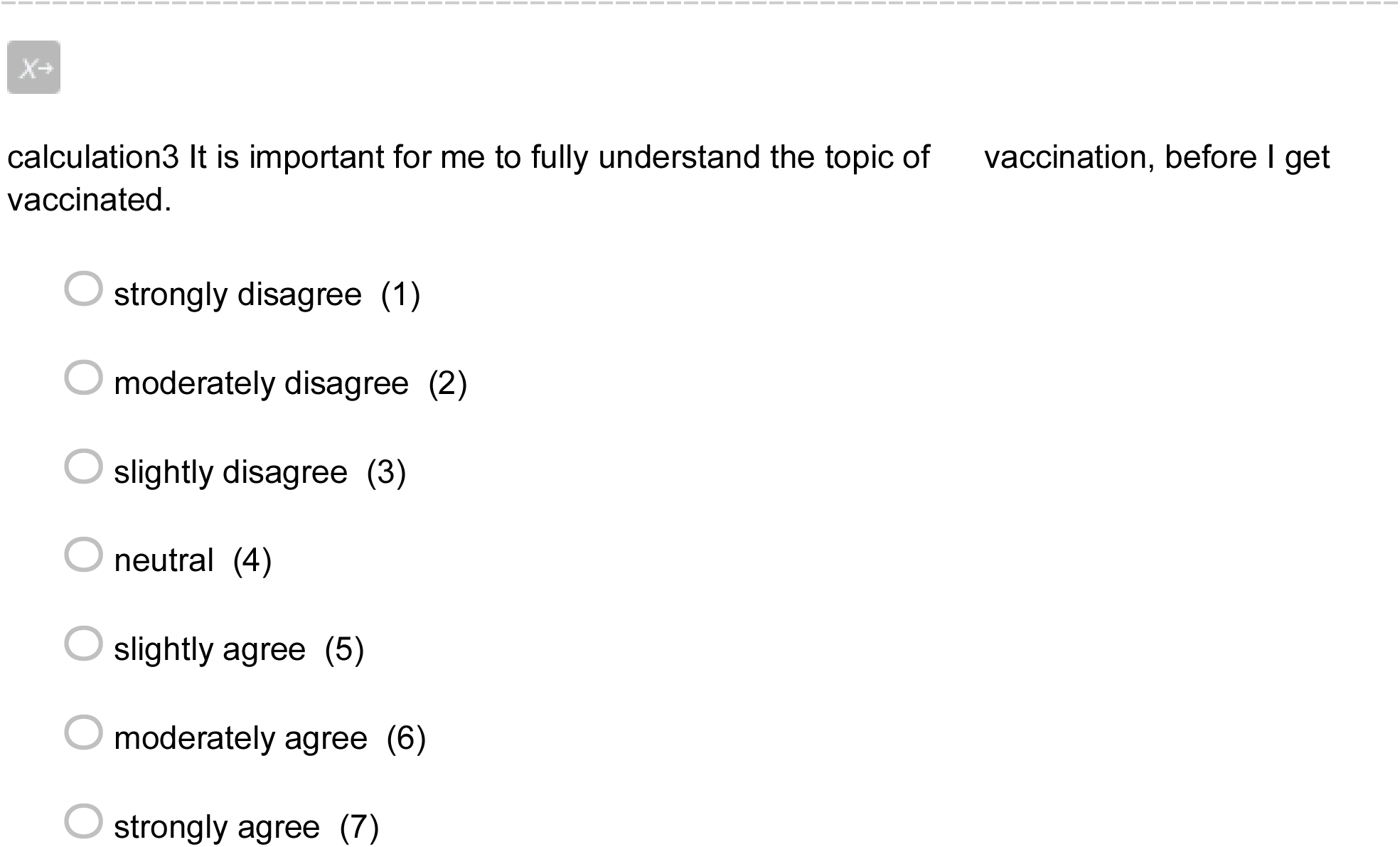
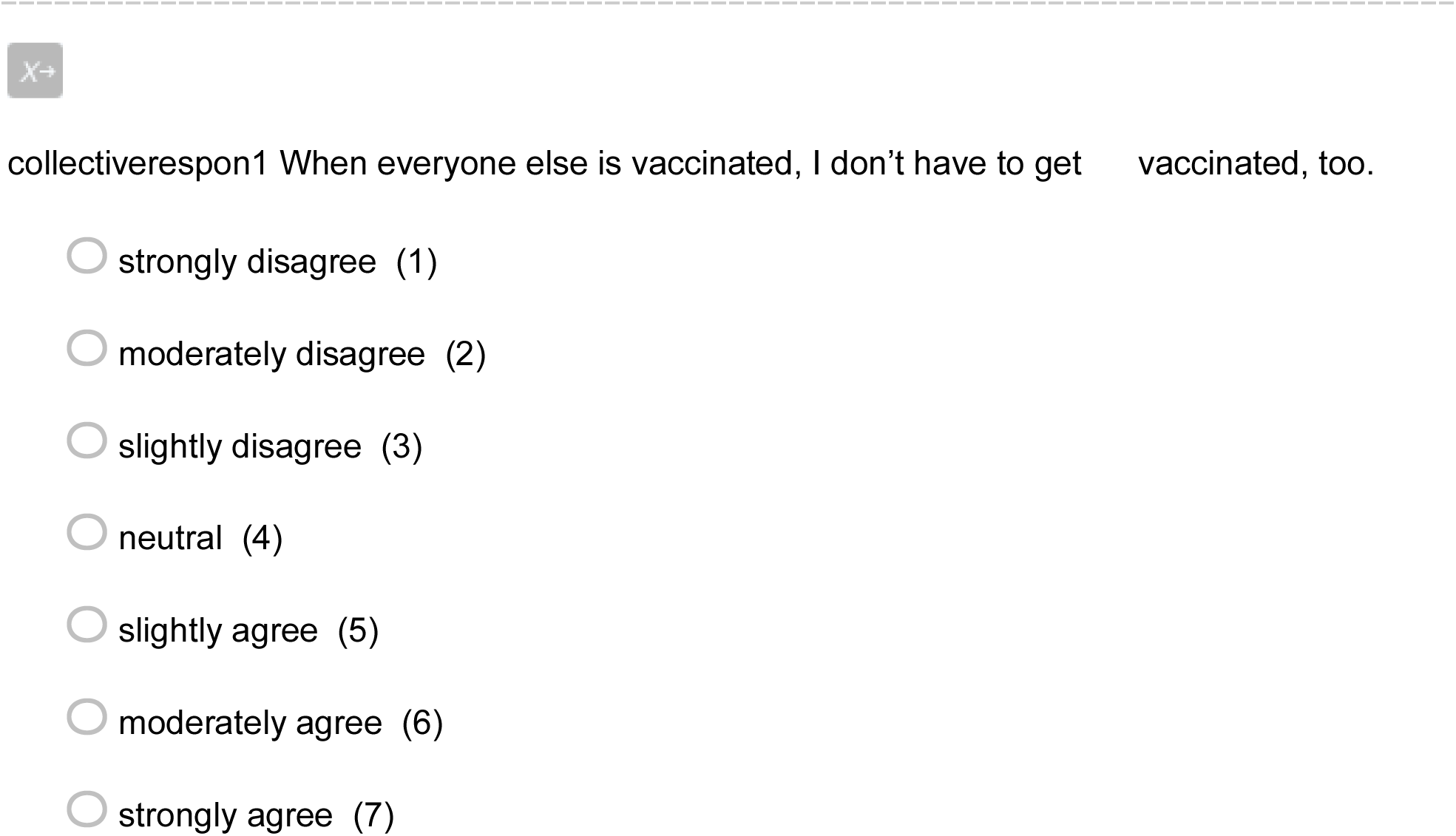
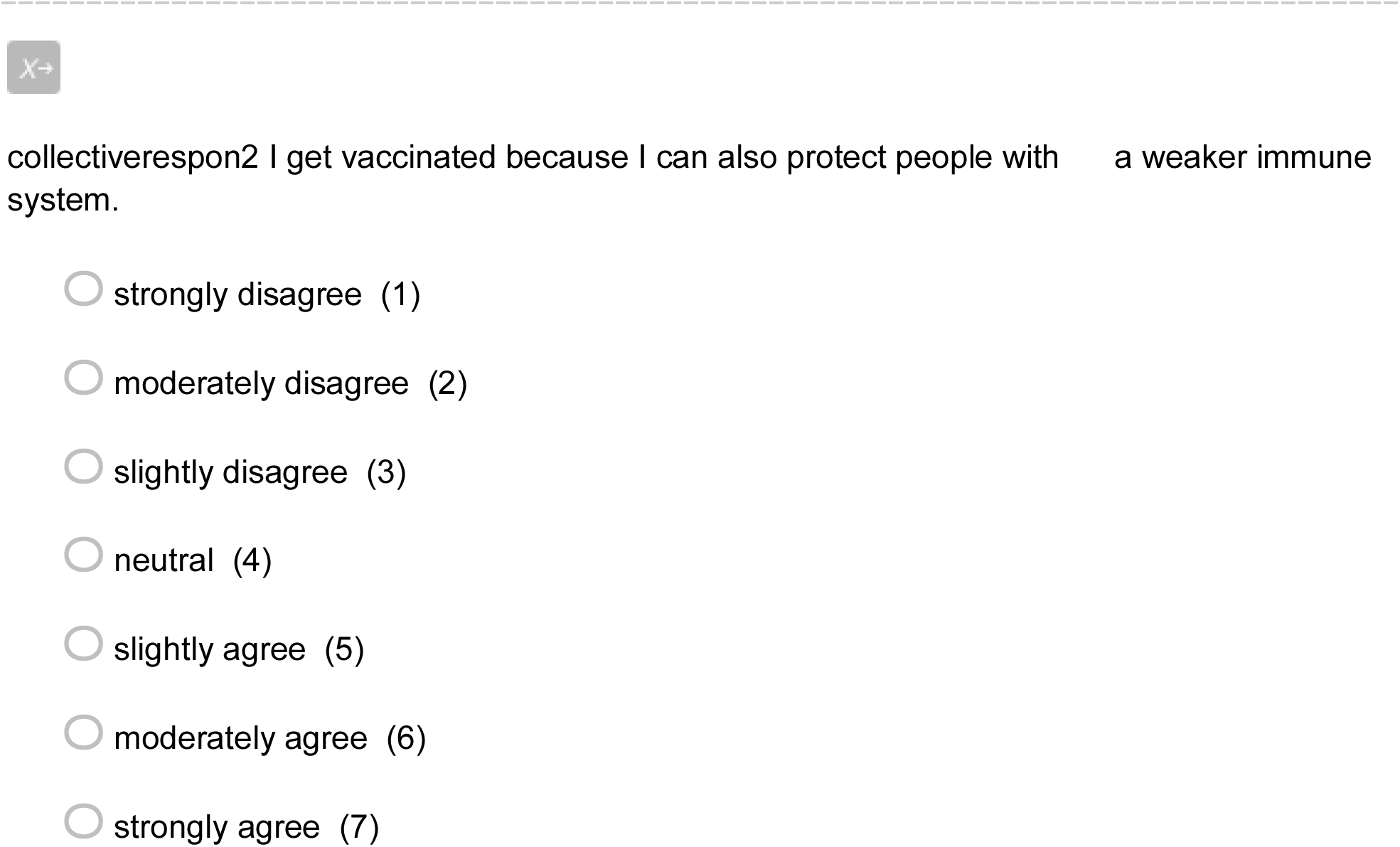
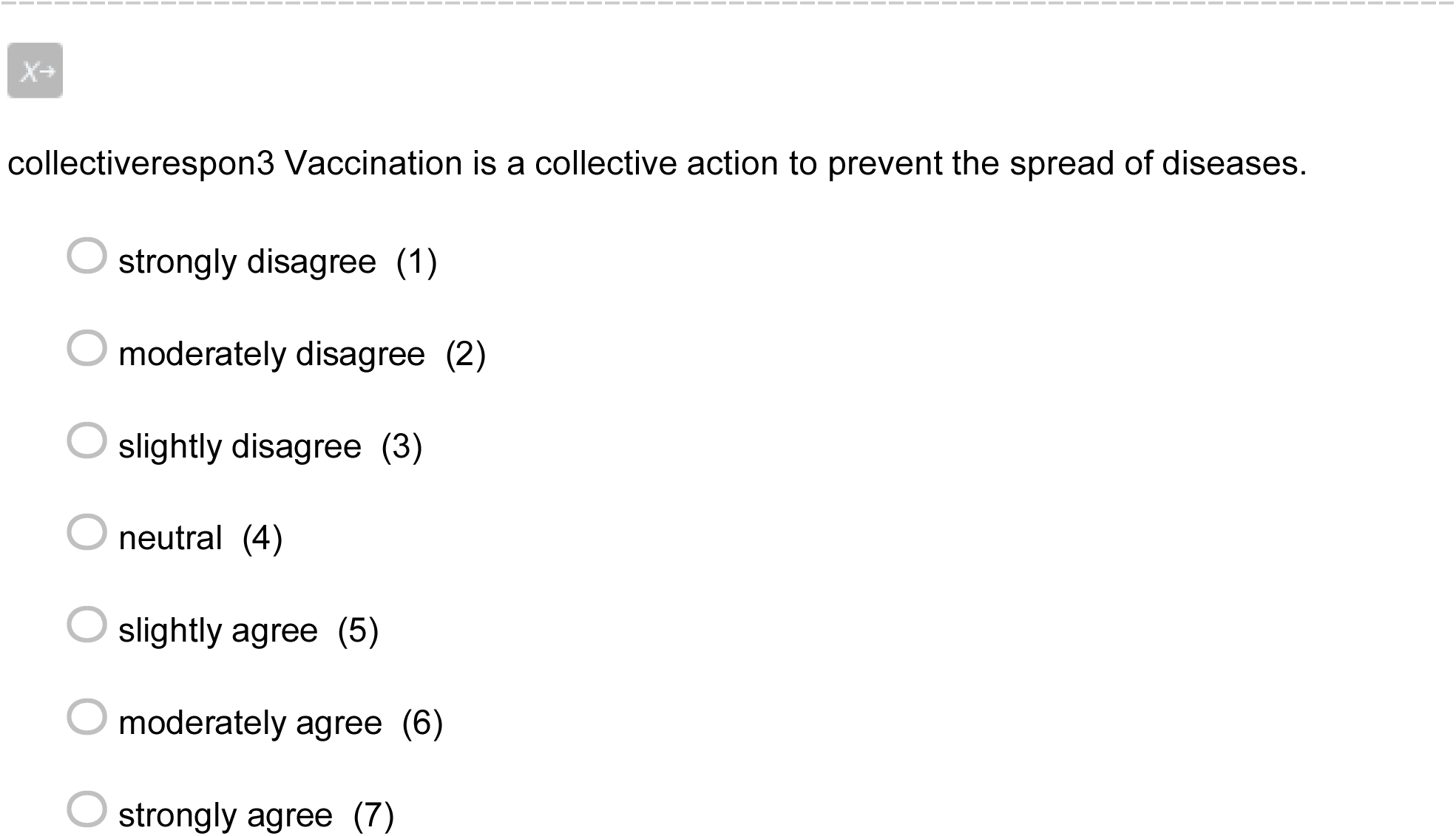
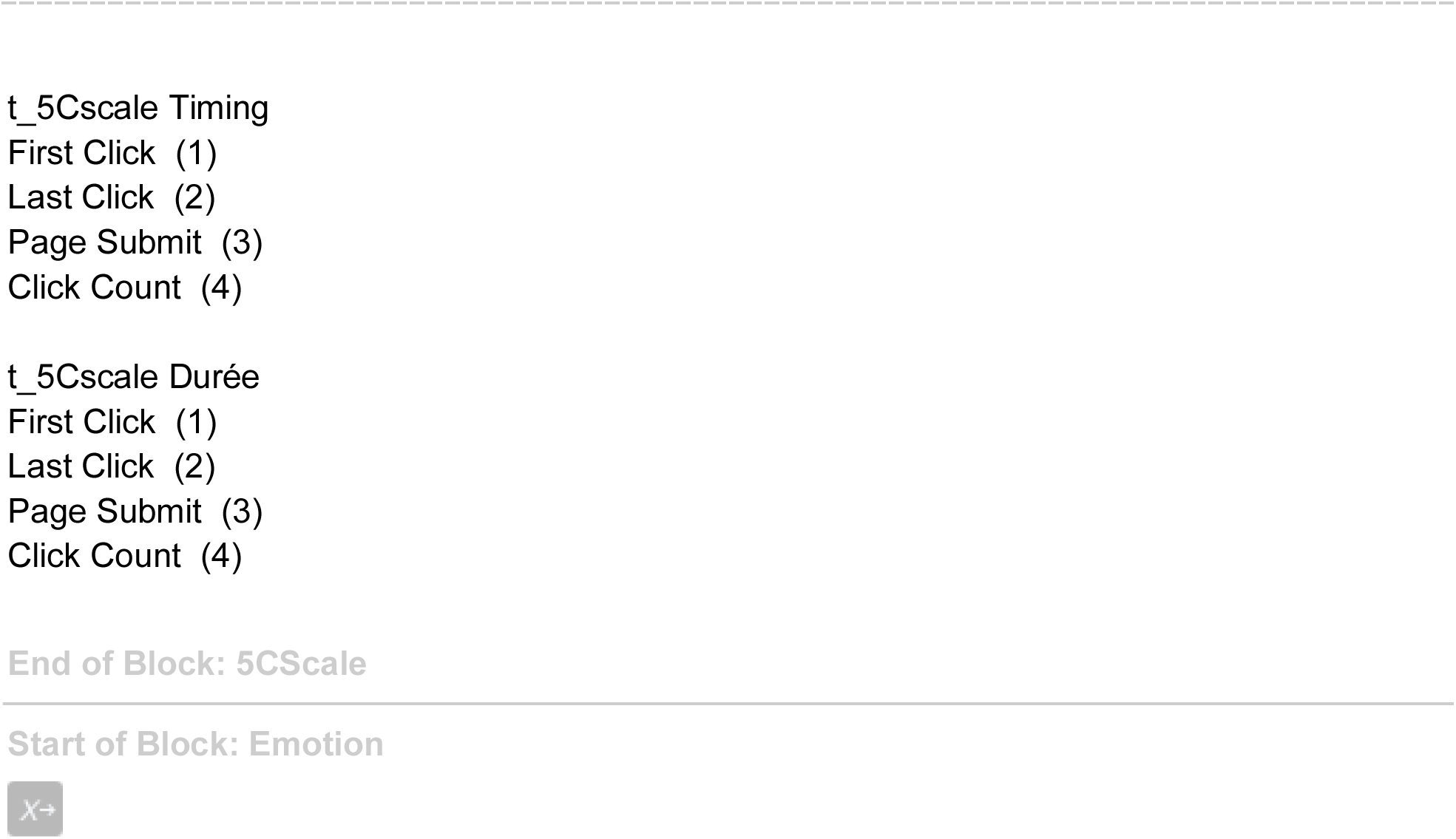
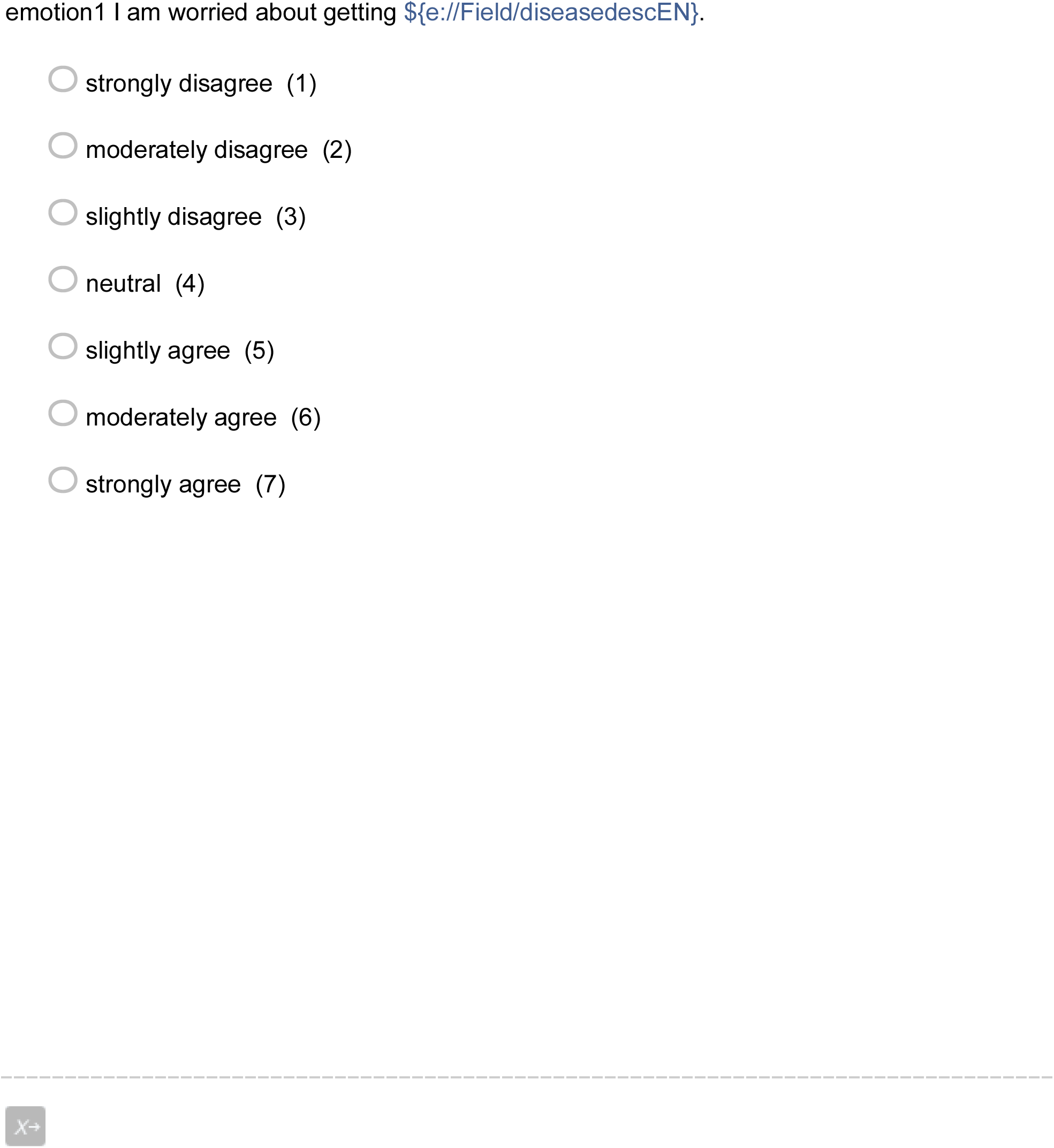
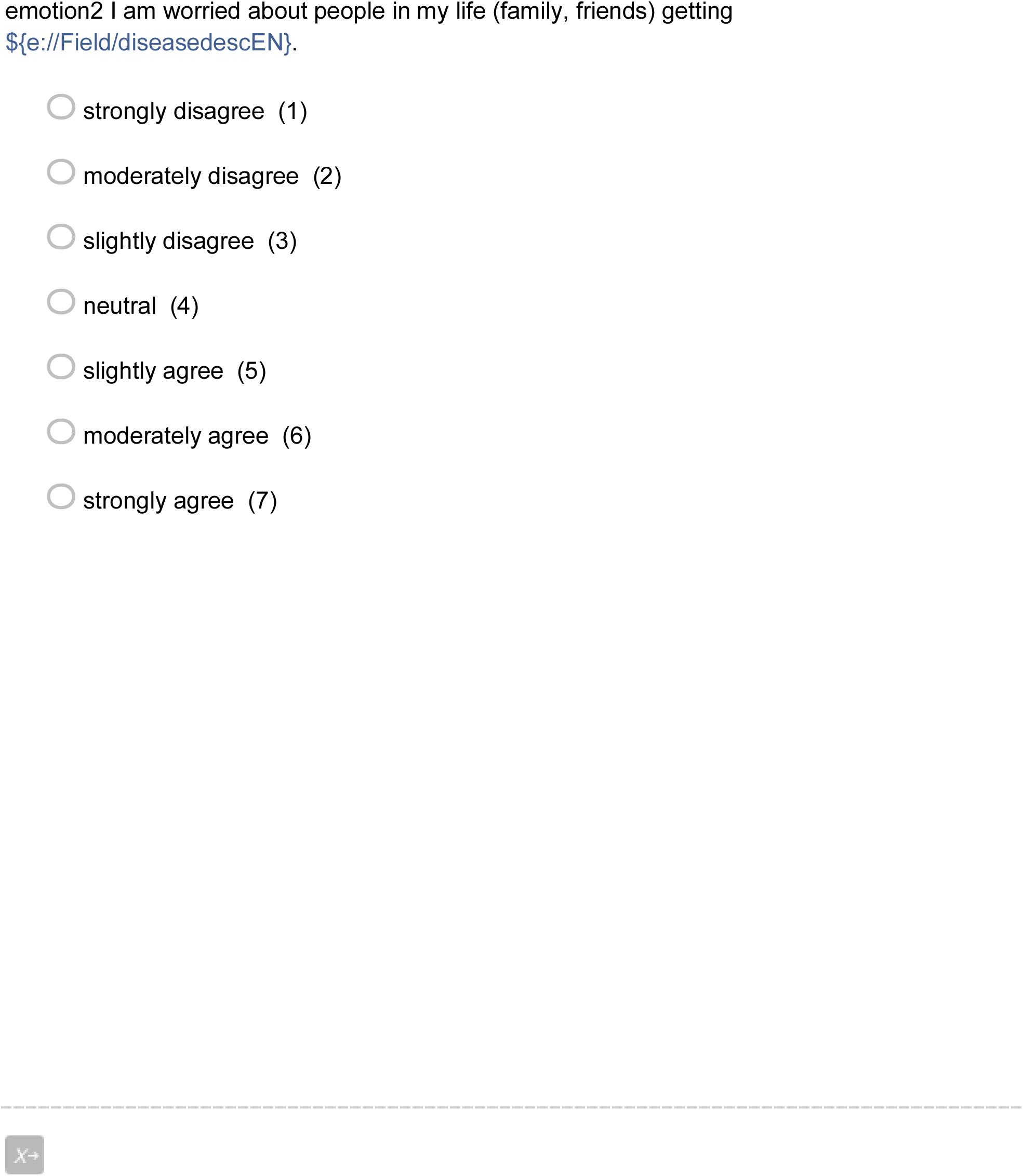
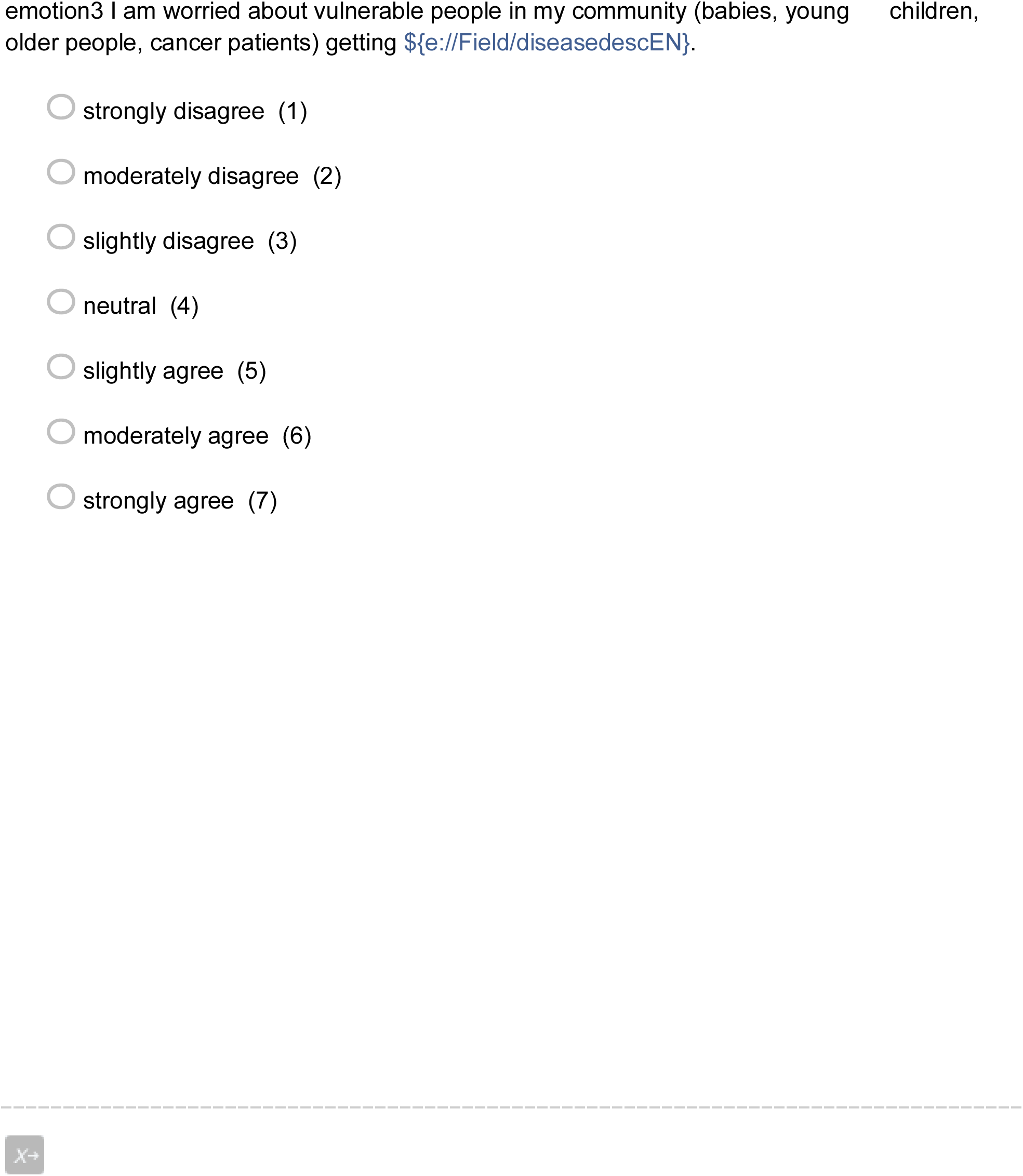
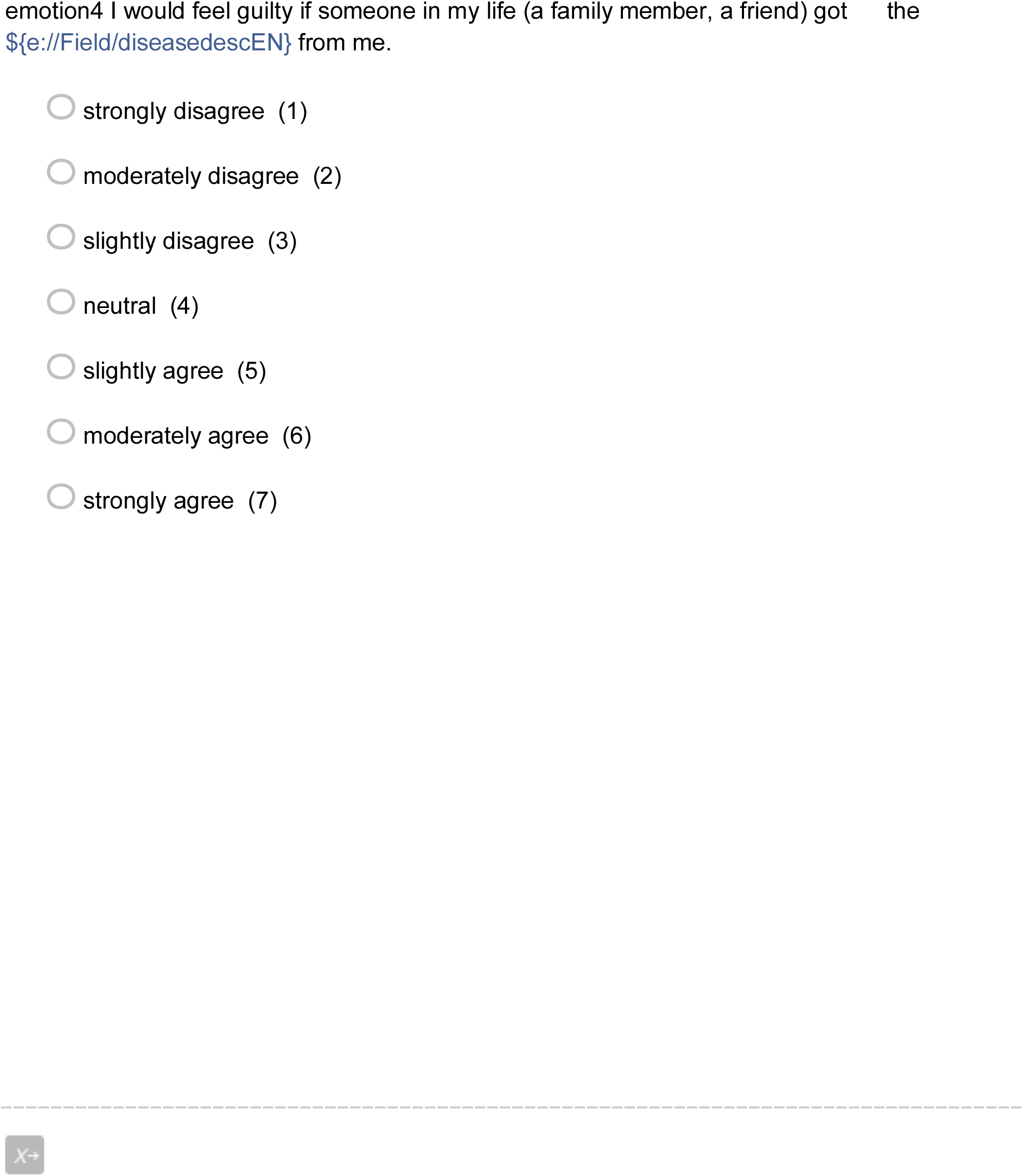
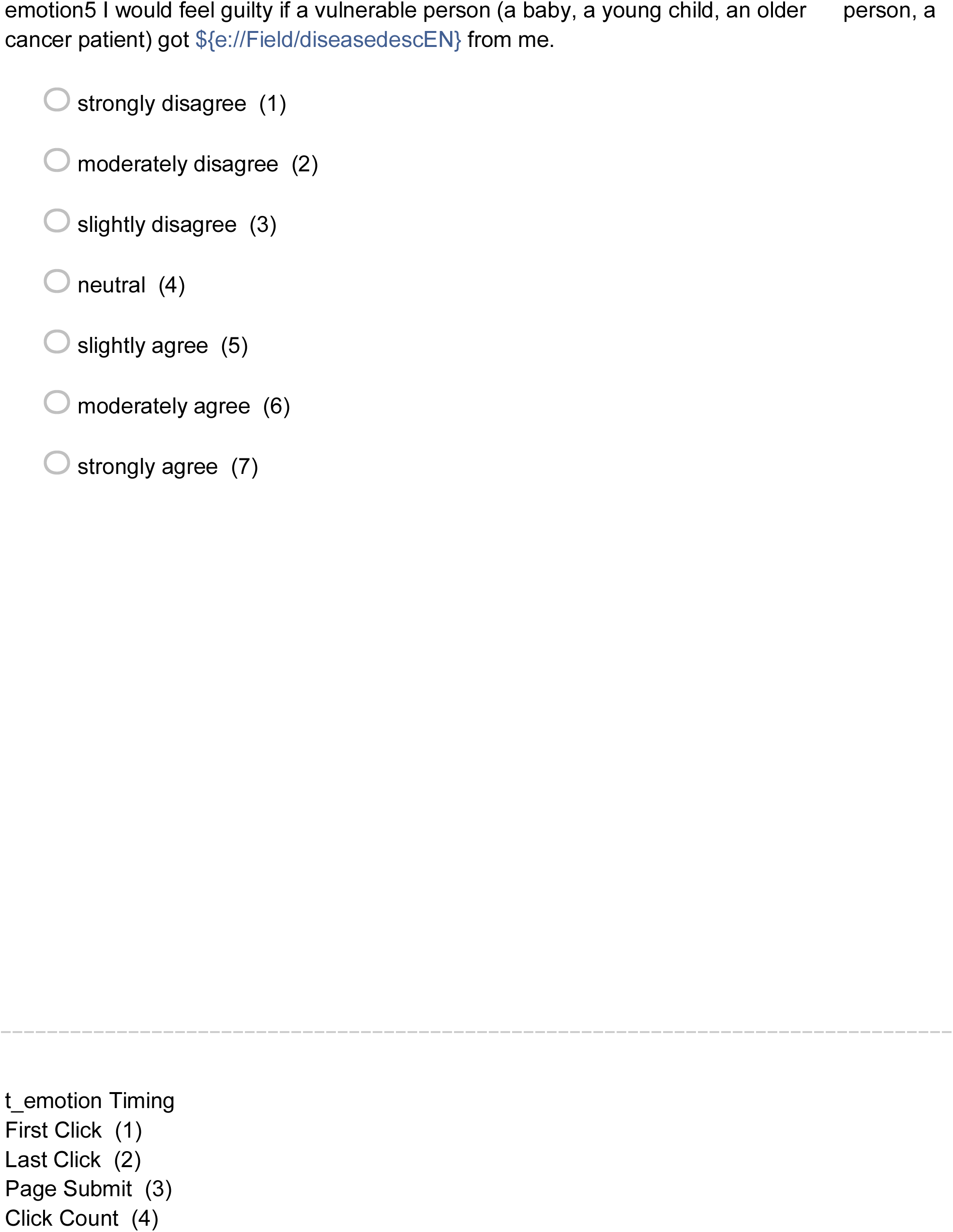
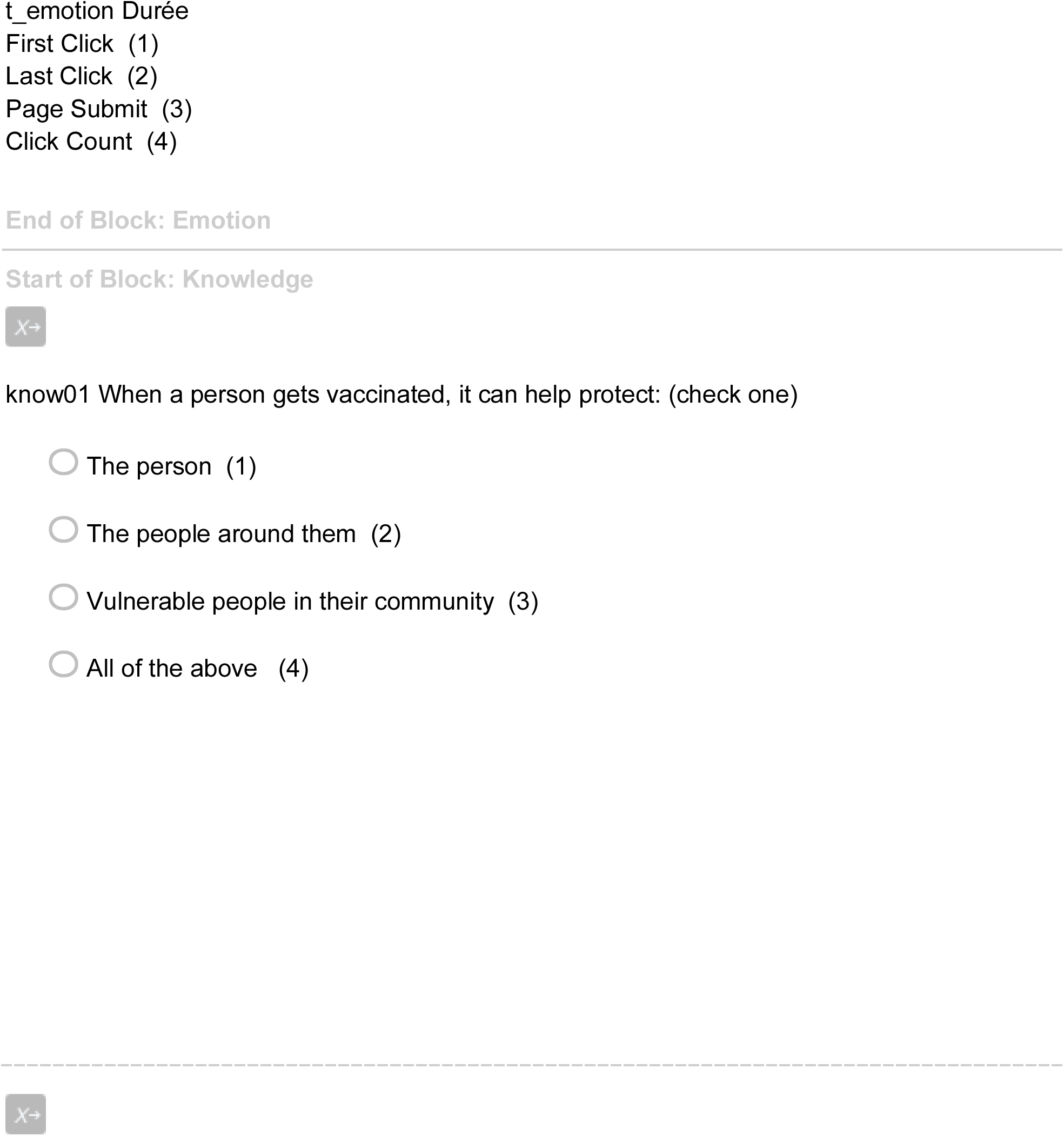
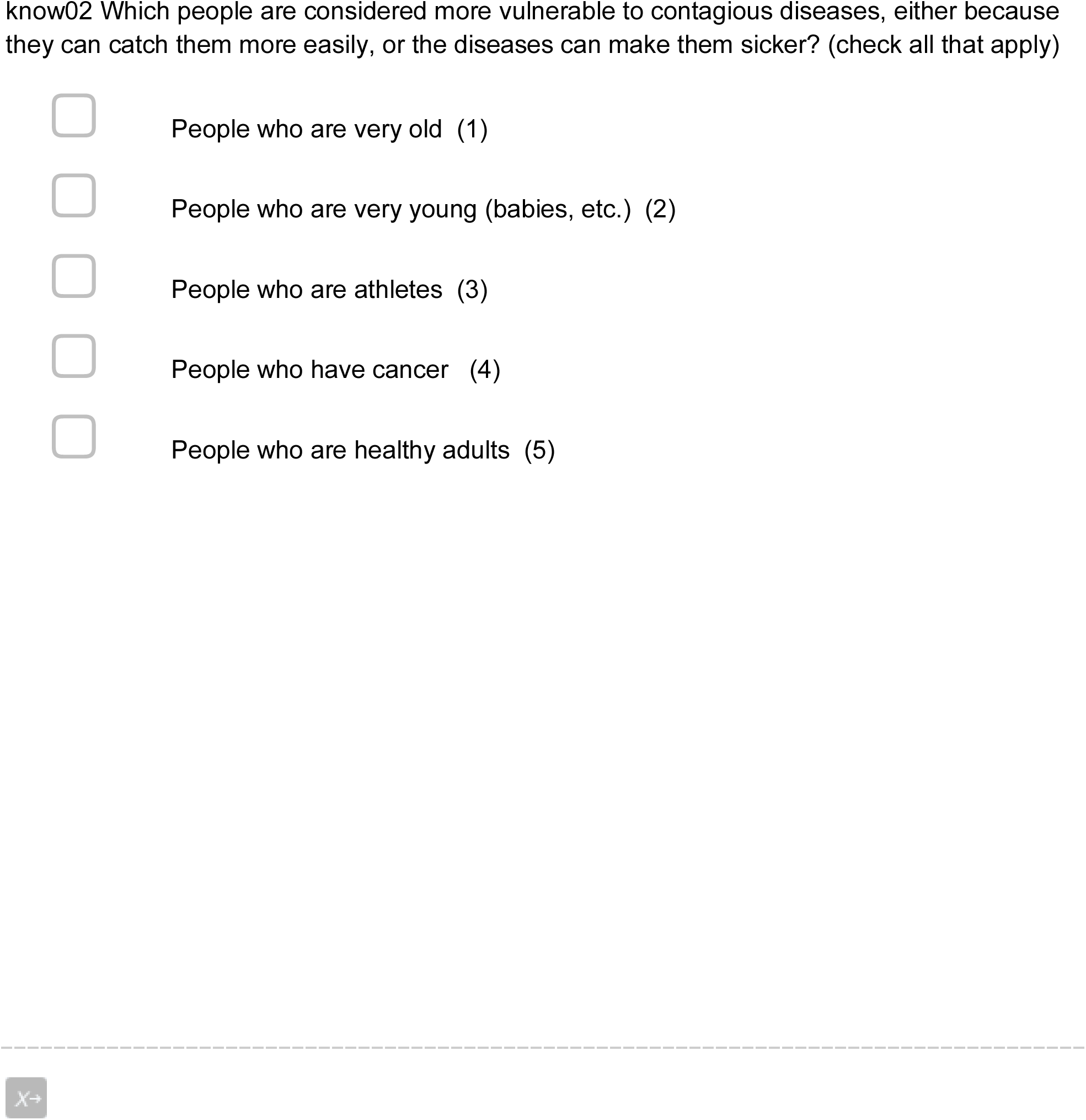
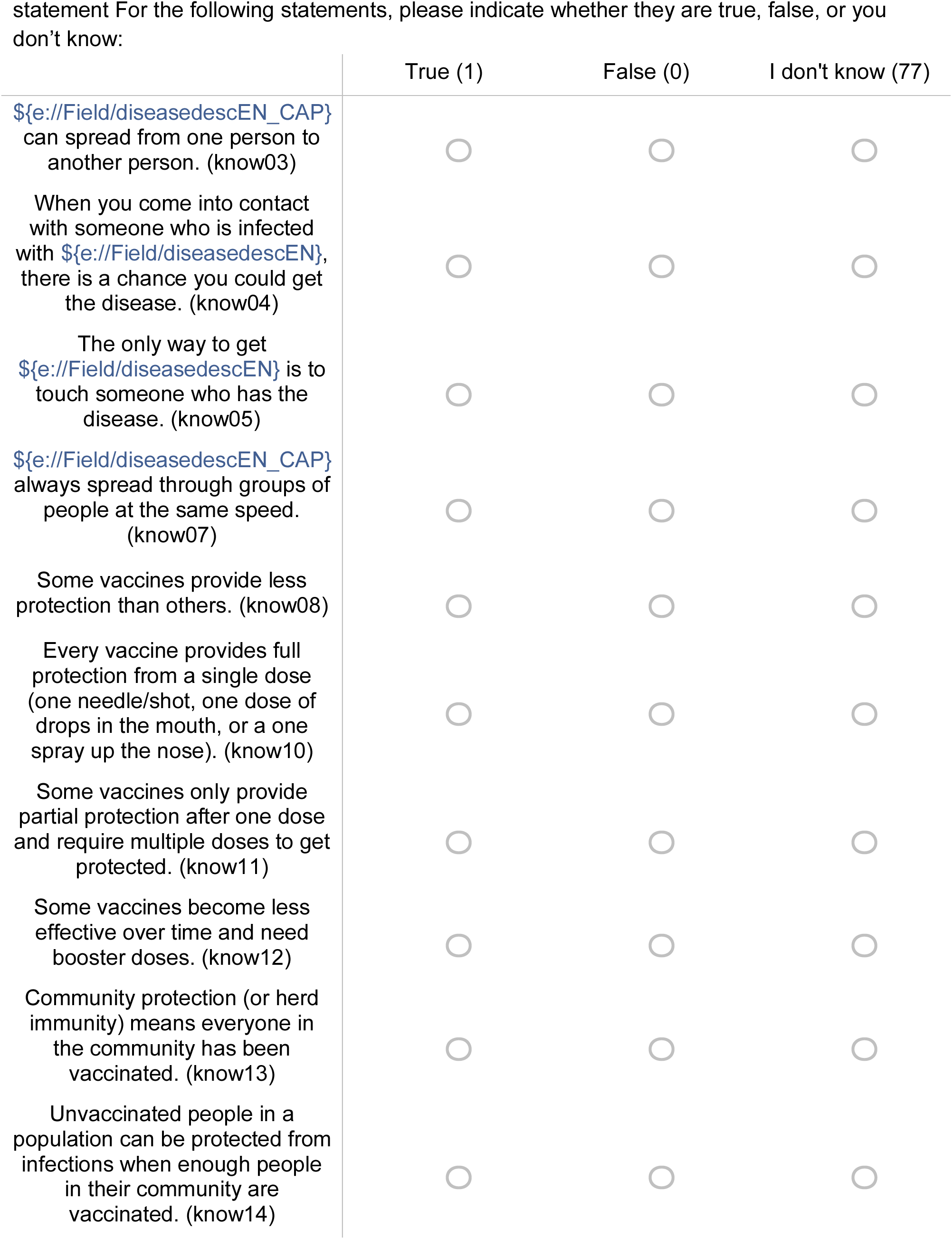
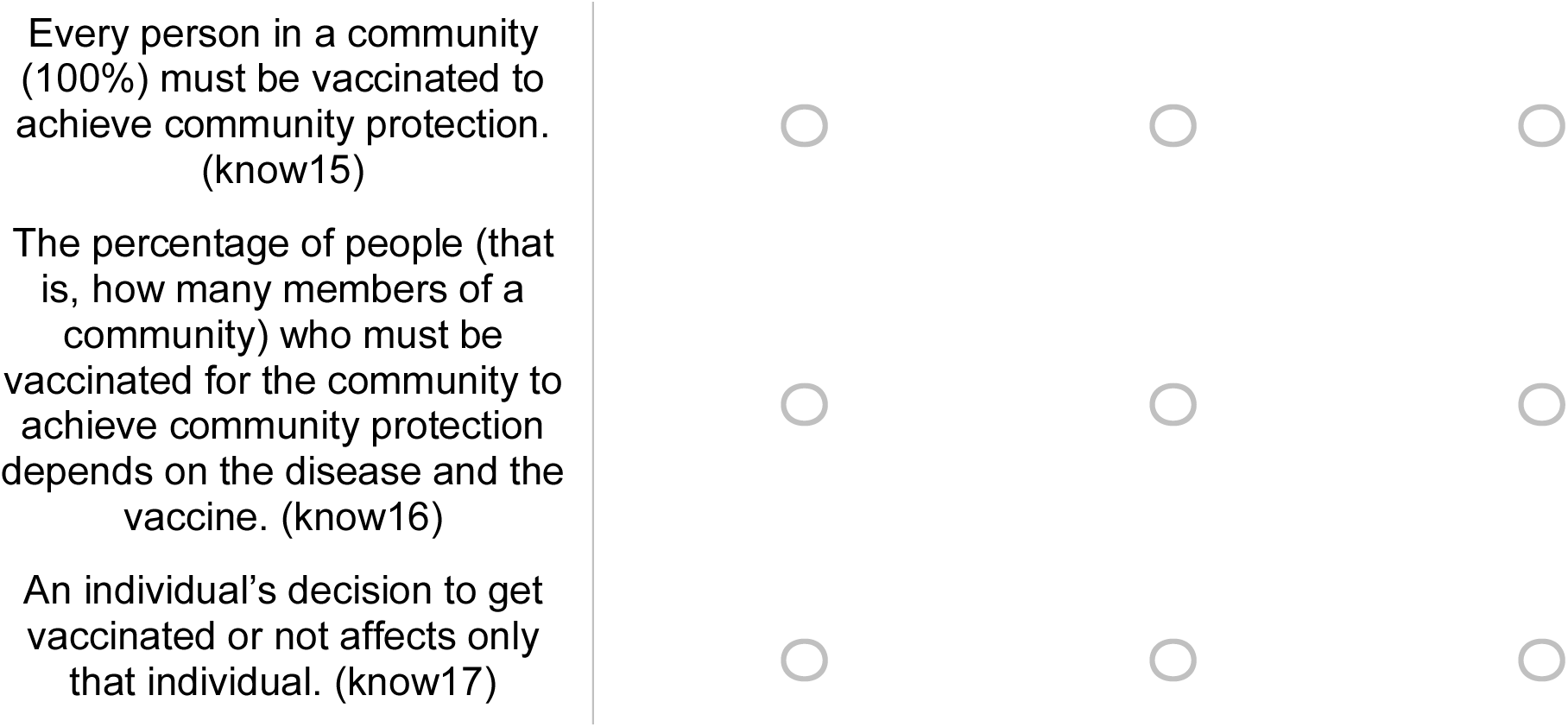
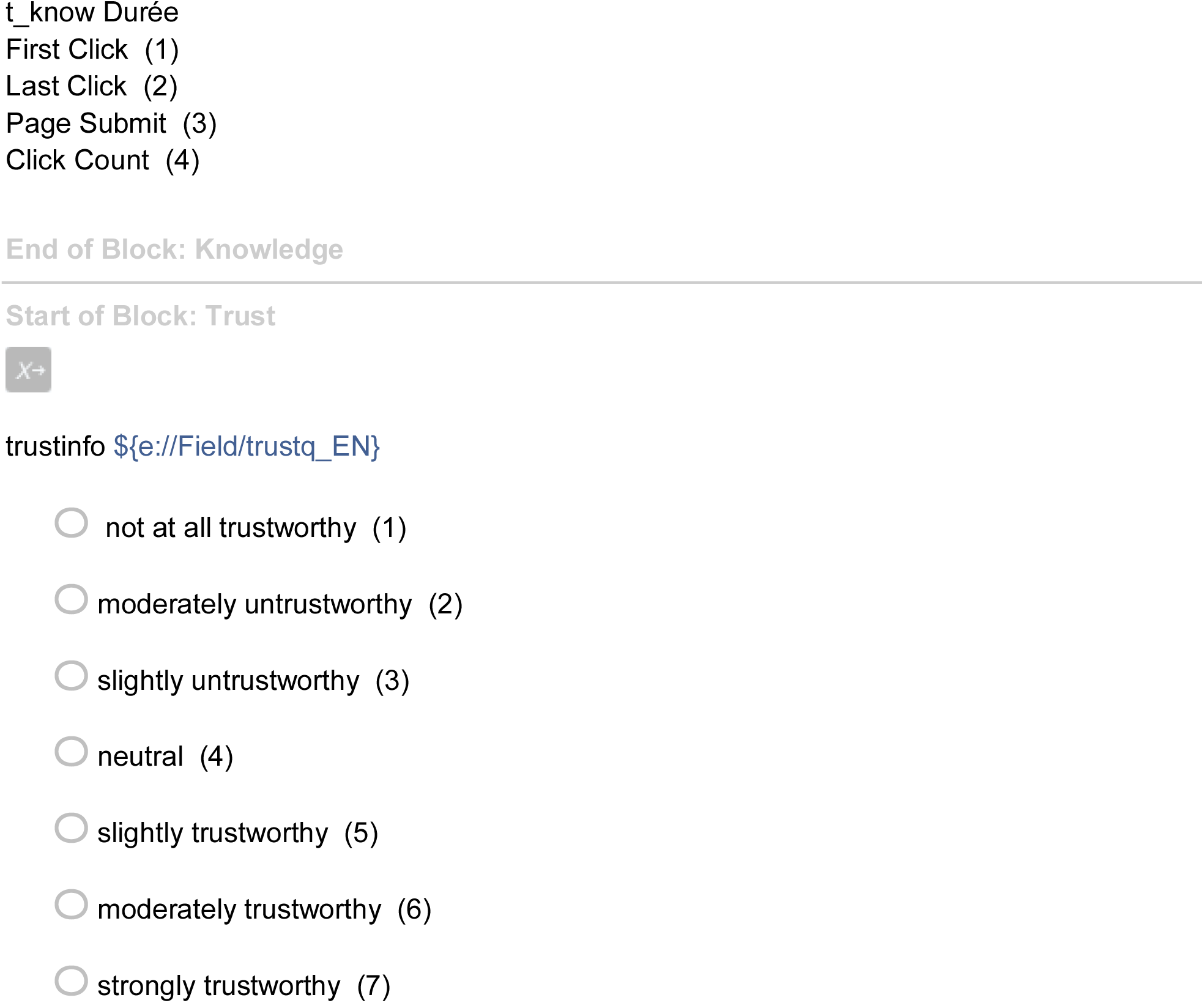
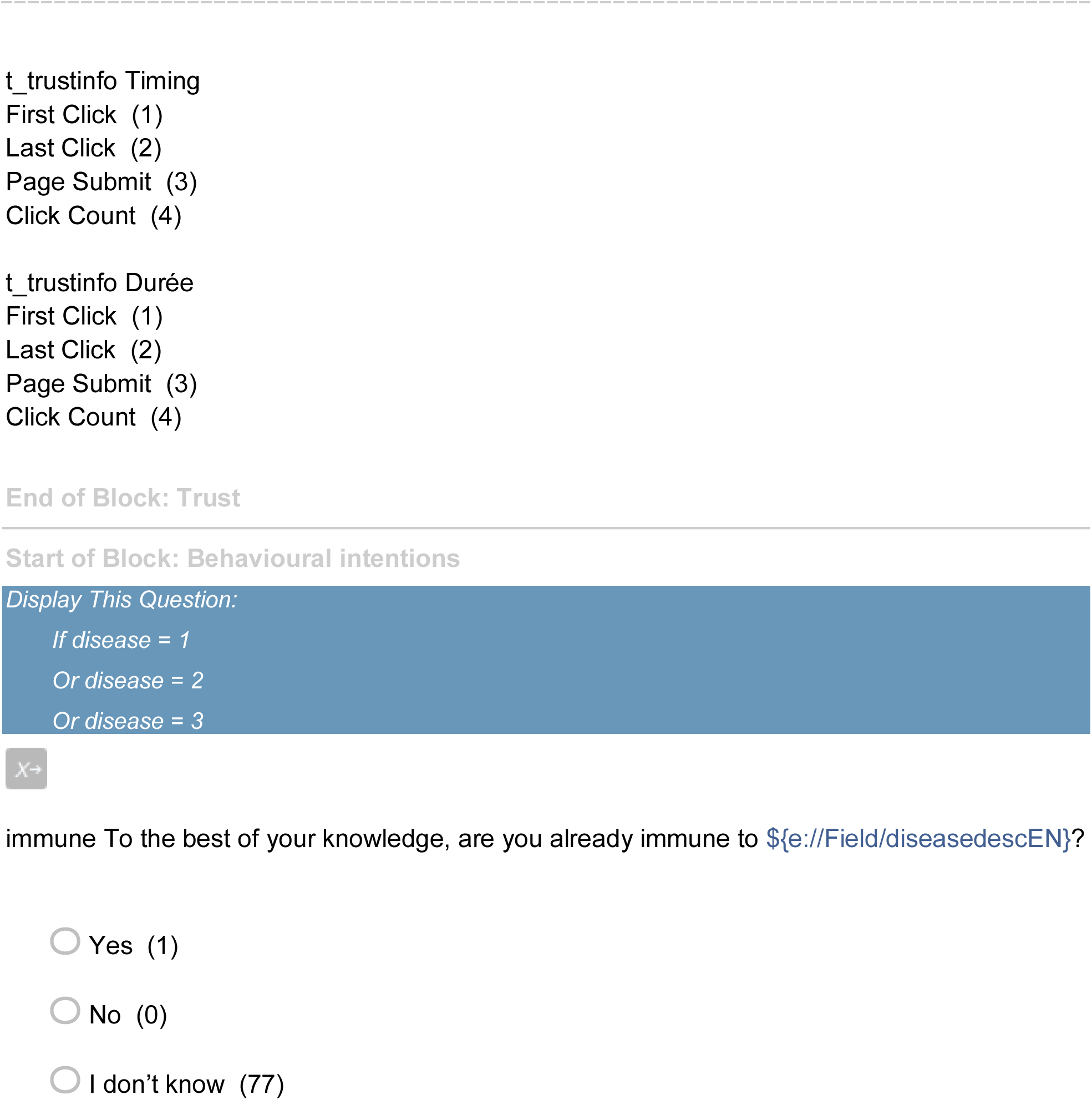
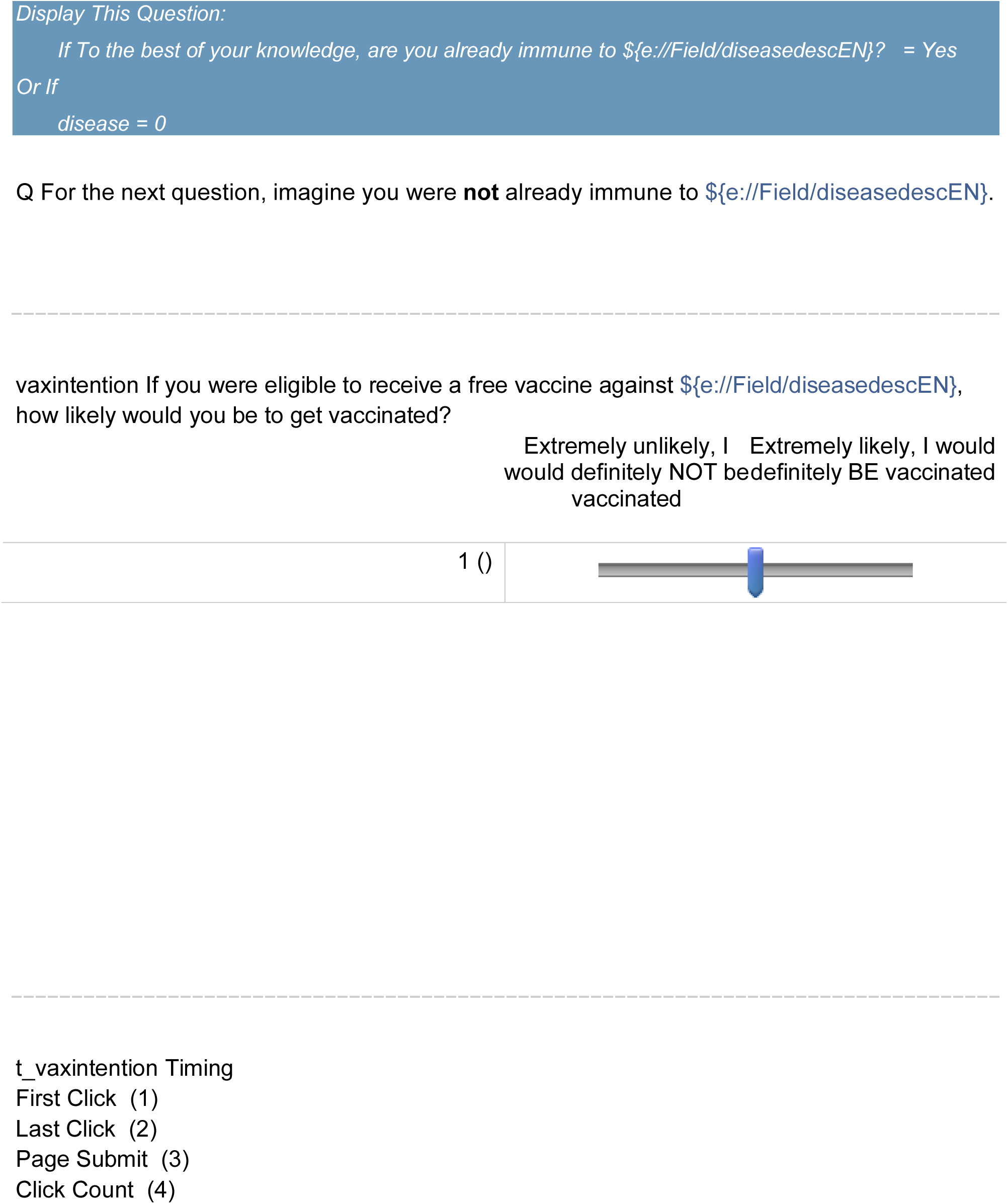
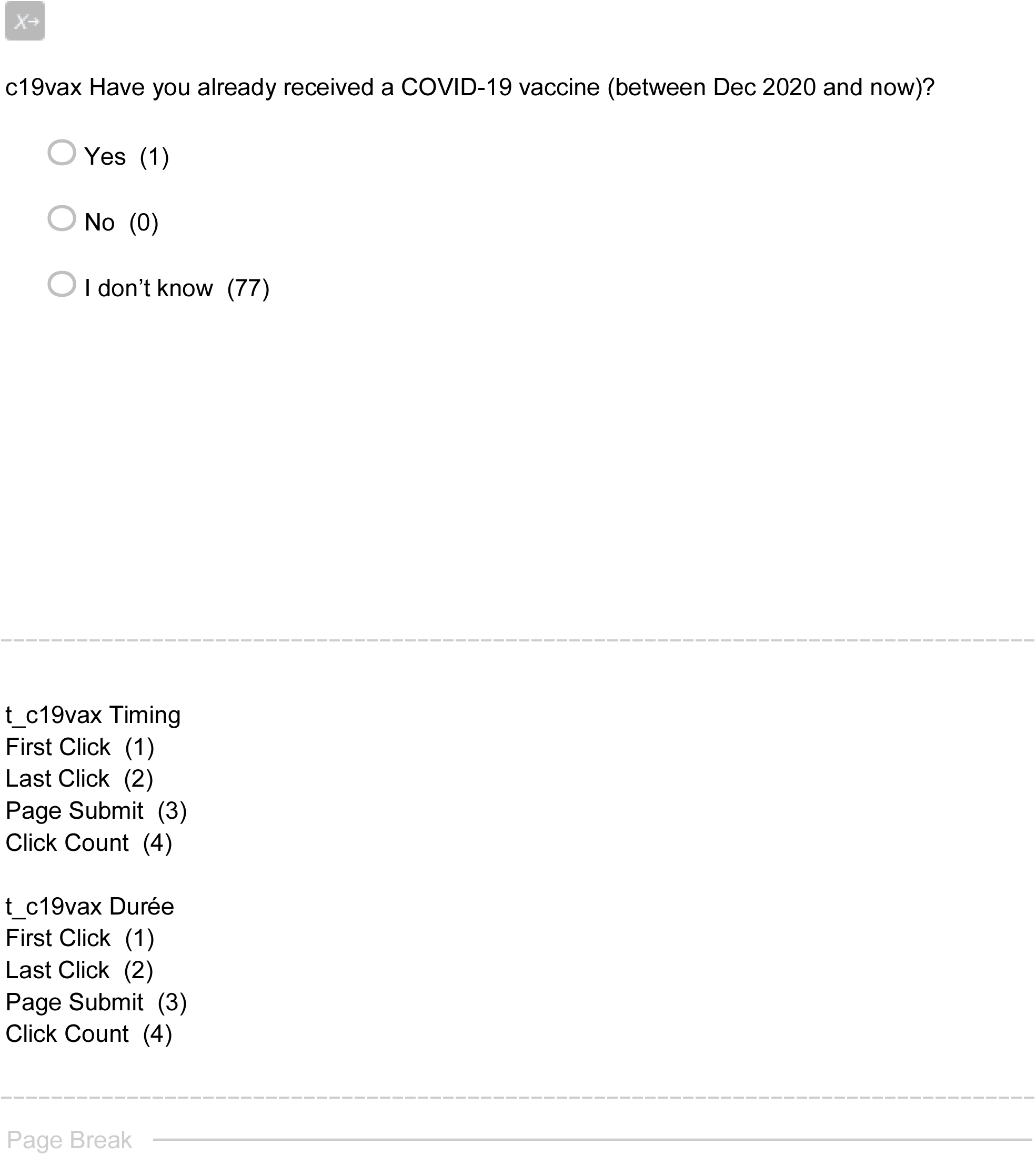
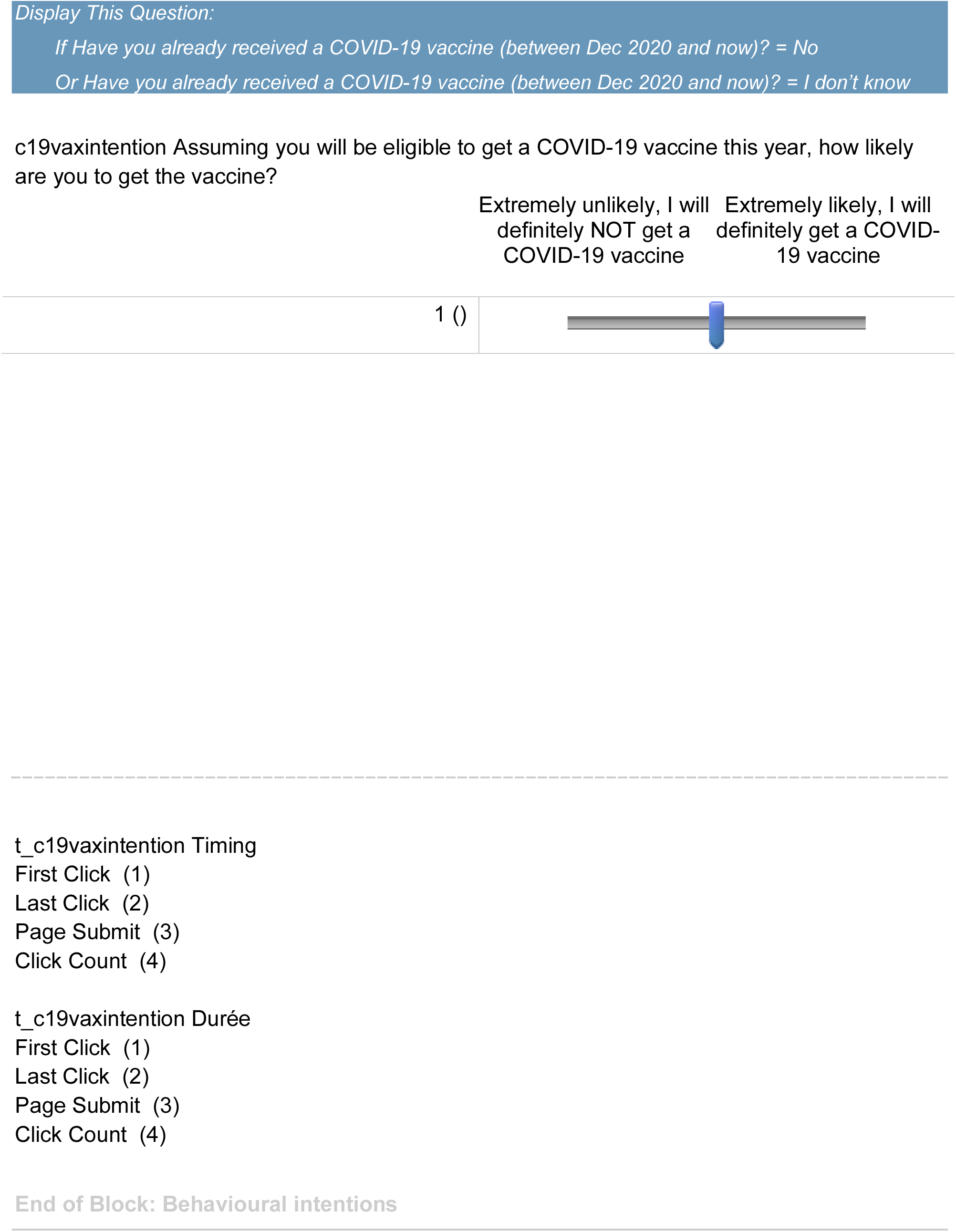

## Appendix 4: Descriptive analysis per arm

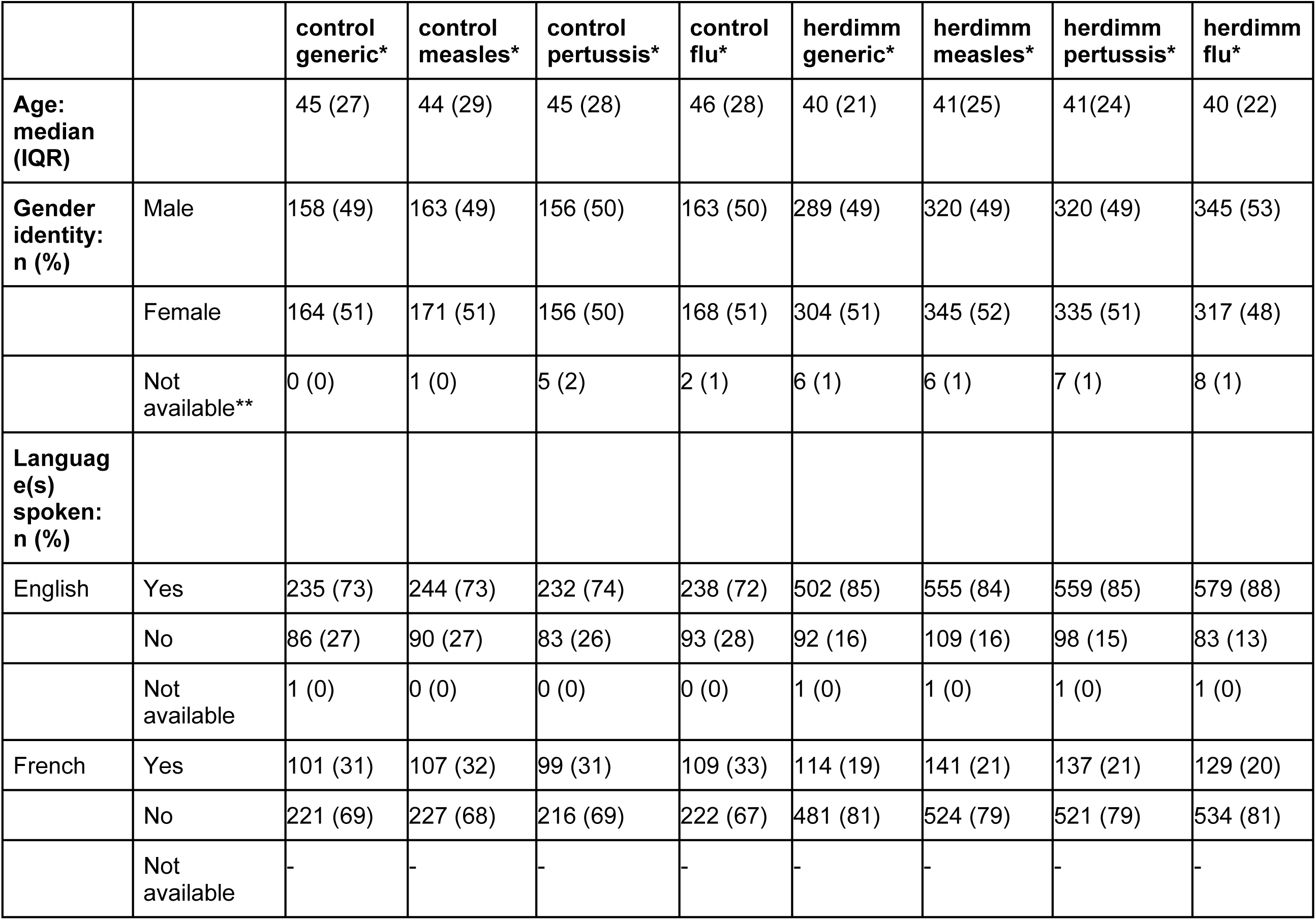
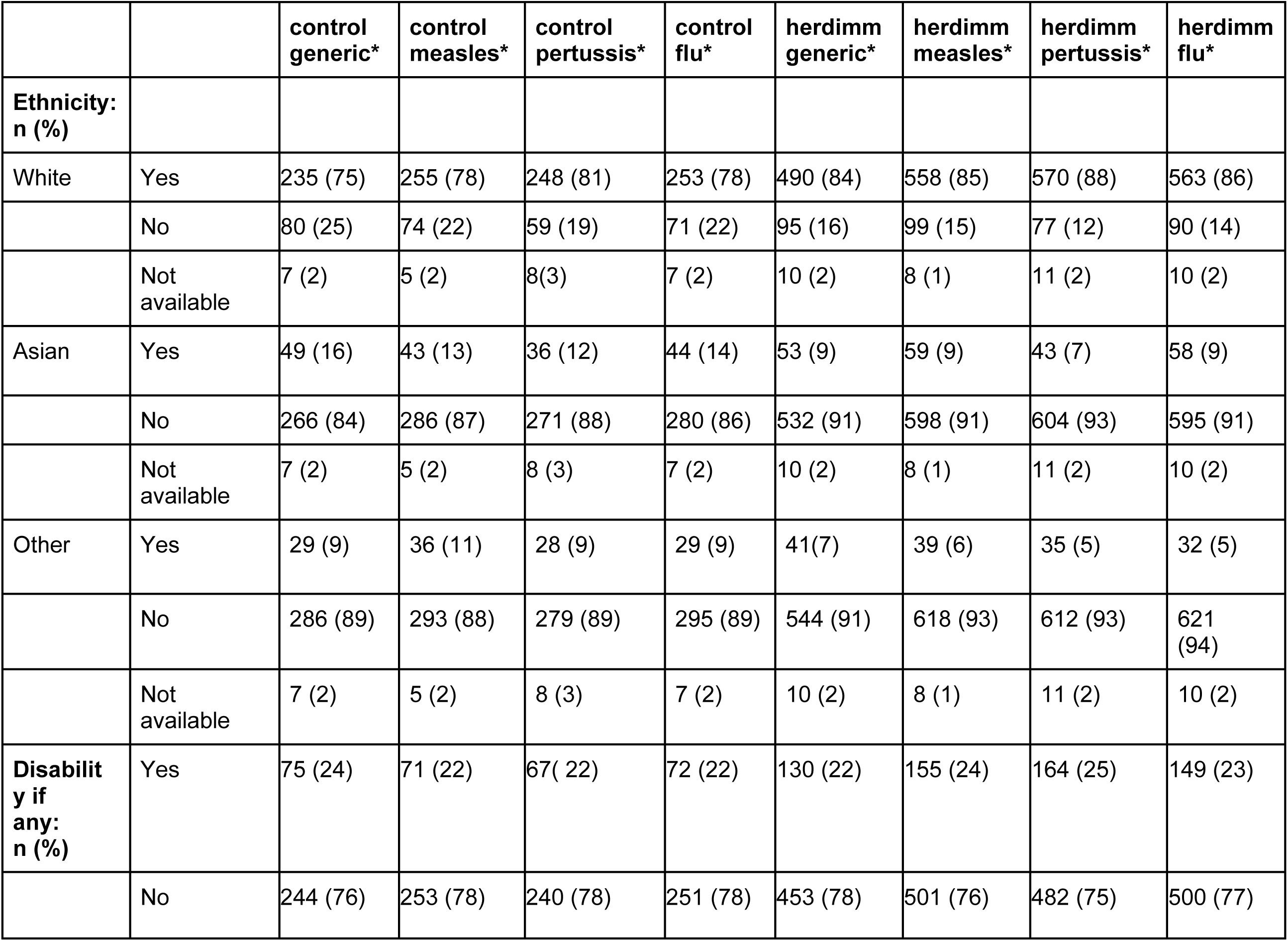
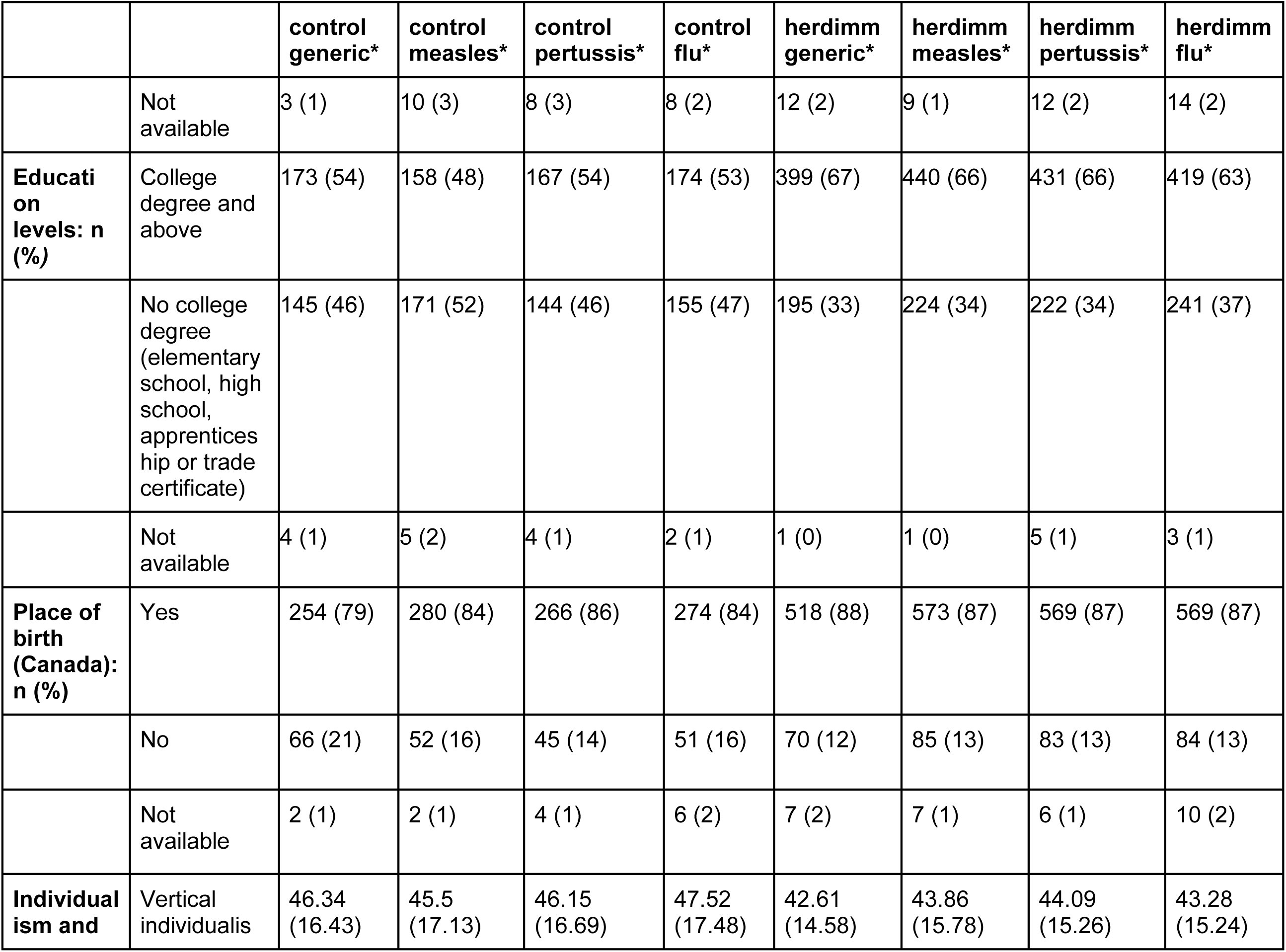
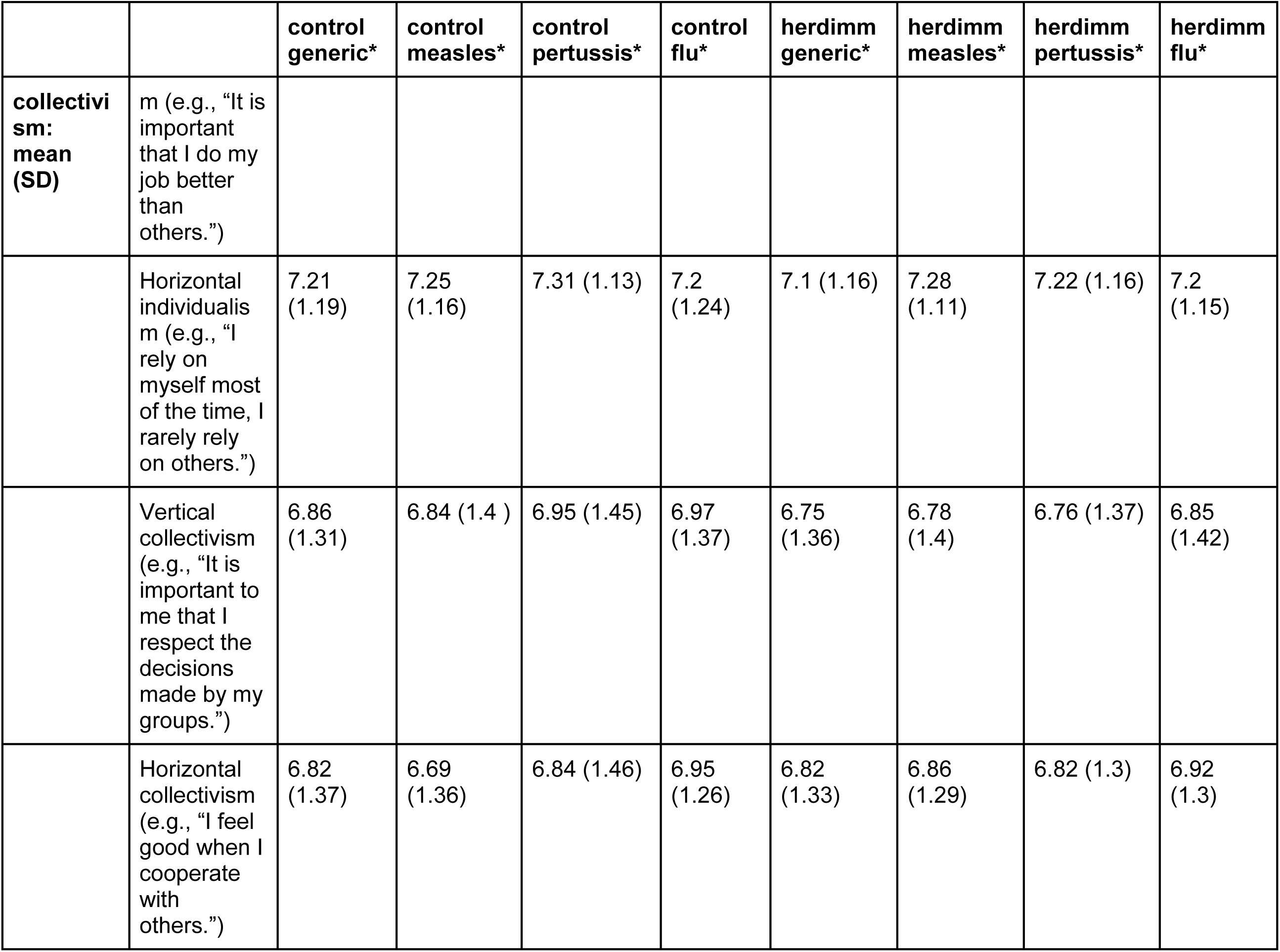
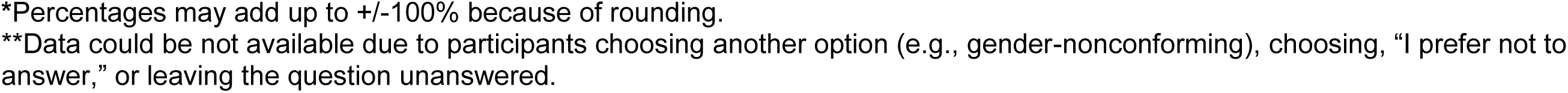

## Appendix 5: Complete results of models

| Non Descriptive labels | labels |
| --- | --- |
| disease1 | Measles |
| disease2 | Pertussis |
| disease3 | Flu |
| viz1 | herdimm |
| language_1 | English |
| language_2 | French |
| edhi | We recoded this covariate as college degree and above and no college degree (elementary school, high school diploma, or apprenticeship or trade certificate or diploma.) |
| mean_indivhorz | Mean horizontal individualism |
| mean_indivvertical | Mean vertical individualism |
| mean_collhorz | Mean horizontal collectivism |
| mean_collvertical | Mean vertical collectivism |

### Risk perception 1 (Objective risk perception)

#### Two-way

##### Model 1: Check for direct effects of factors without any covariates ####

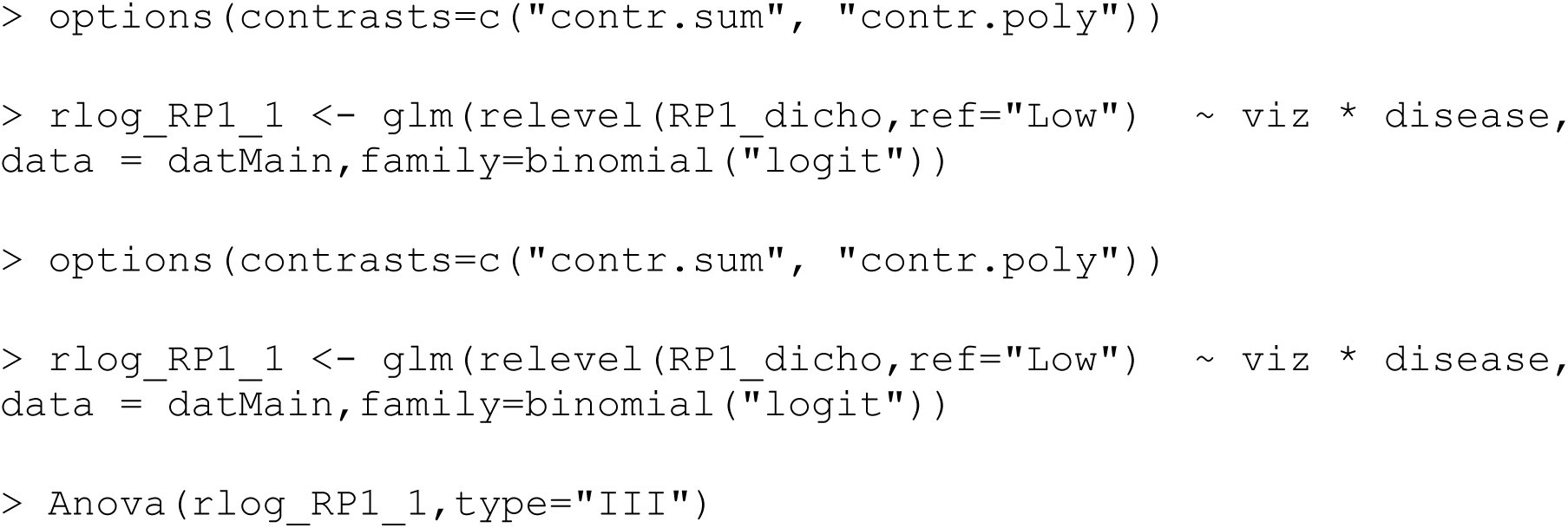
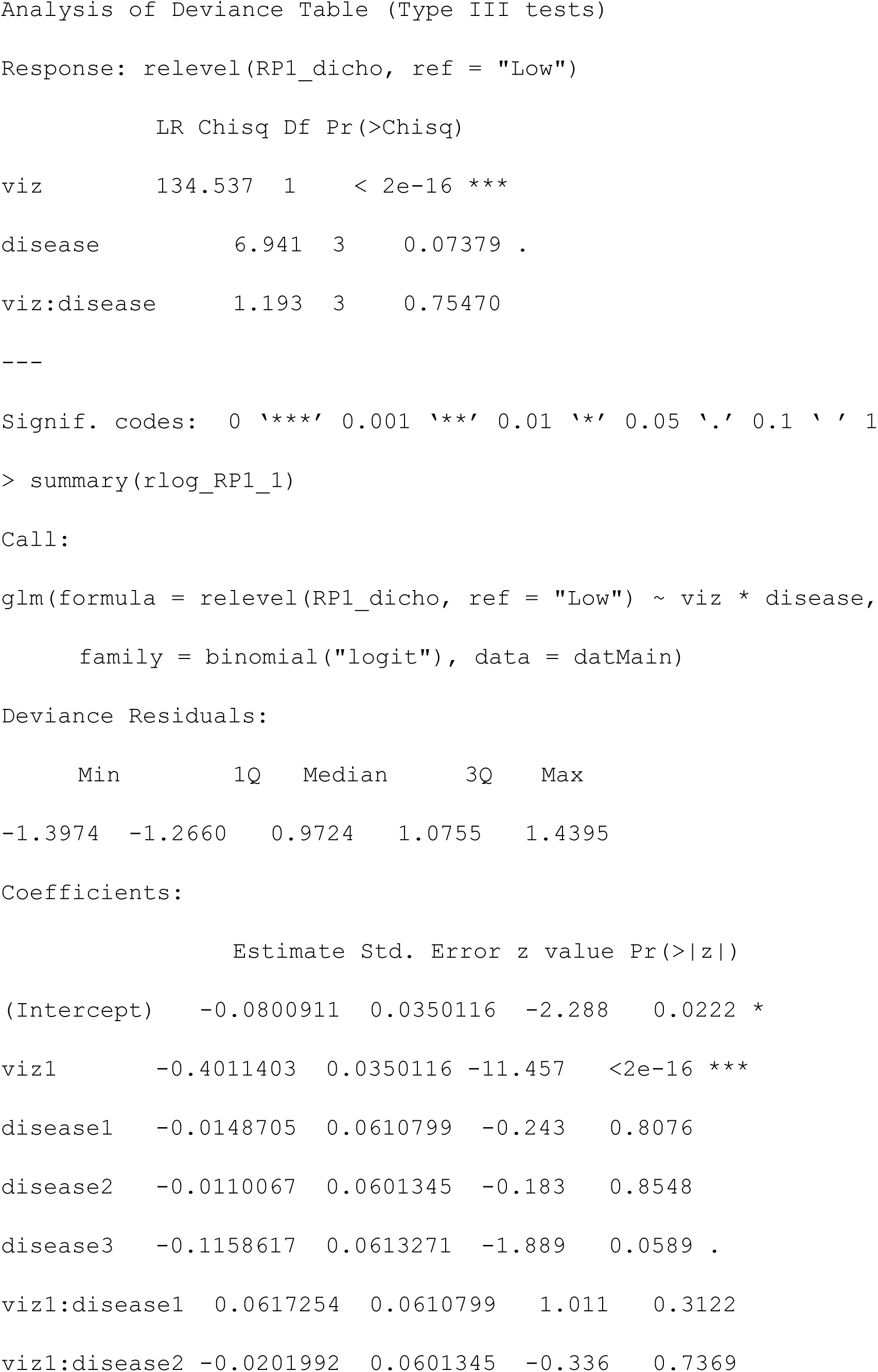
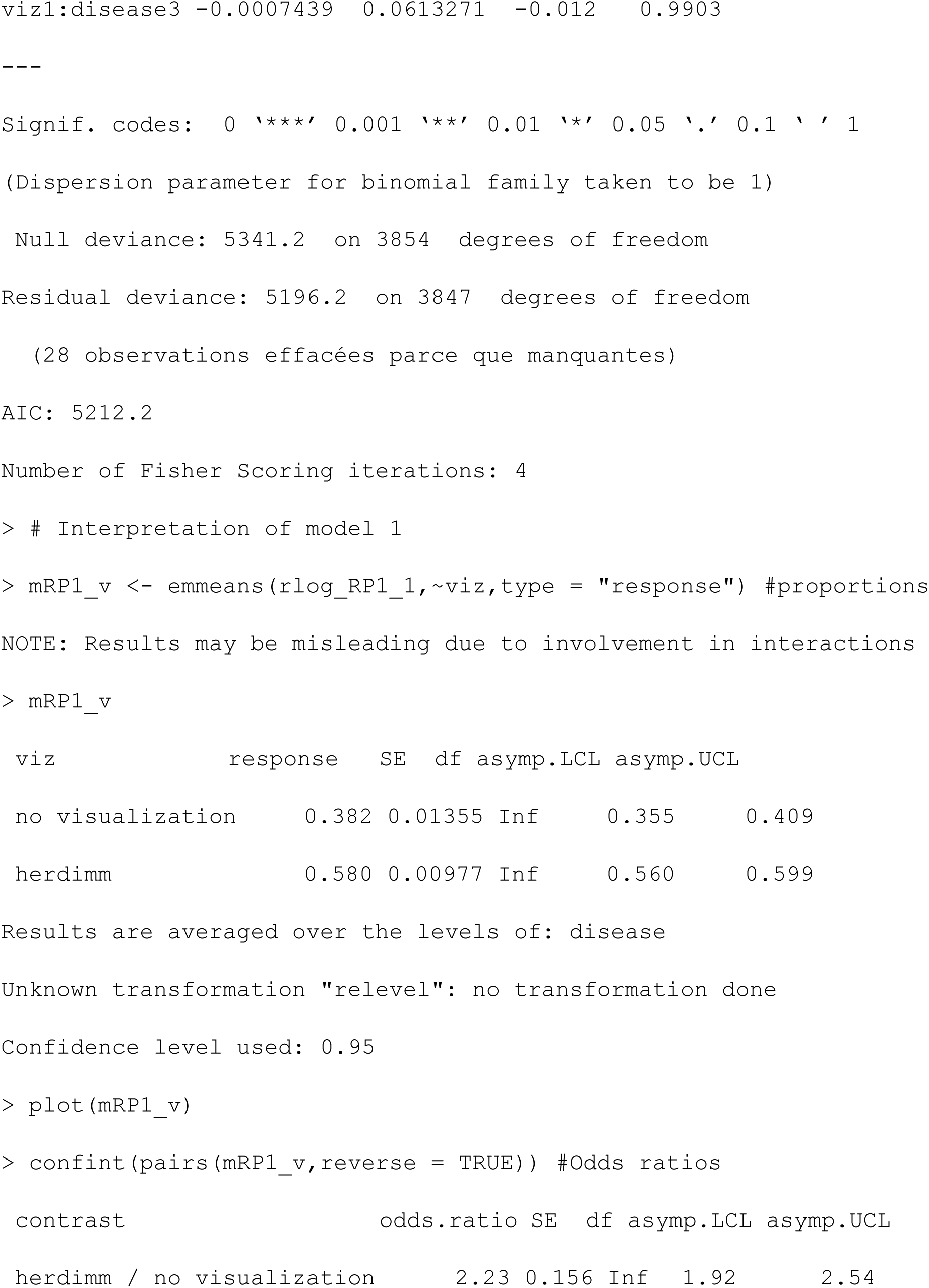
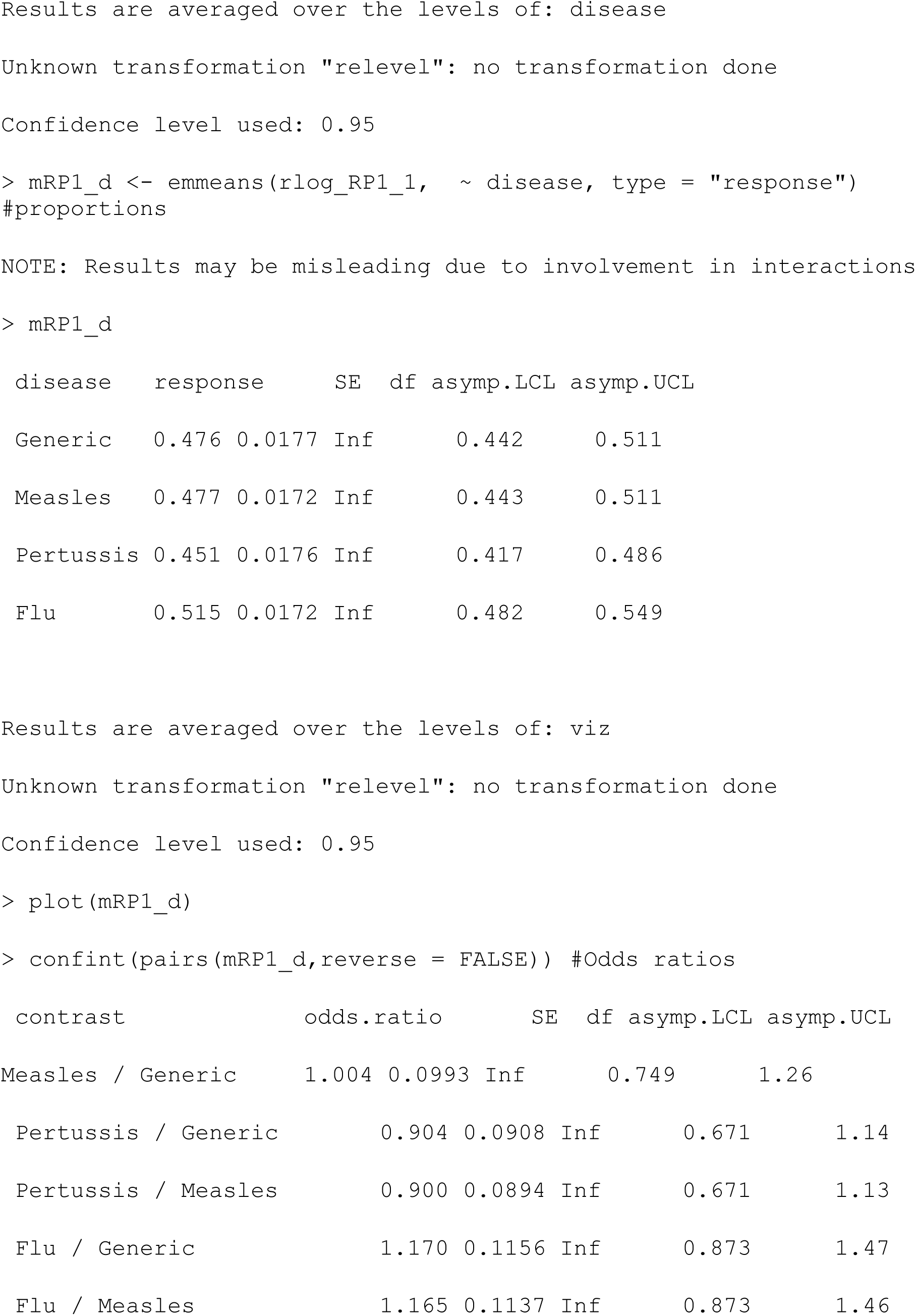
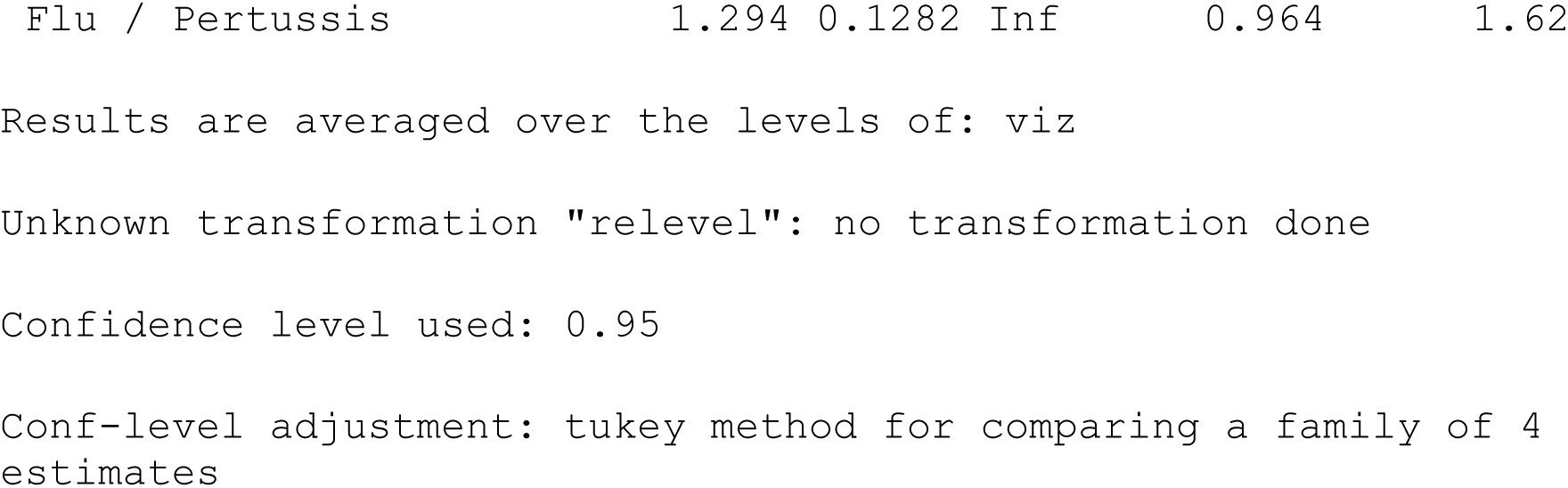

##### Model 2: Check for direct effects of factors with adjustment for other covariates

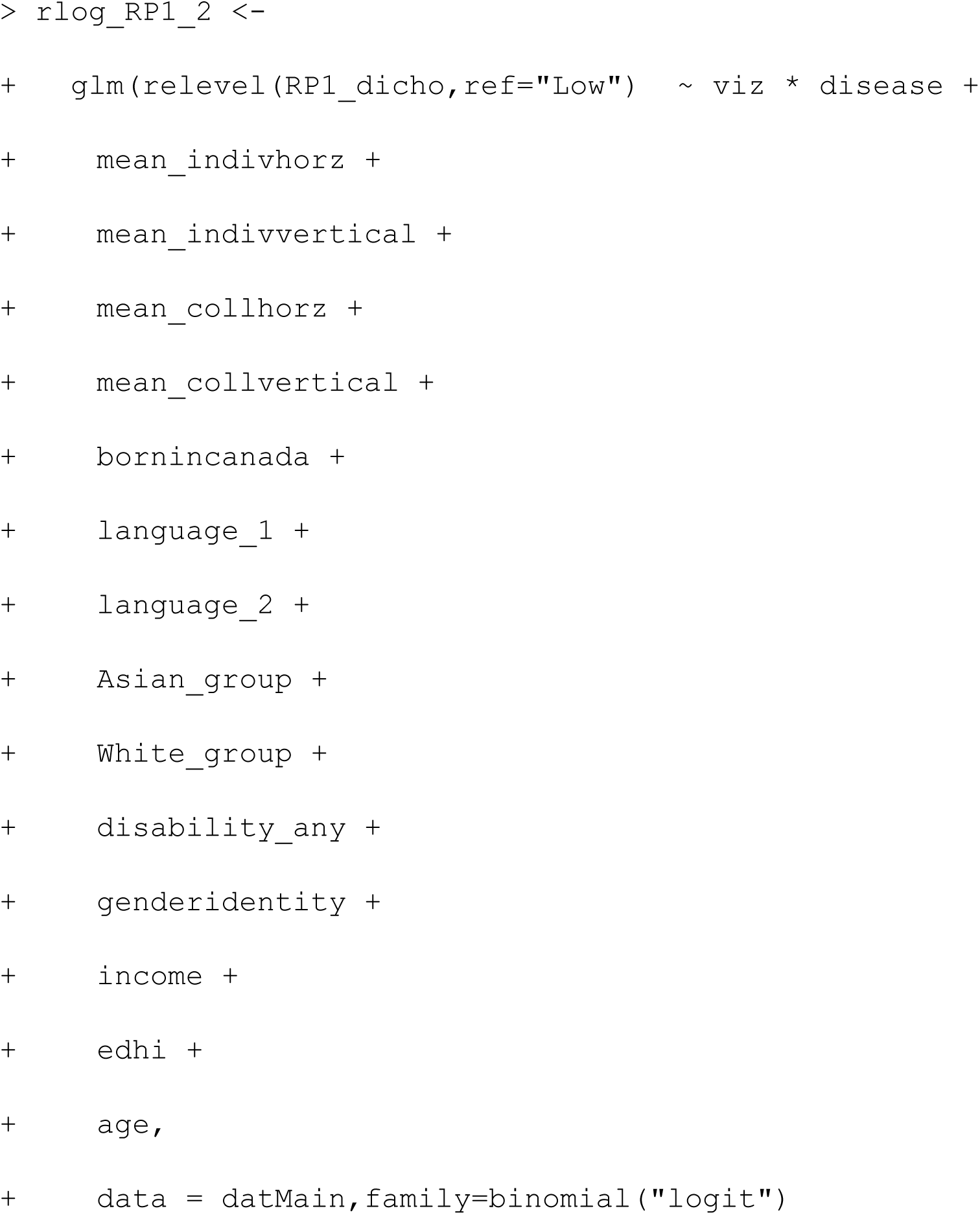
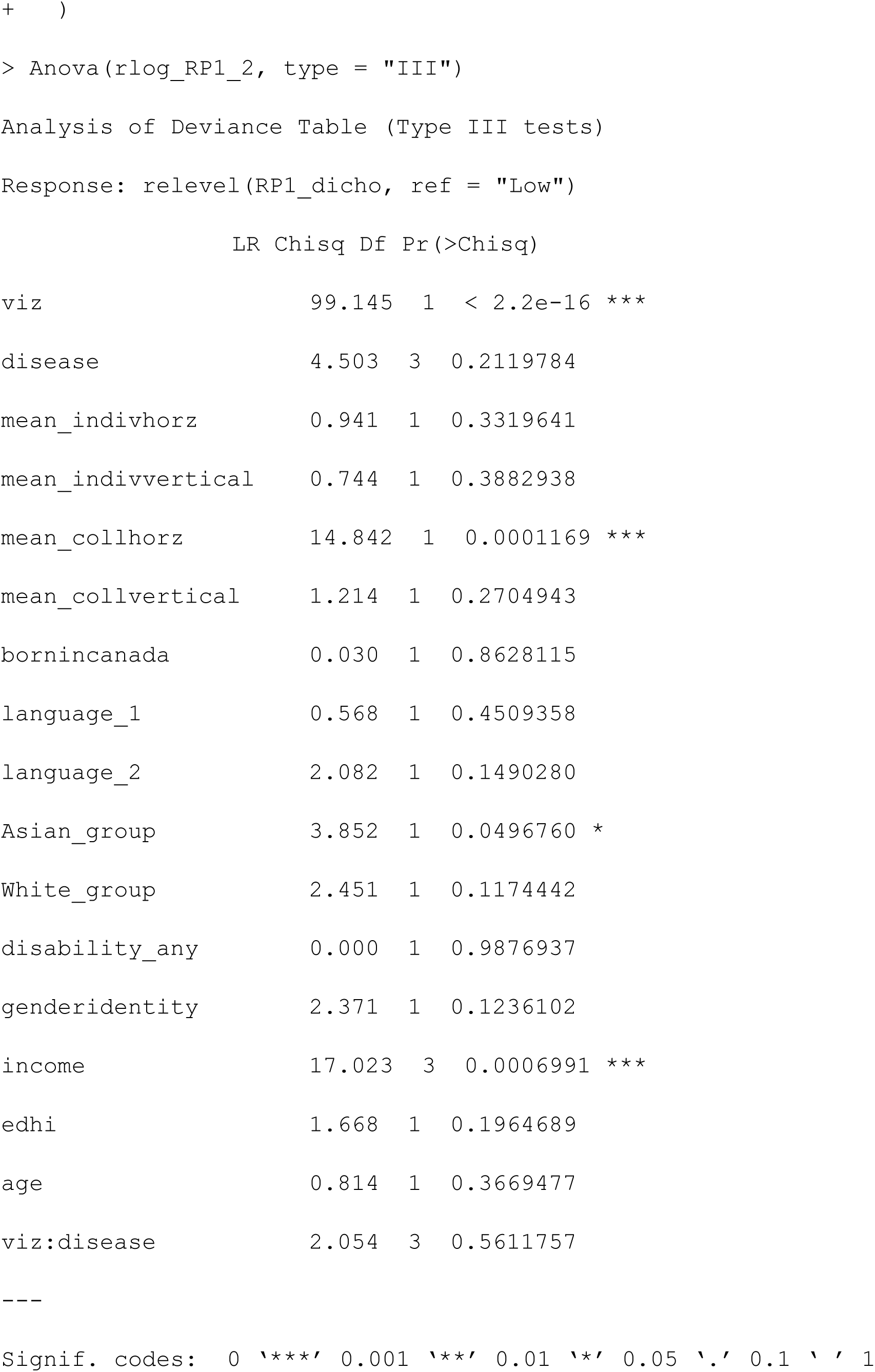
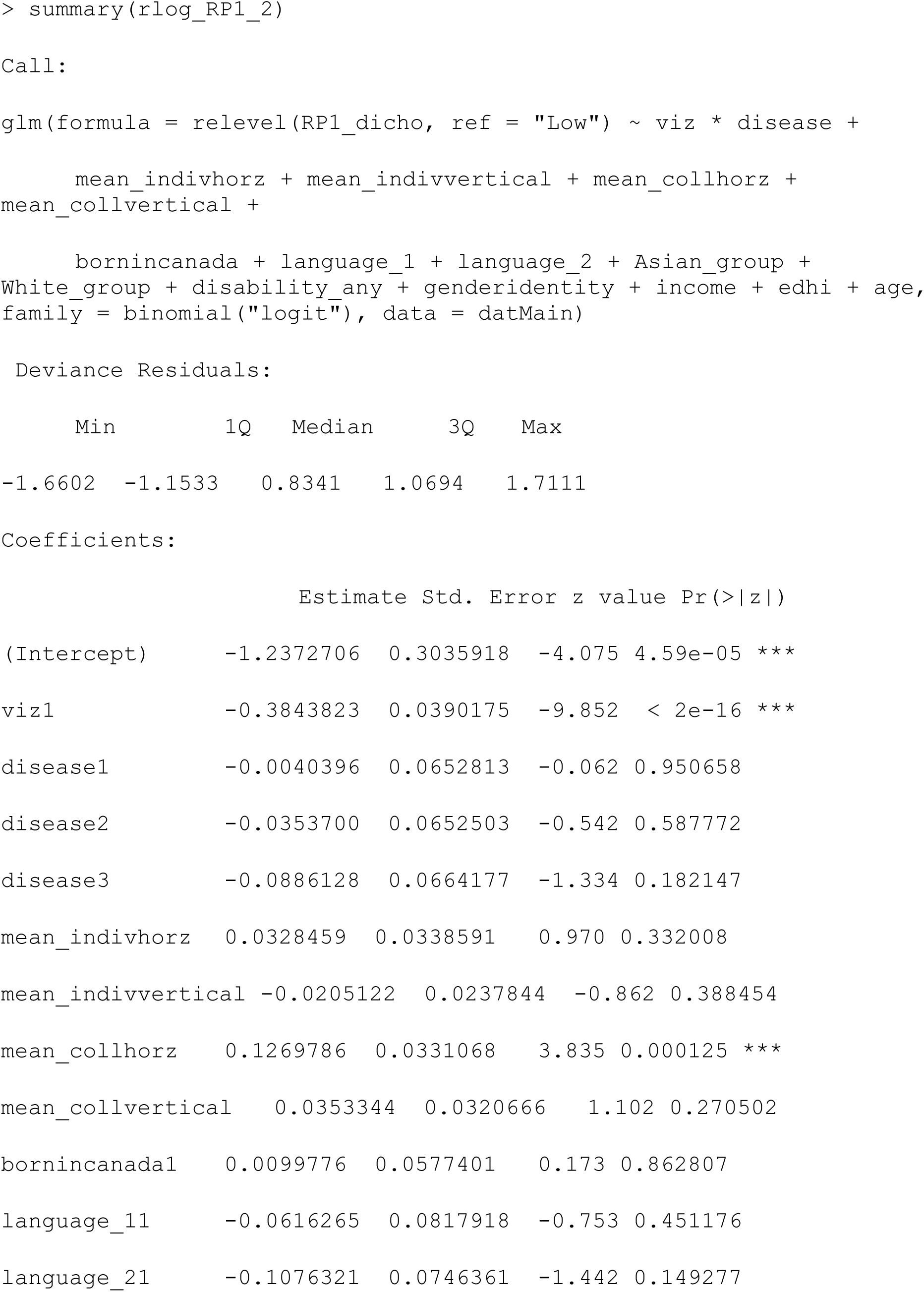
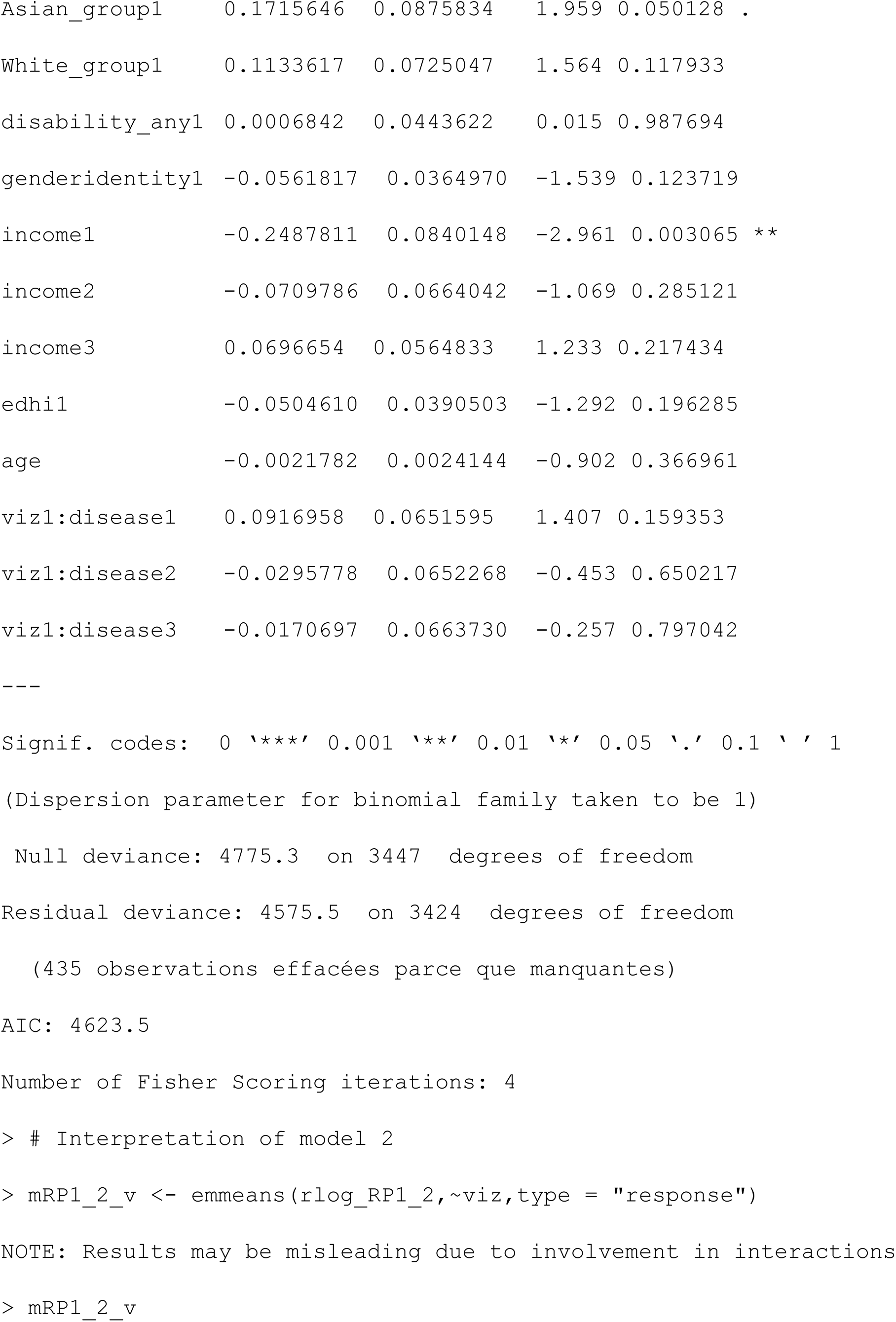
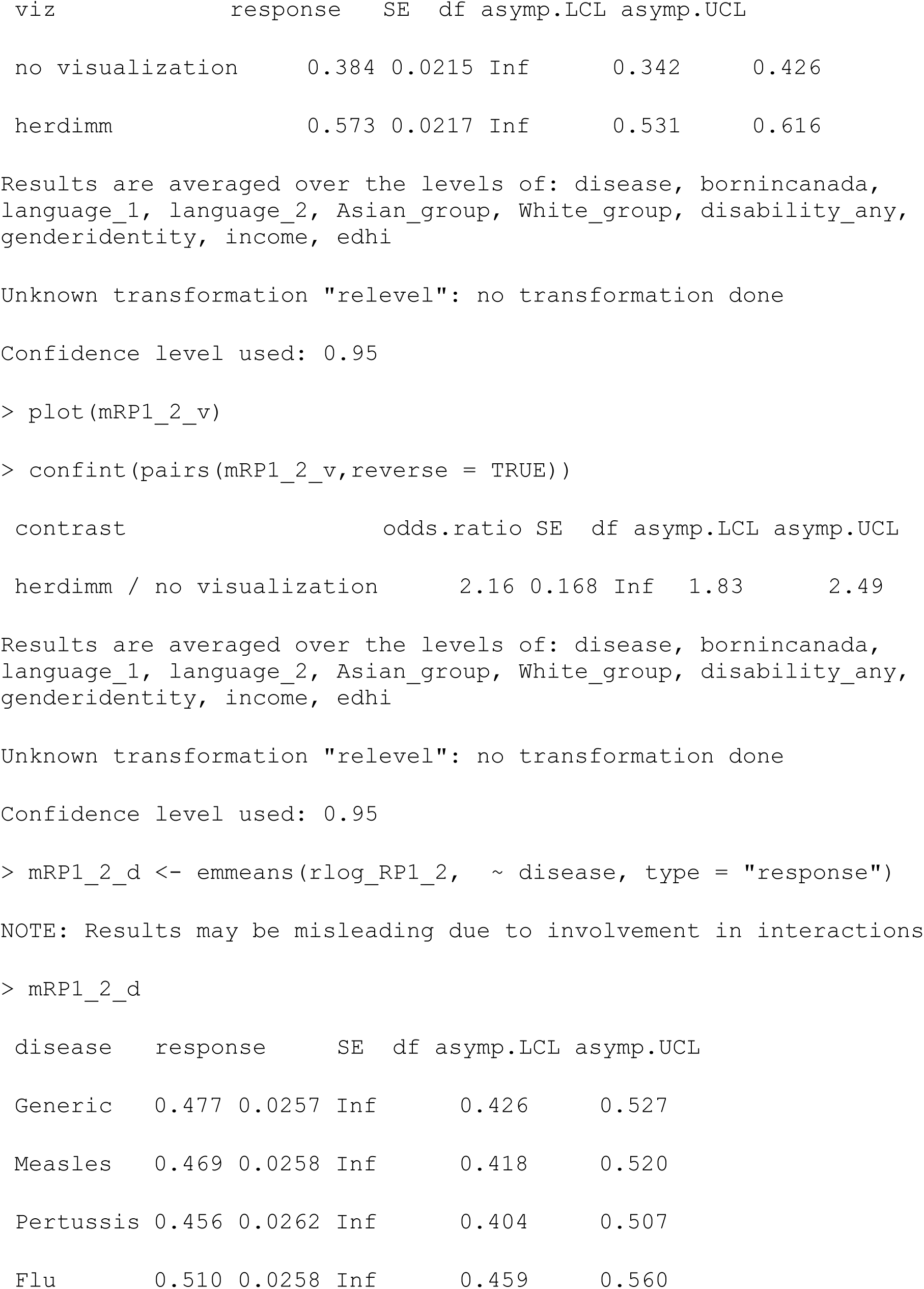
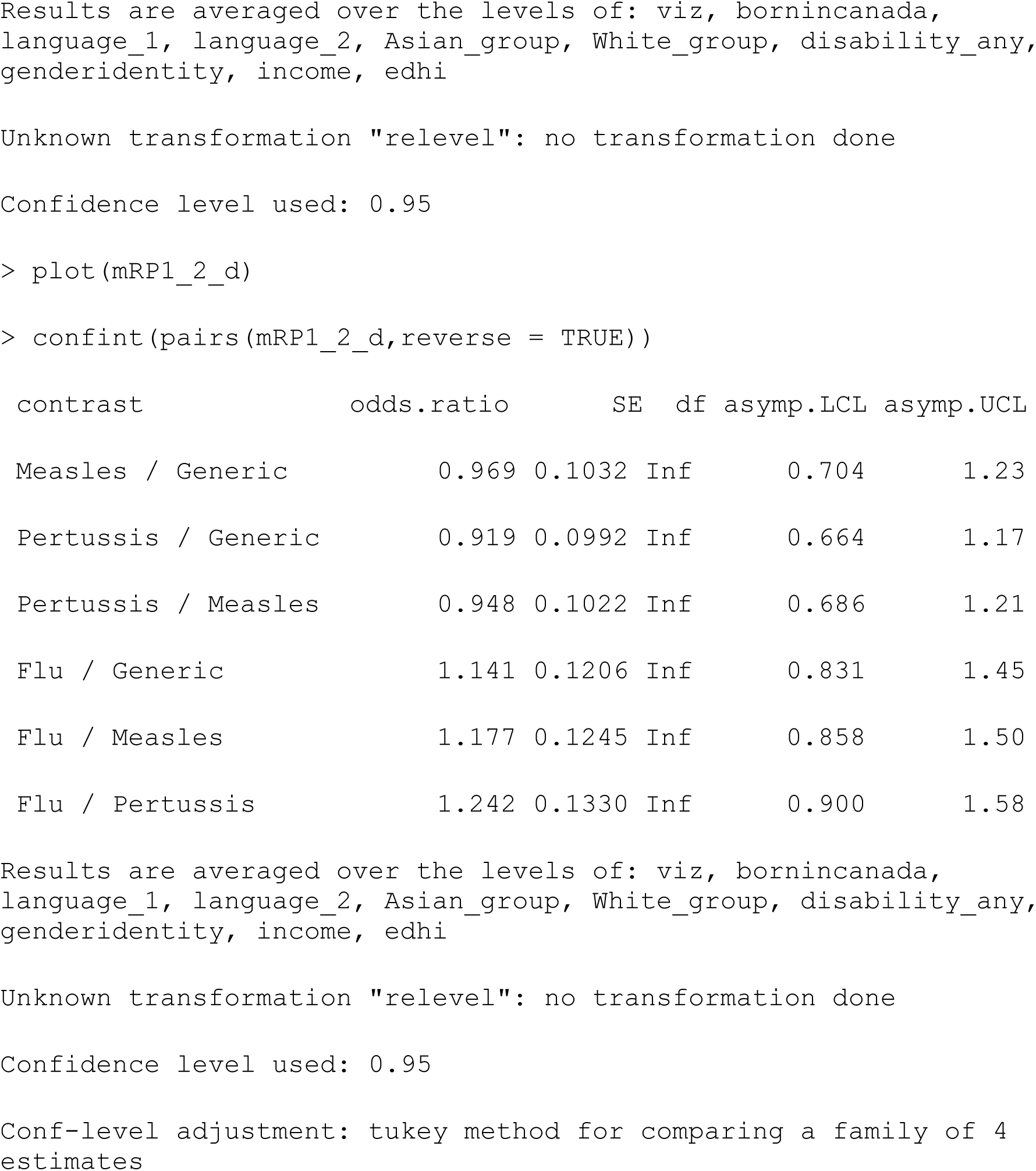

##### Model 3: Check for moderating effects of individualism & collectivism with adjustment for other covariates

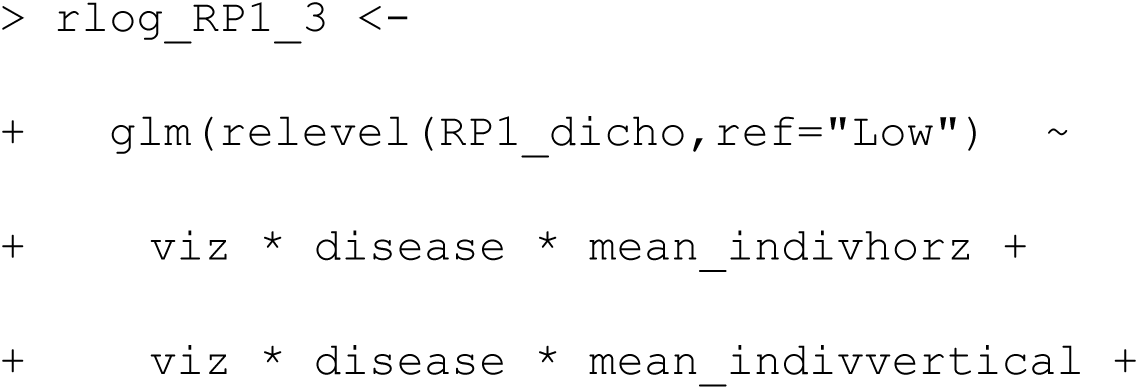
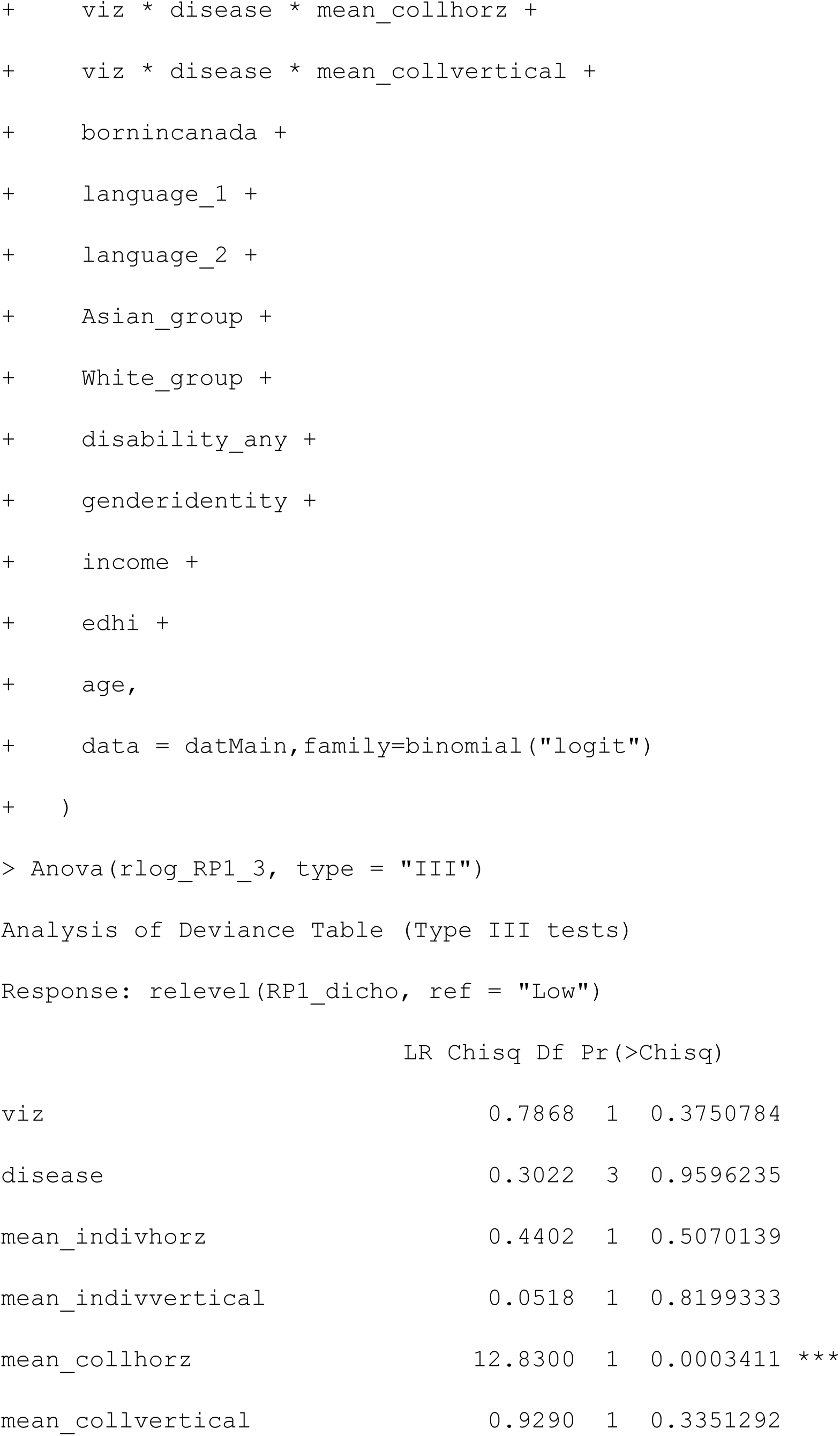
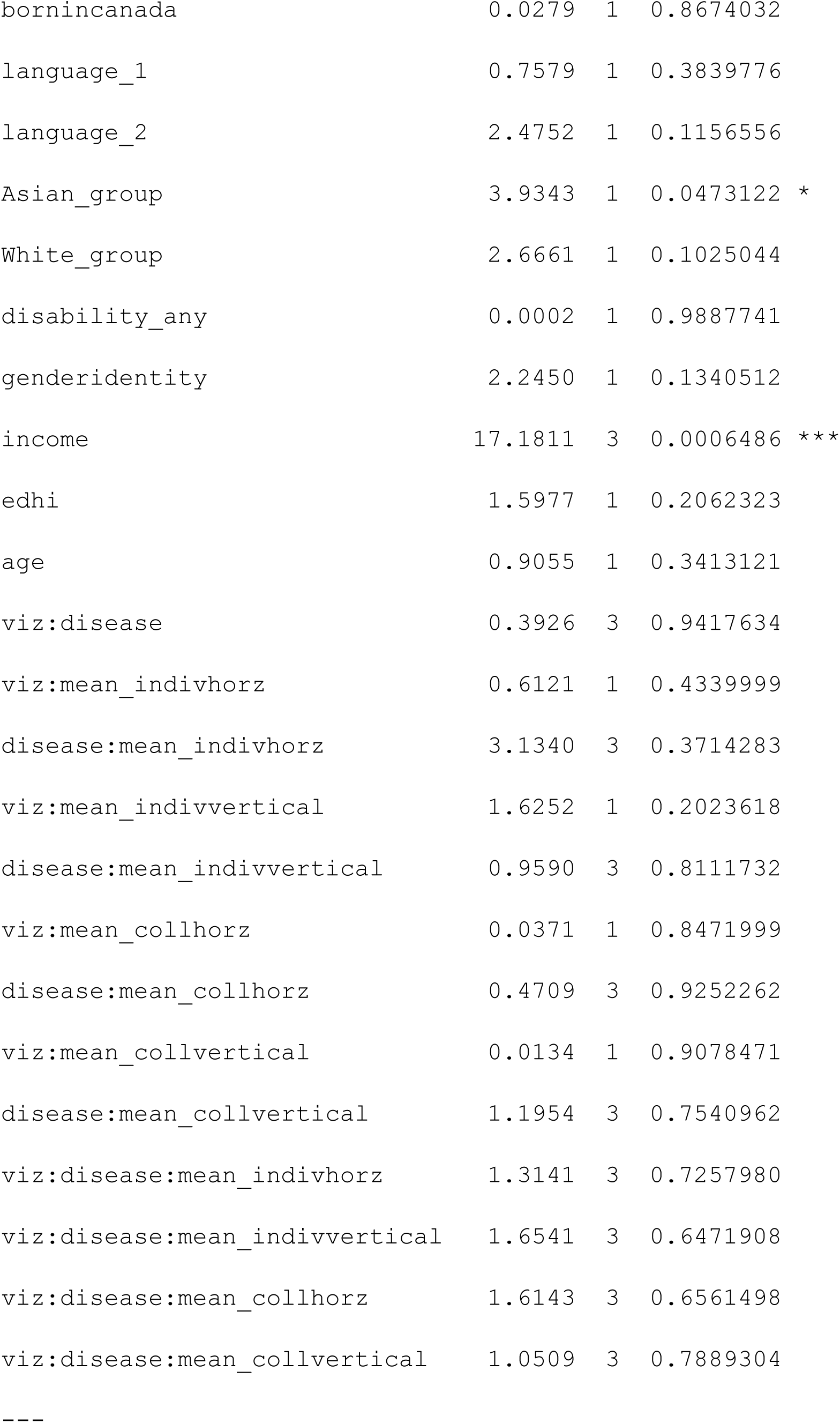
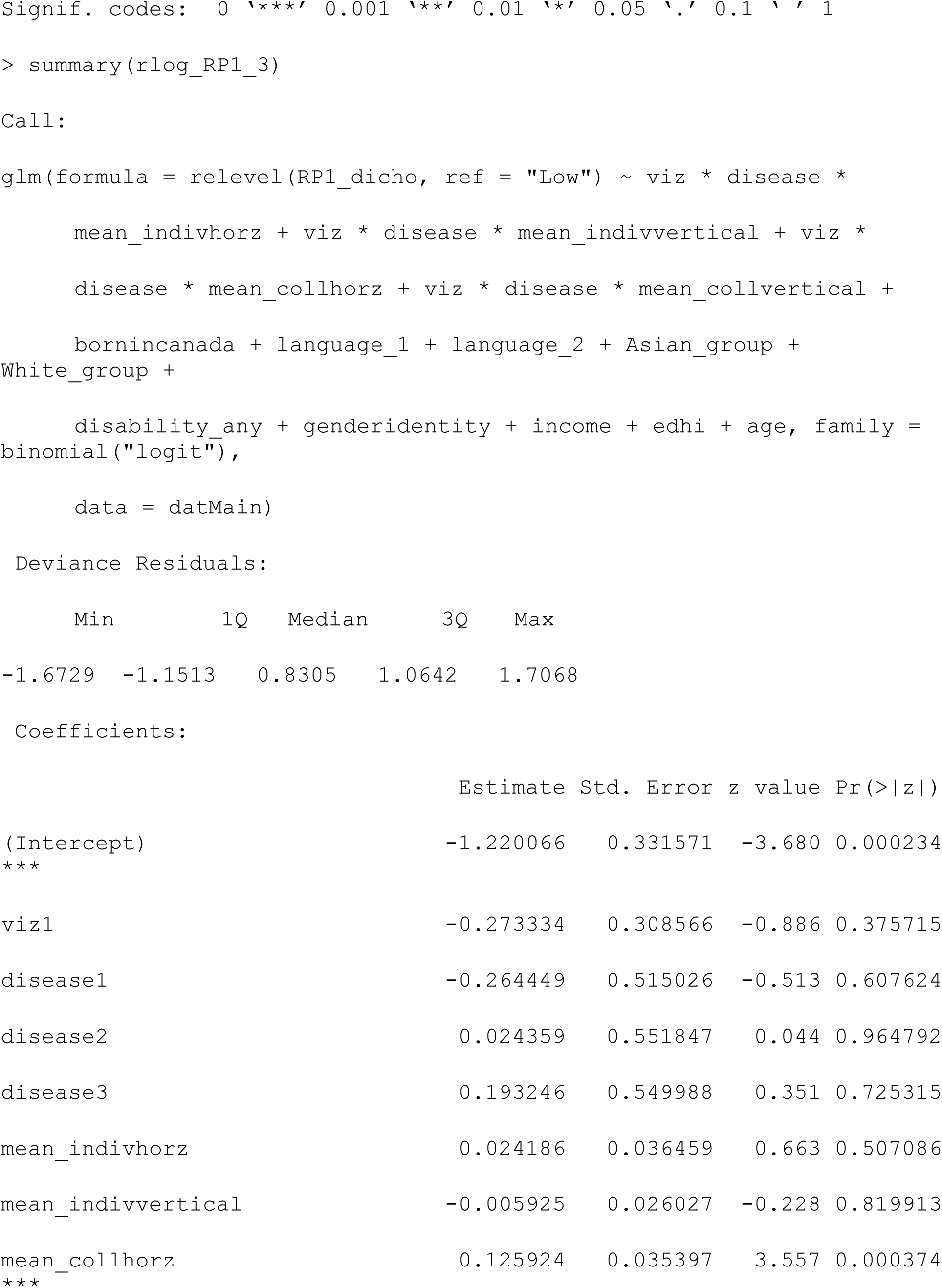
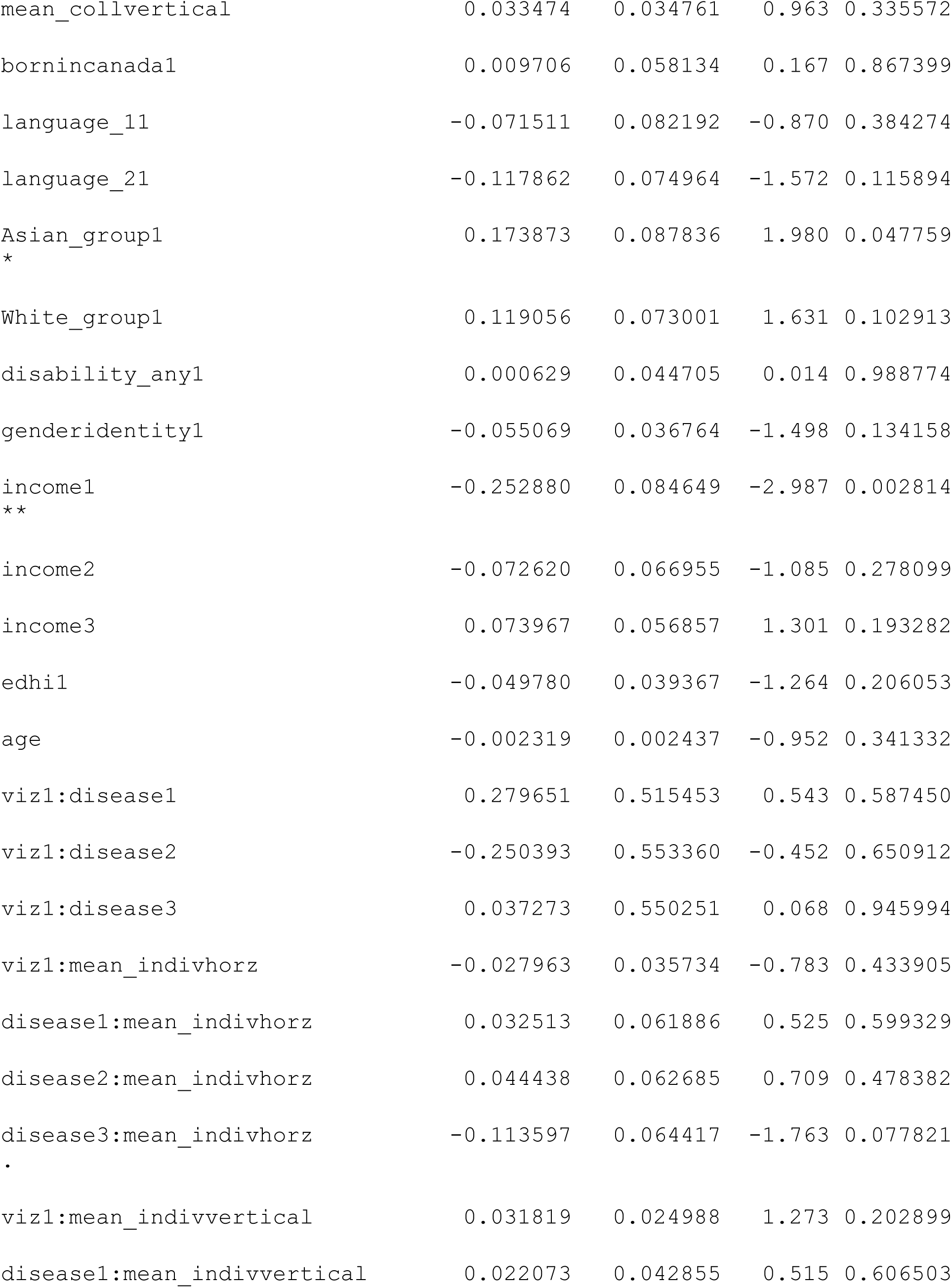
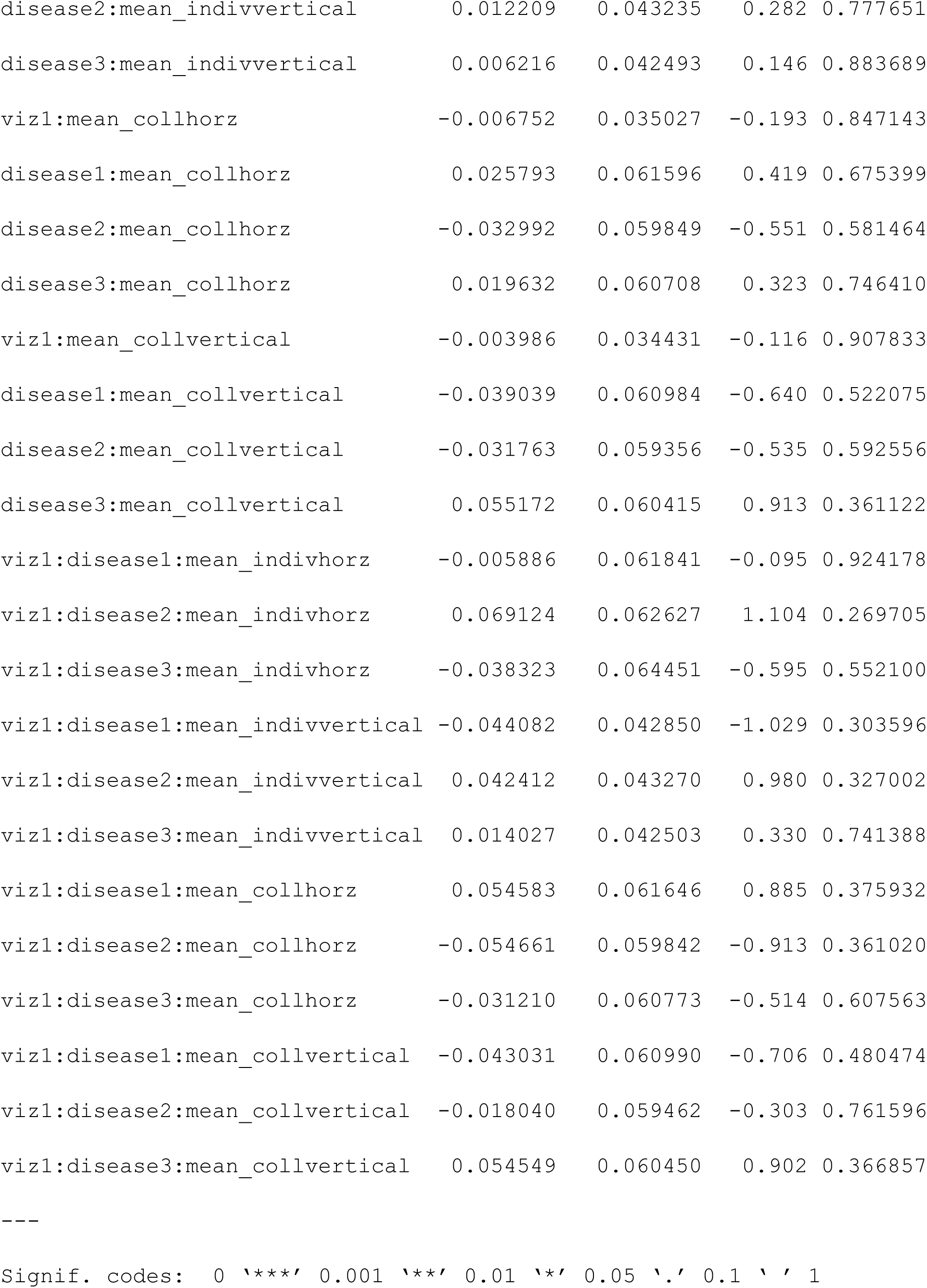
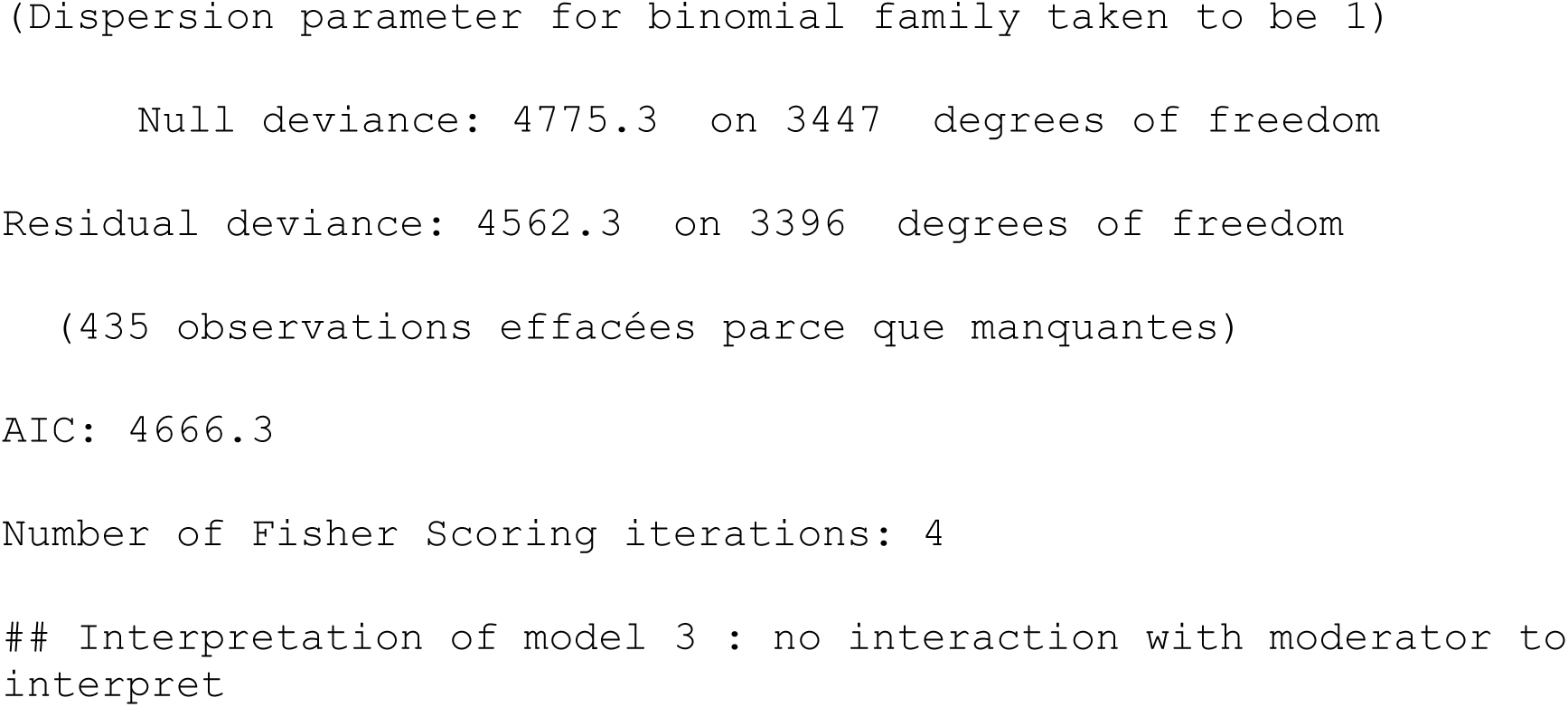

### Risk perception 2-6 (Subjective risk perception)

#### Two-way

##### Model 1: Check for direct effects of factors without any covariates (for risk perception 2 to 6) ####

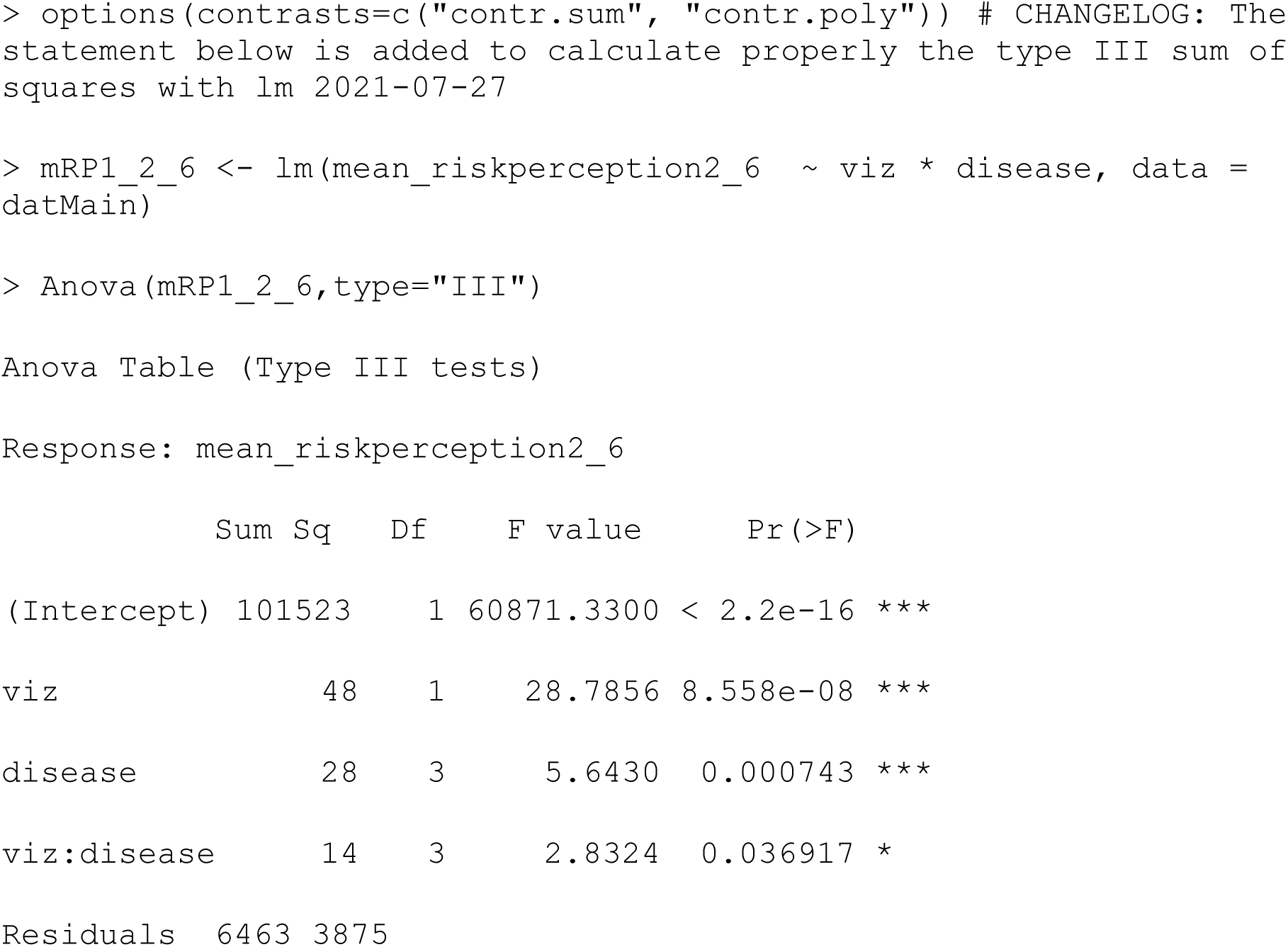
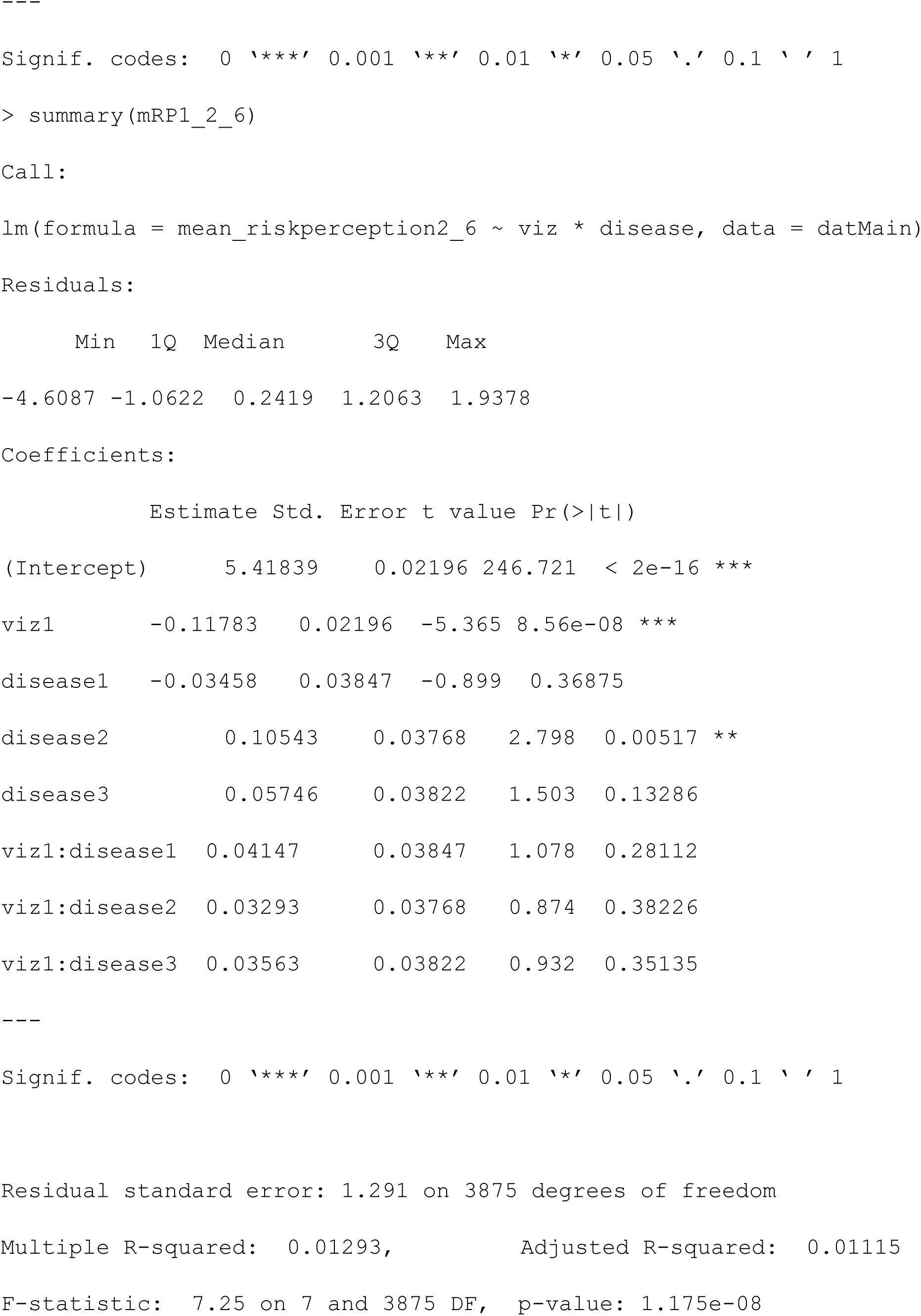
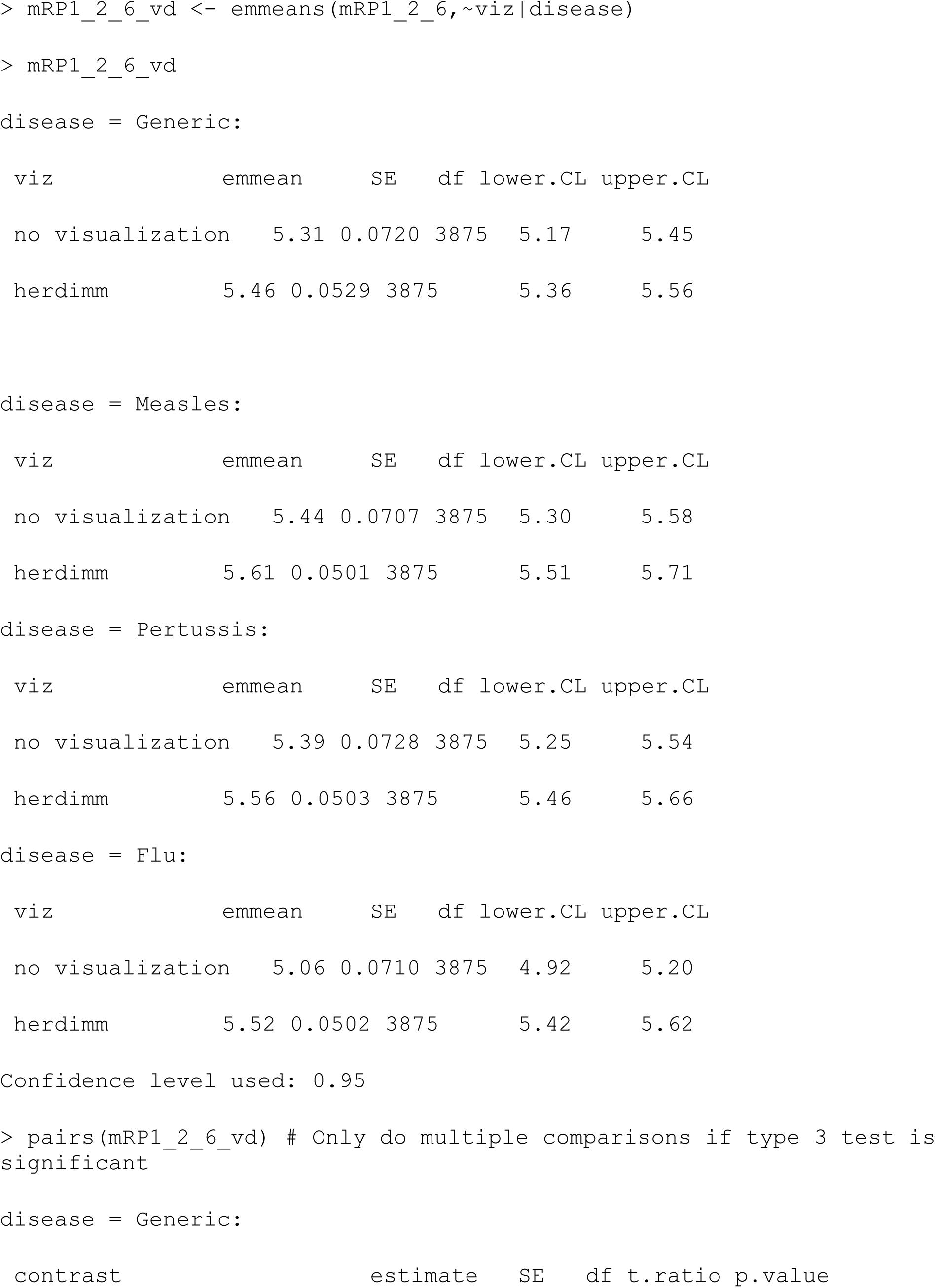
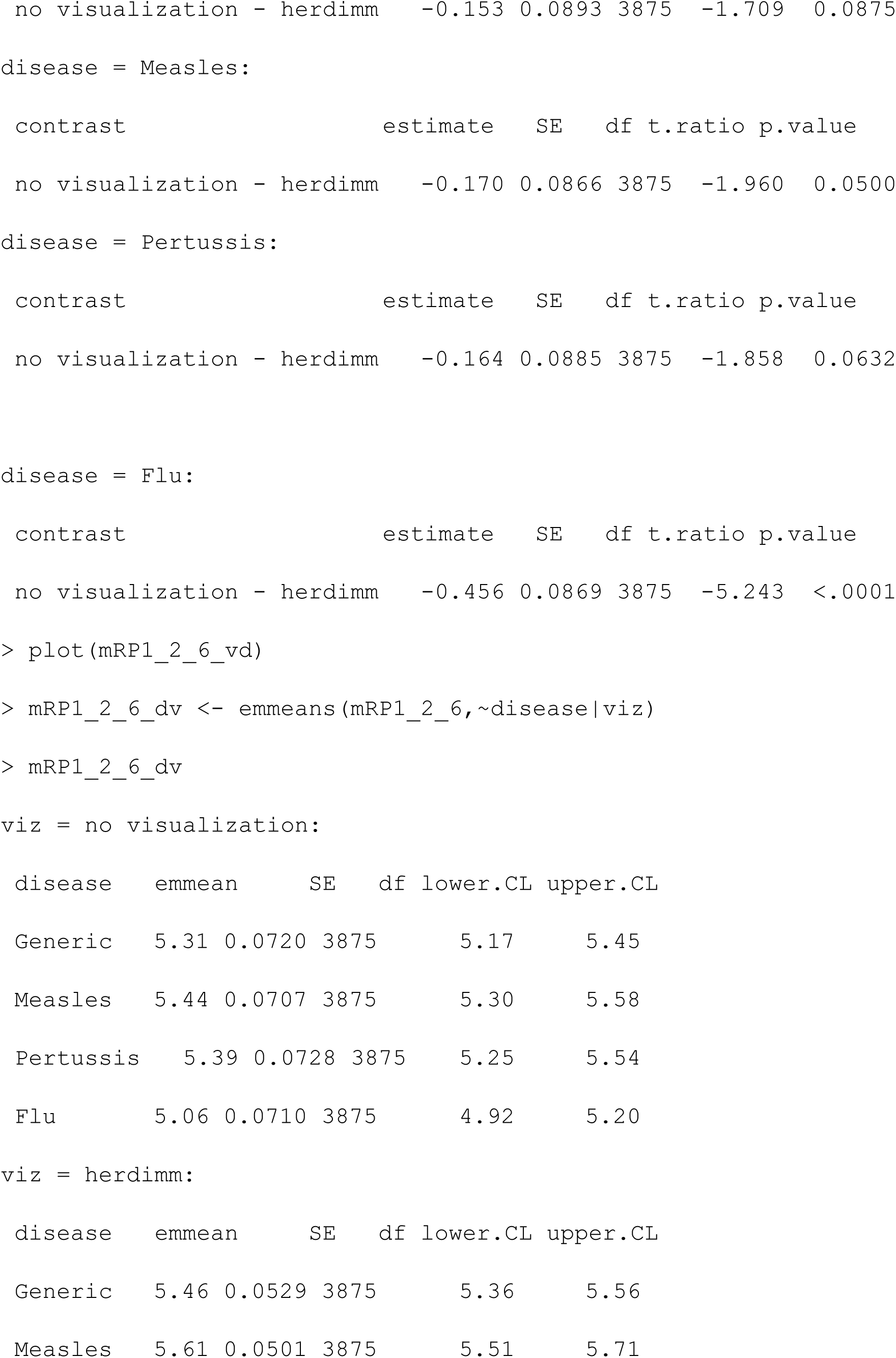
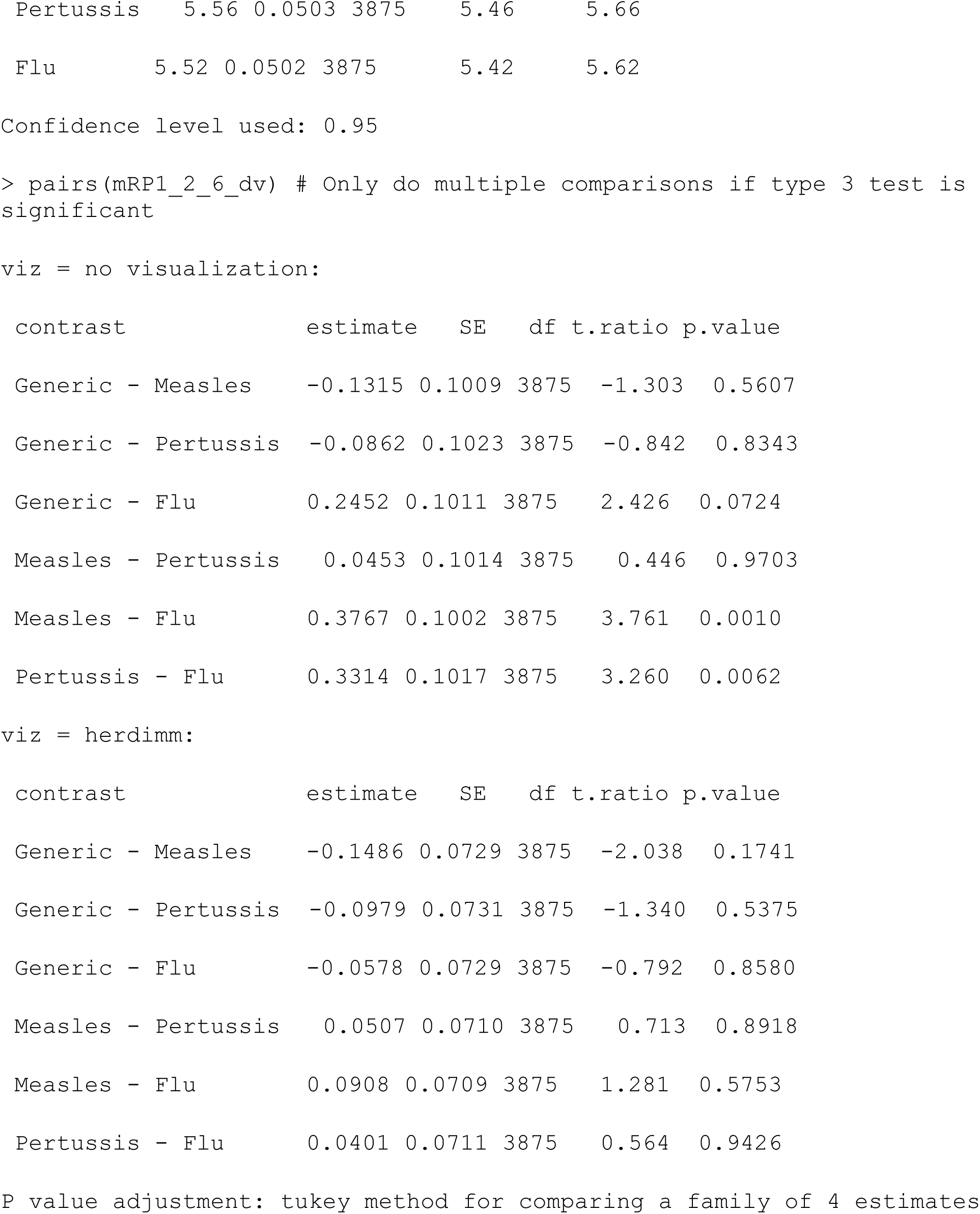

##### Model 2: Check for direct effects of factors with adjustment for other covariates

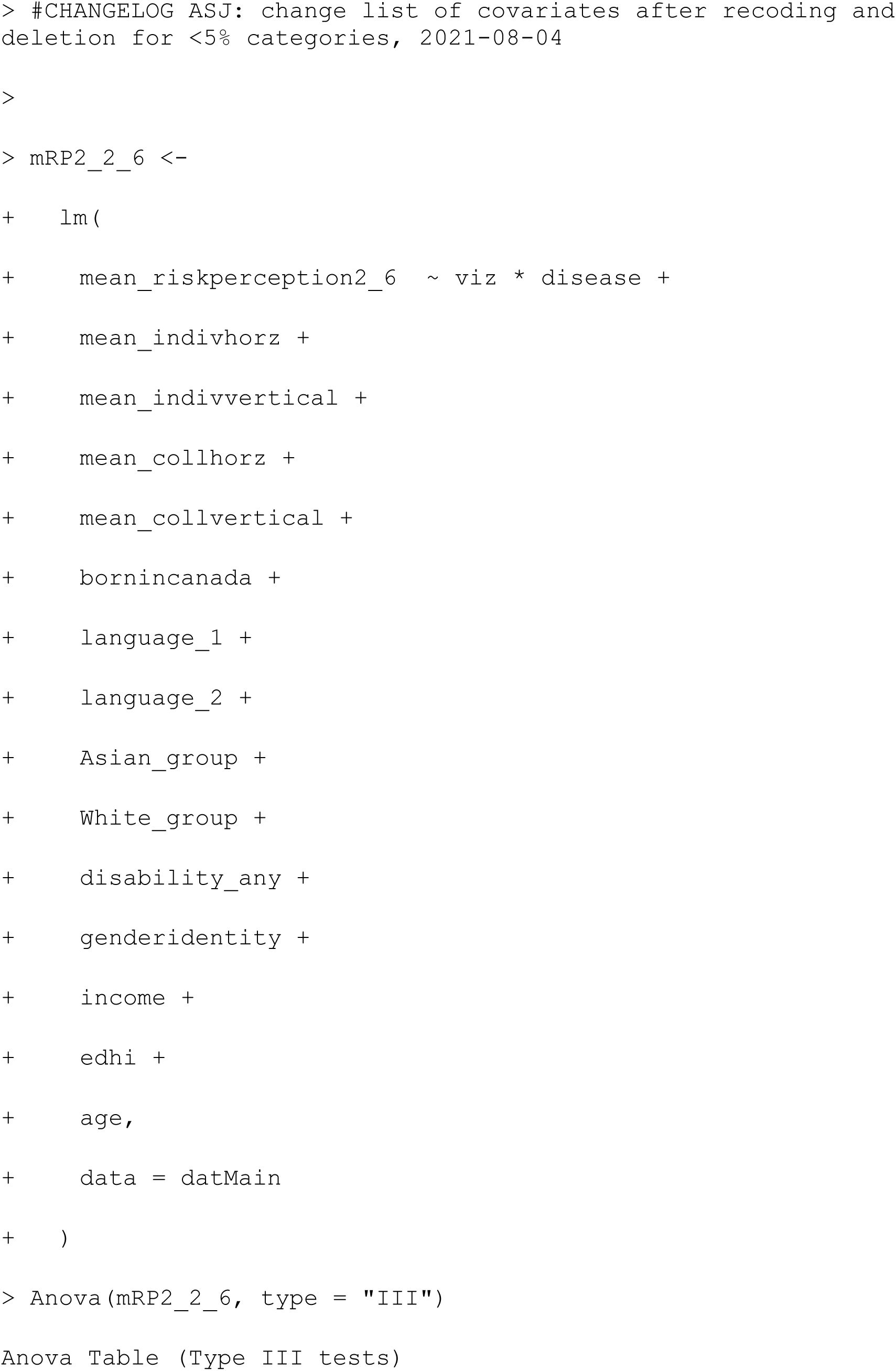
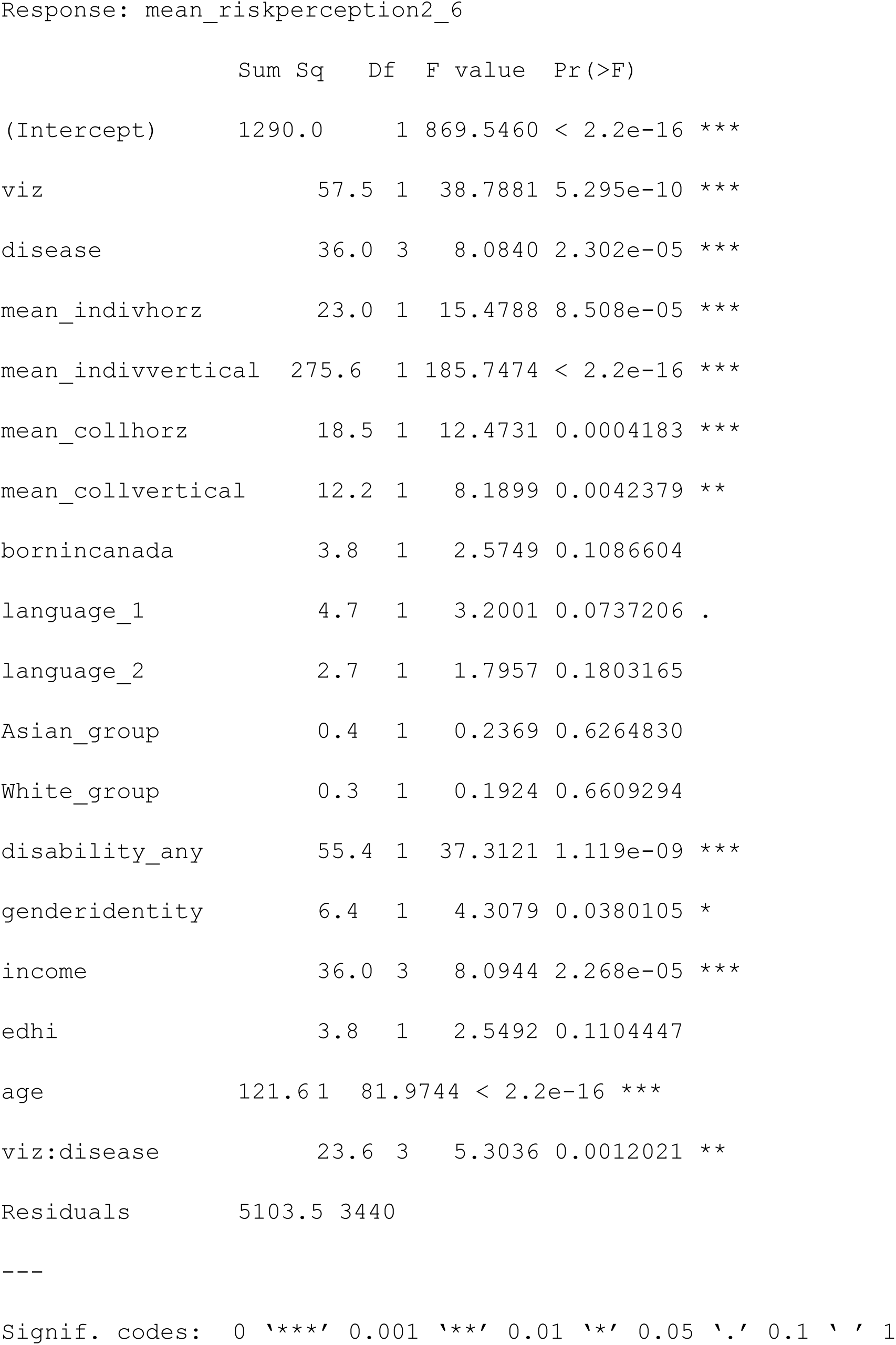
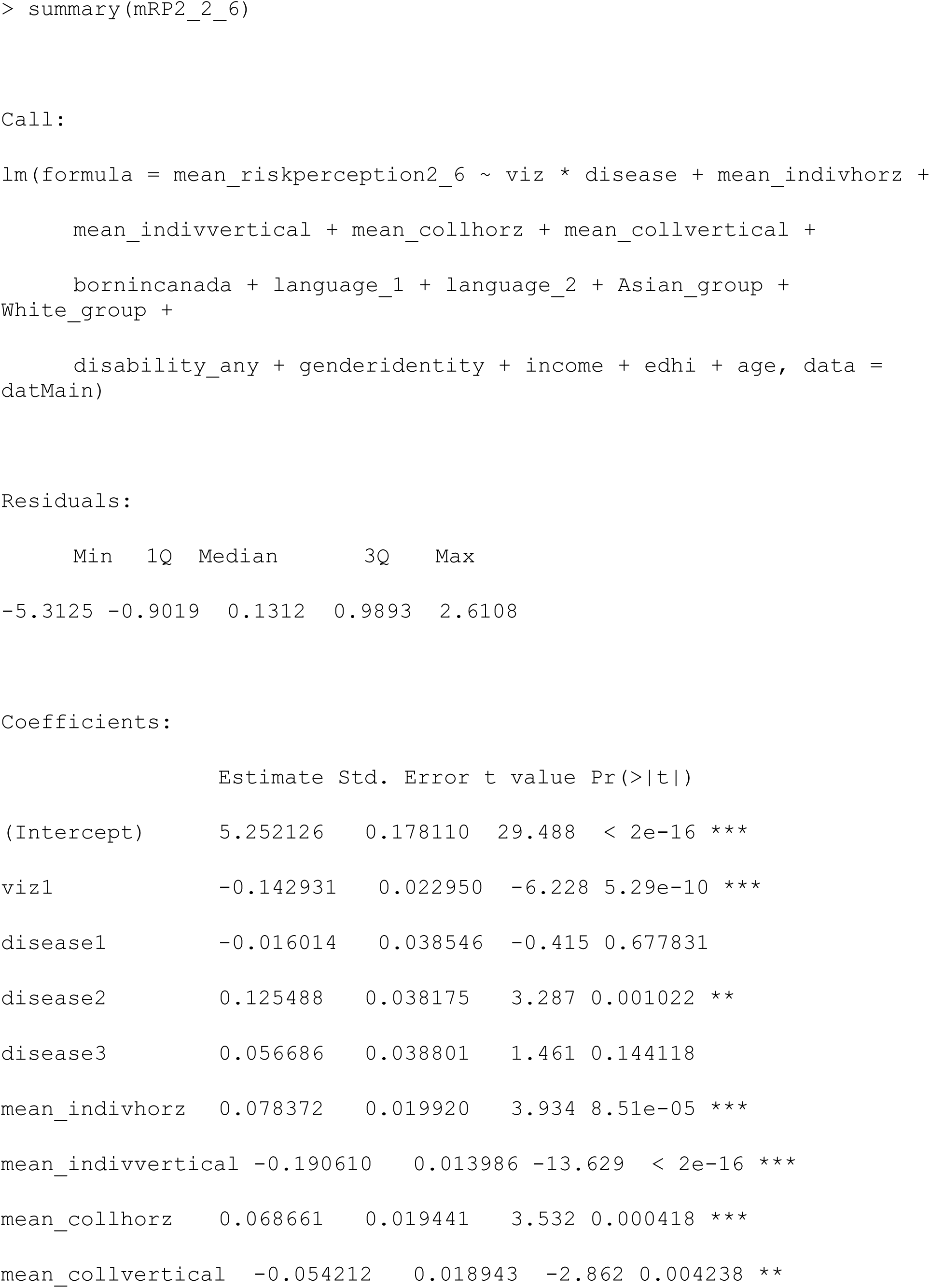
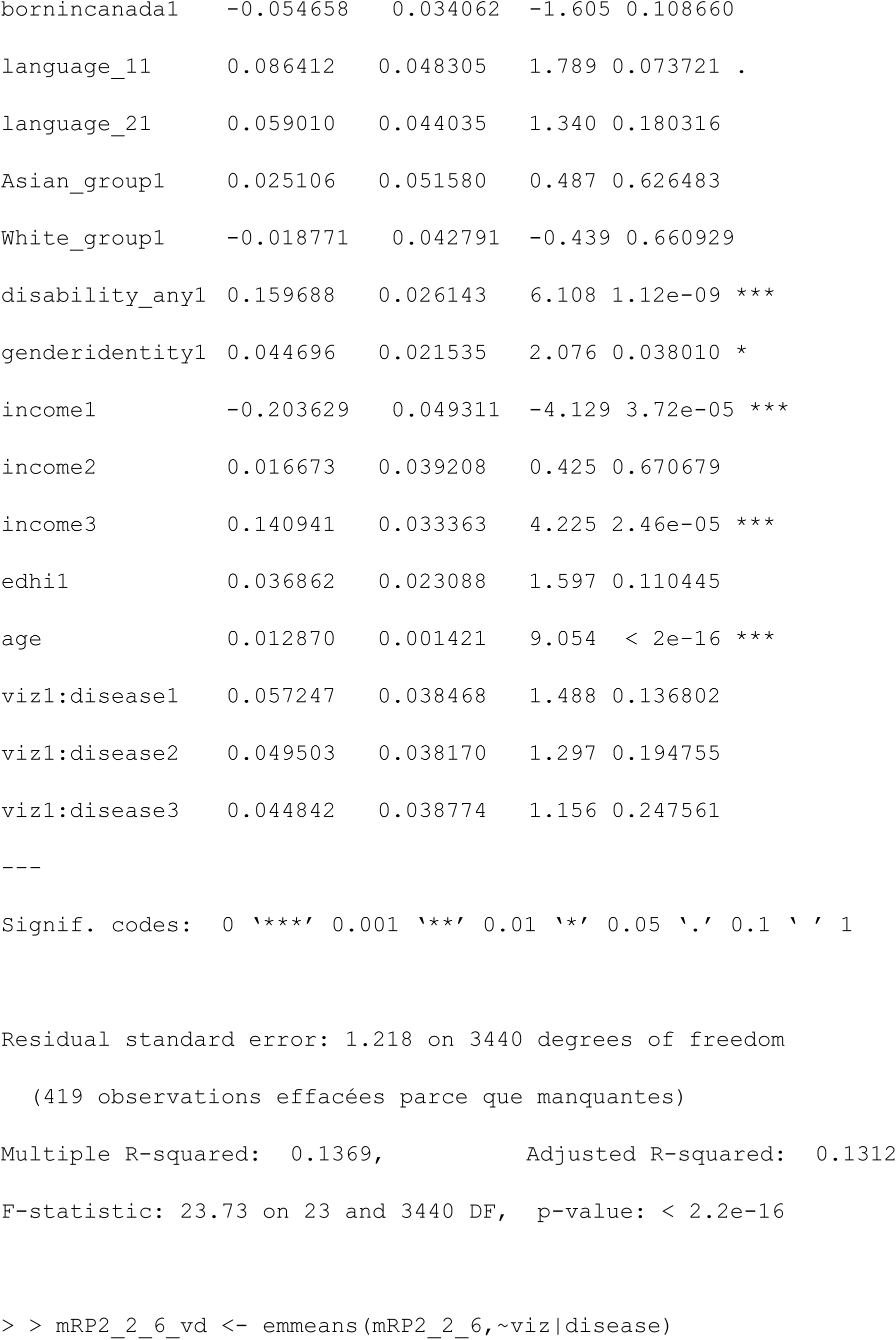
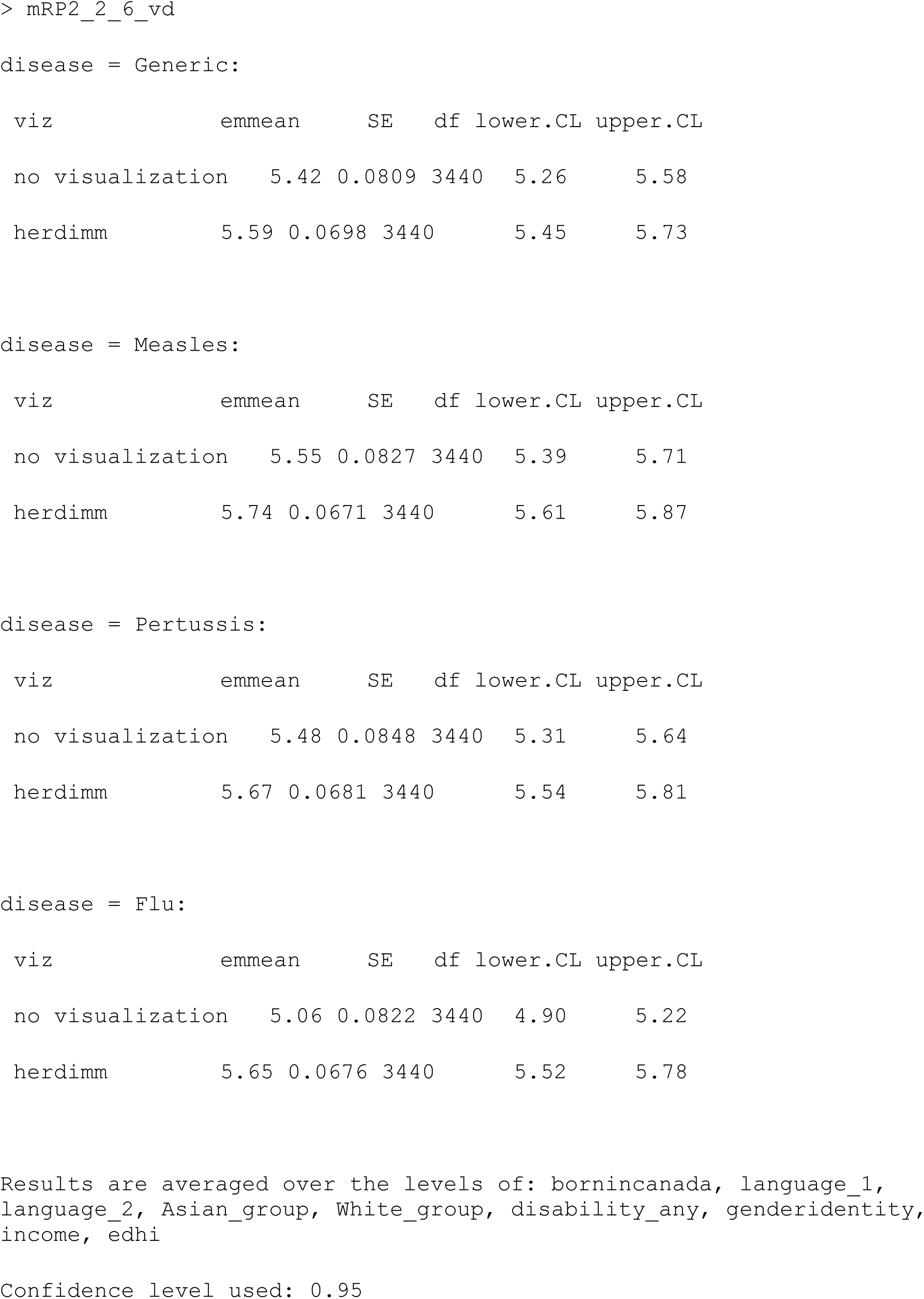
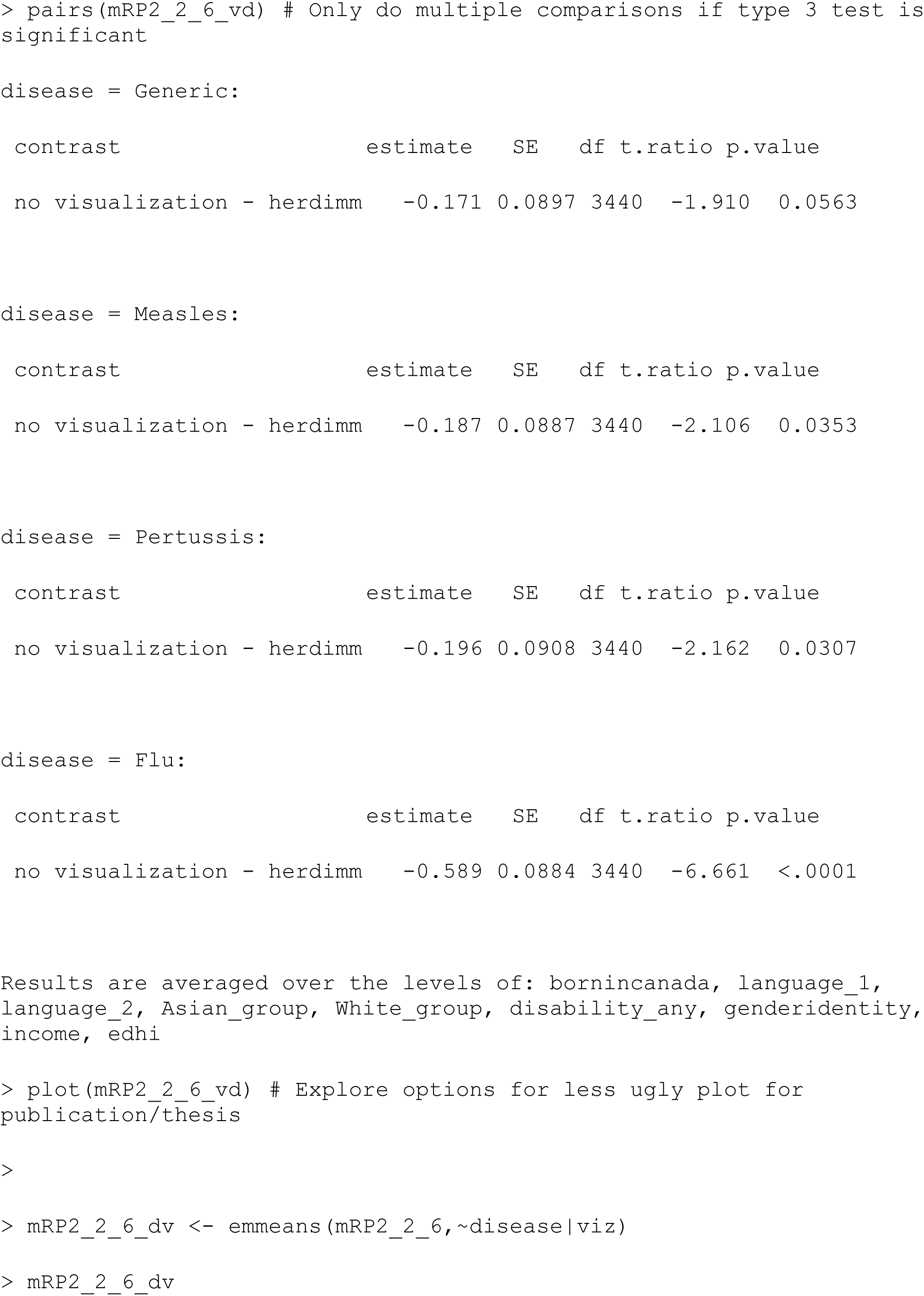
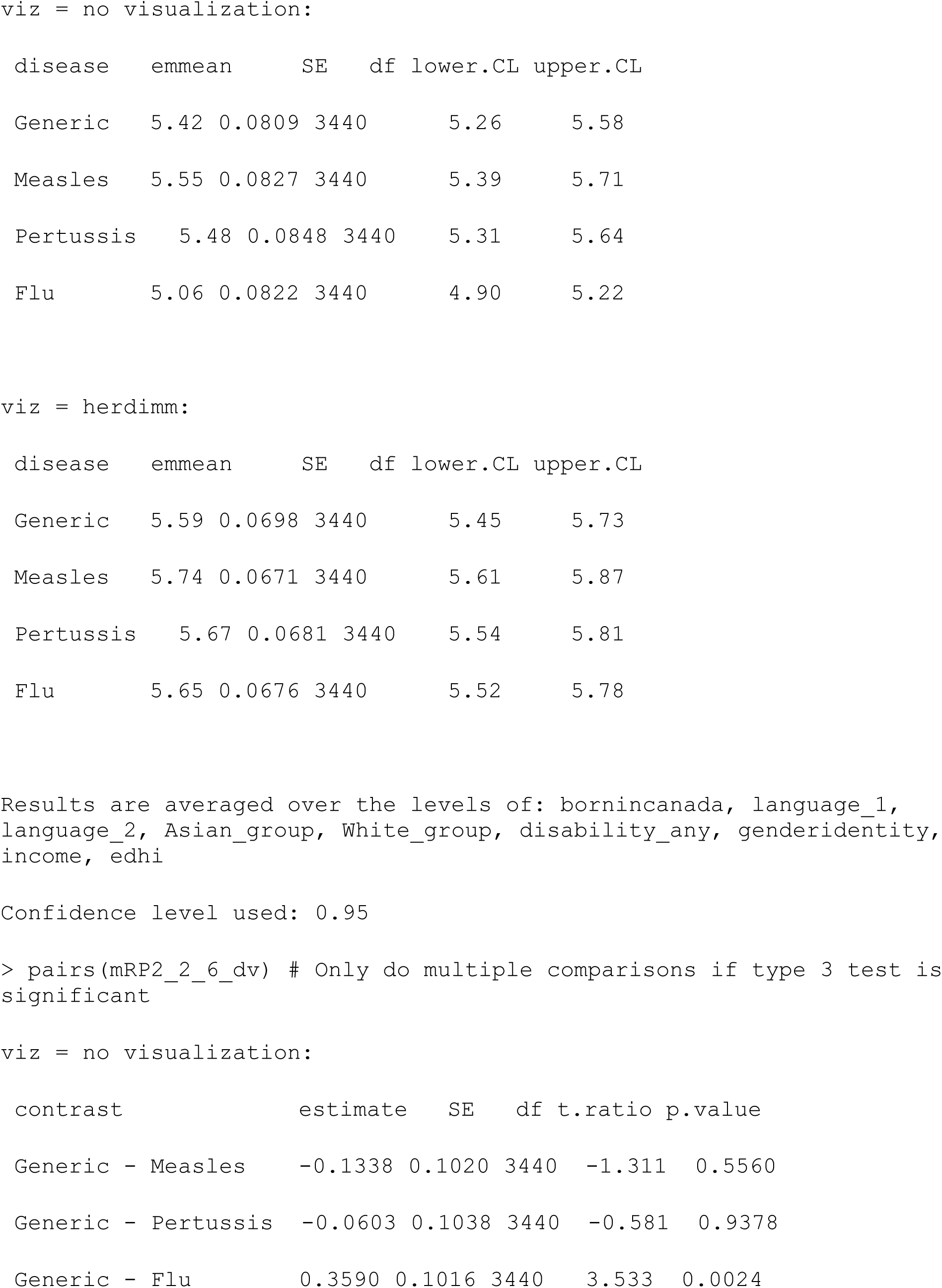
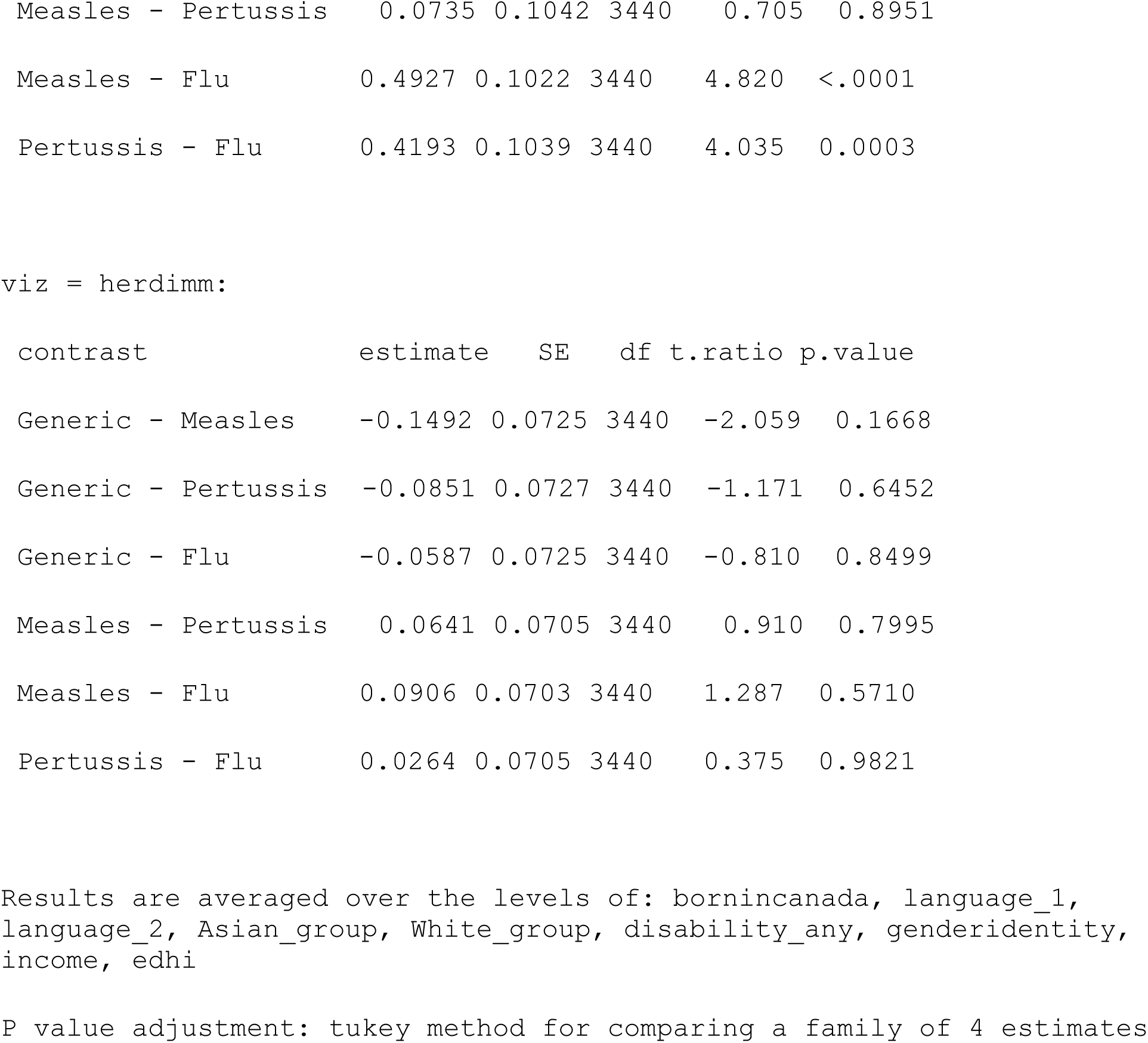

##### Model 3: Check for moderating effects of individualism & collectivism with adjustment for other covariates

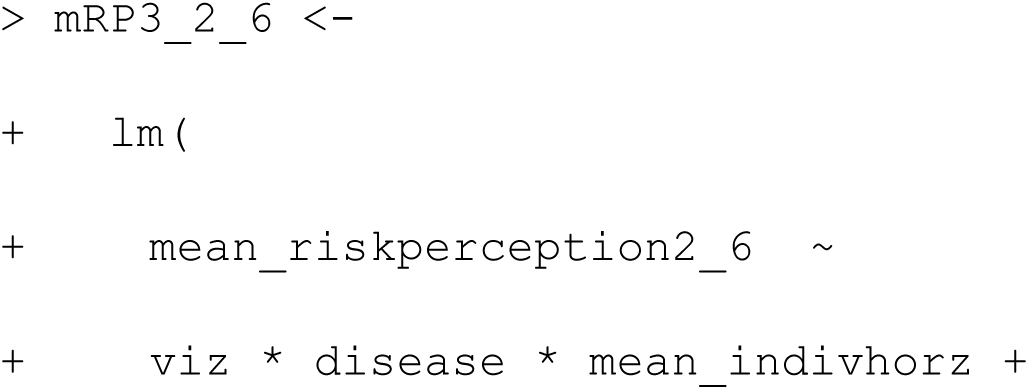
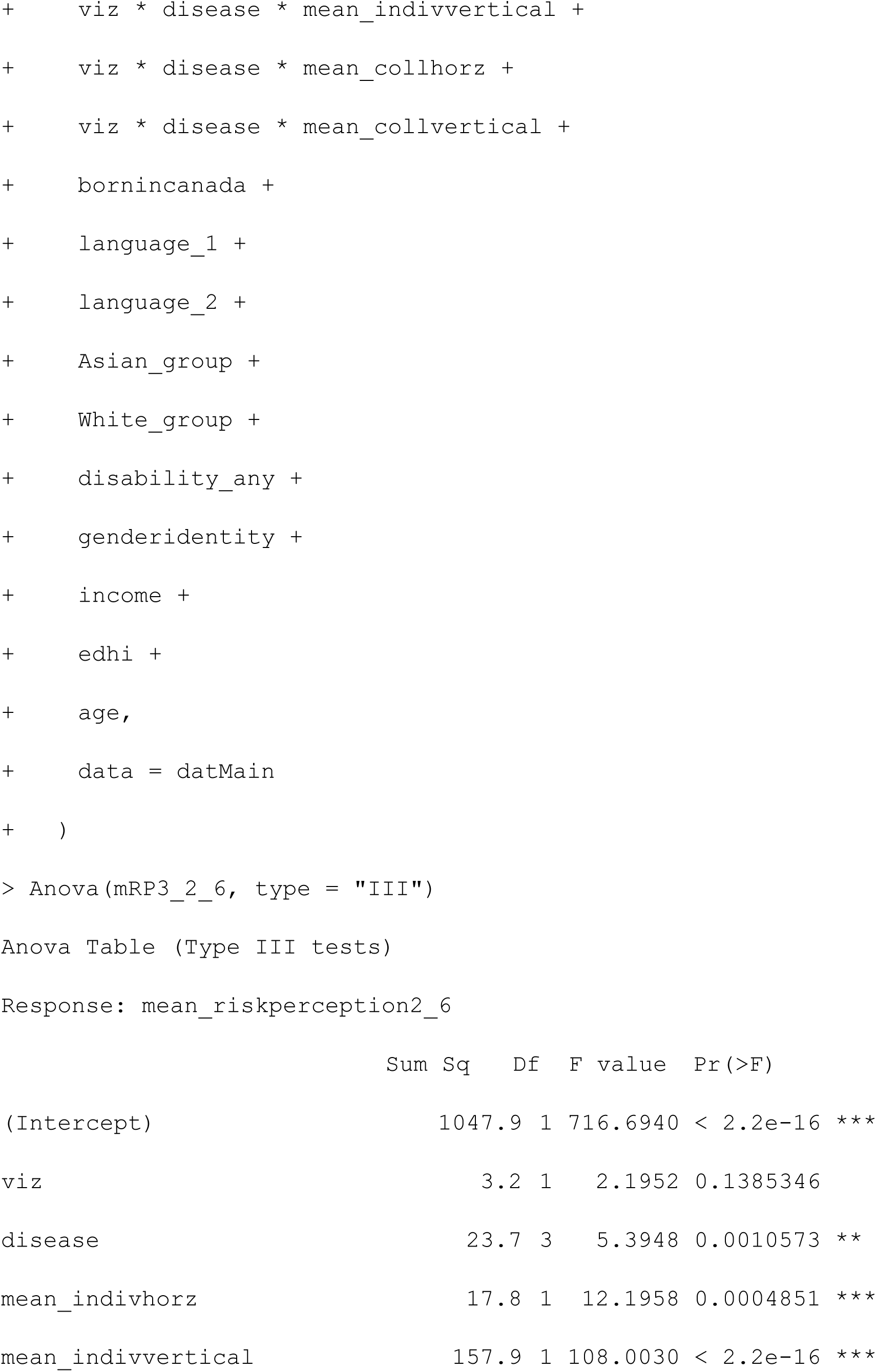
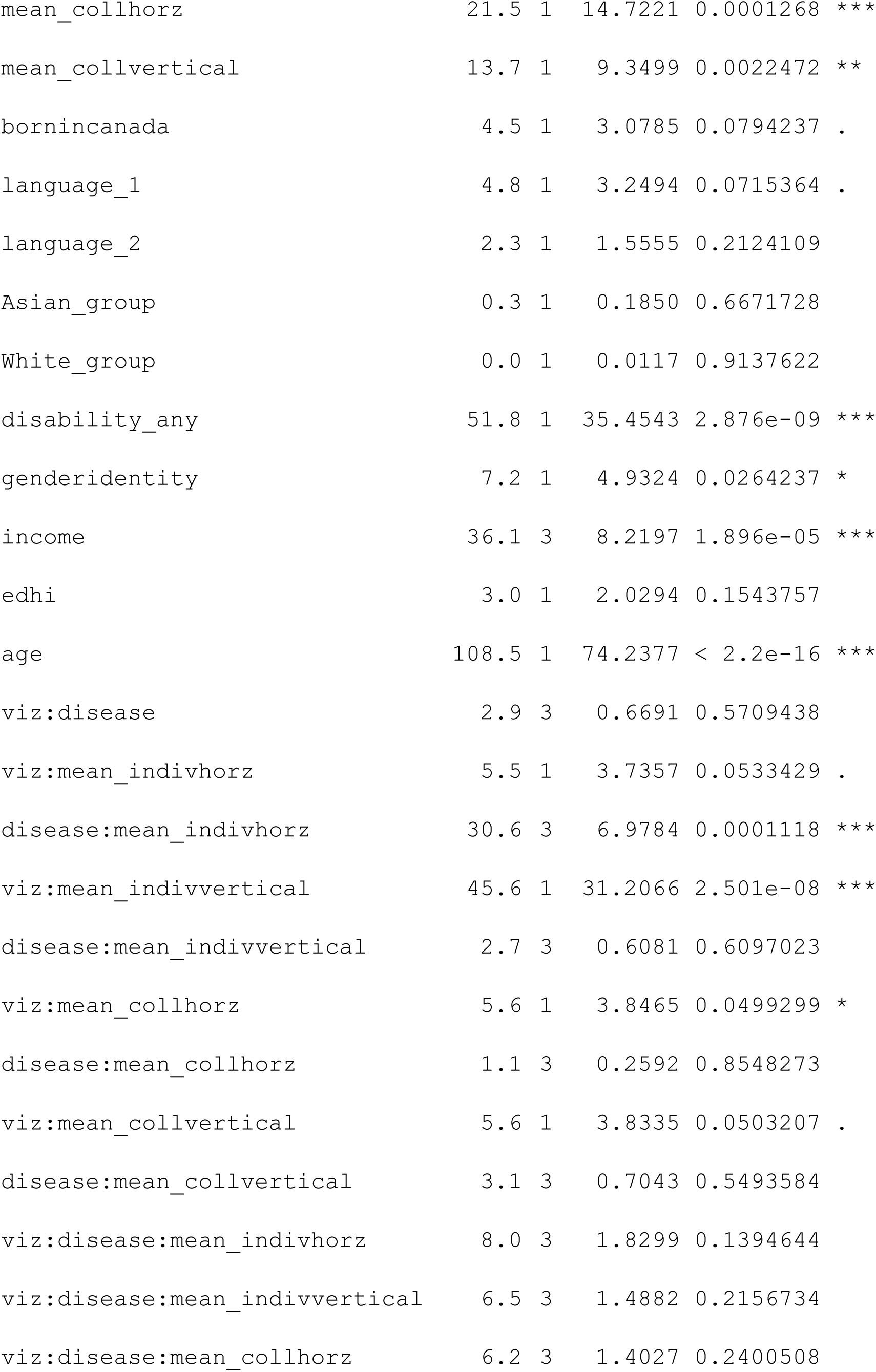
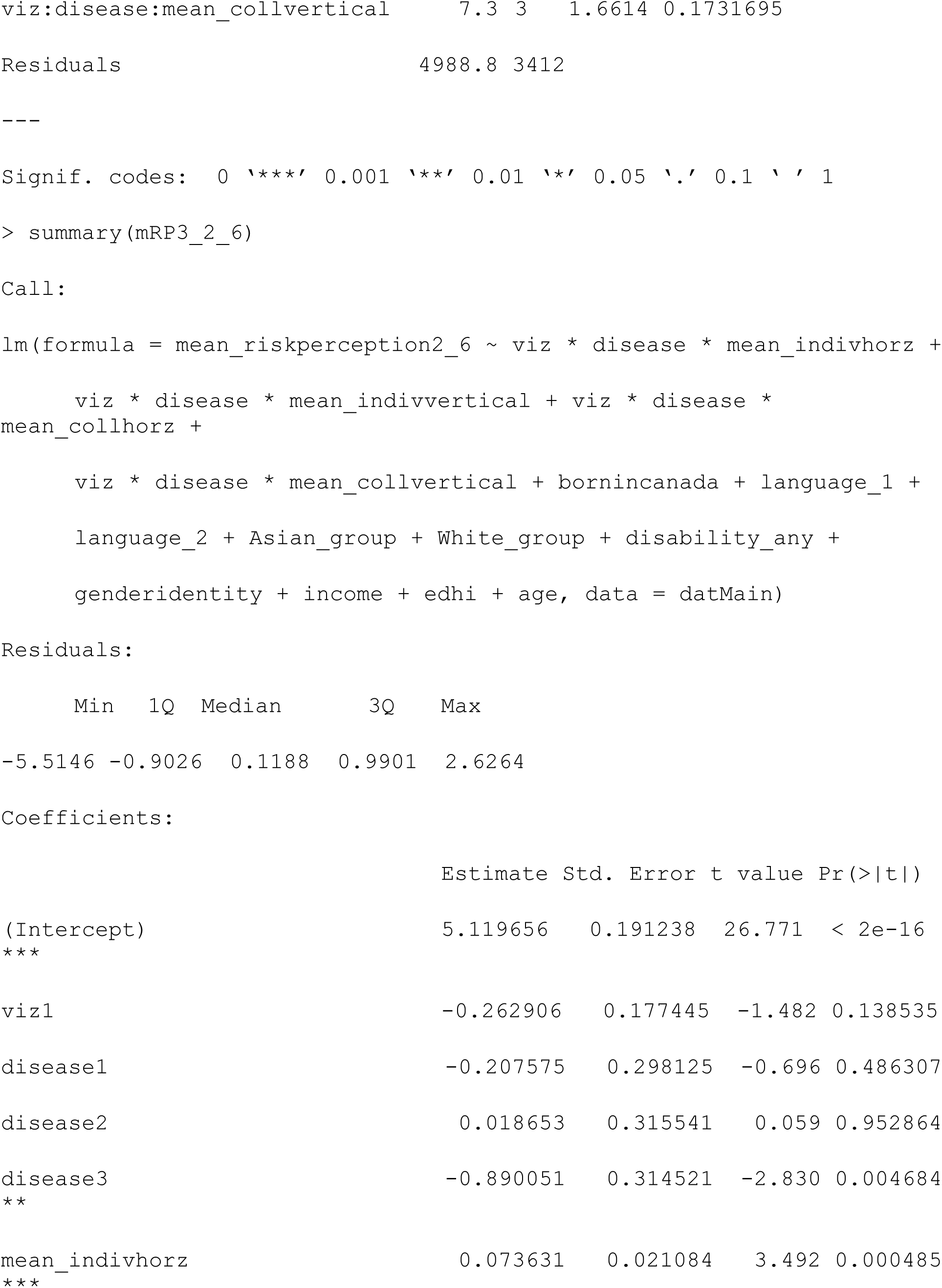
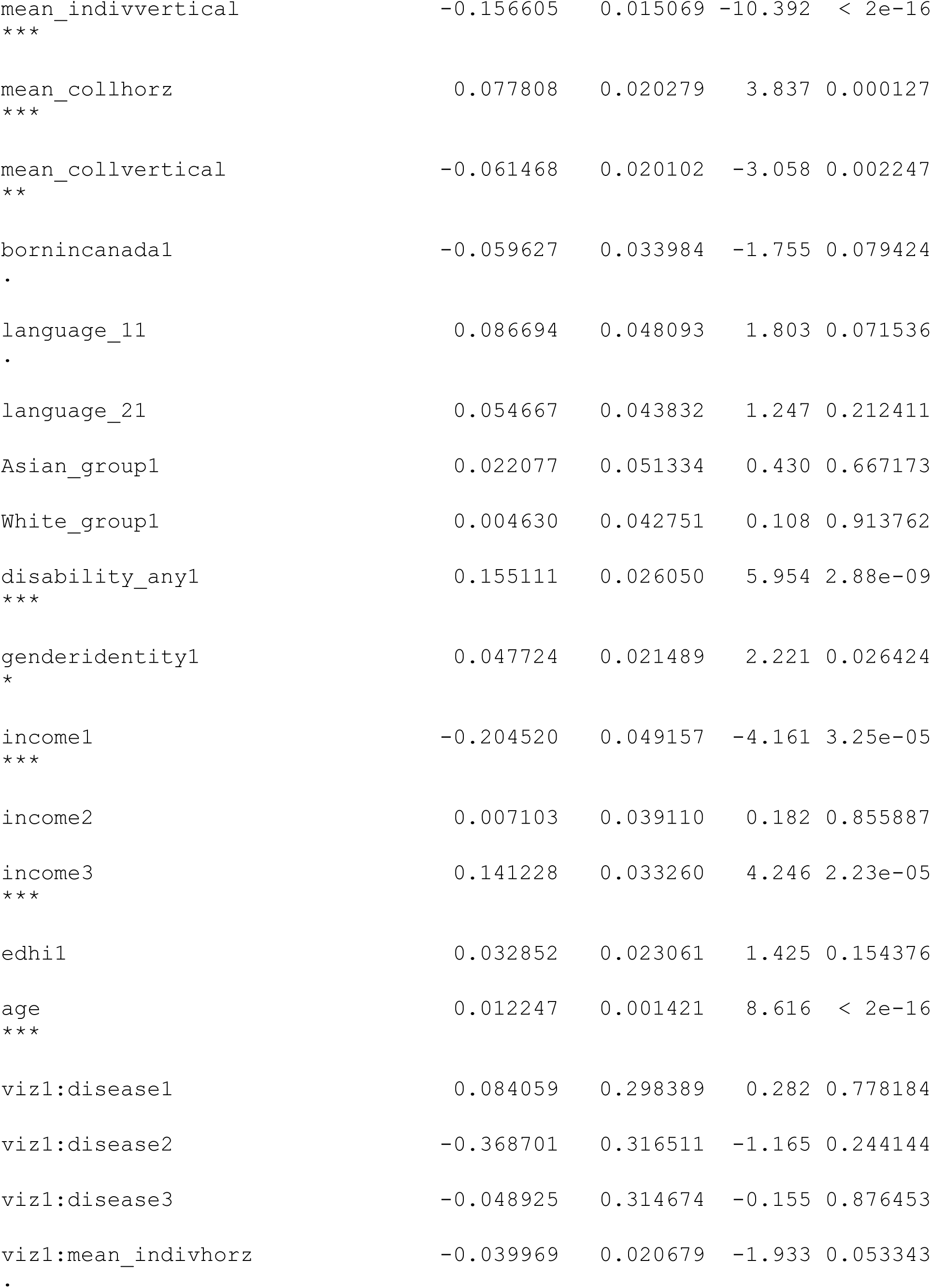
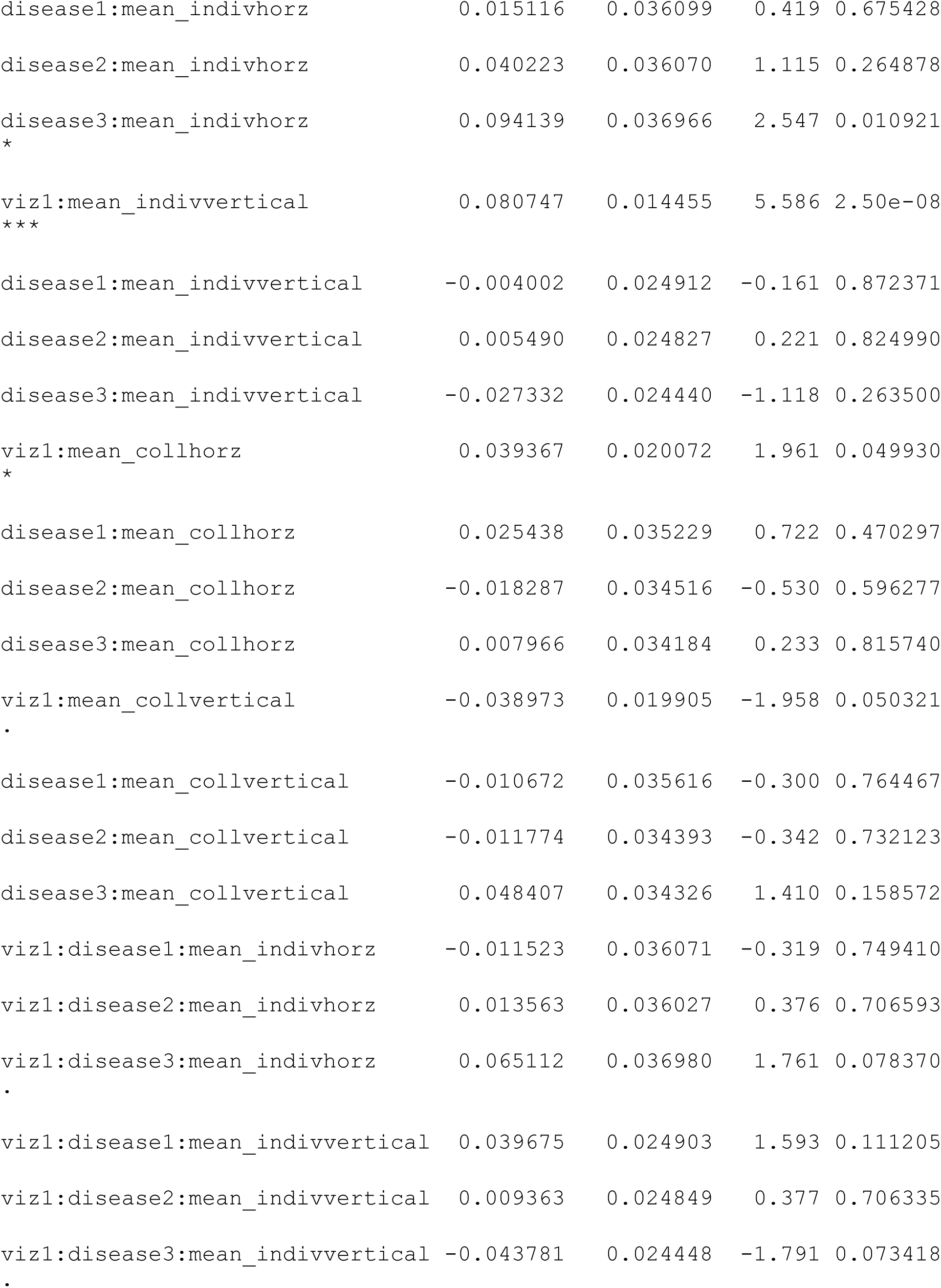
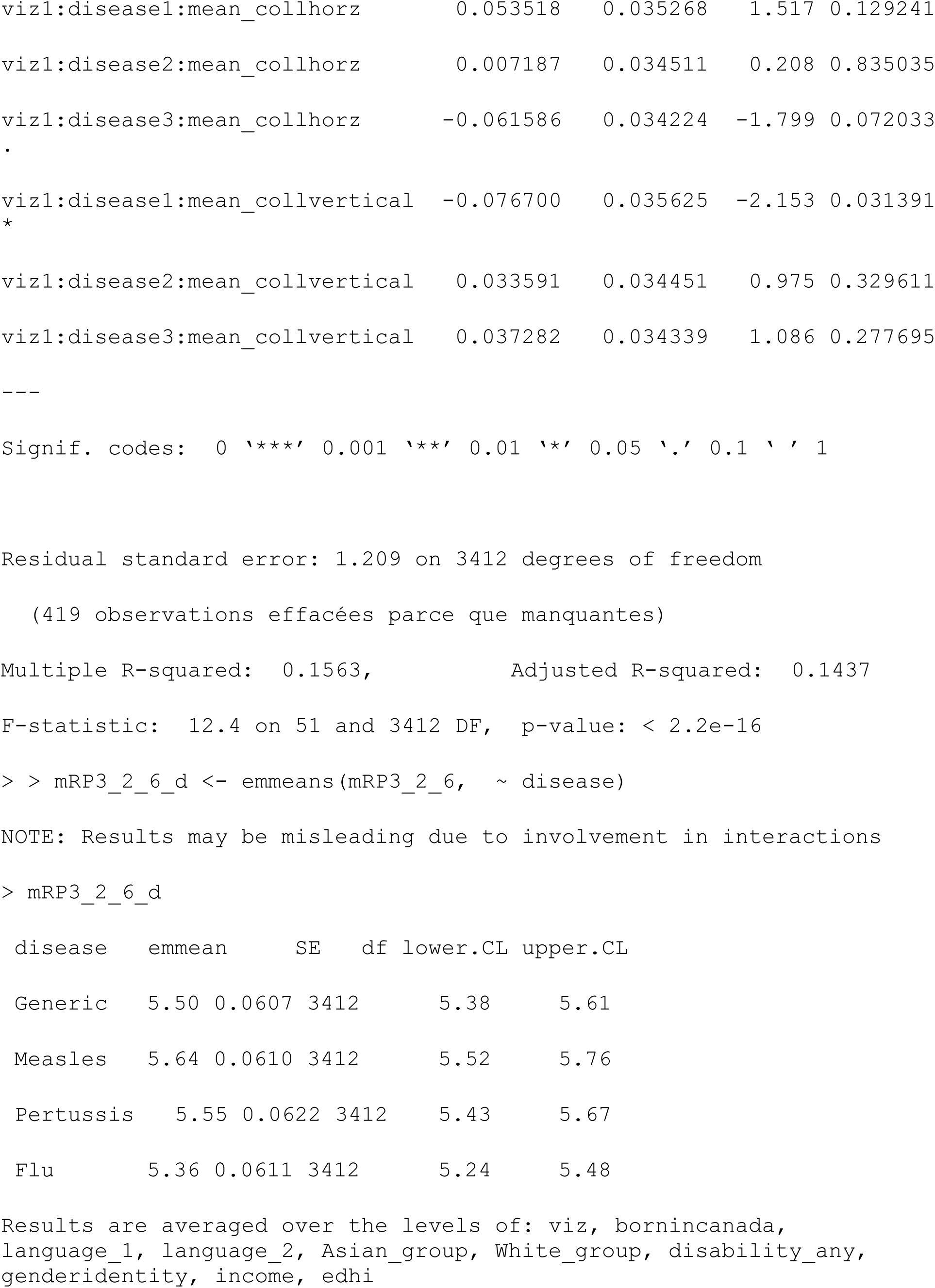
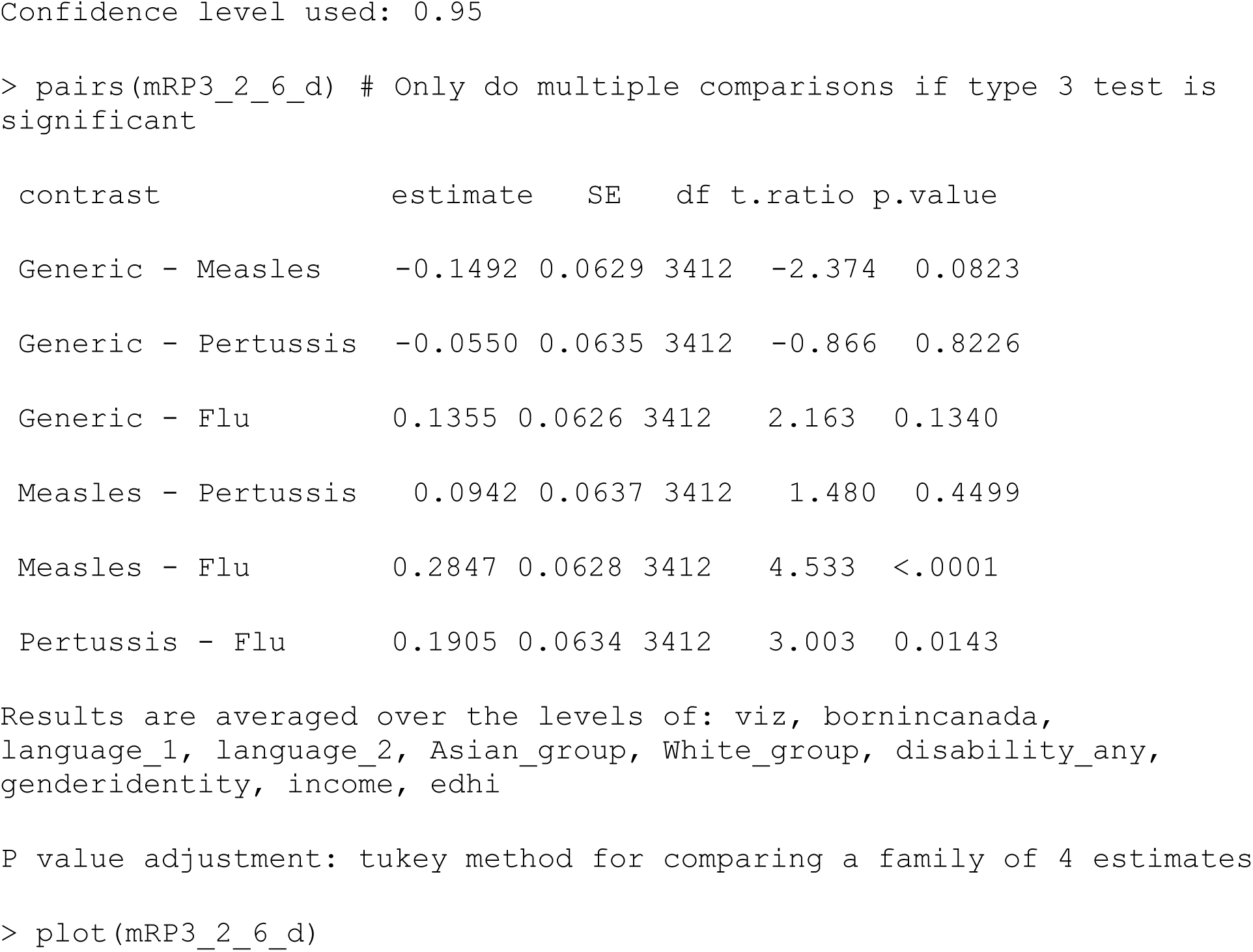

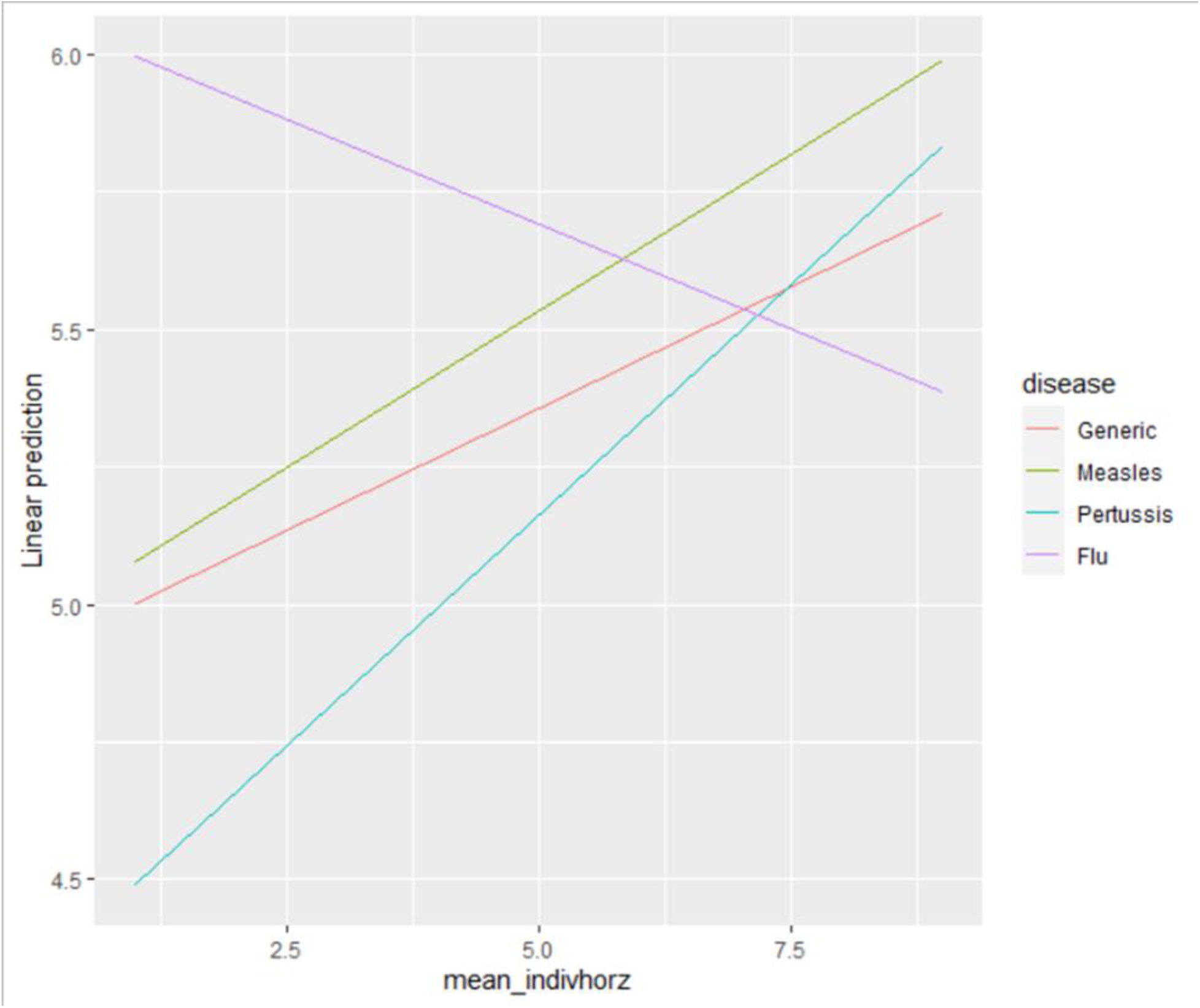

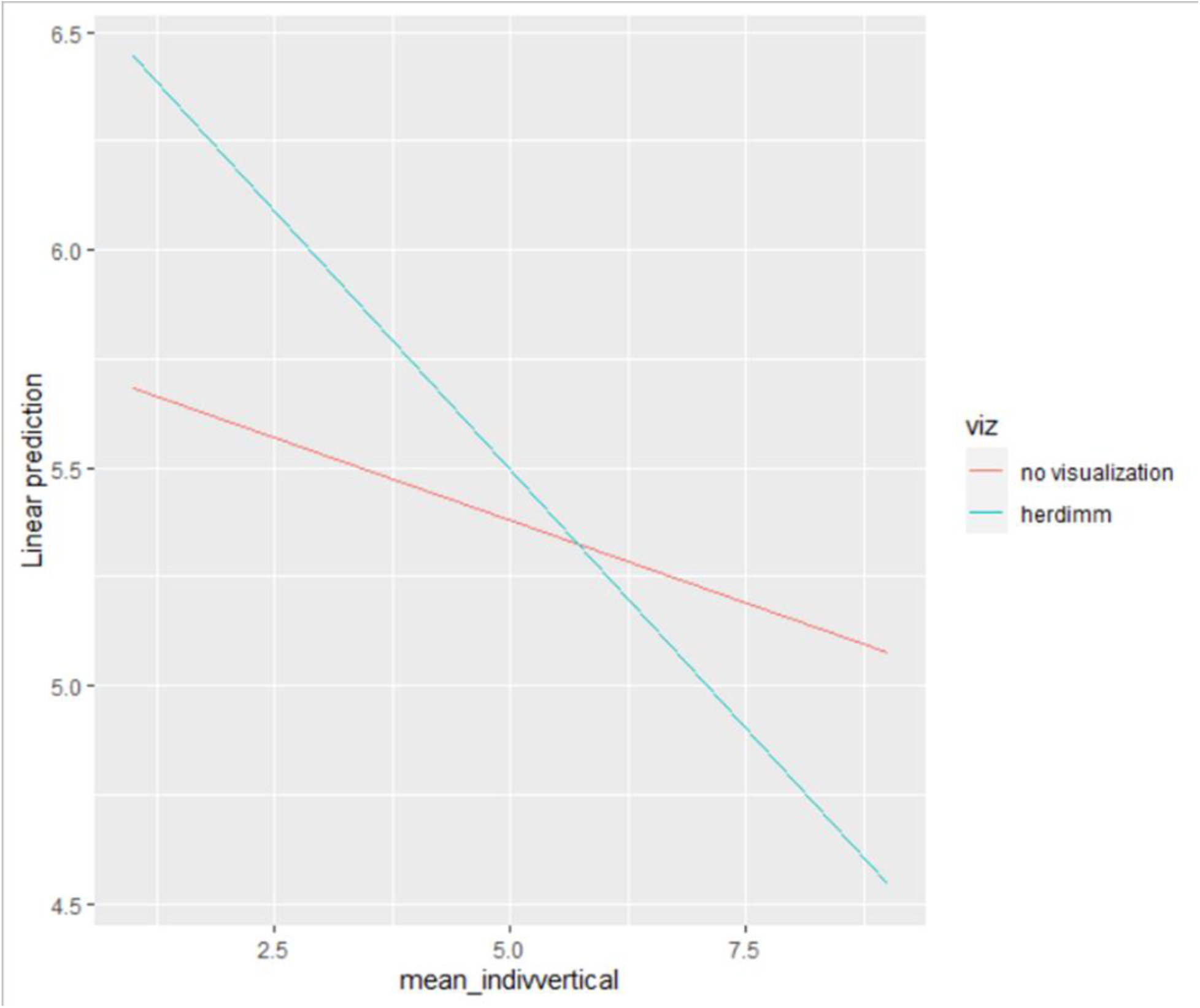

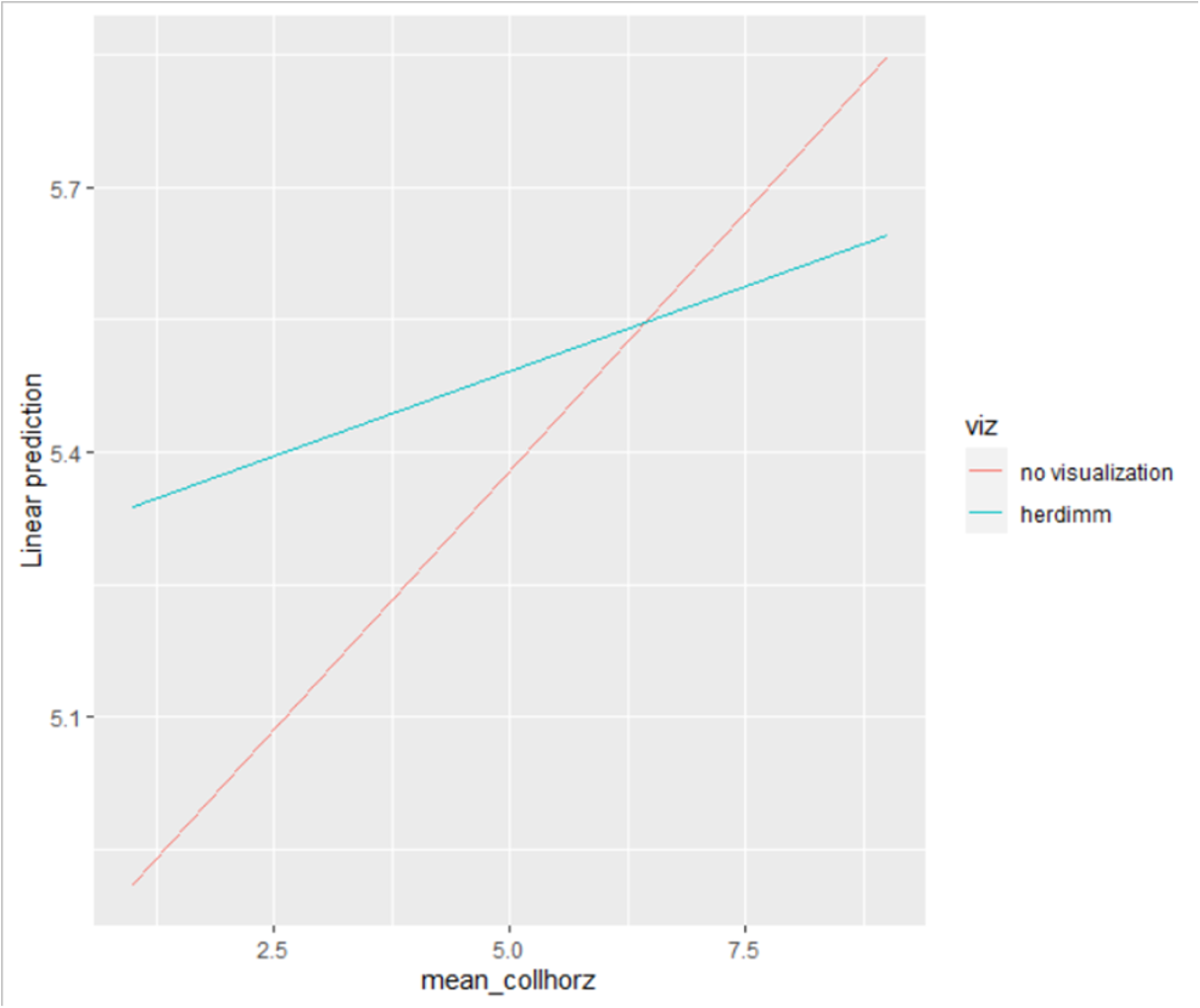

### Emotions

#### Two-way

##### Model 1: Check for direct effects of factors without any covariates ####

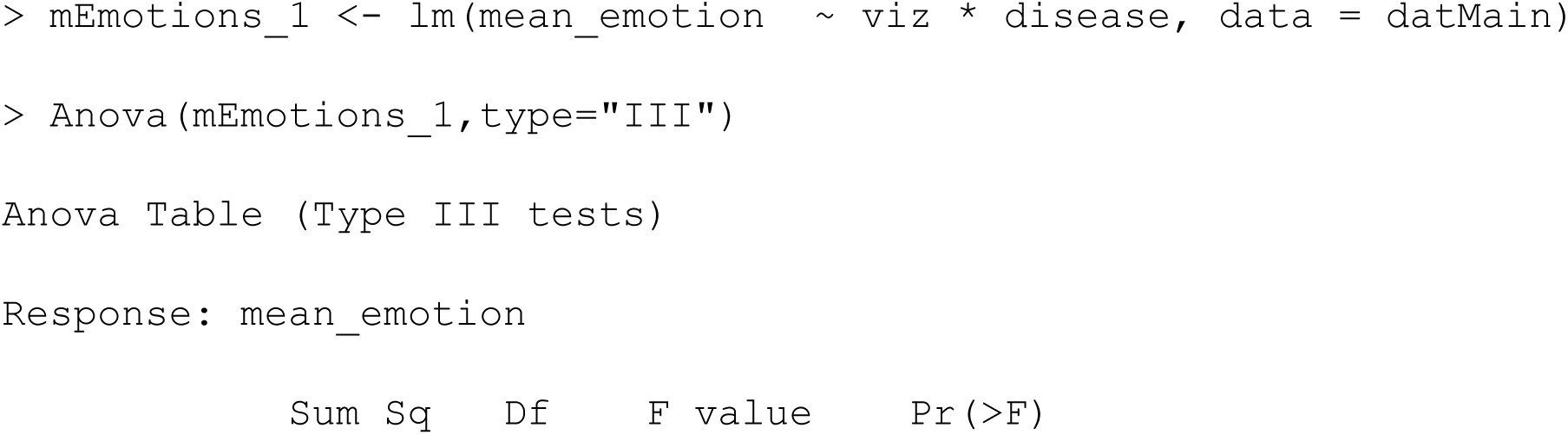
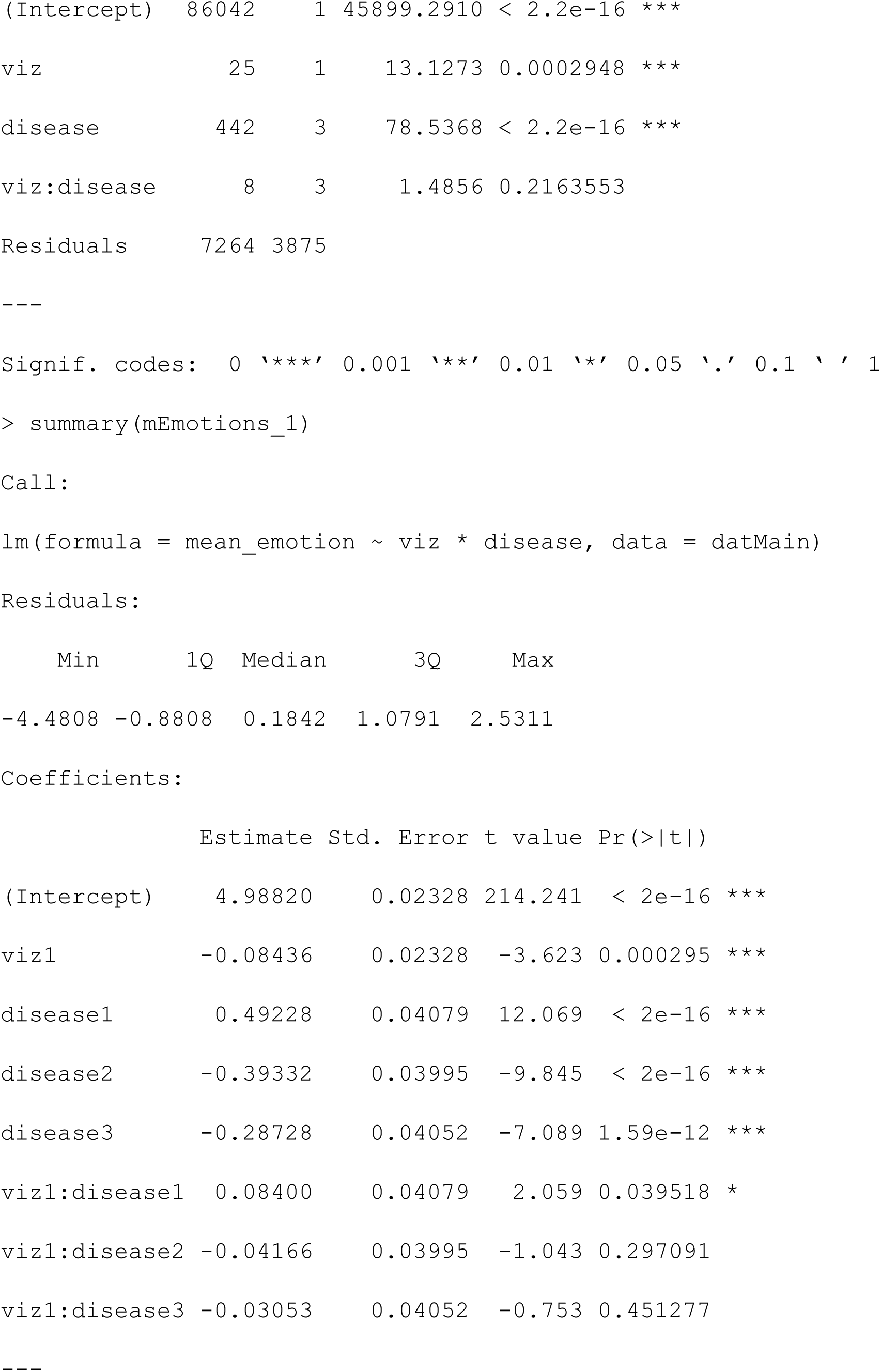
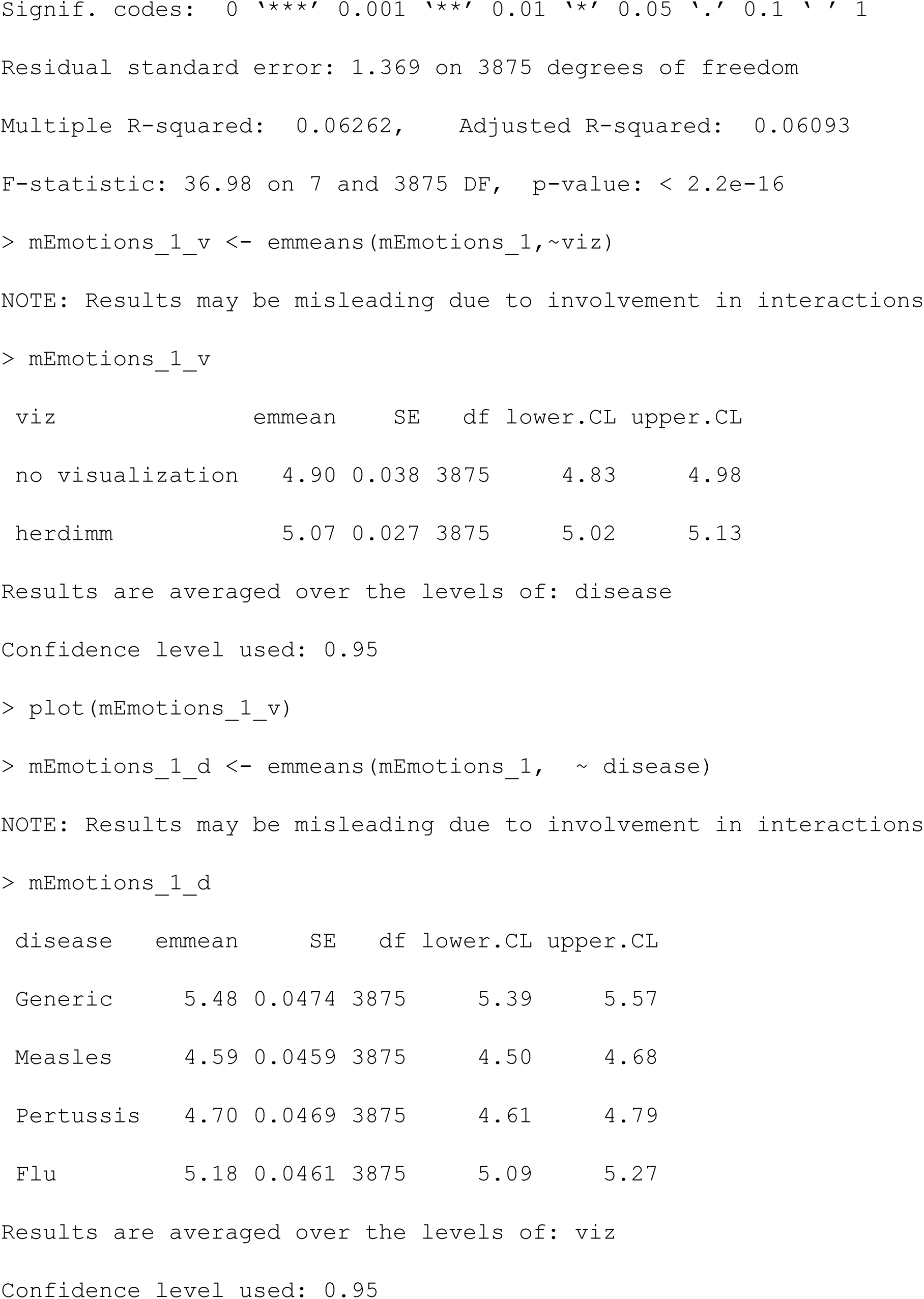
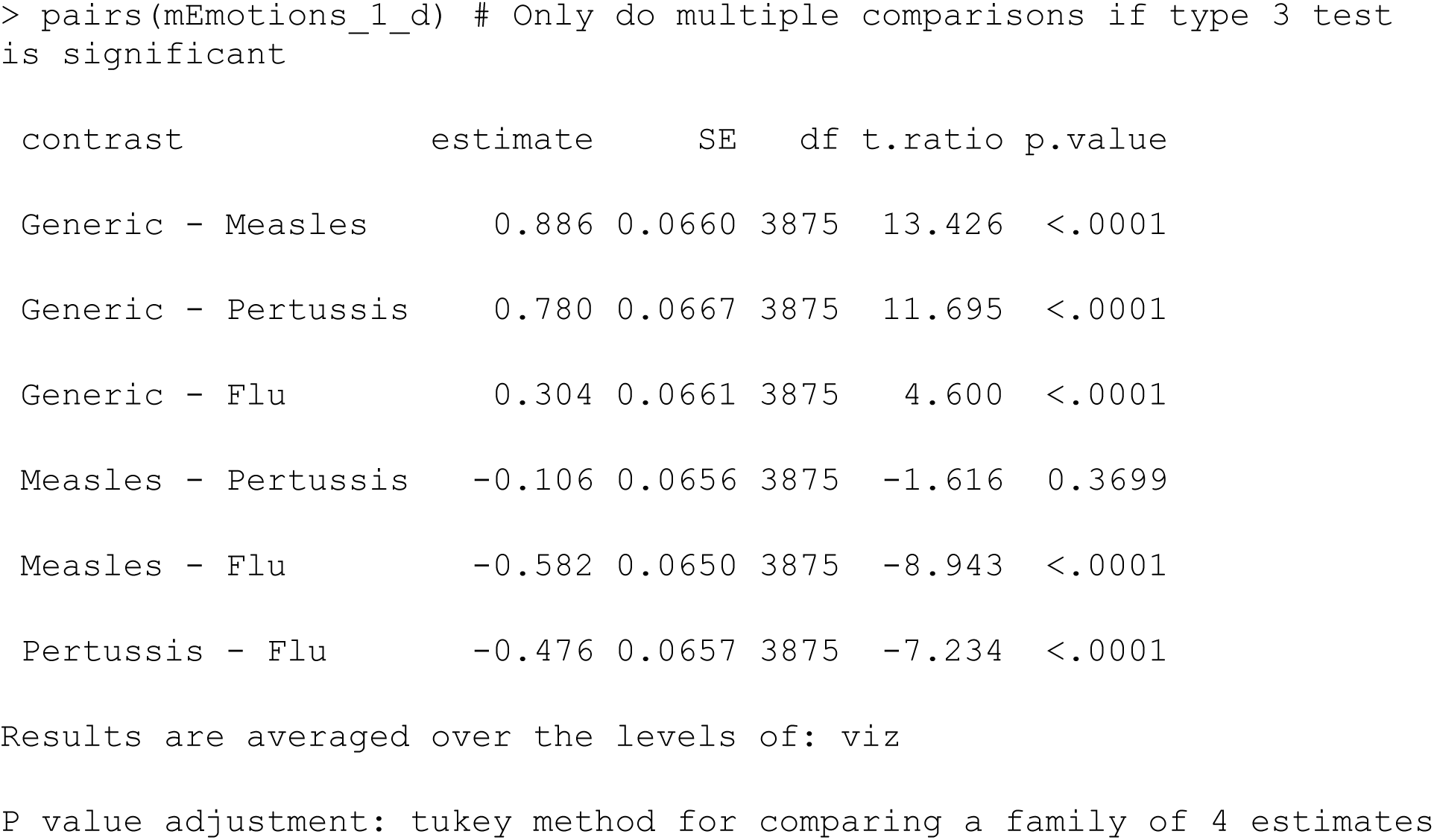

##### Model 2: Check for direct effects of factors with adjustment for other covariates

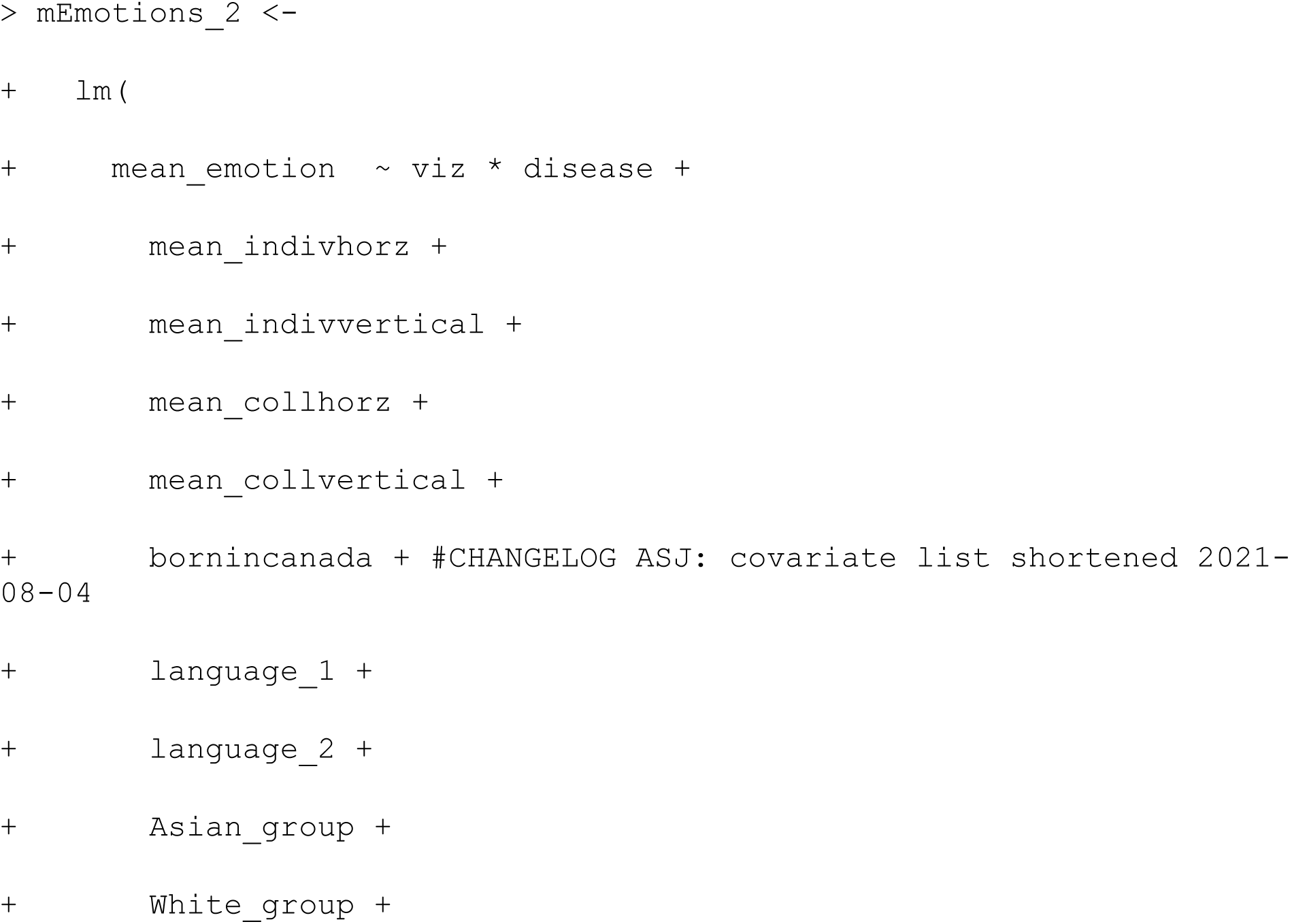
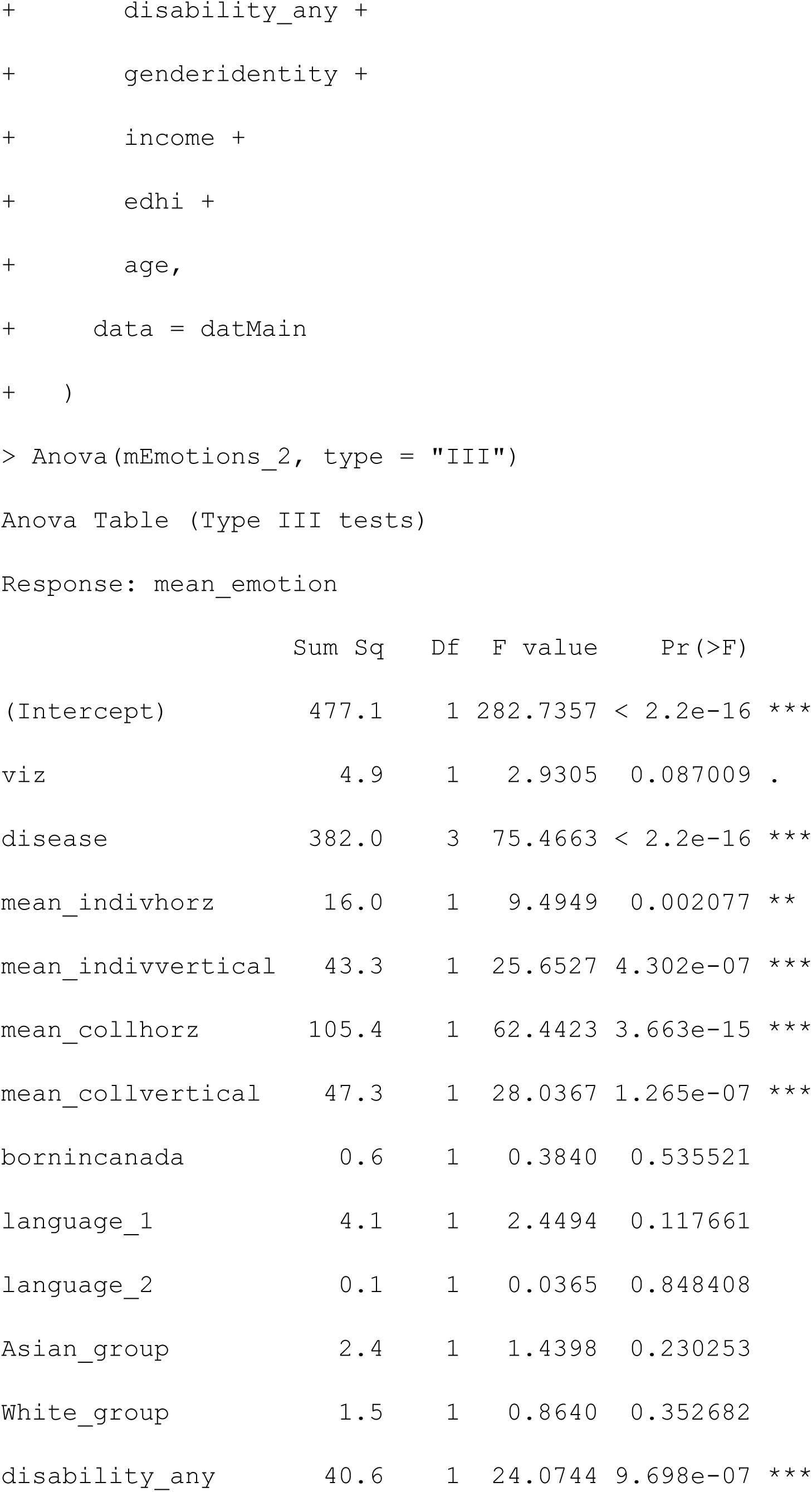
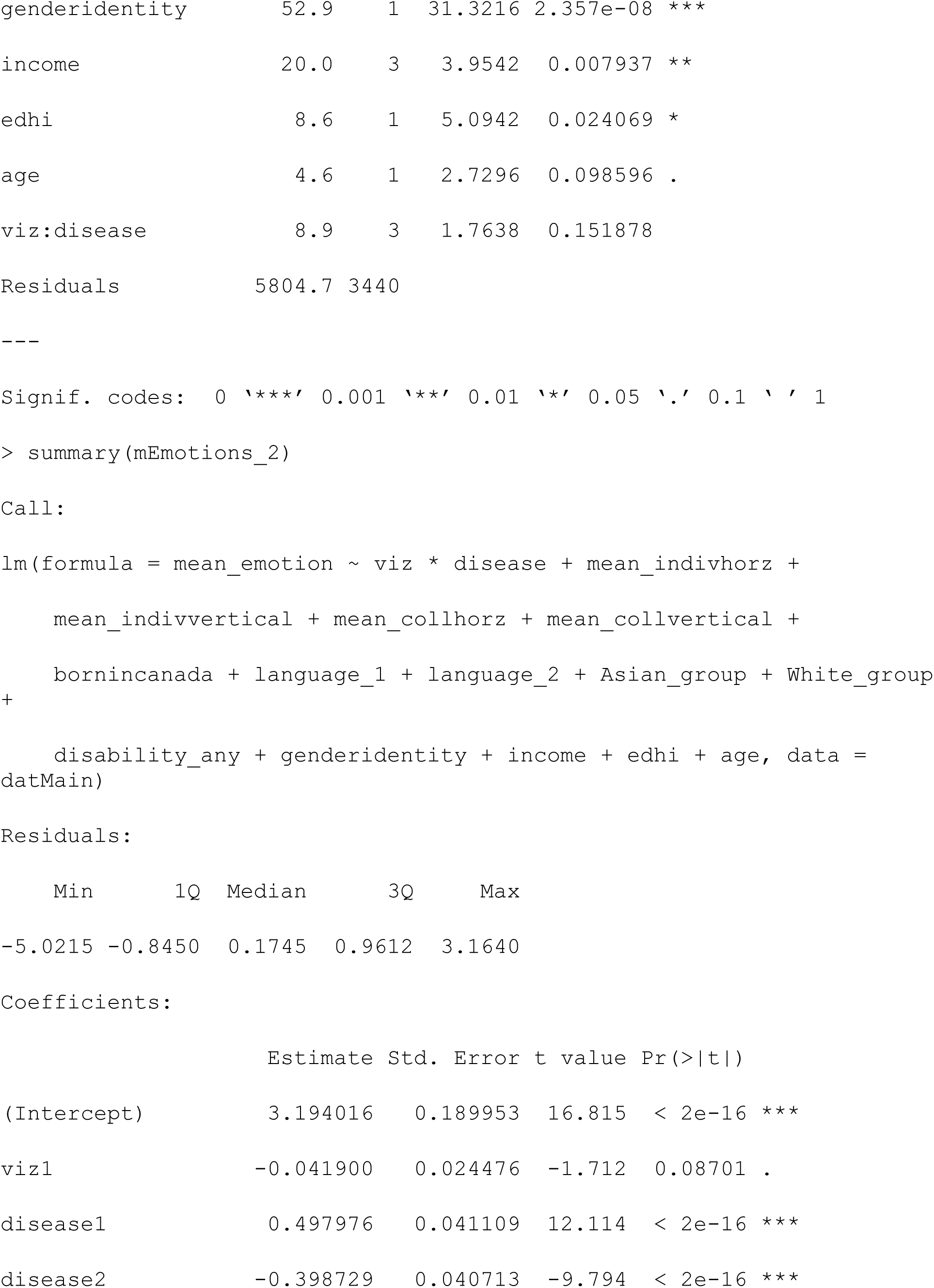
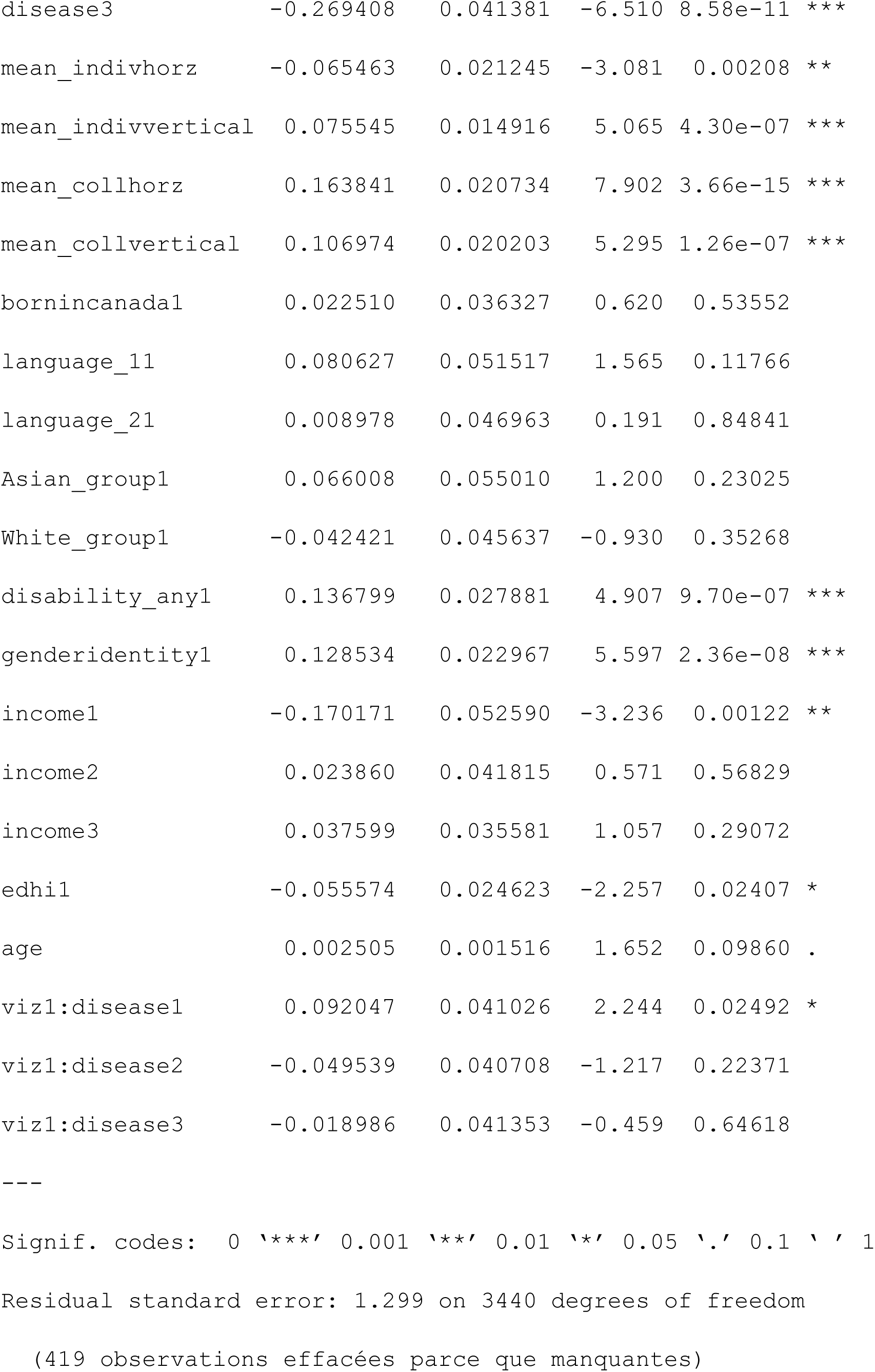
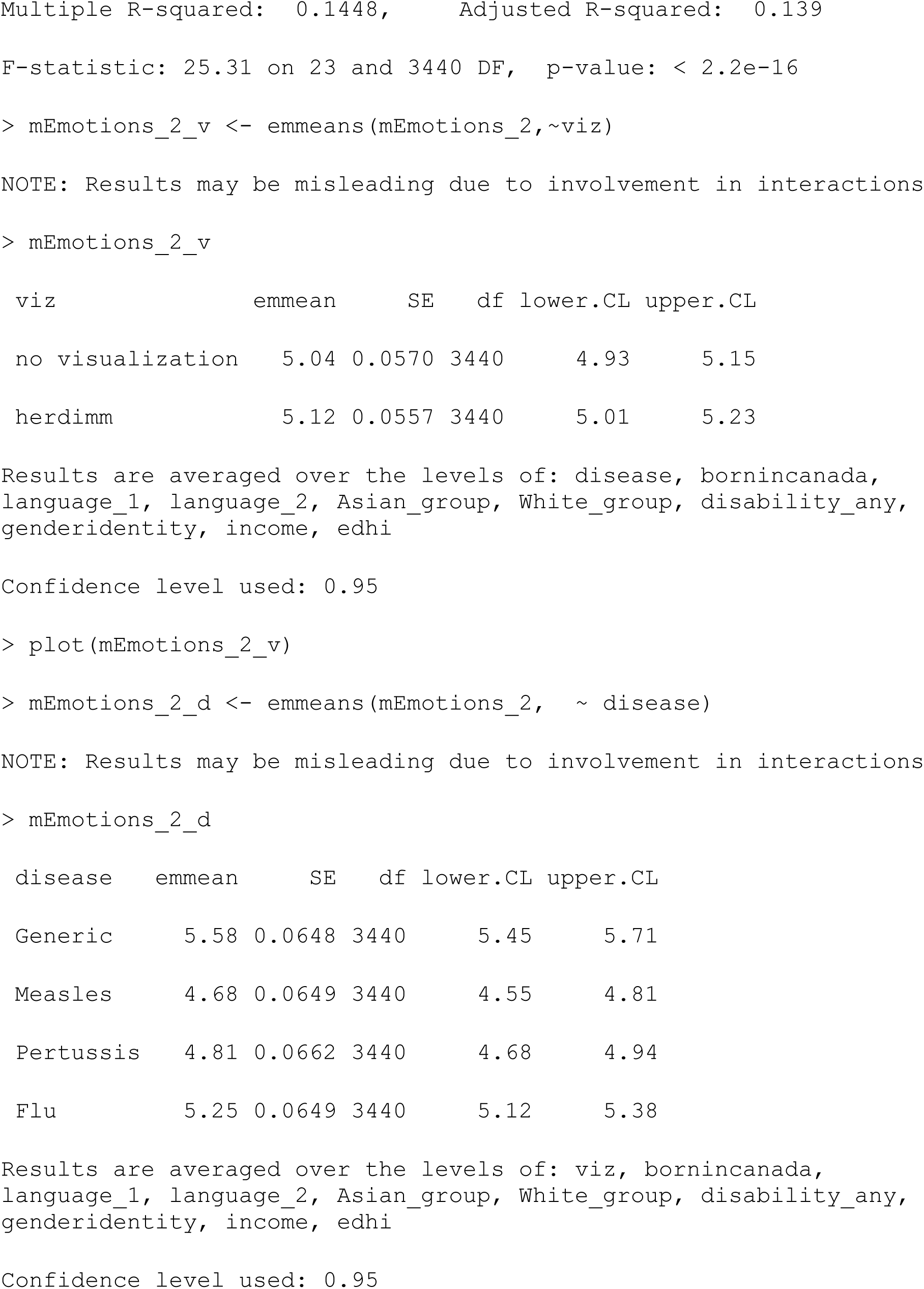
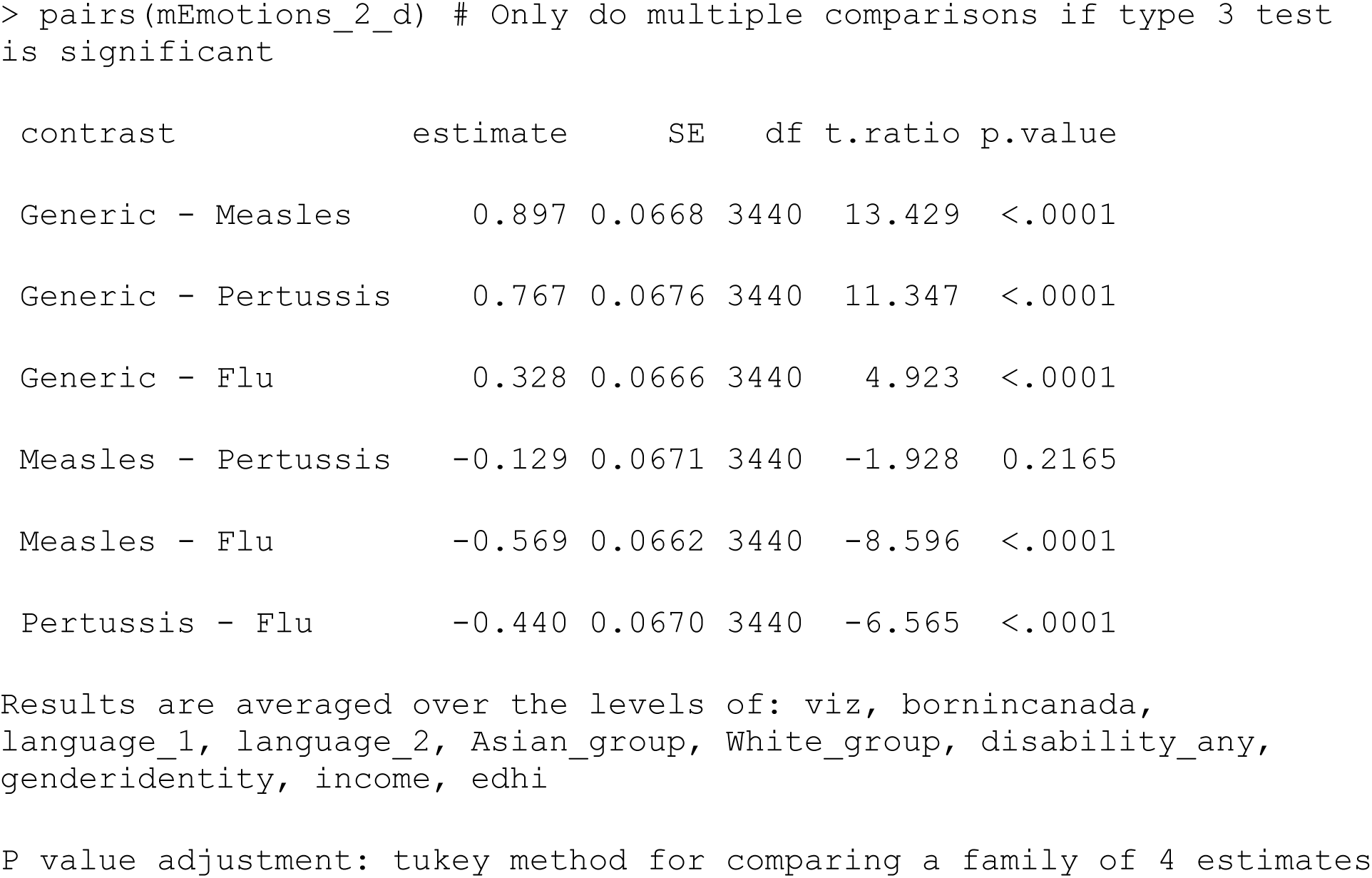

##### Model 3: Check for moderating effects of individualism & collectivism with adjustment for other covariates

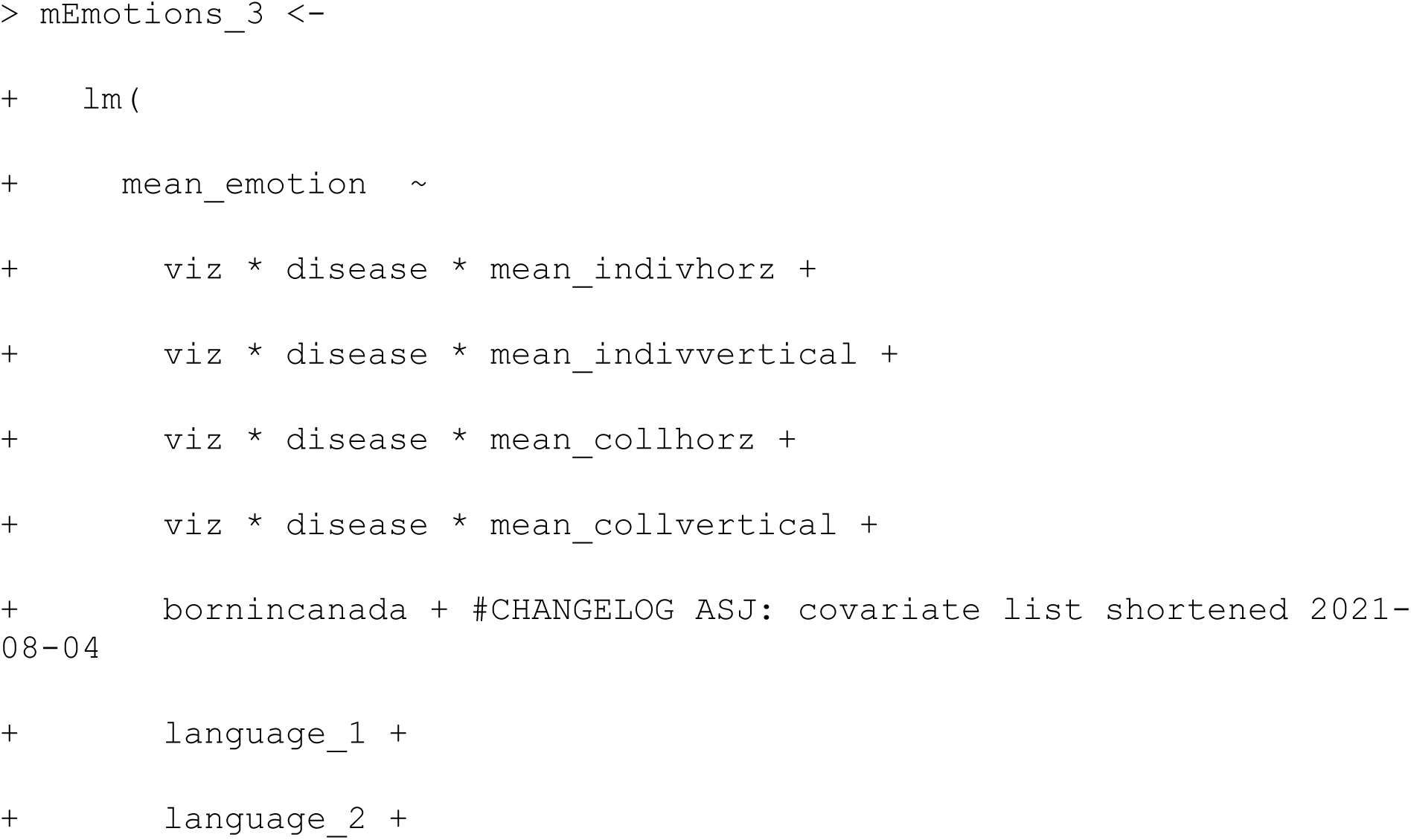
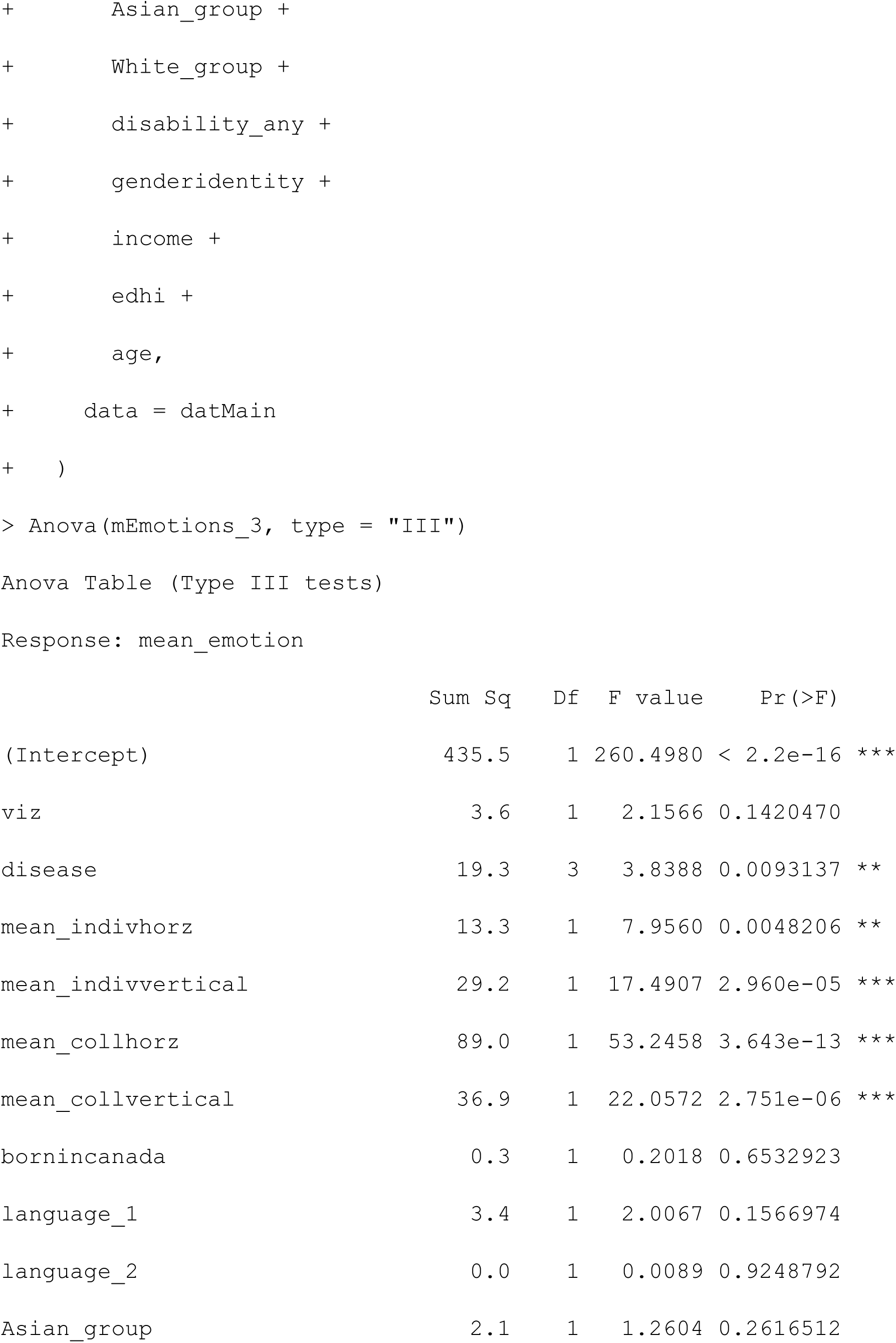
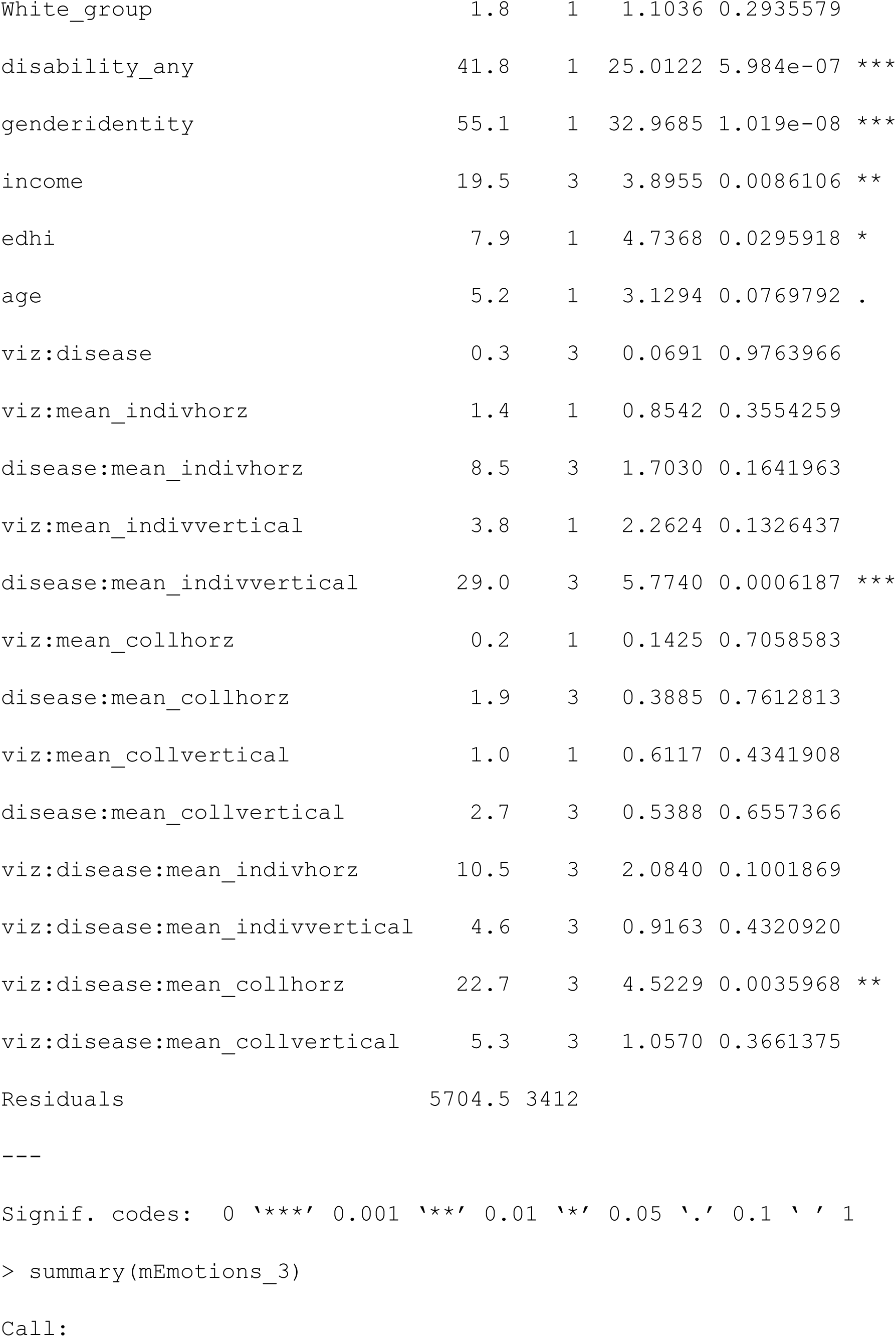
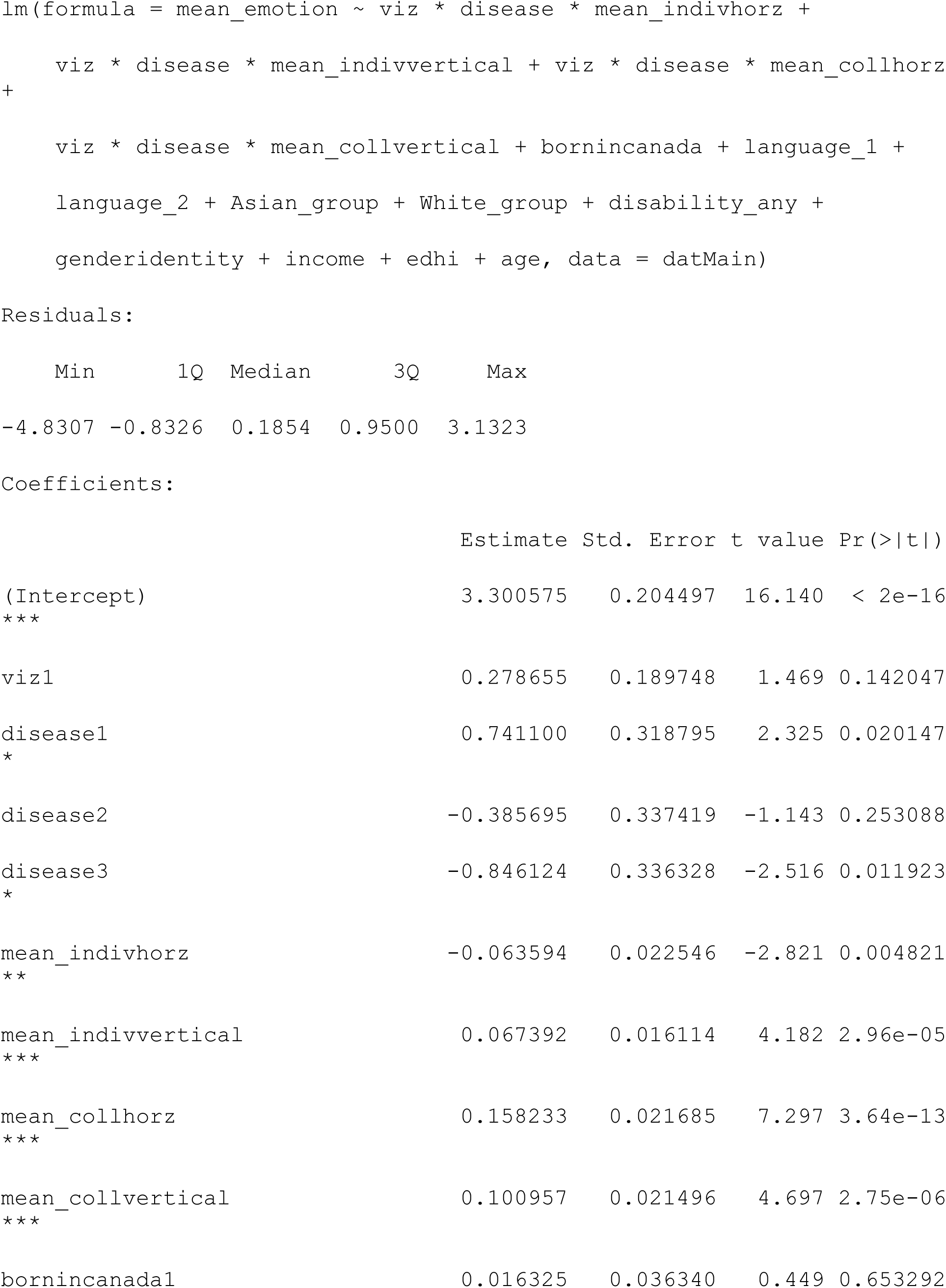
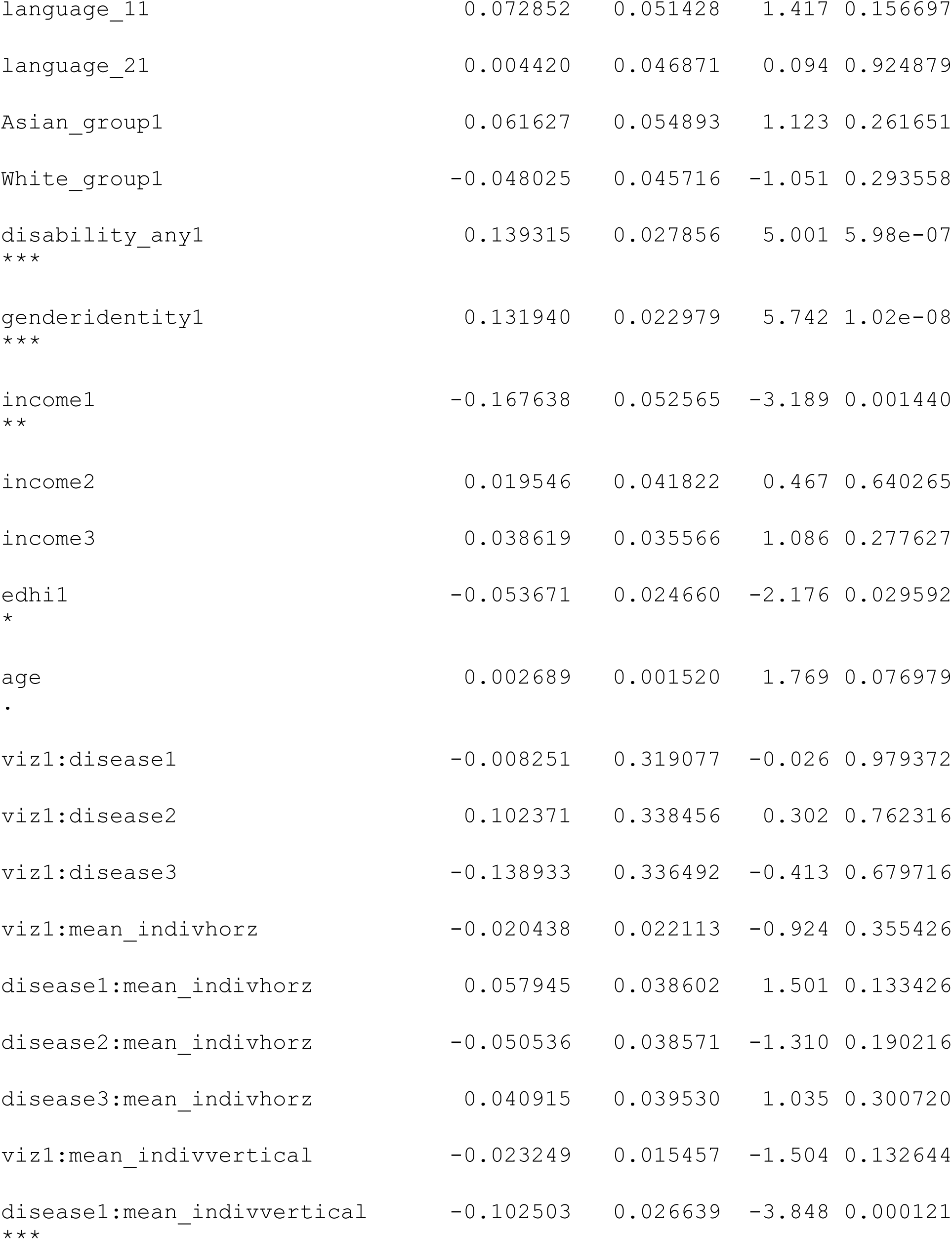
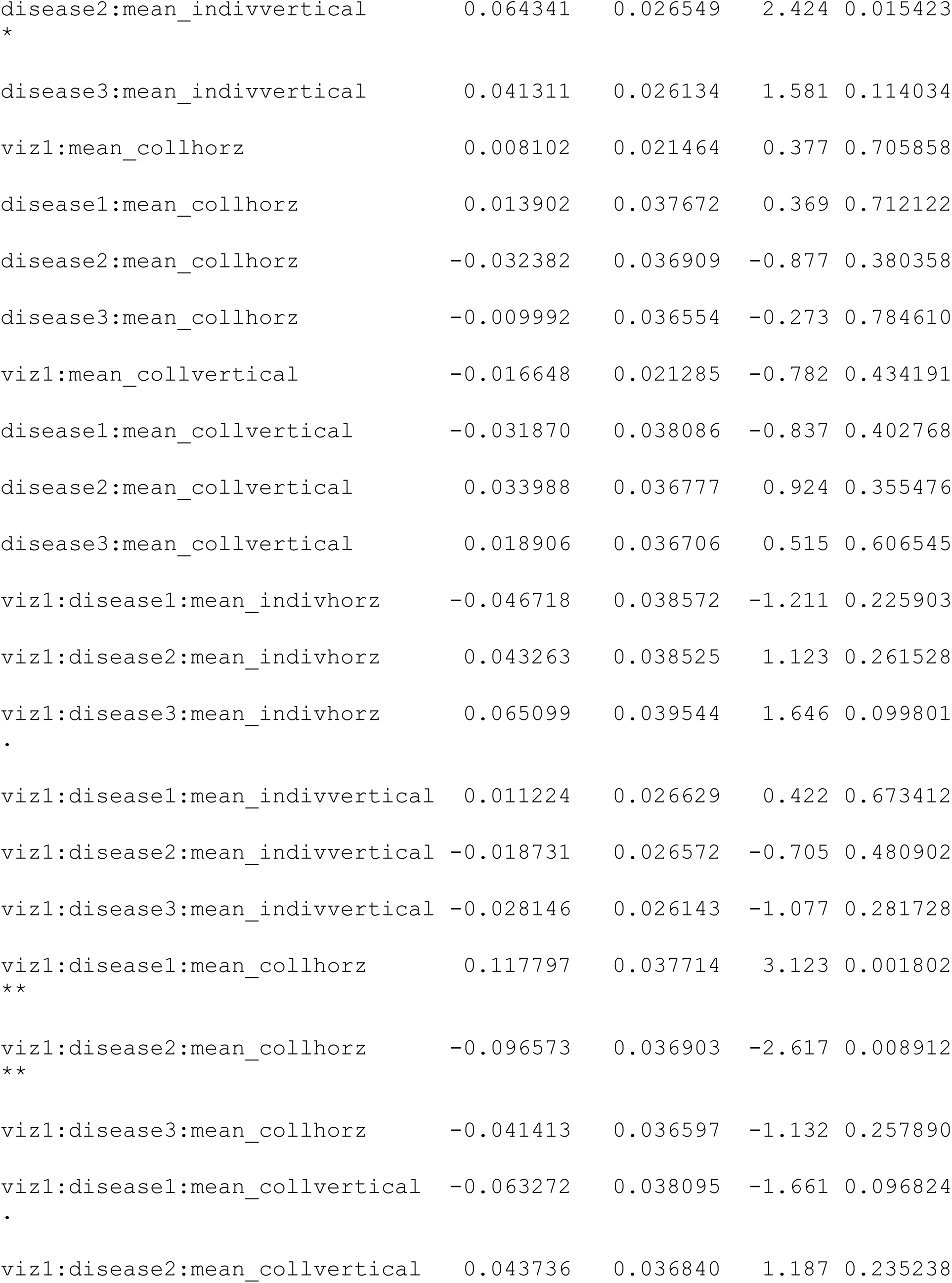
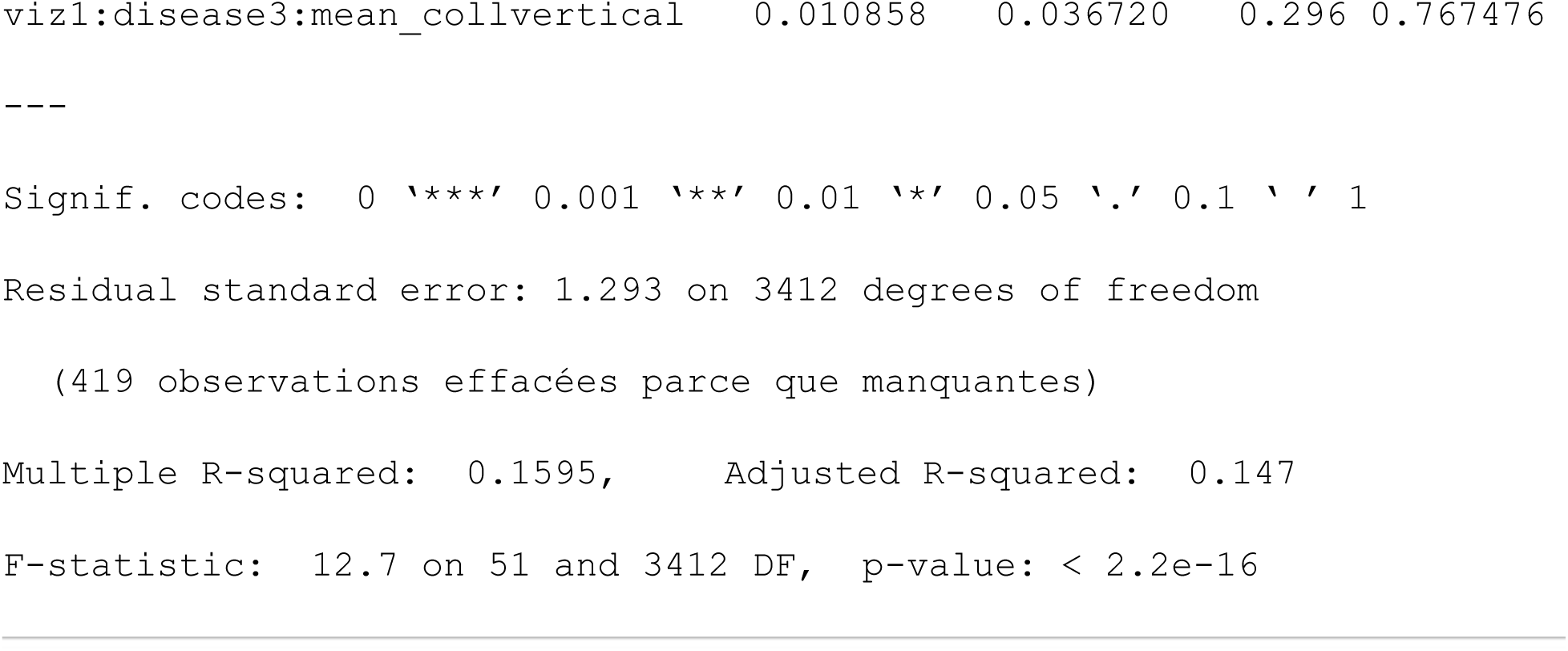

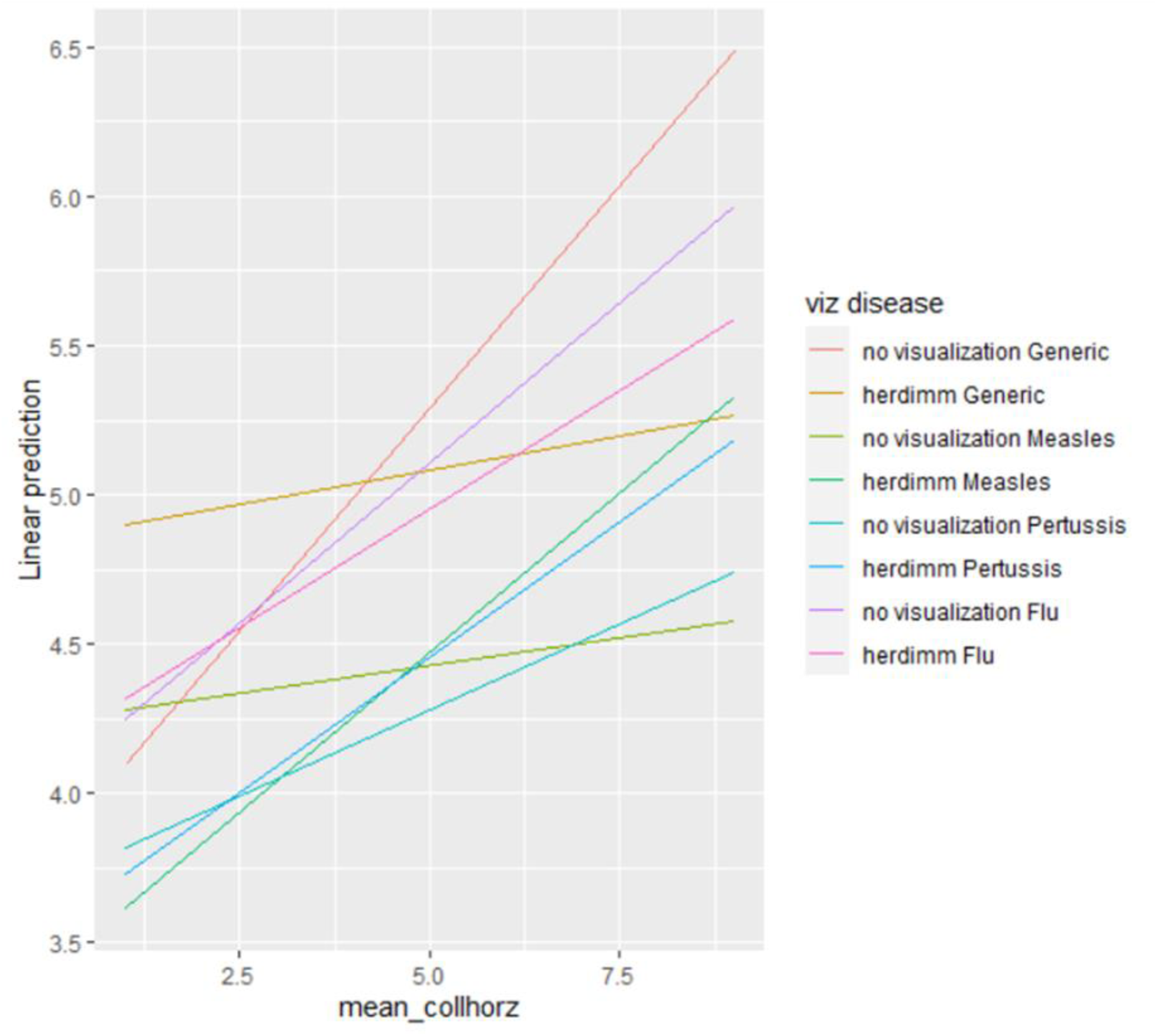

### Knowledge

#### Two-way

##### Model 1: Check for direct effects of factors without any covariates ####

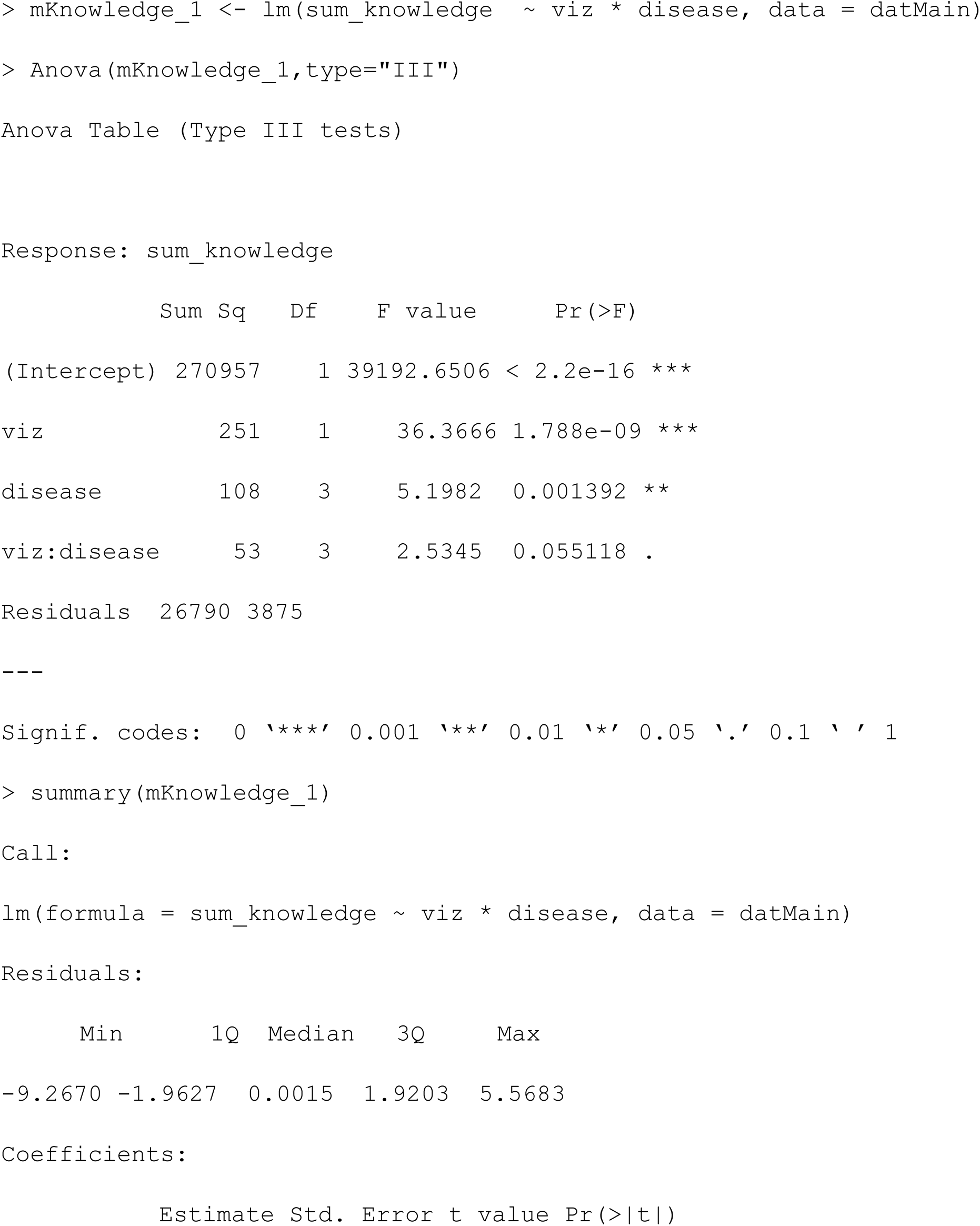
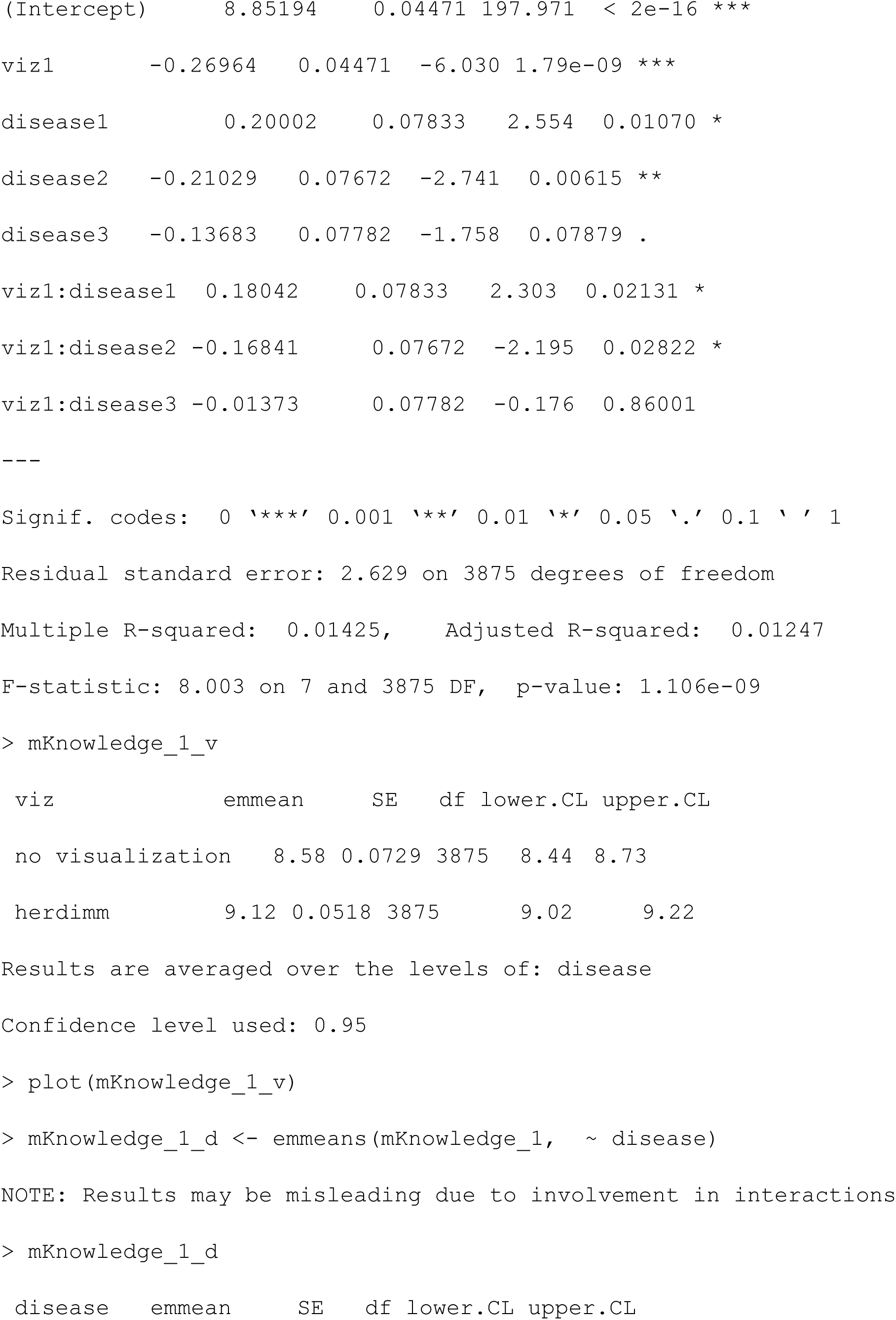
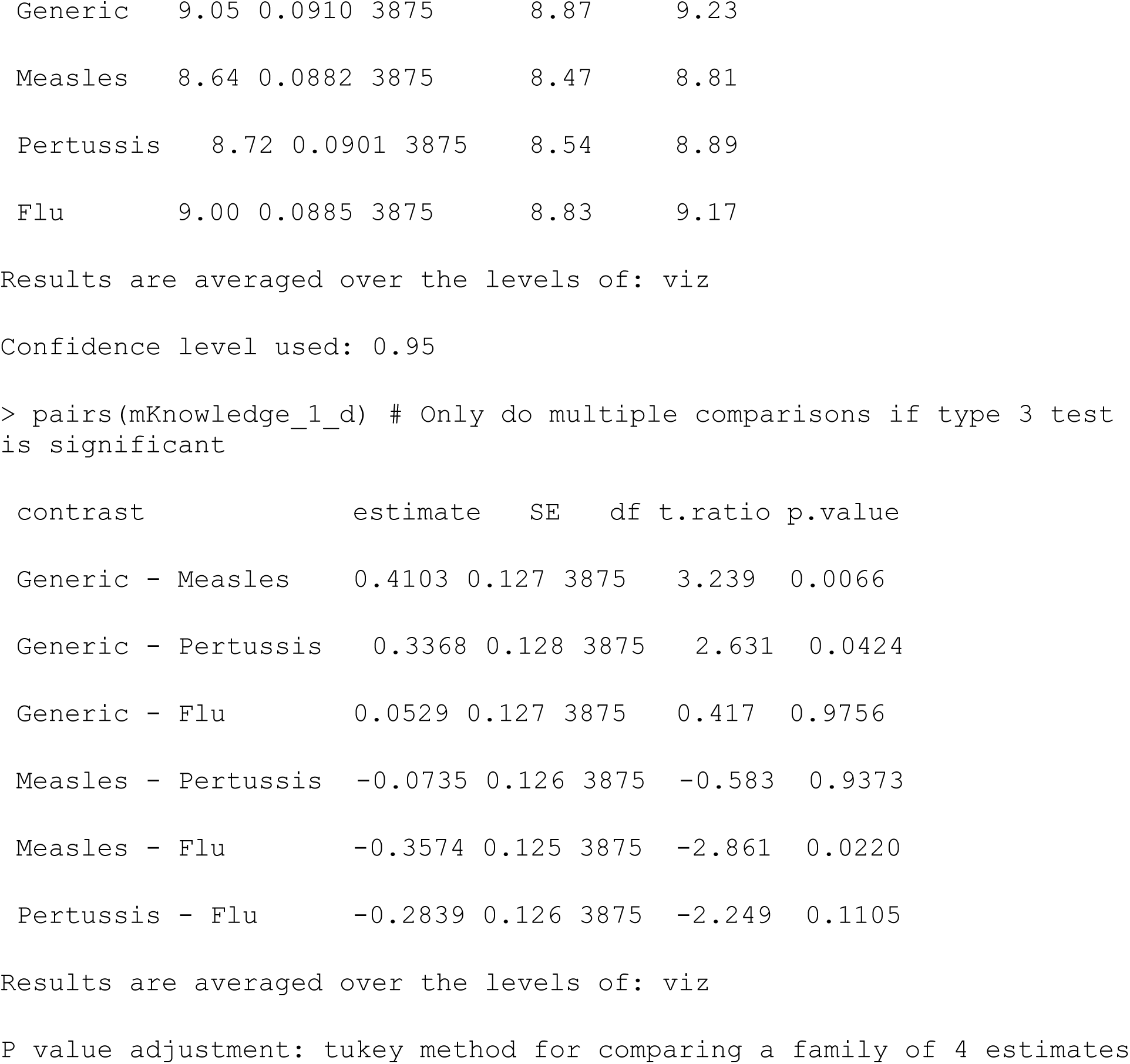

|  | Sum Sq | Df | F value | Pr(>F) |
| --- | --- | --- | --- | --- |
| (Intercept) | 270957 | 1 | 39192.6506 | < 2.2e-16 *** |
| viz | 251 | 1 | 36.3666 | 1.788e-09 *** |
| disease | 108 | 3 | 5.1982 | 0.001392 ** |
| viz:disease | 53 | 3 | 2.5345 | 0.055118 . |
| Residuals | 26790 | 3875 |  |  |

| Min | 1Q | Median | 3Q | Max |
| --- | --- | --- | --- | --- |
| -9.2670 | -1.9627 | 0.0015 | 1.9203 | 5.5683 |

Coefficients:
| Estimate | Std. Error | t value | Pr(>\|t\|) |
| --- | --- | --- | --- |

| viz | emmean | SE | df | lower.CL | upper.CL |
| --- | --- | --- | --- | --- | --- |
| no visualization | 8.58 | 0.0729 | 3875 | 8.44 | 8.73 |
| herdimm | 9.12 | 0.0518 | 3875 | 9.02 | 9.22 |

| disease | emmean | SE | df | lower.CL | upper.CL |
| --- | --- | --- | --- | --- | --- |
| Generic | 9.05 | 0.0910 | 3875 | 8.87 | 9.23 |
| Measles | 8.64 | 0.0882 | 3875 | 8.47 | 8.81 |
| Pertussis | 8.72 | 0.0901 | 3875 | 8.54 | 8.89 |
| Flu | 9.00 | 0.0885 | 3875 | 8.83 | 9.17 |

| contrast | estimate | SE | df | t.ratio | p.value |
| --- | --- | --- | --- | --- | --- |
| Generic - Measles | 0.4103 | 0.127 | 3875 | 3.239 | 0.0066 |
| Generic - Pertussis | 0.3368 | 0.128 | 3875 | 2.631 | 0.0424 |
| Generic - Flu | 0.0529 | 0.127 | 3875 | 0.417 | 0.9756 |
| Measles - Pertussis | -0.0735 | 0.126 | 3875 | -0.583 | 0.9373 |
| Measles - Flu | -0.3574 | 0.125 | 3875 | -2.861 | 0.0220 |
| Pertussis - Flu | -0.2839 | 0.126 | 3875 | -2.249 | 0.1105 |

Results are averaged over the levels of: viz
P value adjustment: tukey method for comparing a family of 4 estimates

##### Model 2: Check for direct effects of factors with adjustment for other covariates

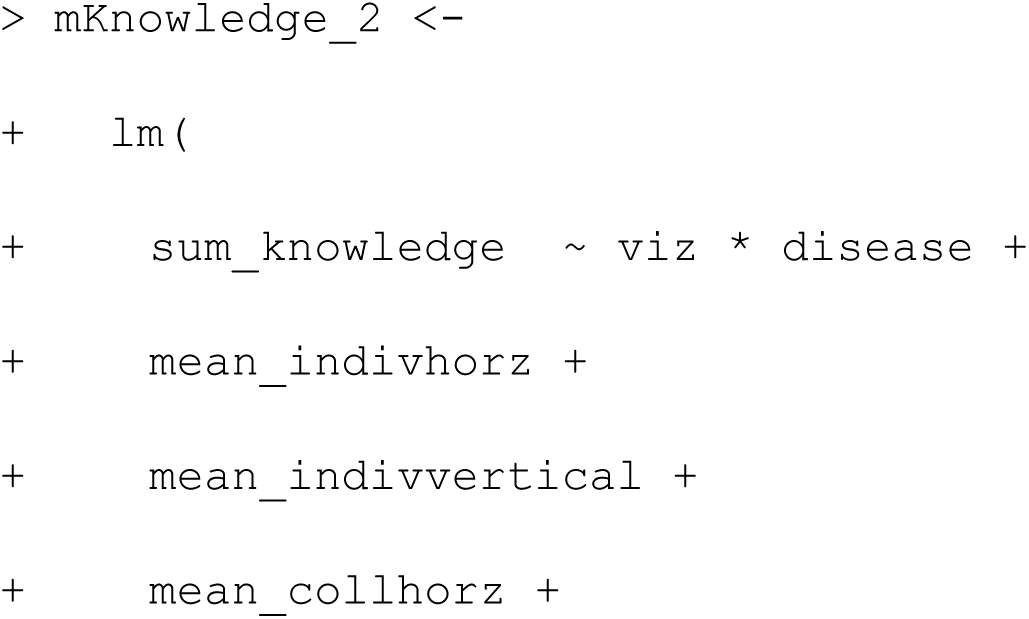
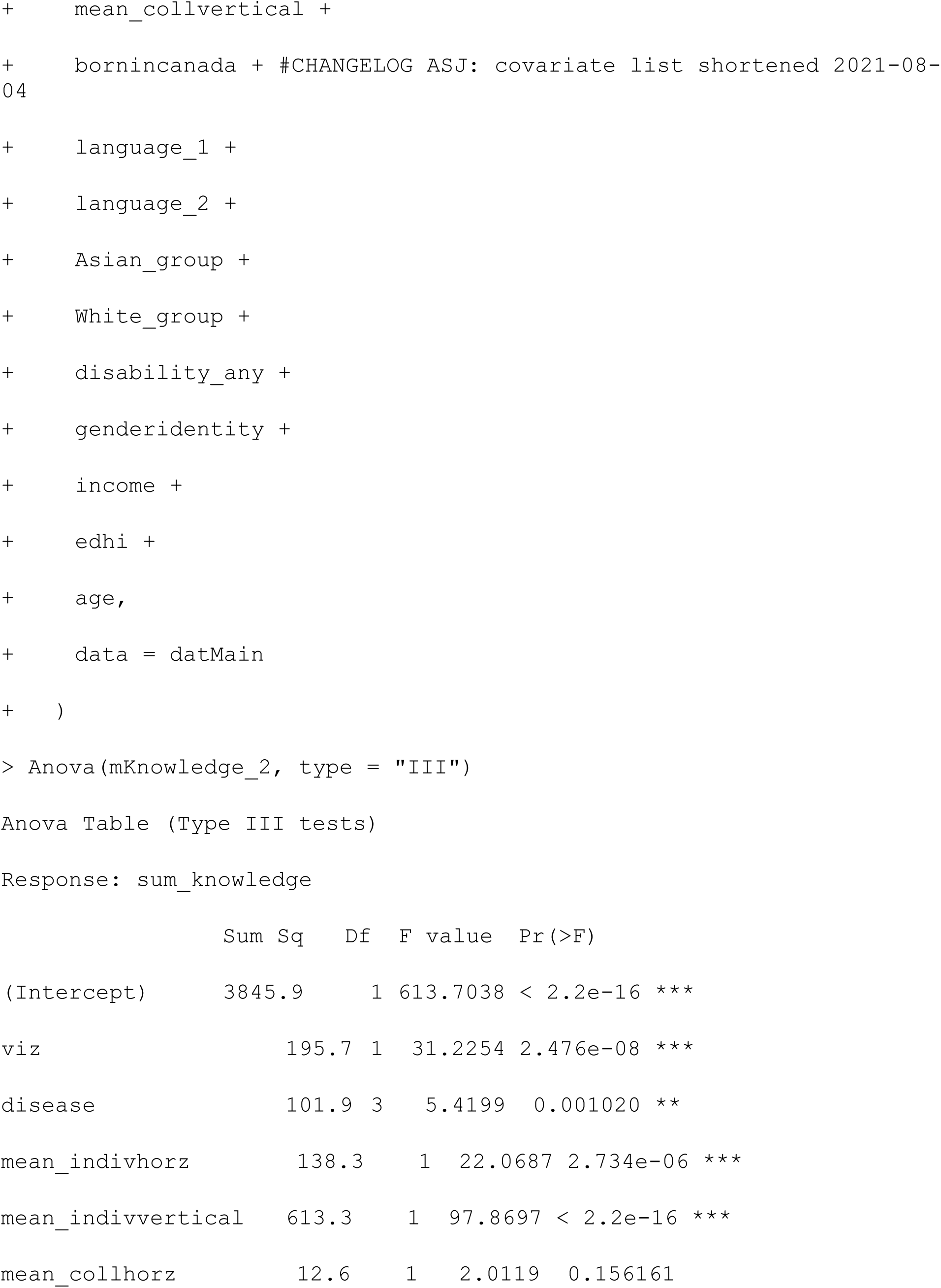
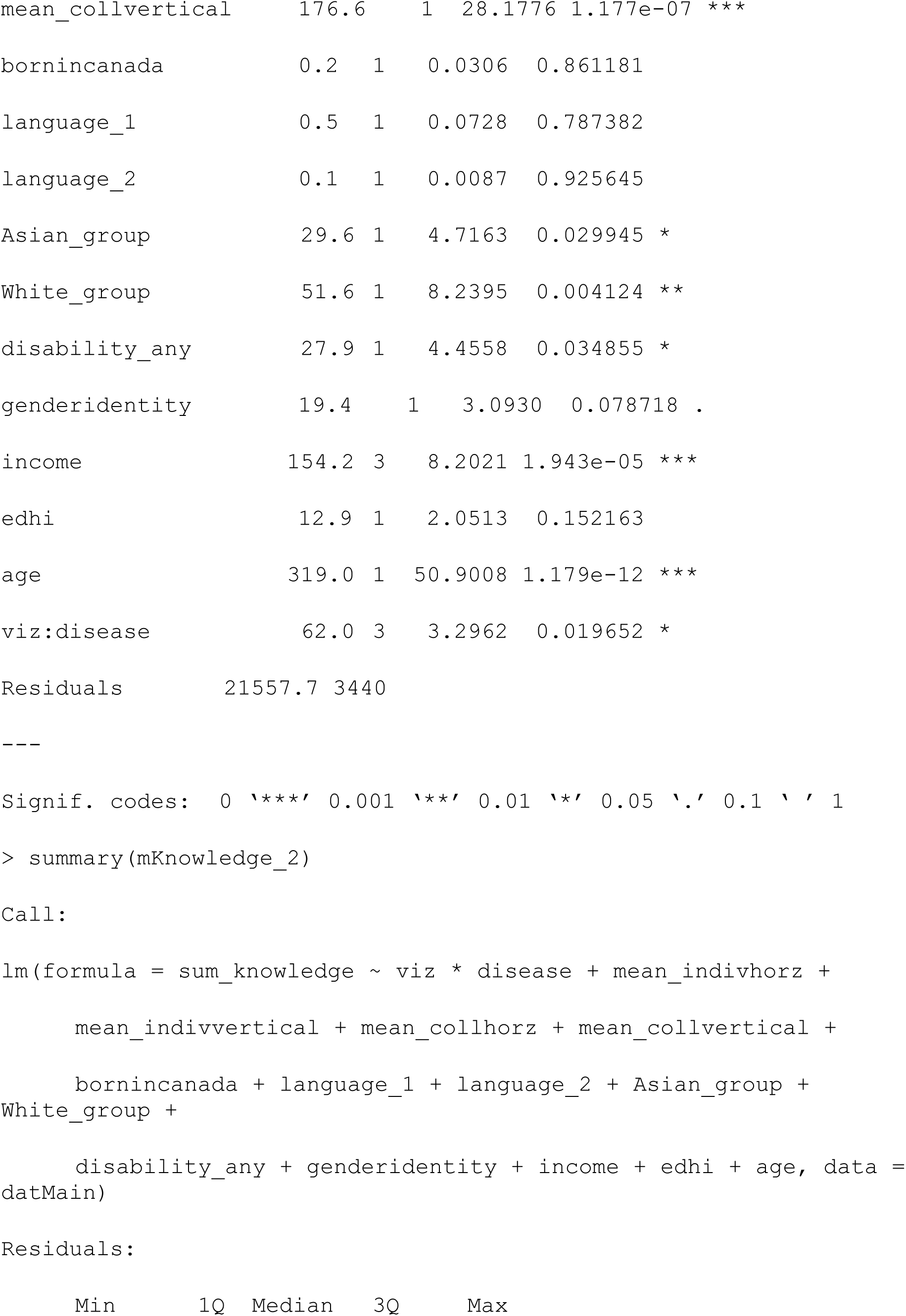
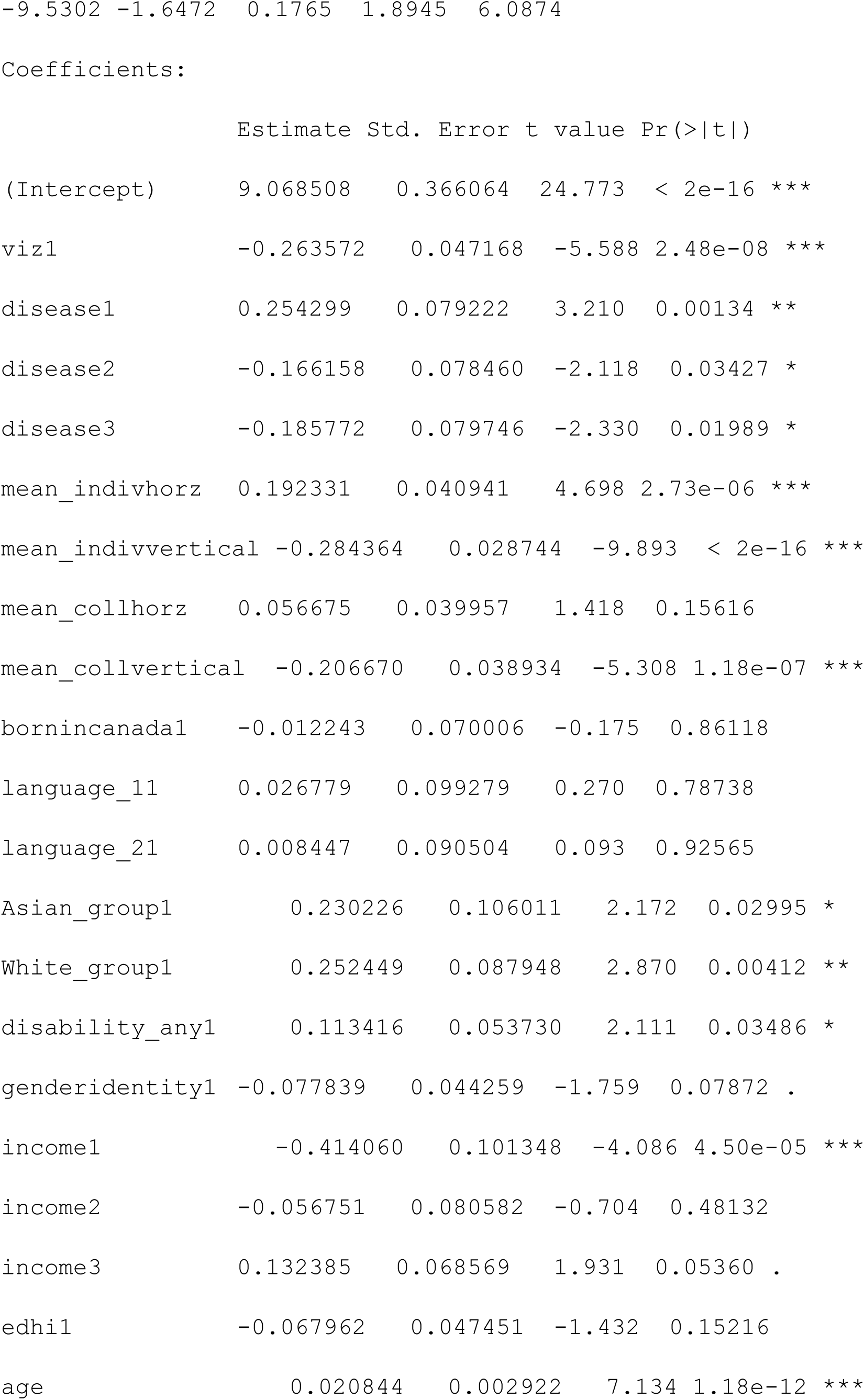
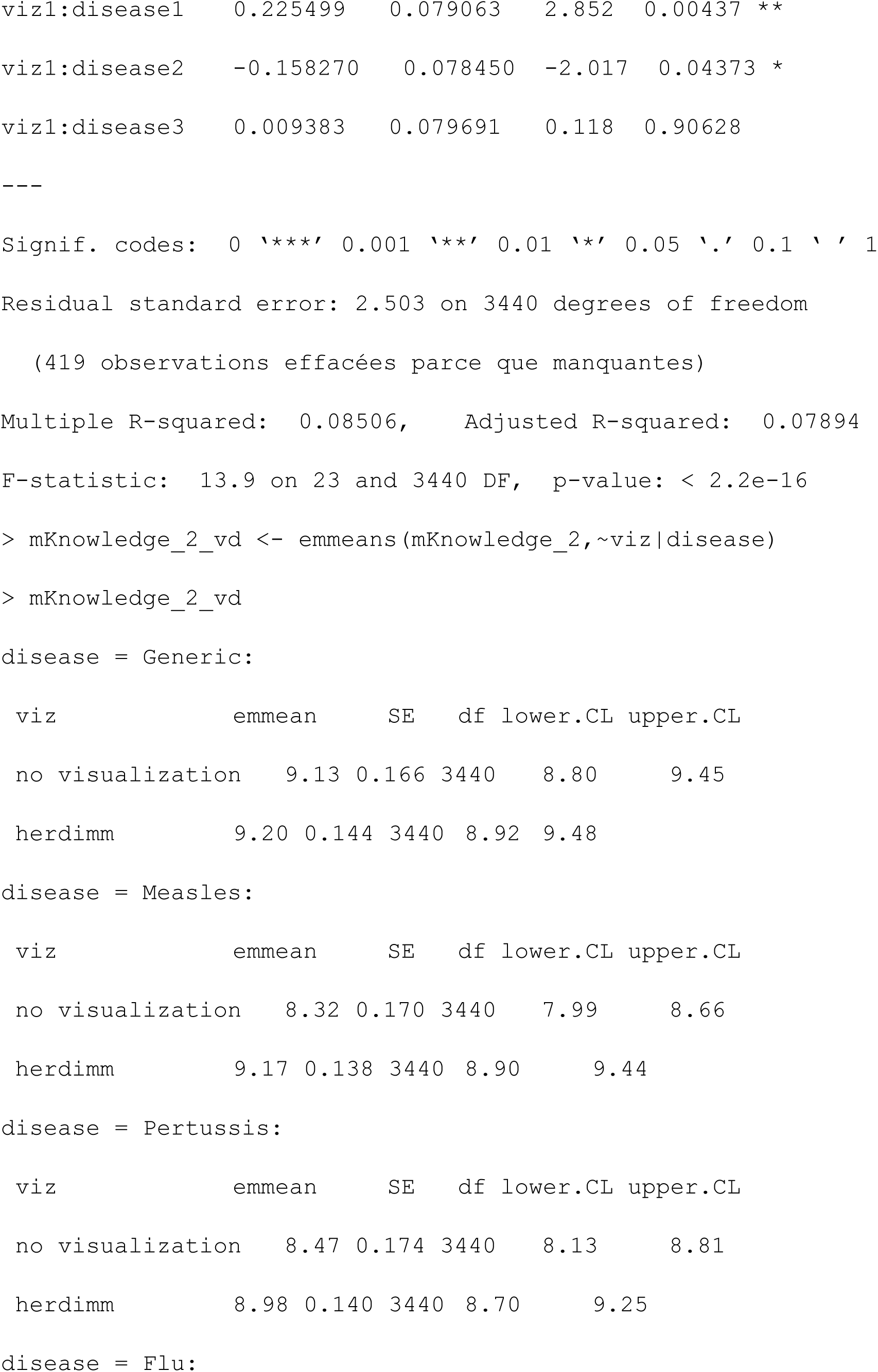
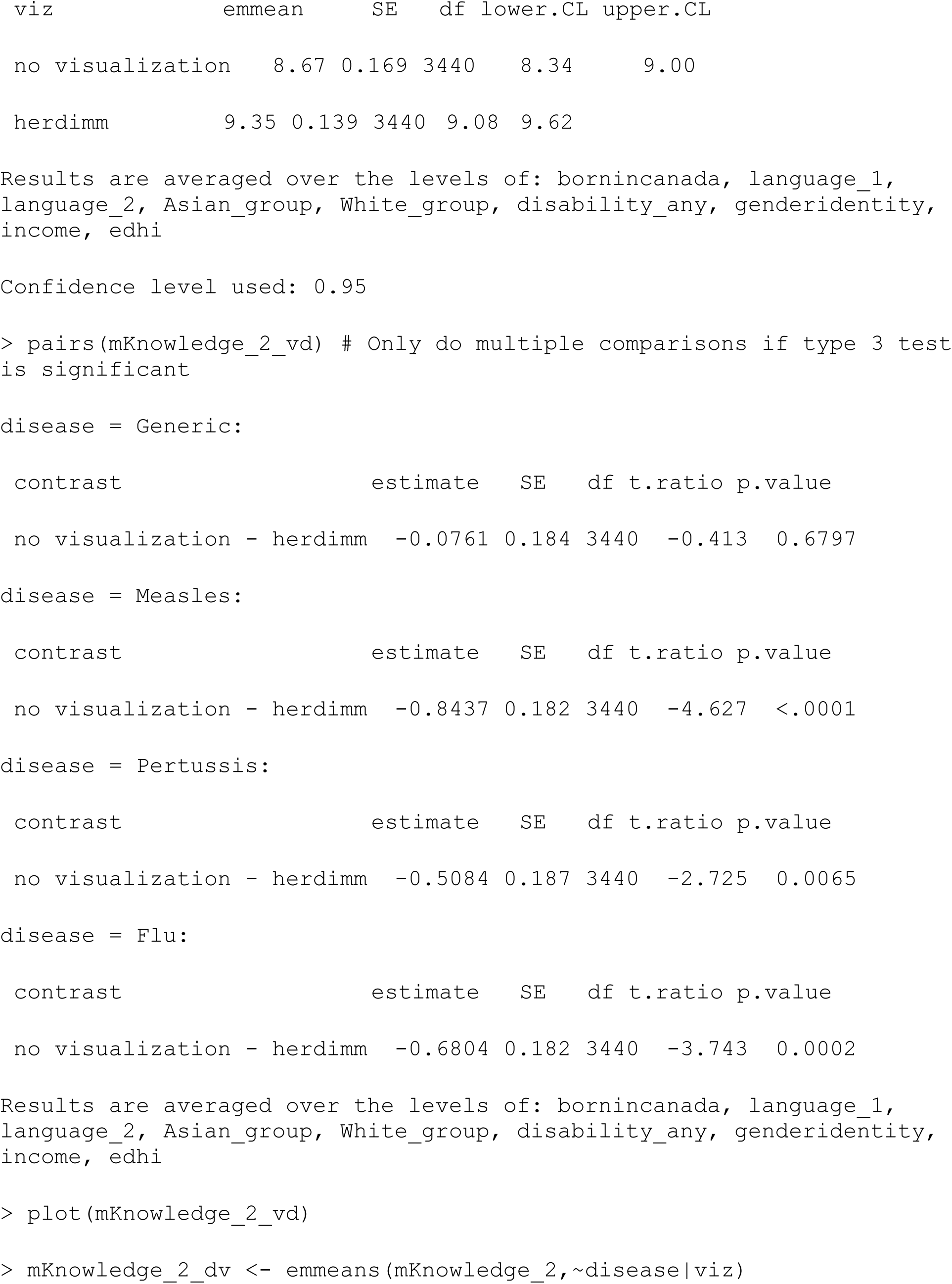
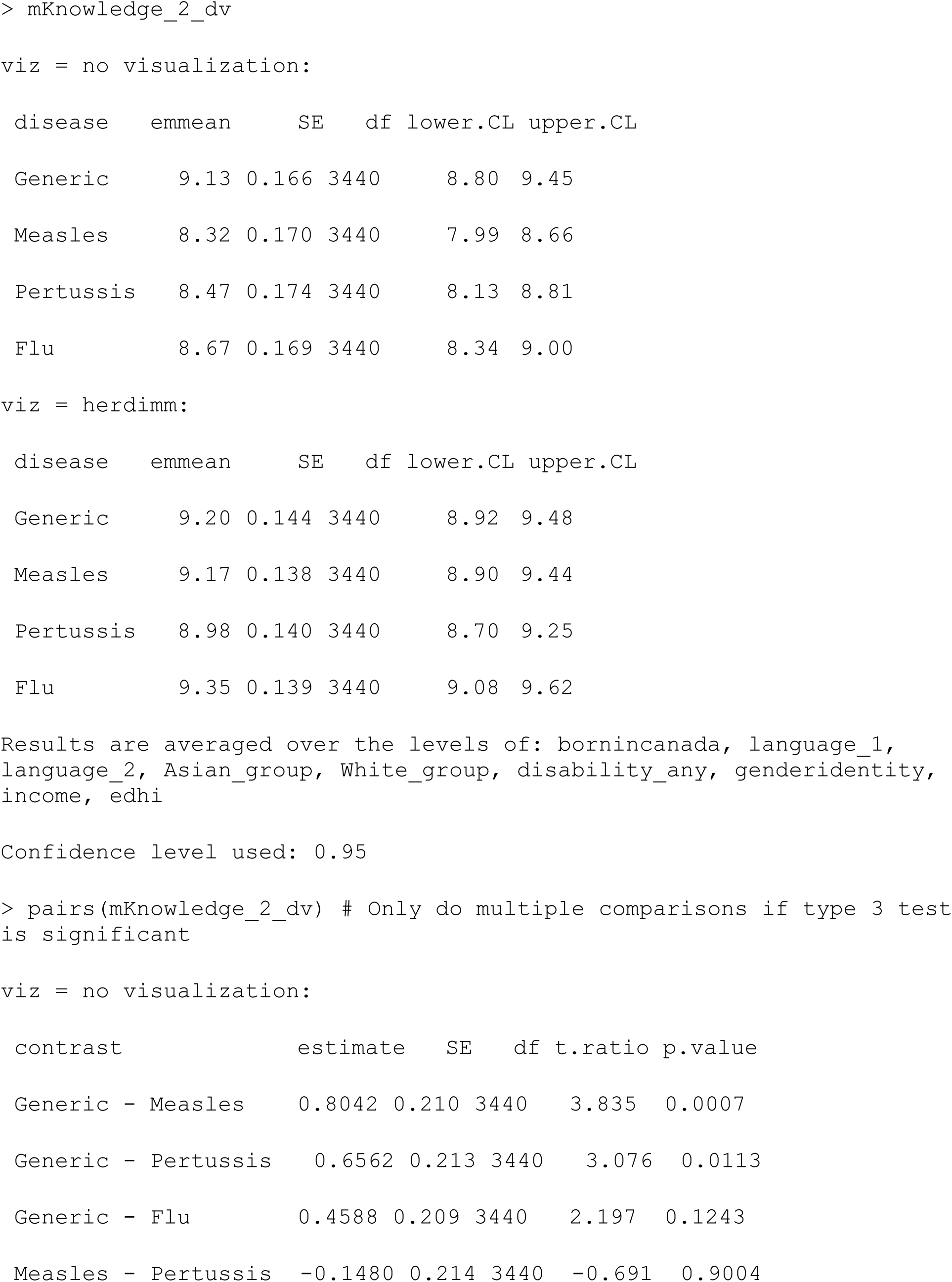
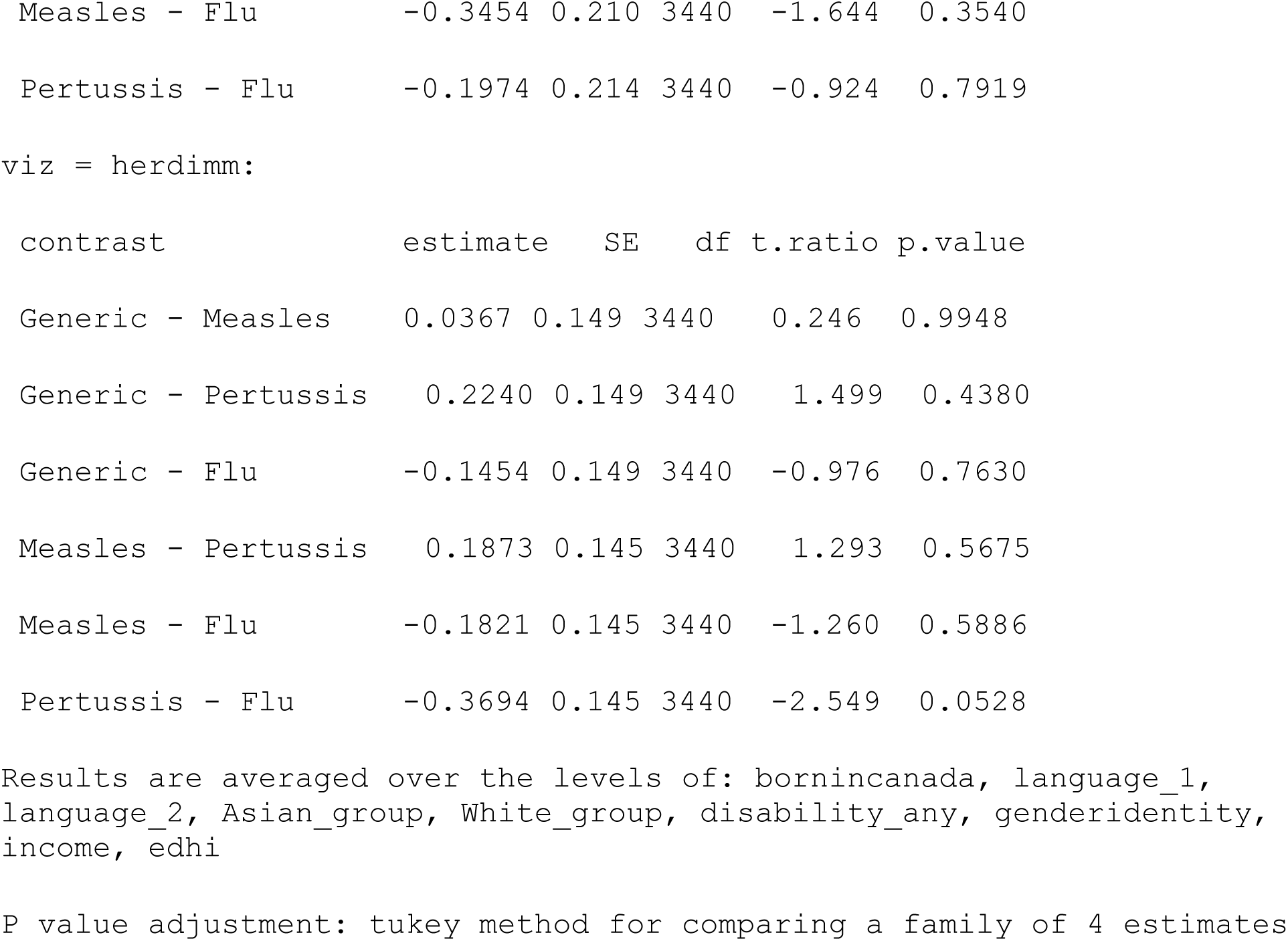

##### Model 3: Check for moderating effects of individualism & collectivism with adjustment for other covariates

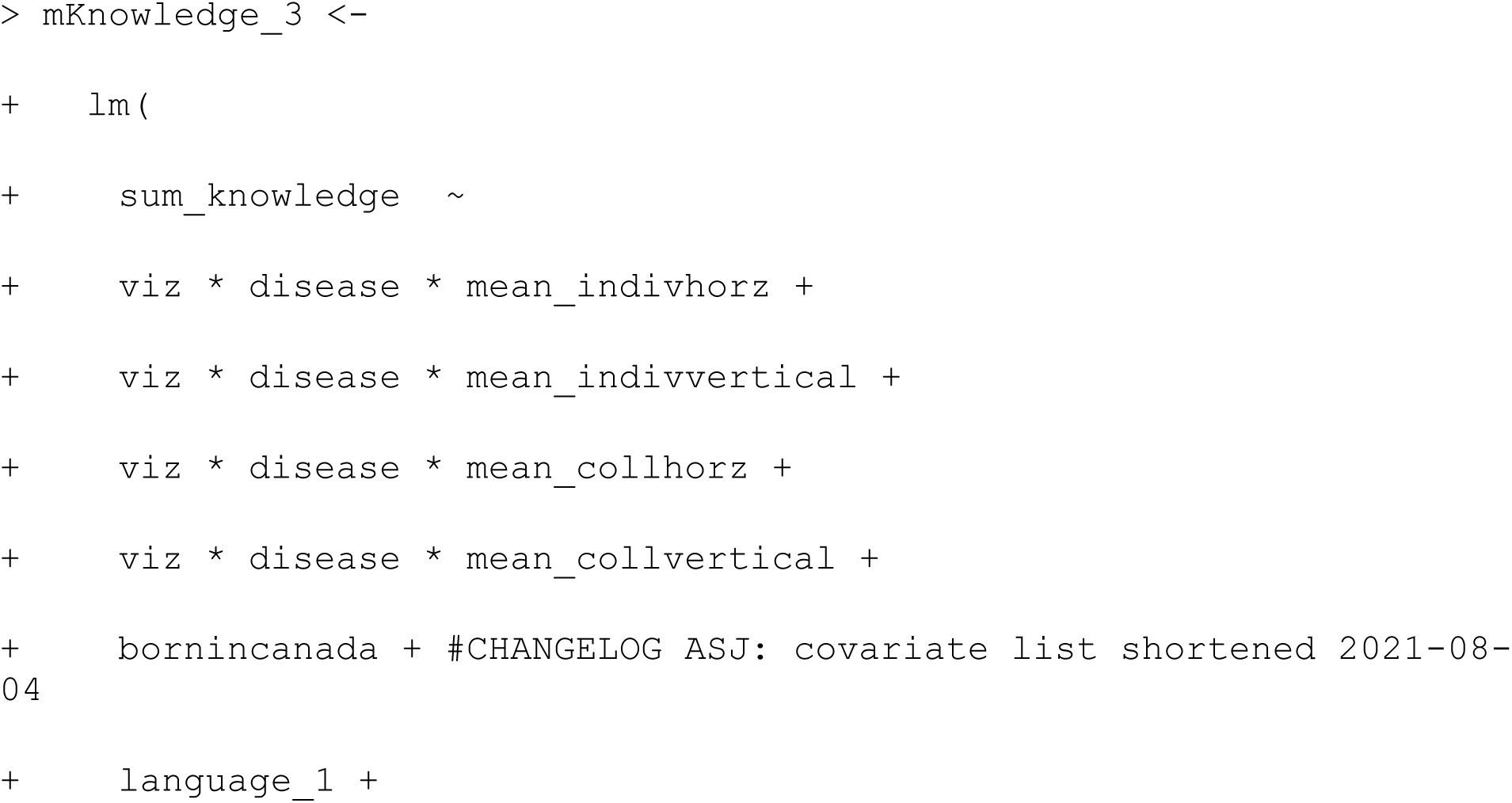
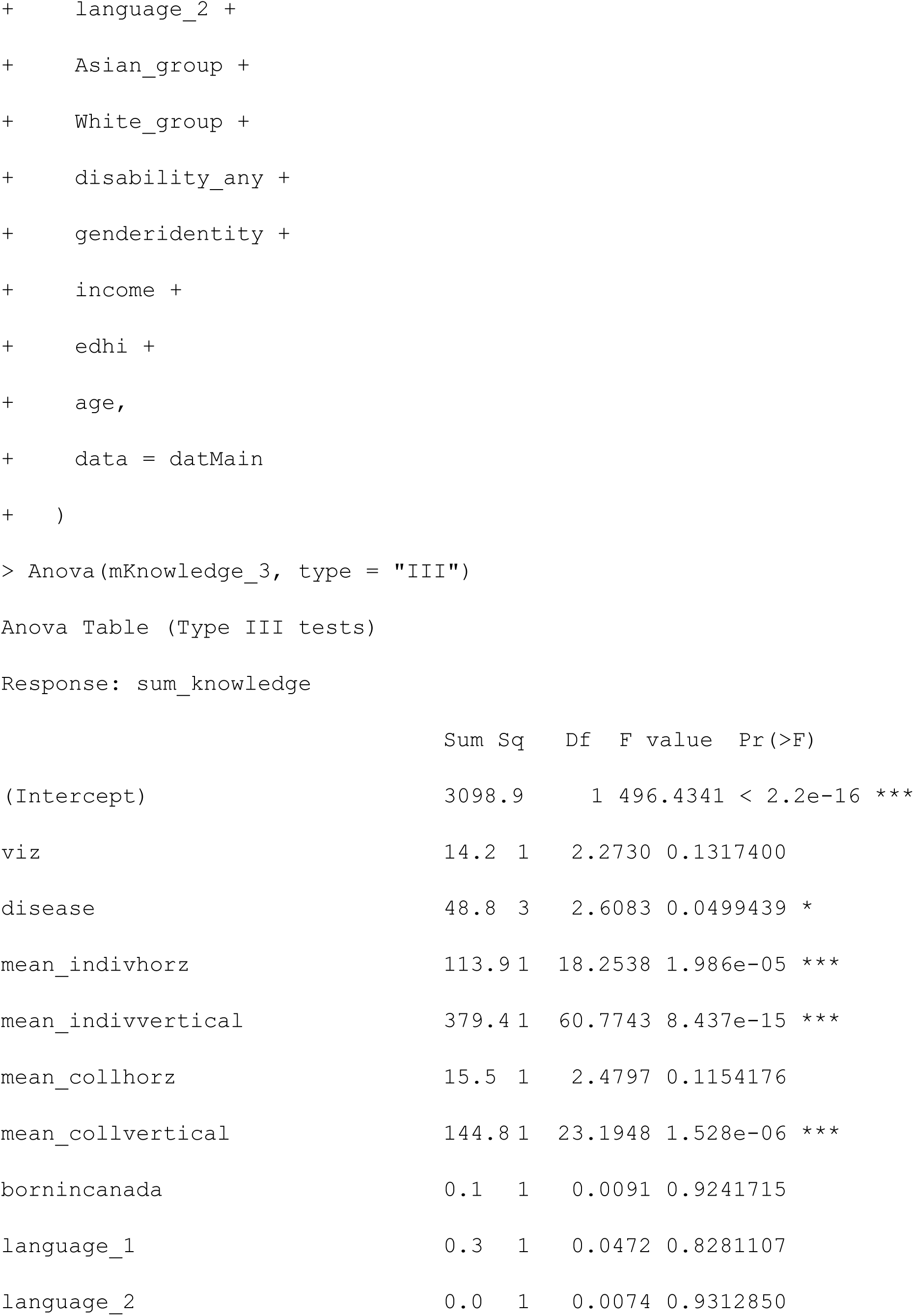
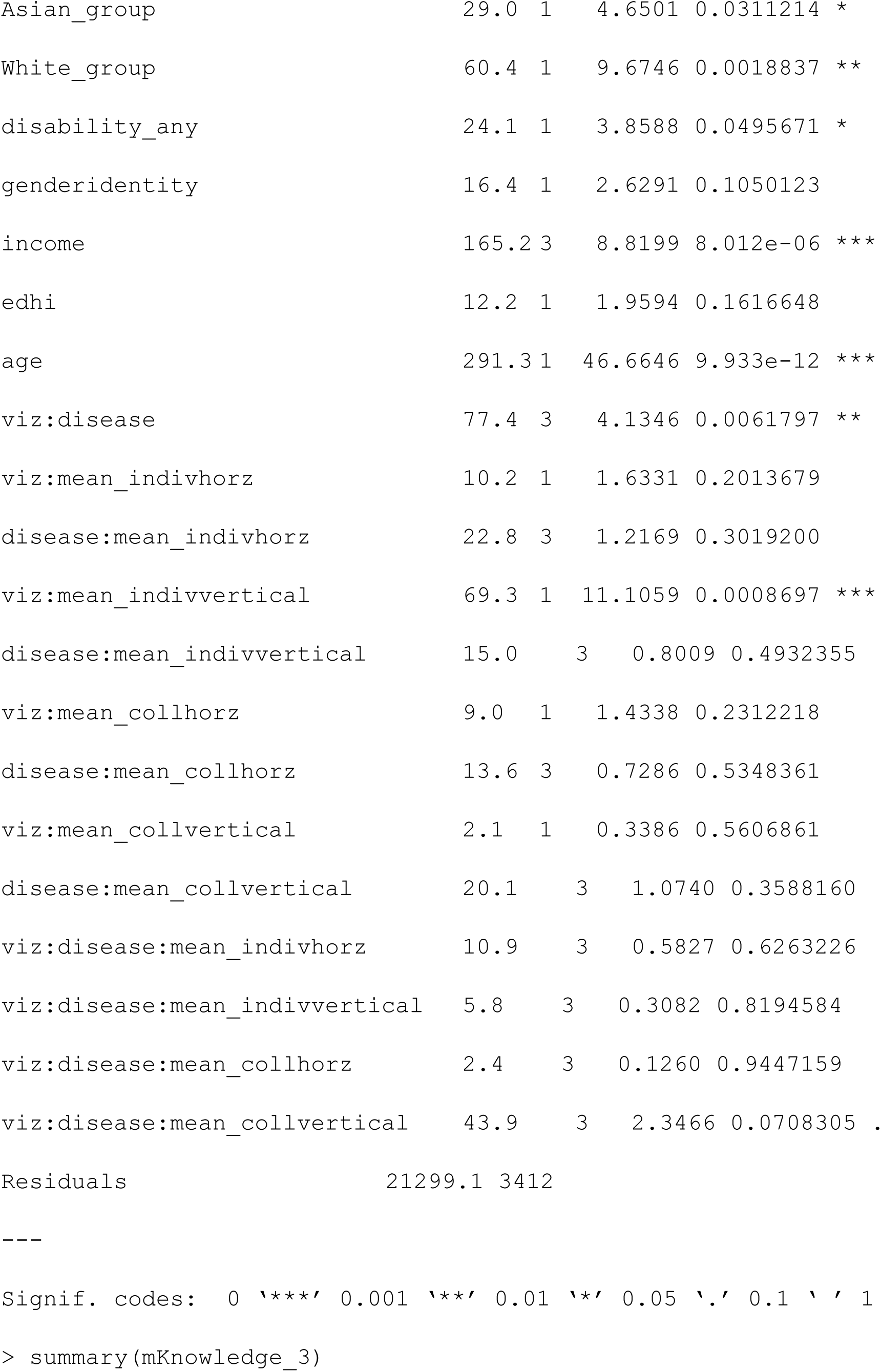
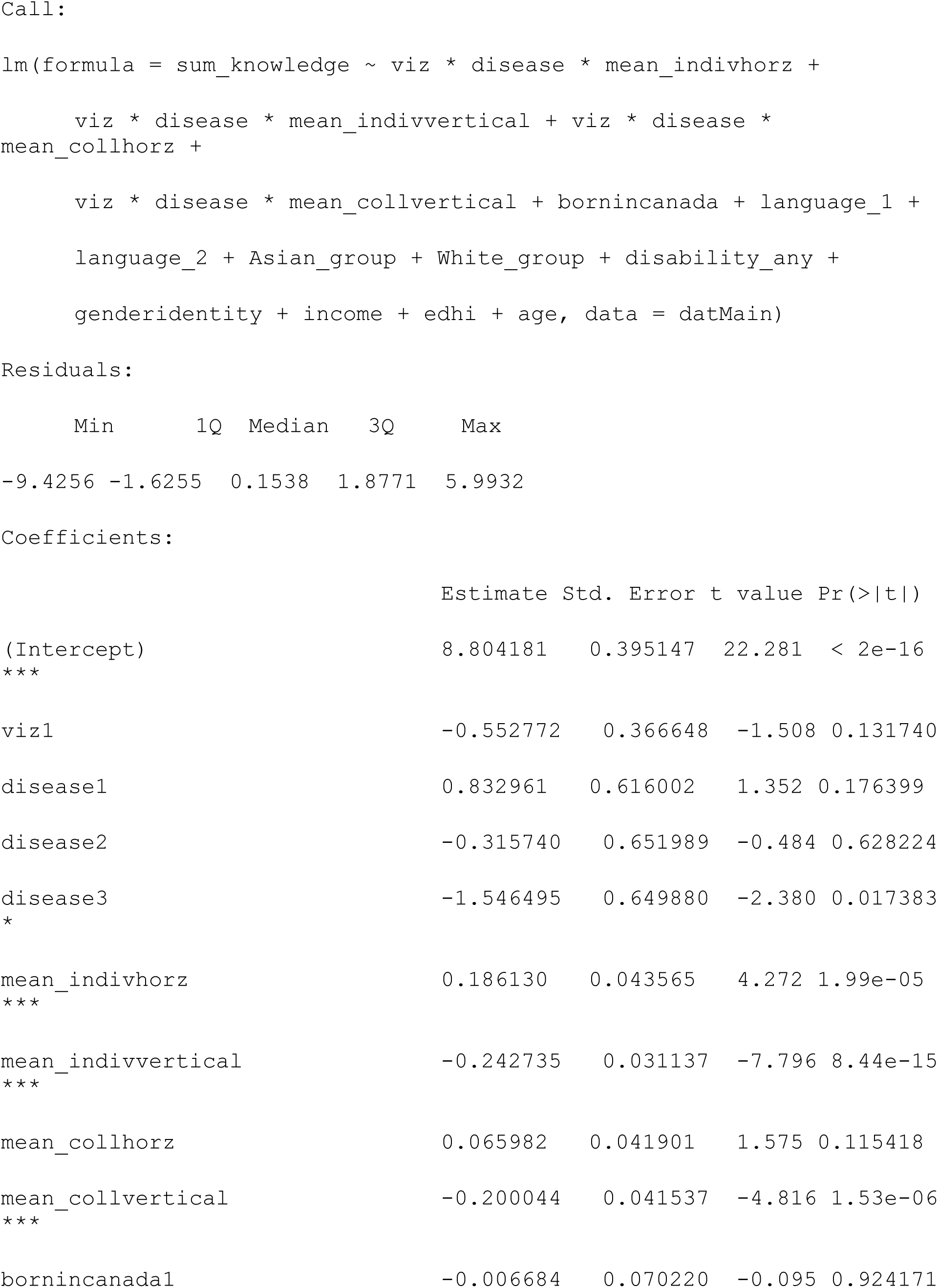
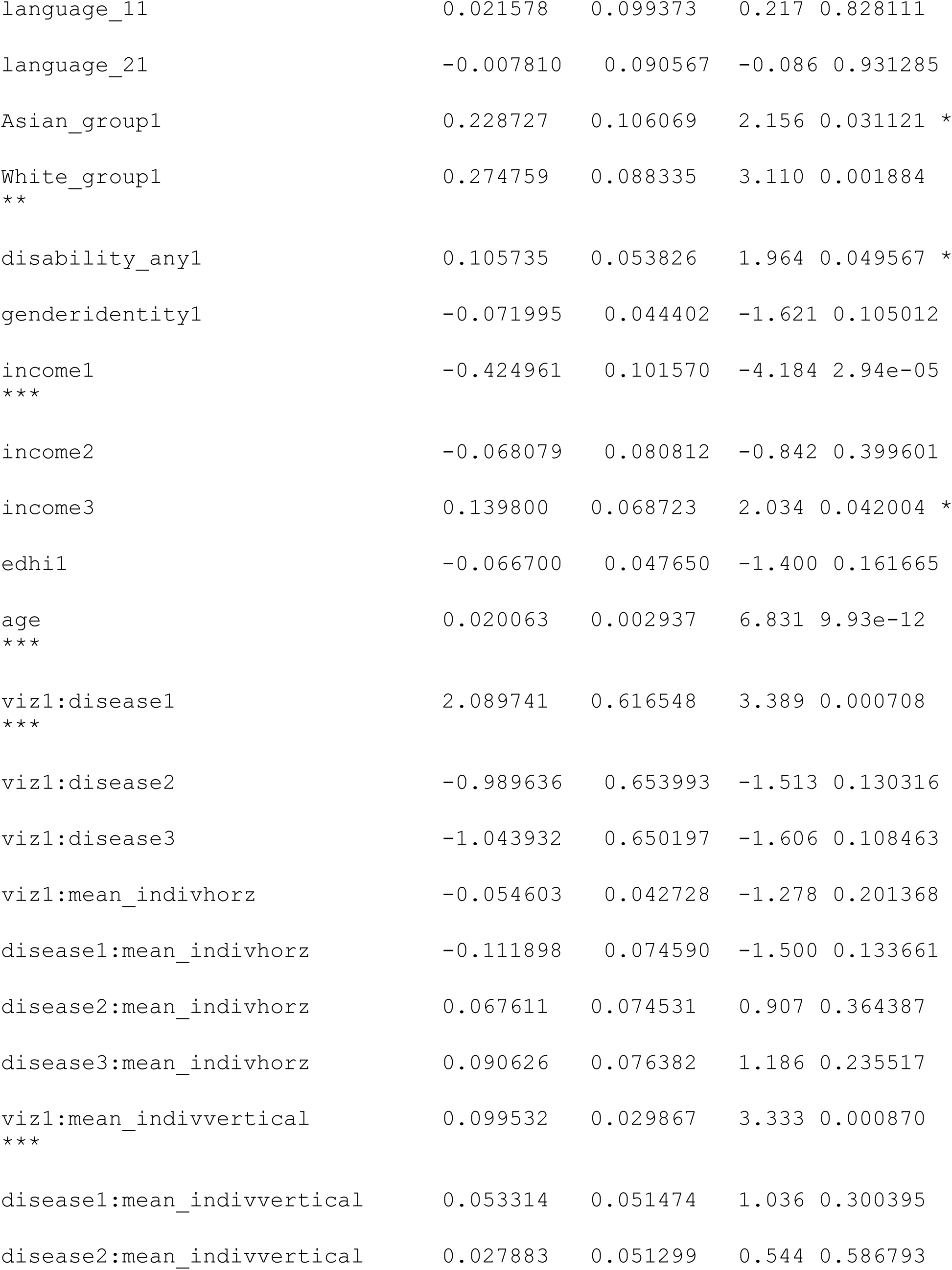
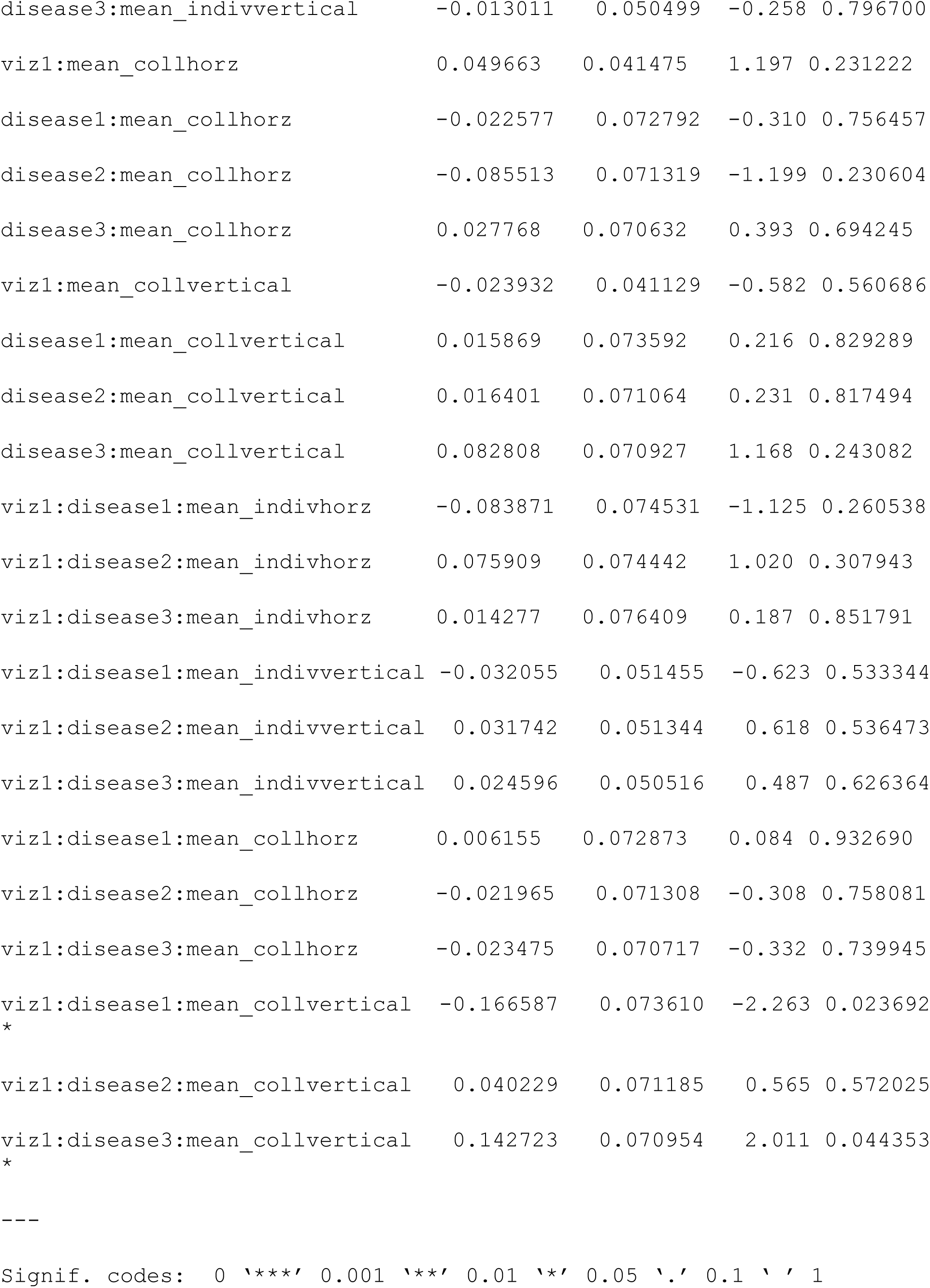
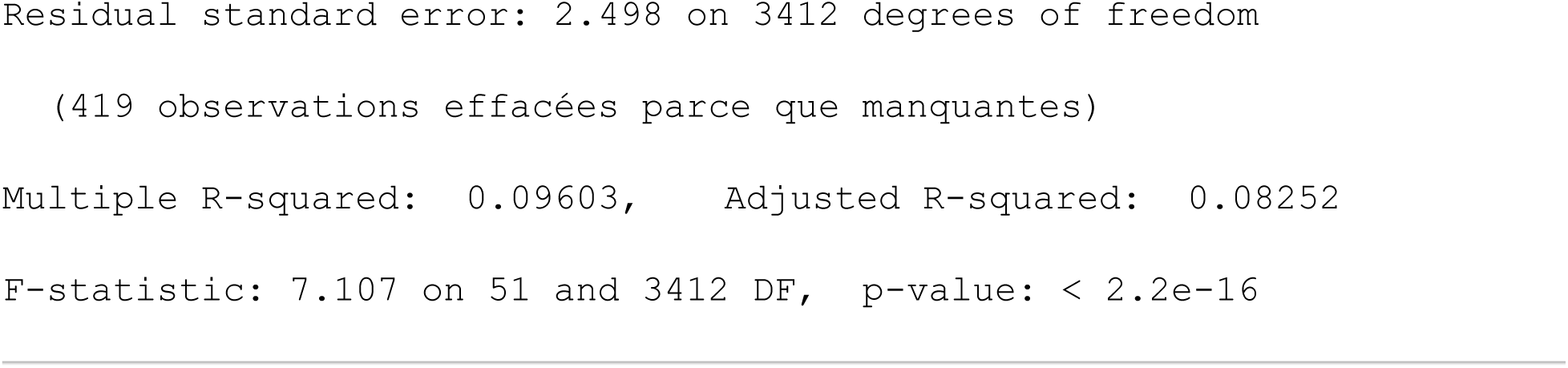

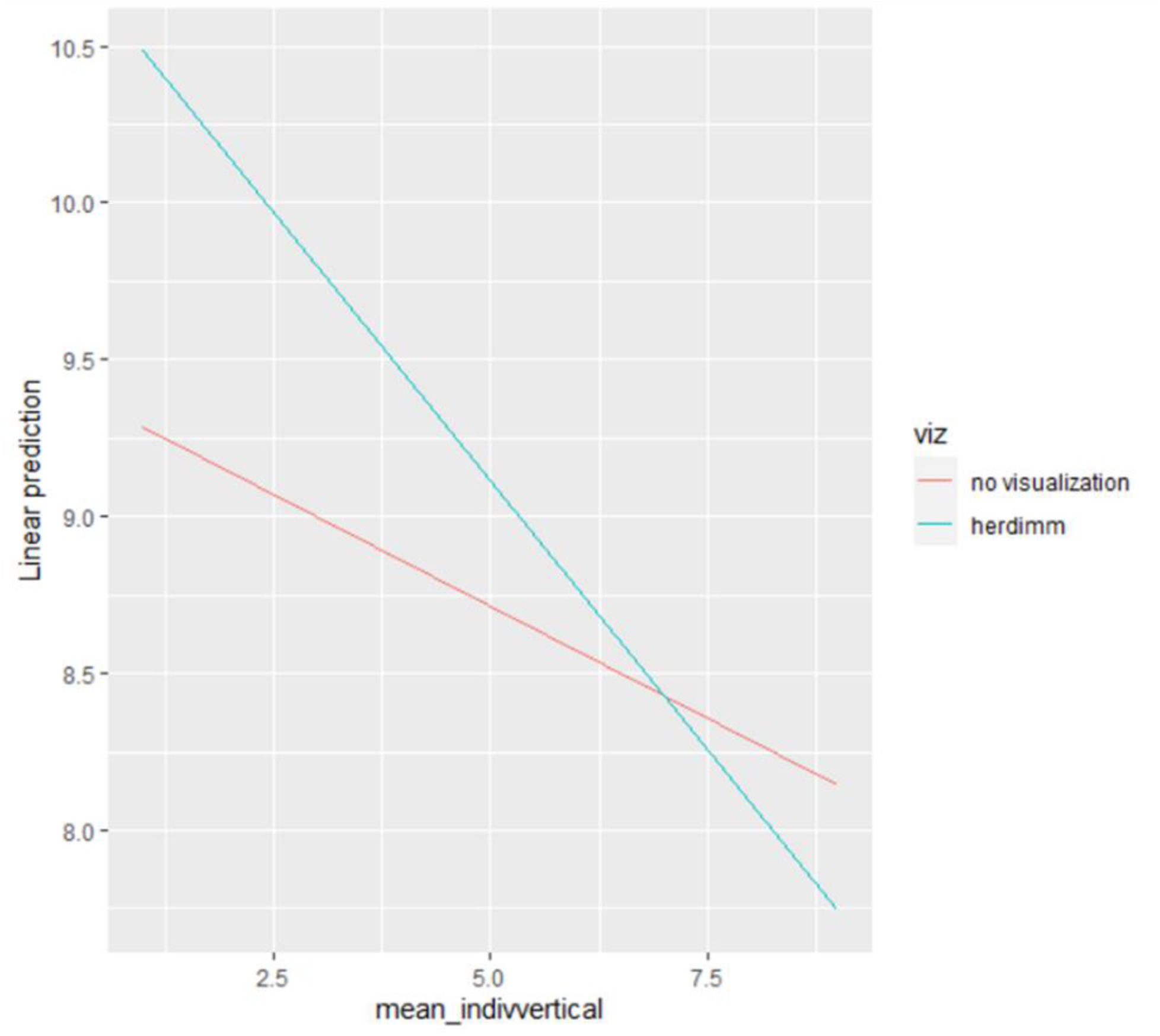

### Vax intention

#### Two-way

##### Model 1

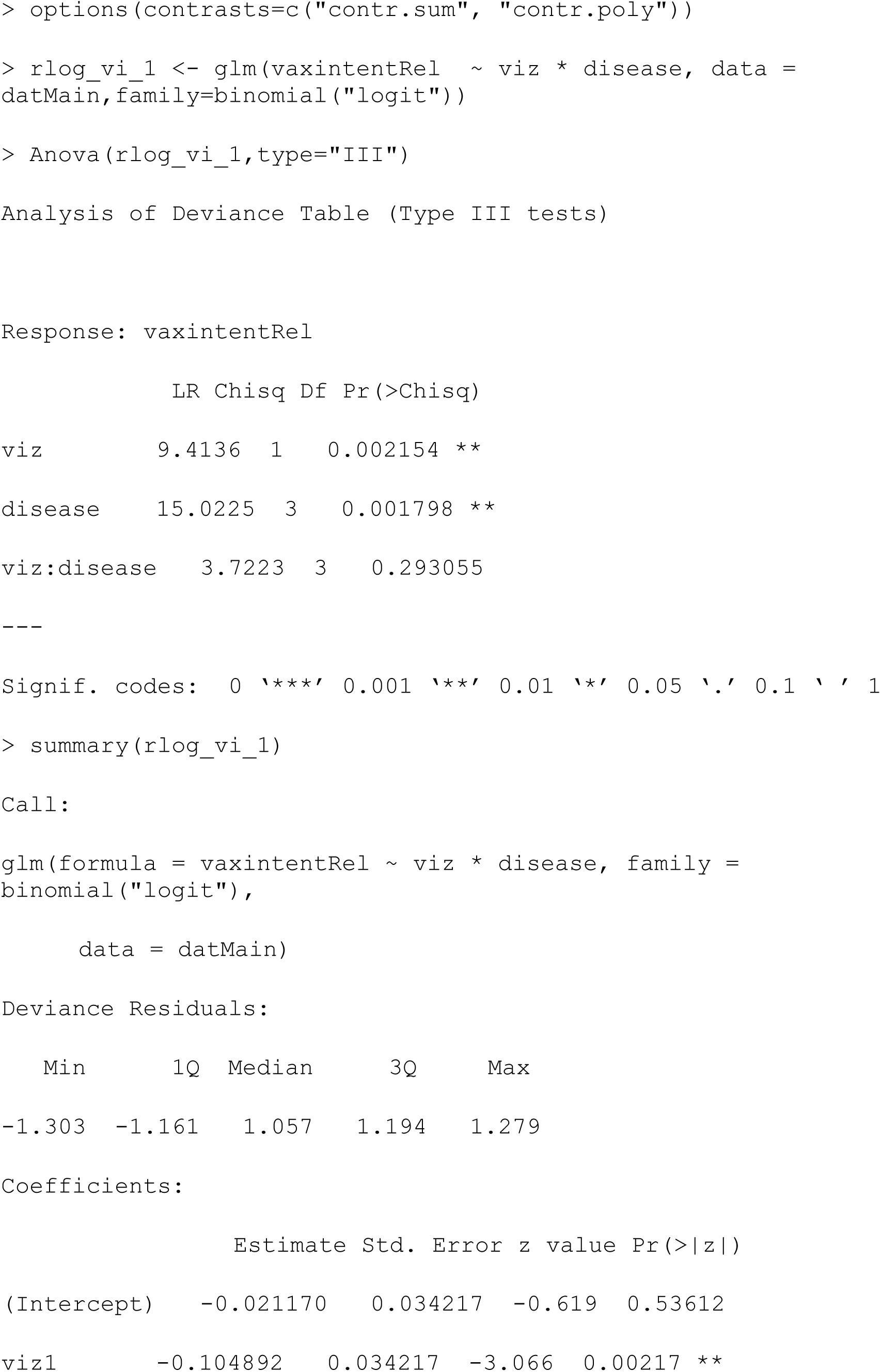
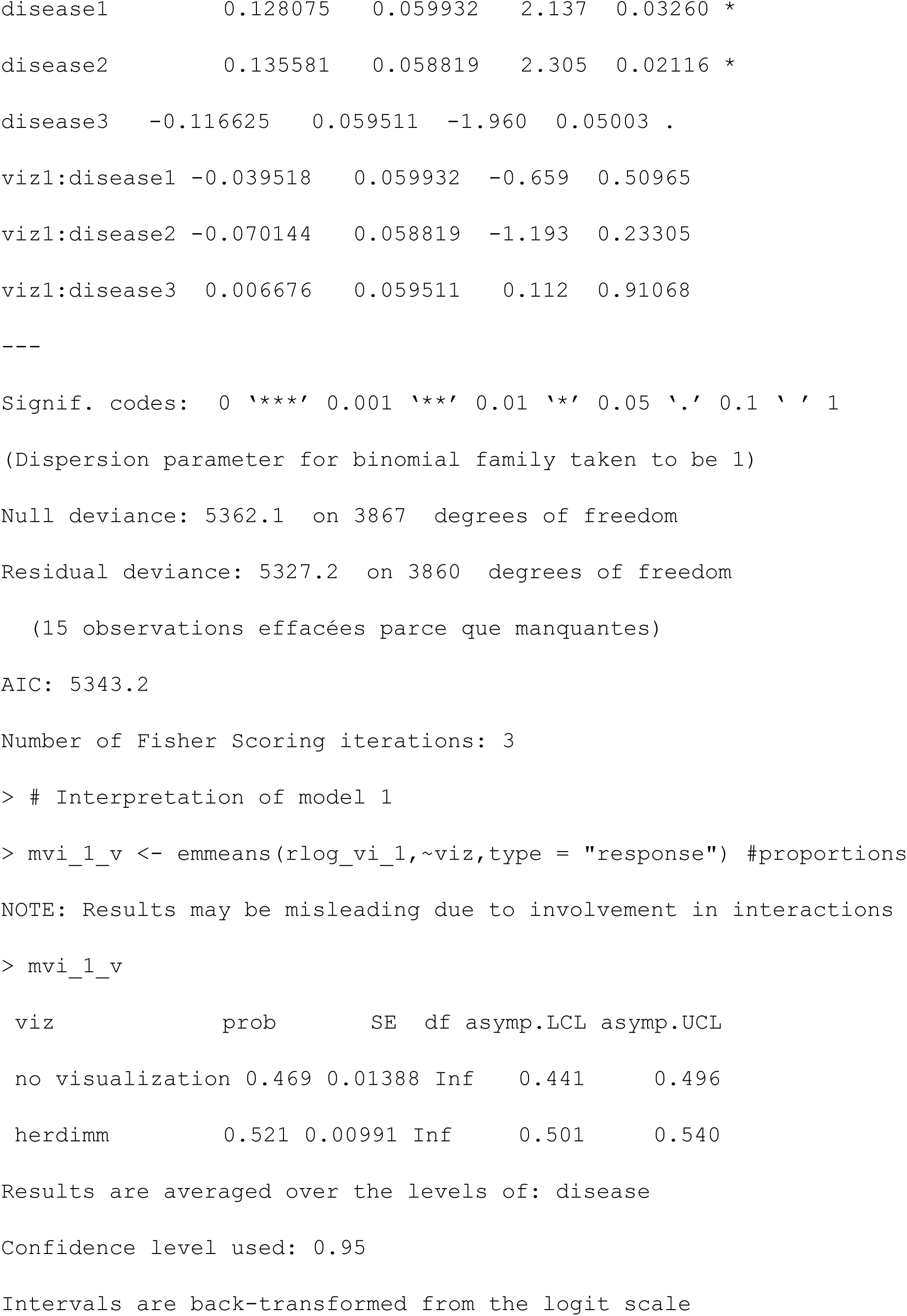
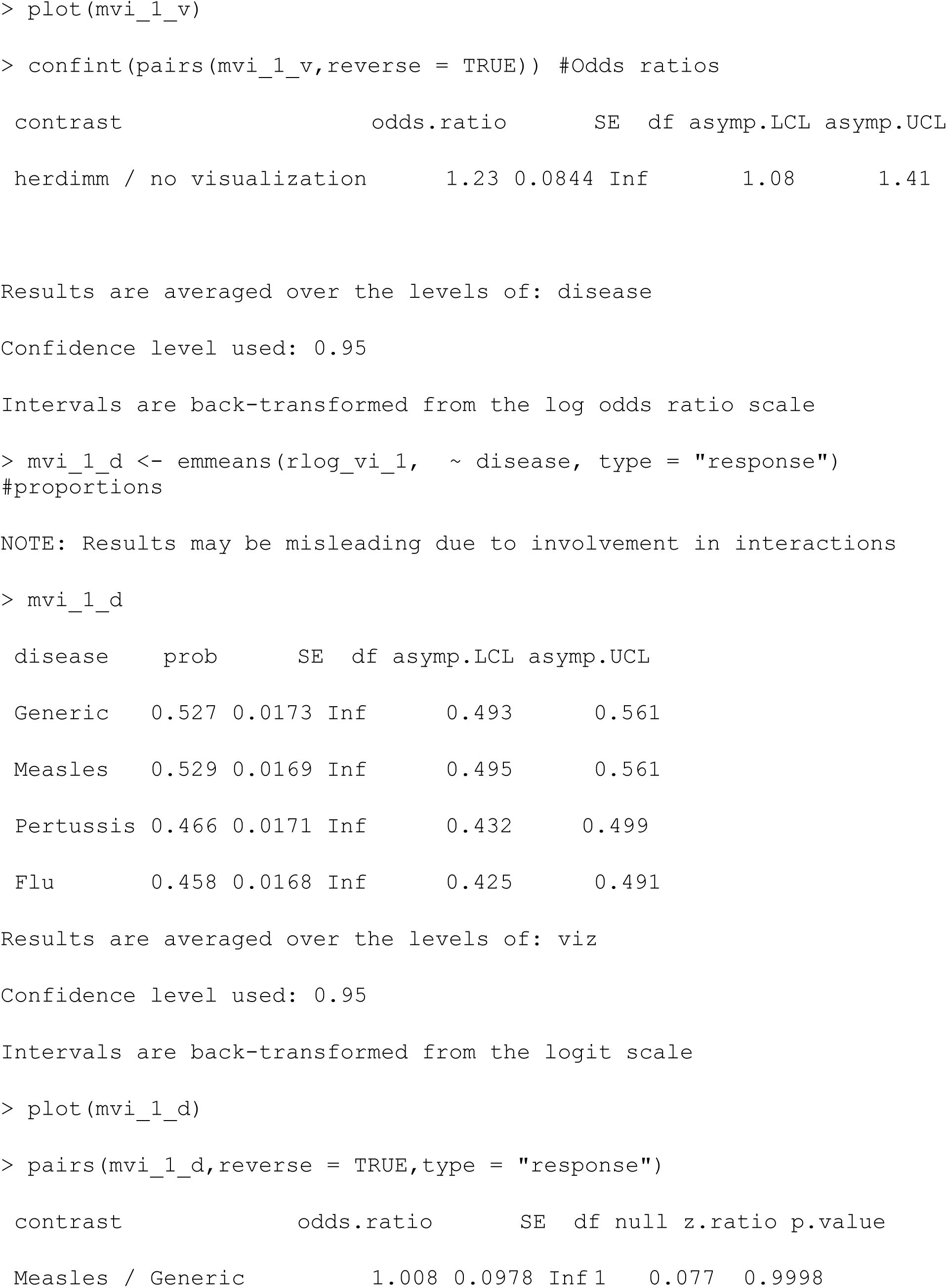
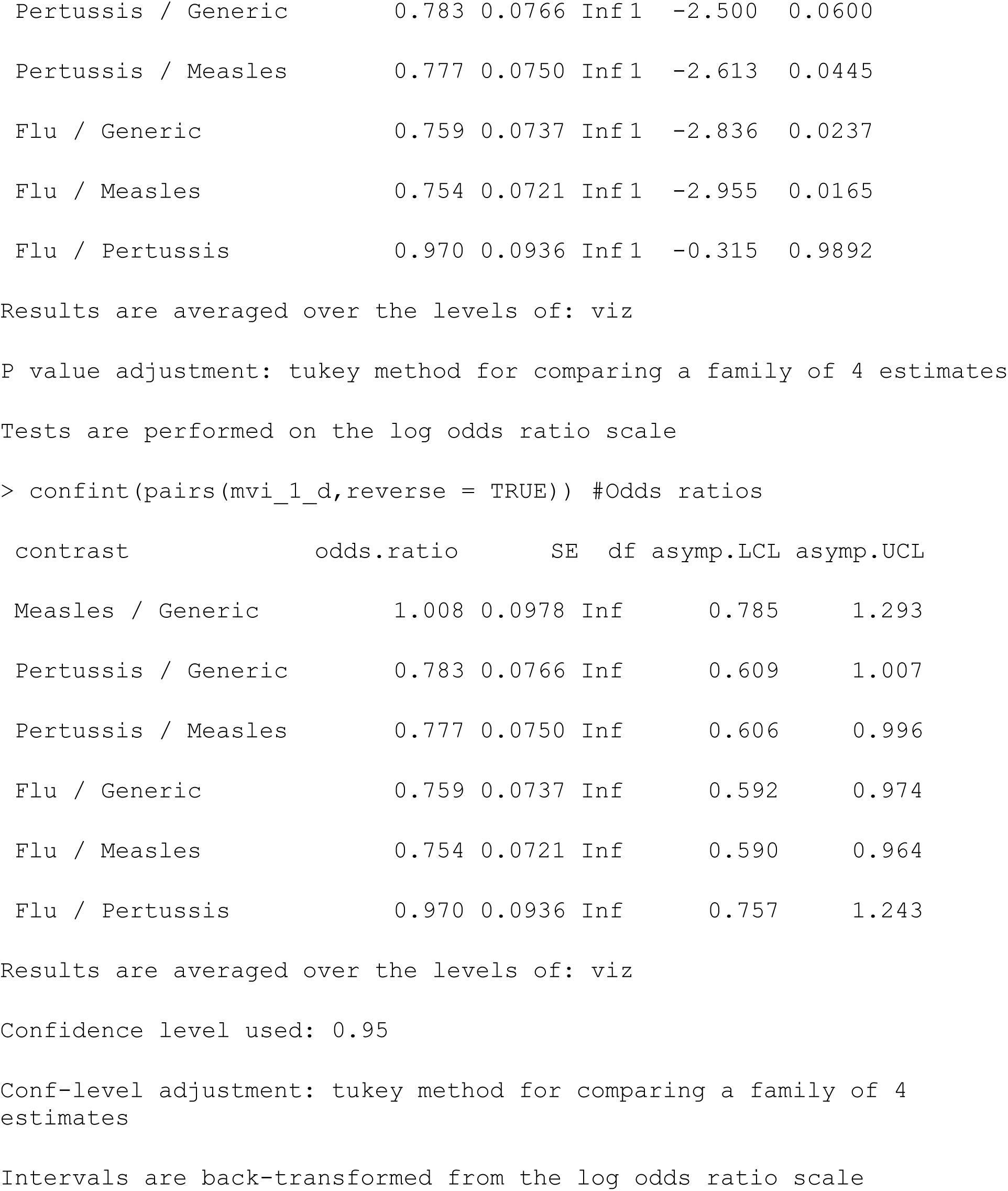

##### Model 2: Check for direct effects of factors with adjustment for other covariates

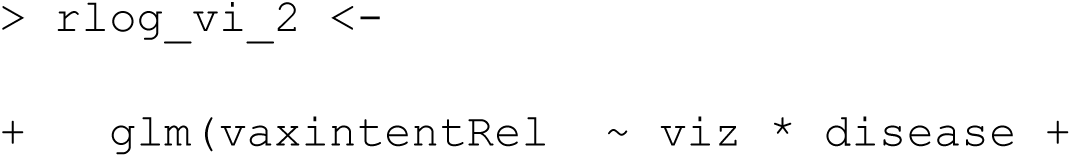
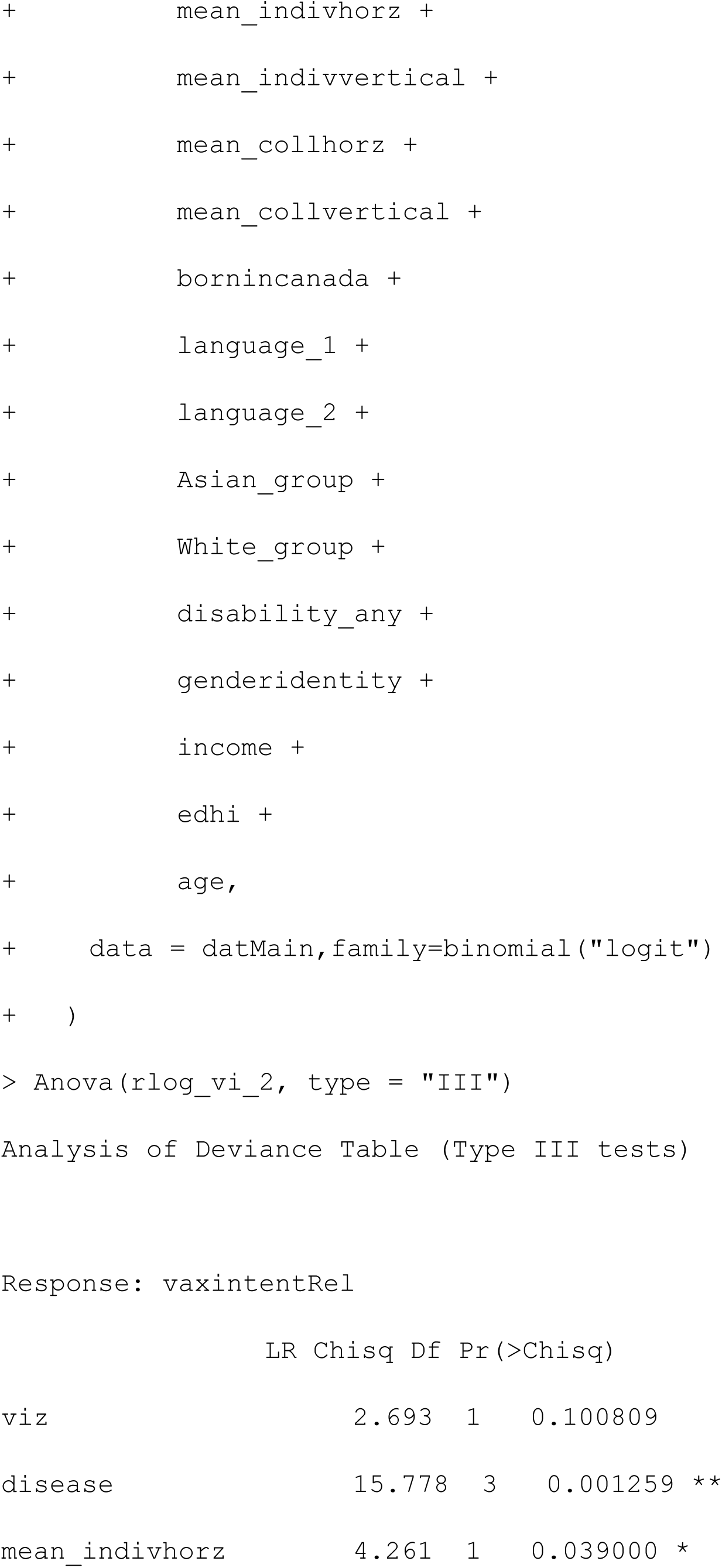
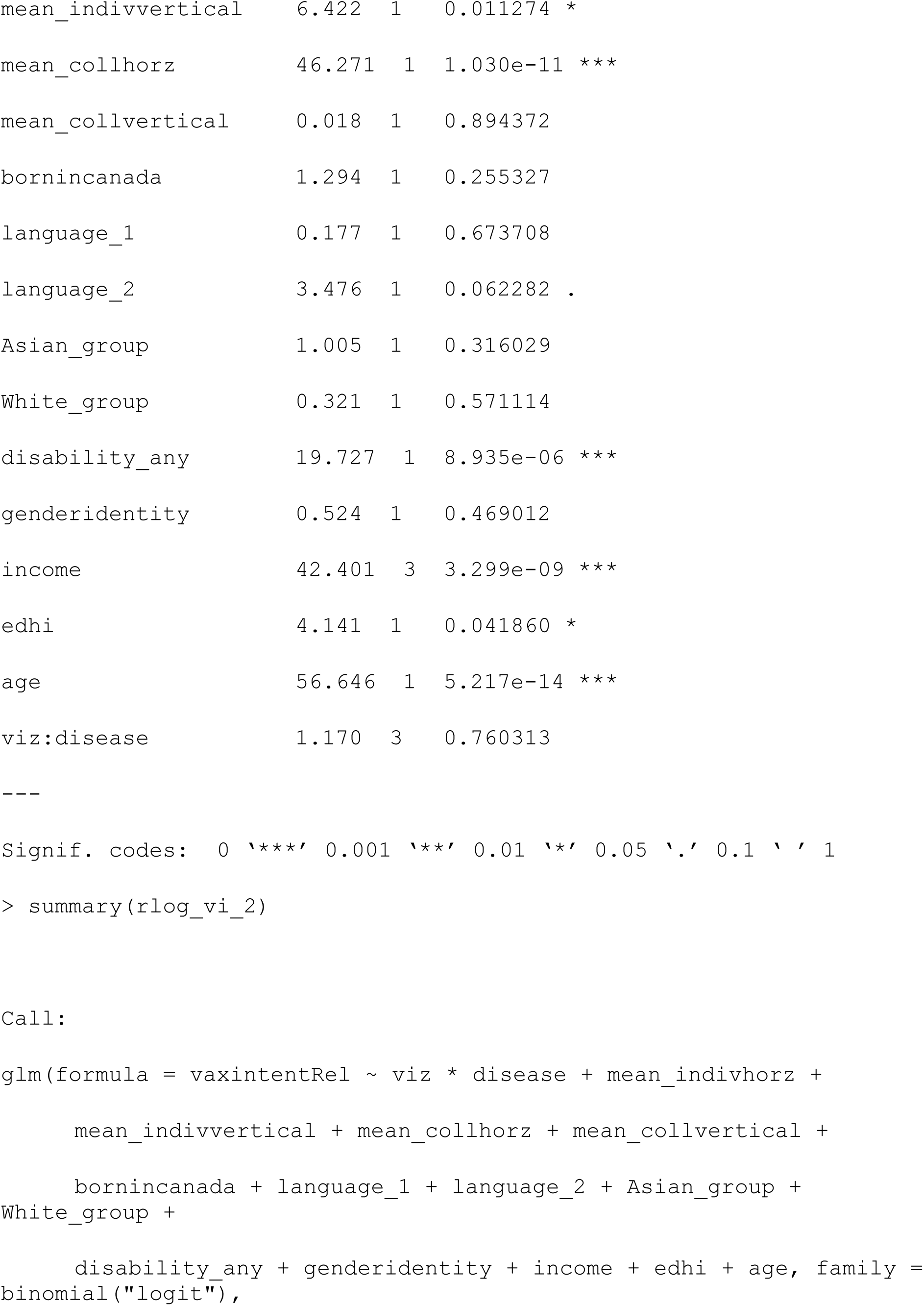
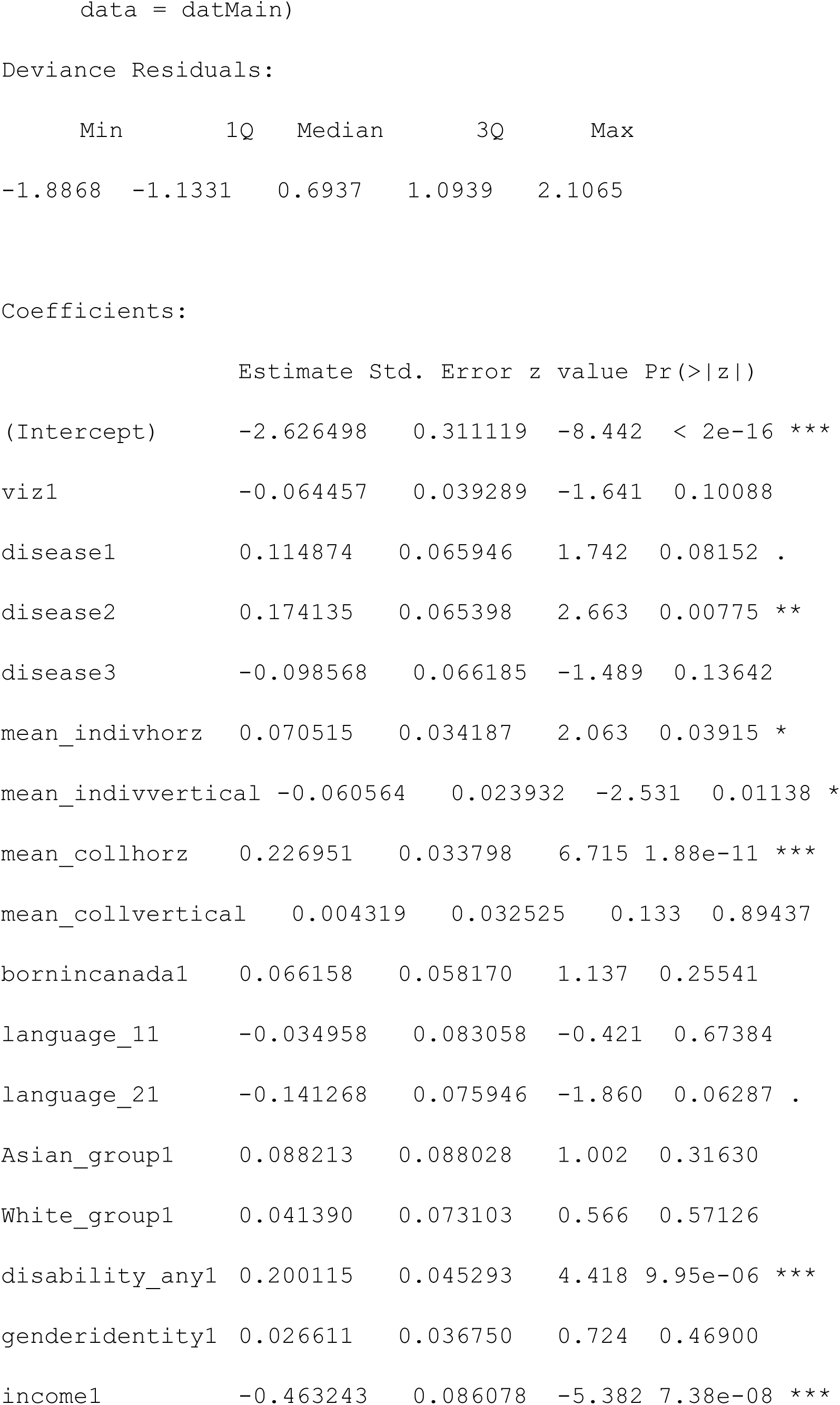
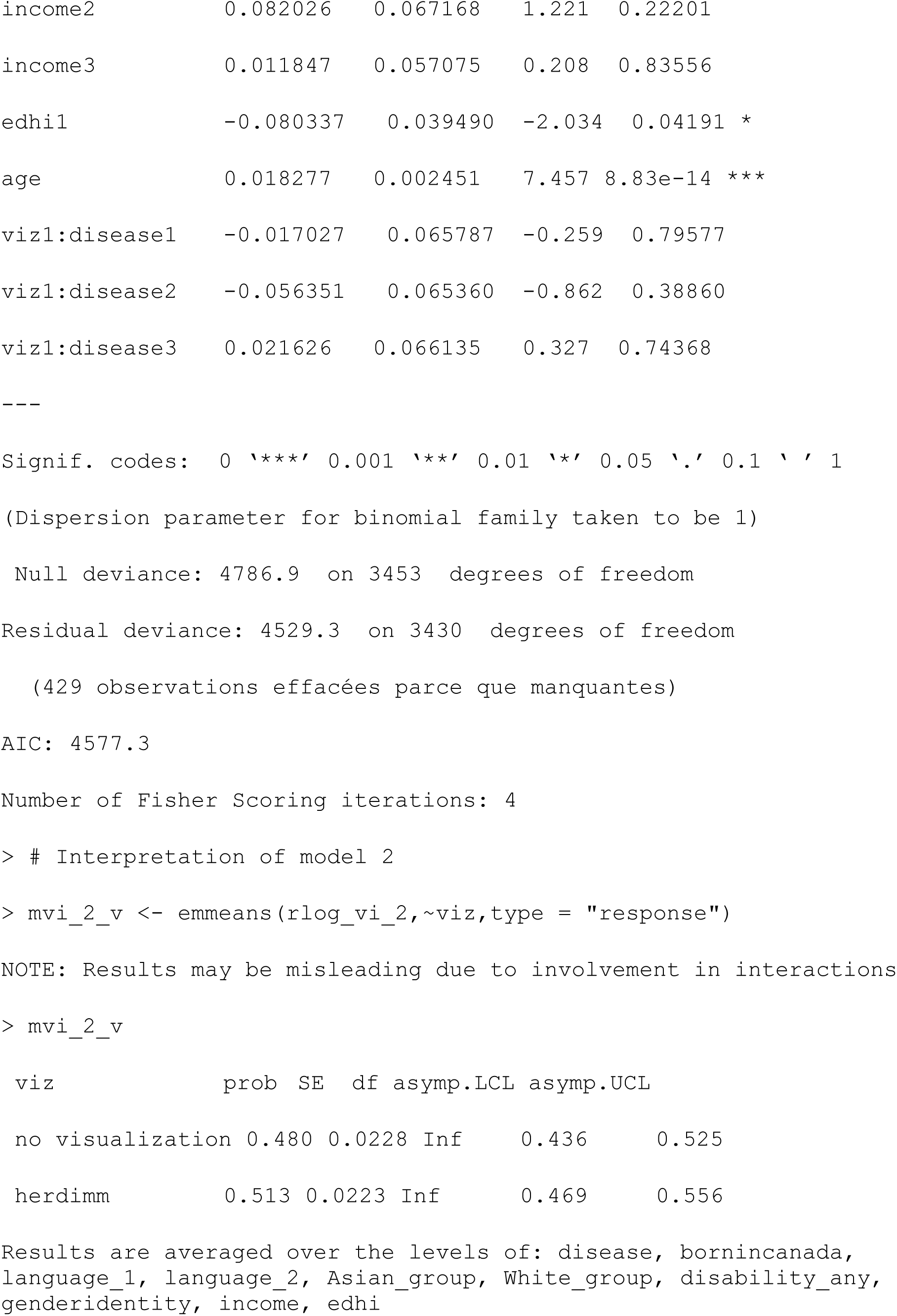
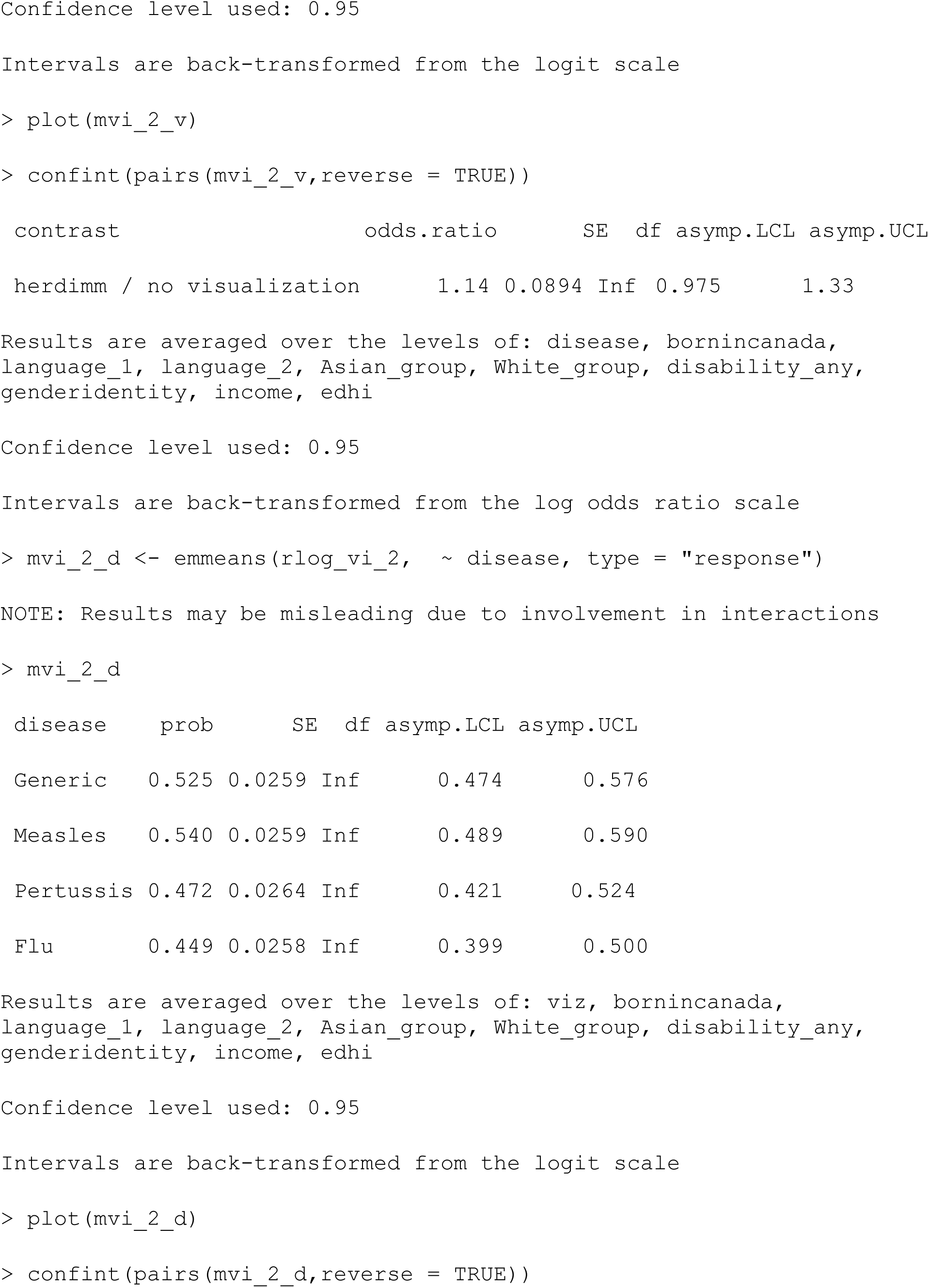
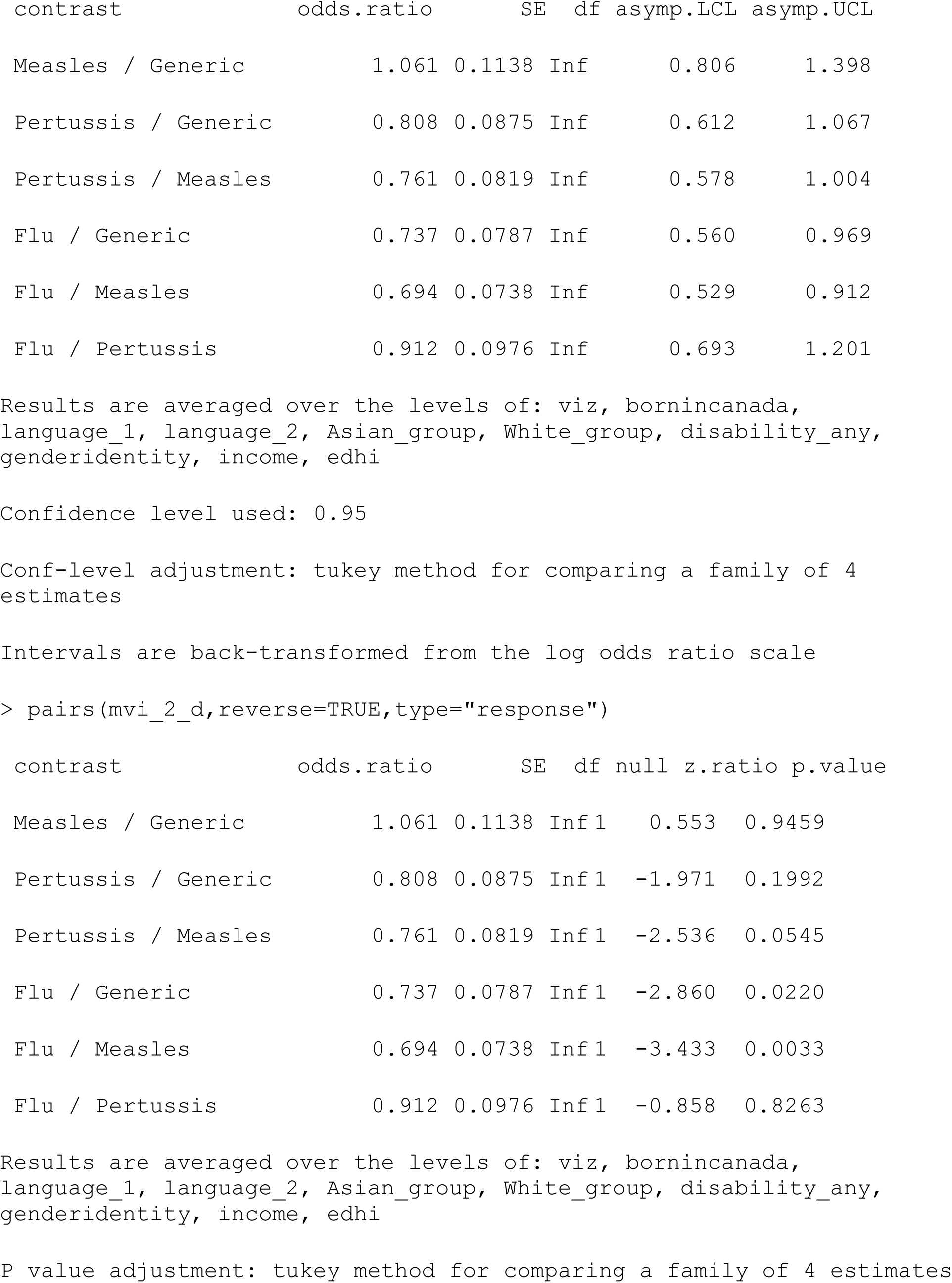
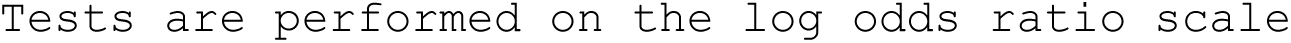

##### Model 3: Check for moderating effects of individualism & collectivism with adjustment for other covariates

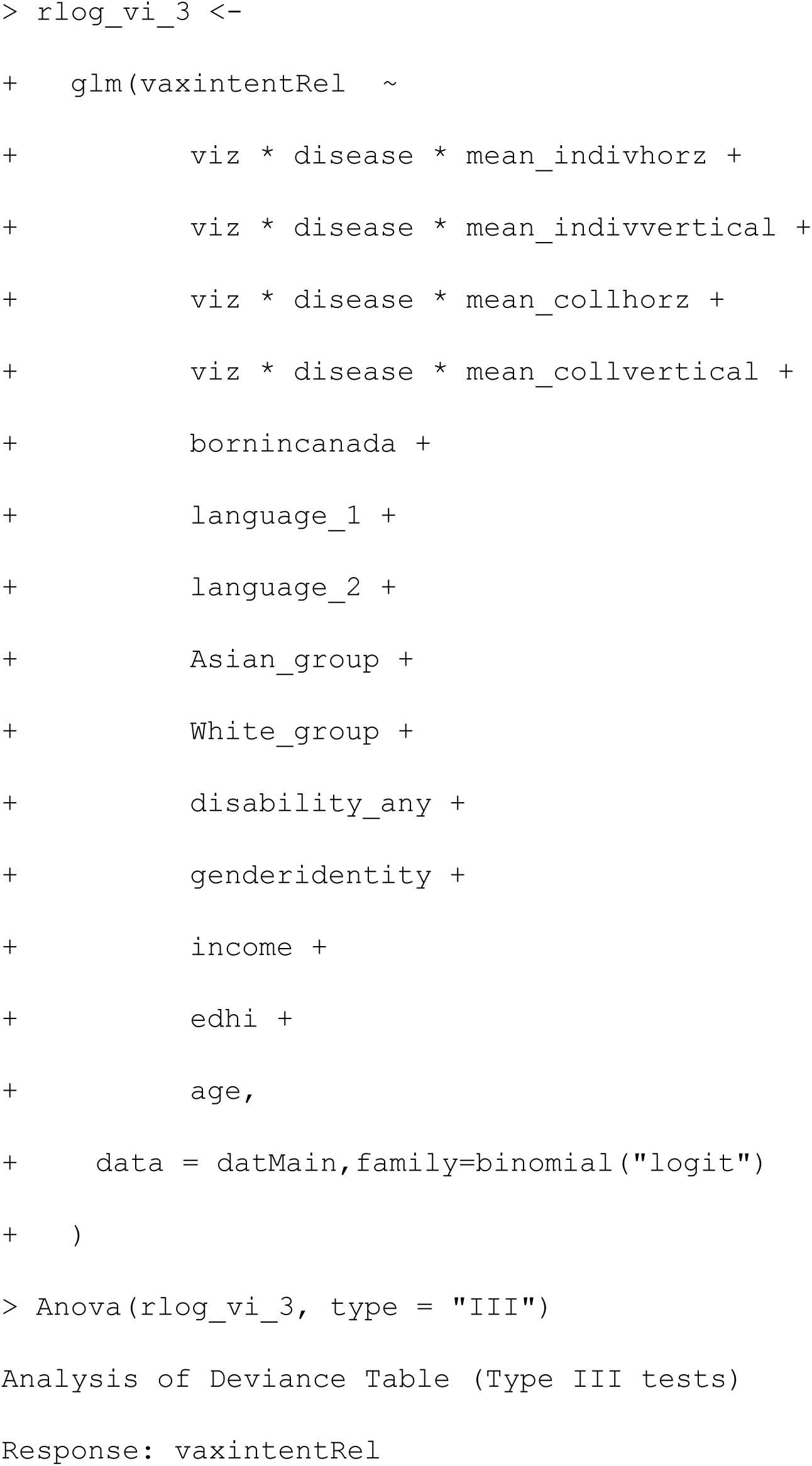
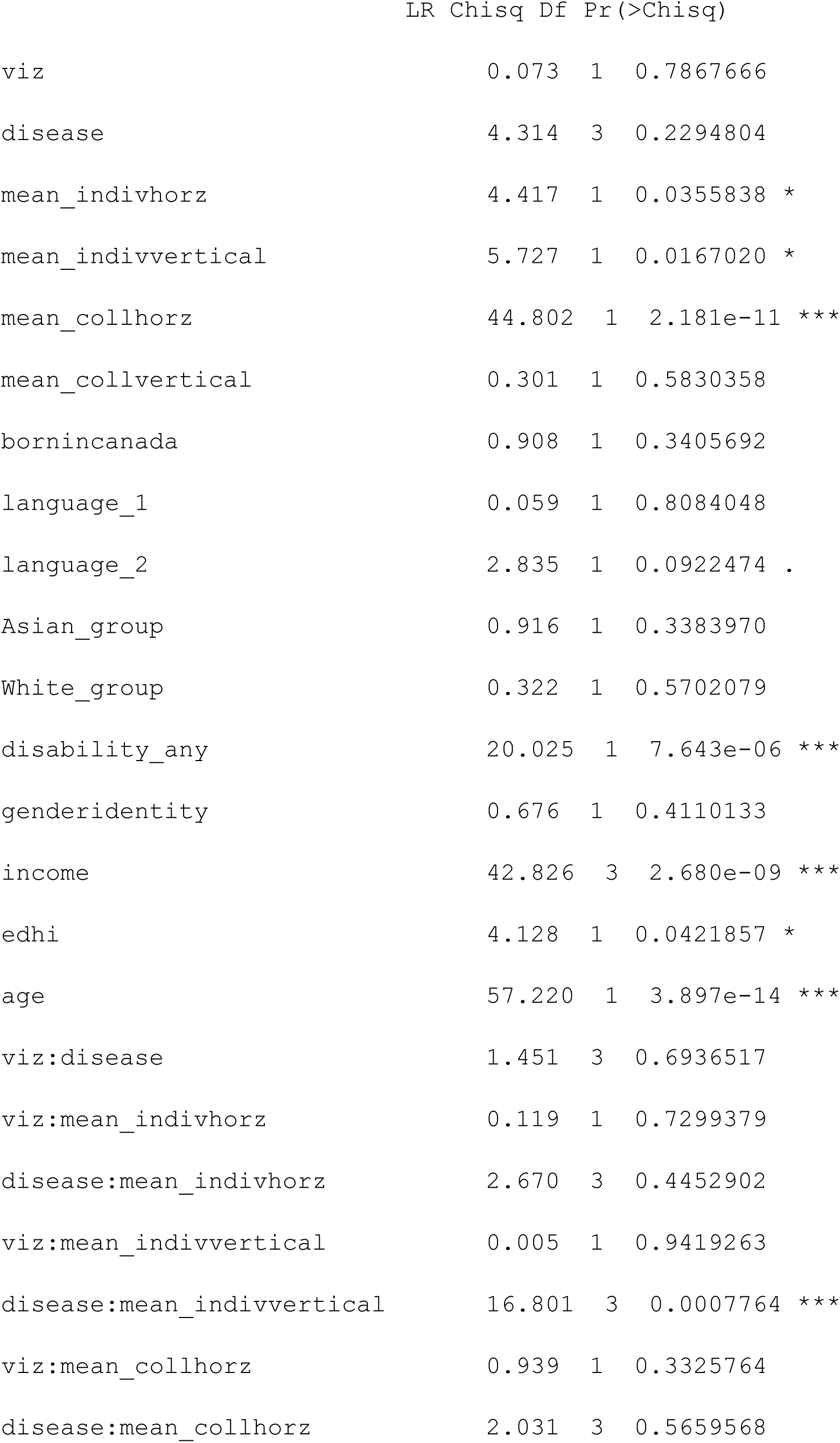
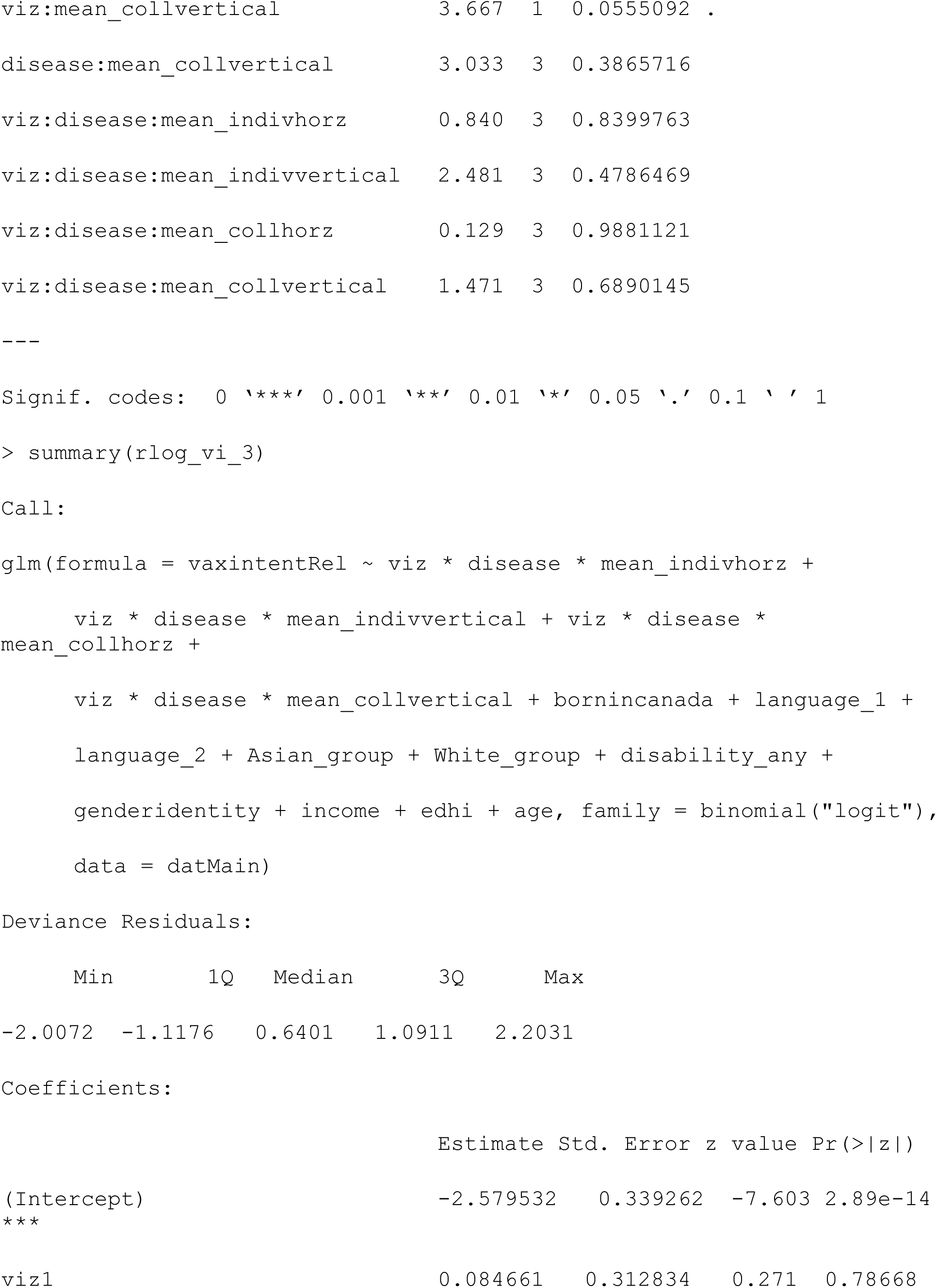
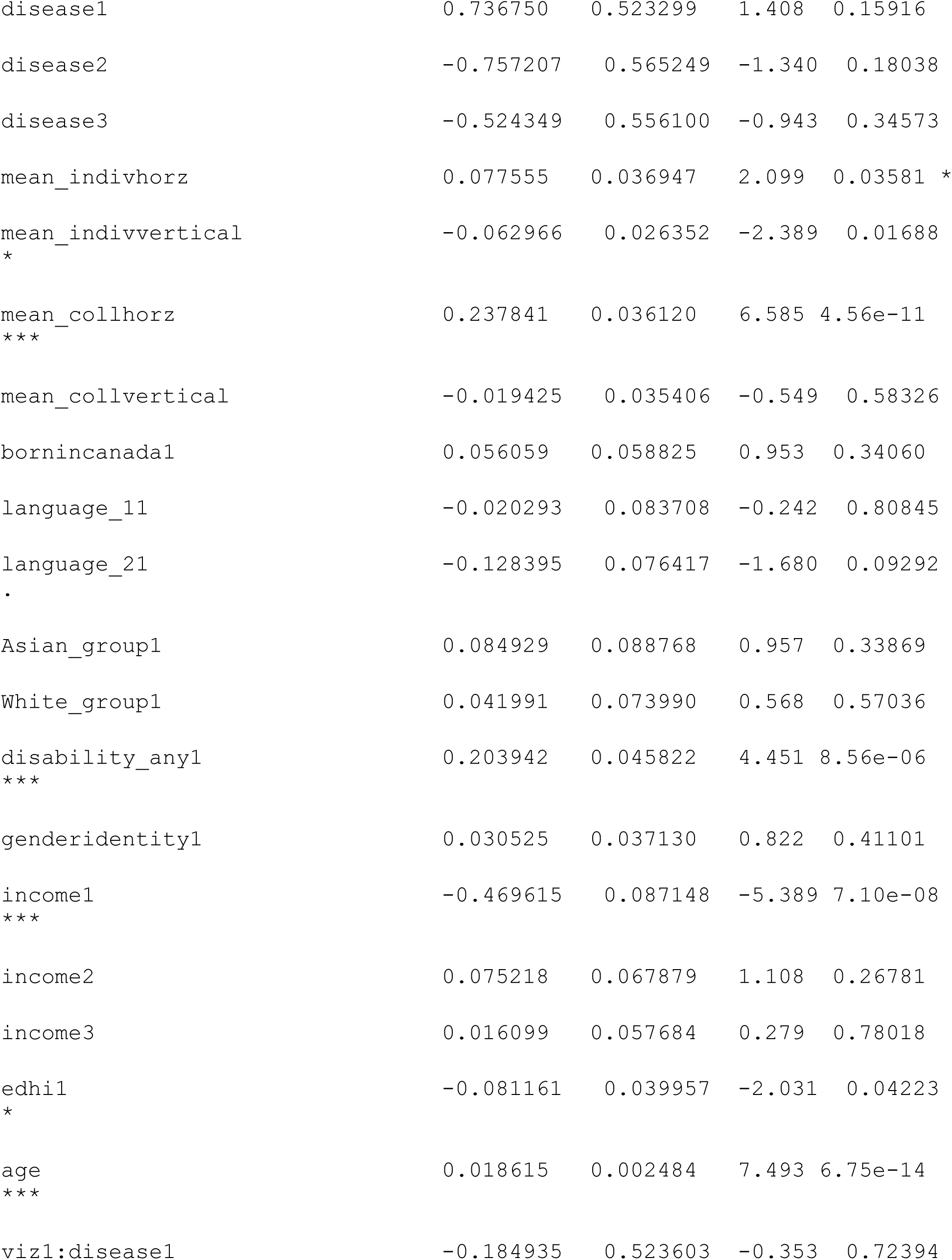
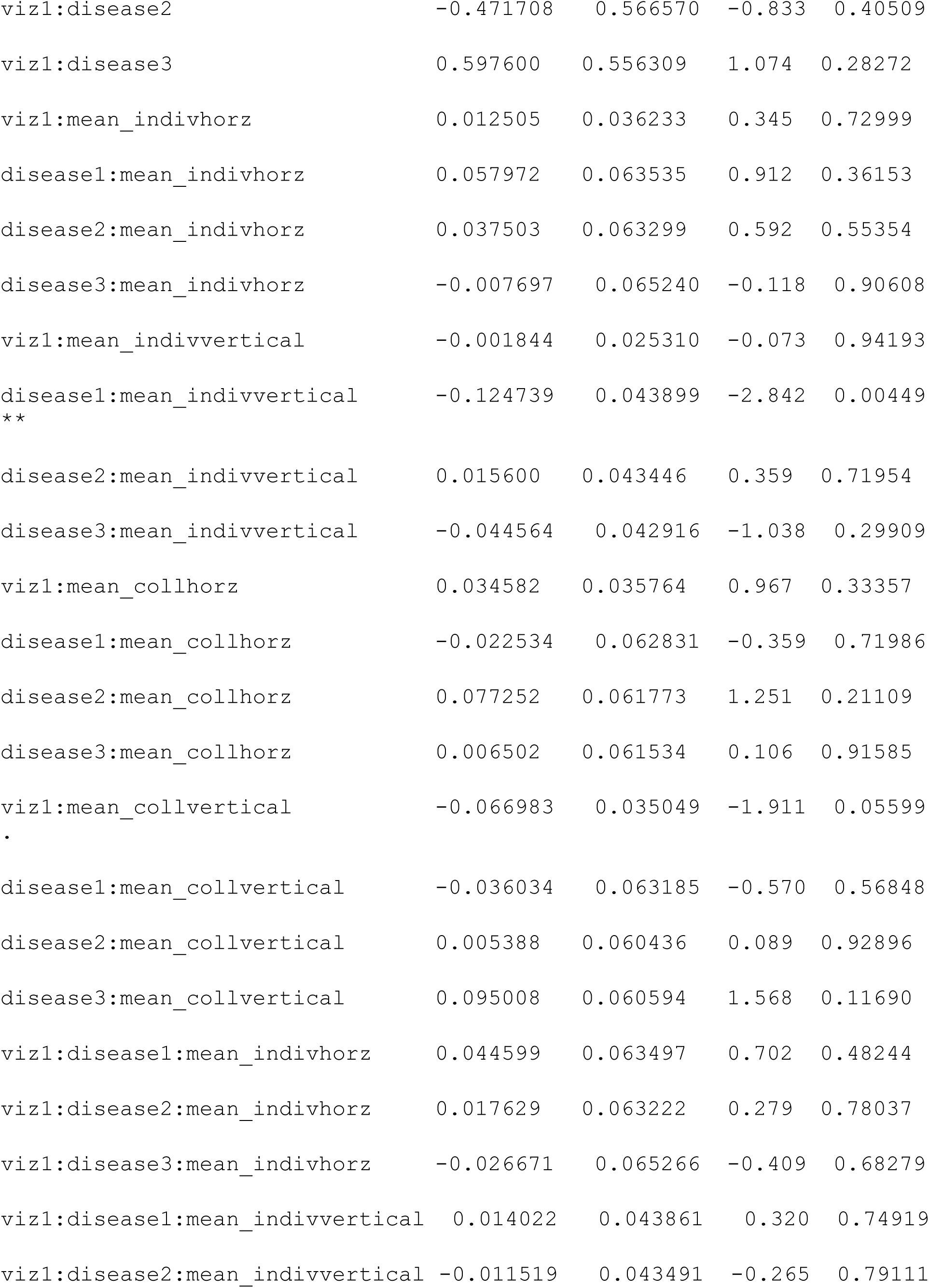
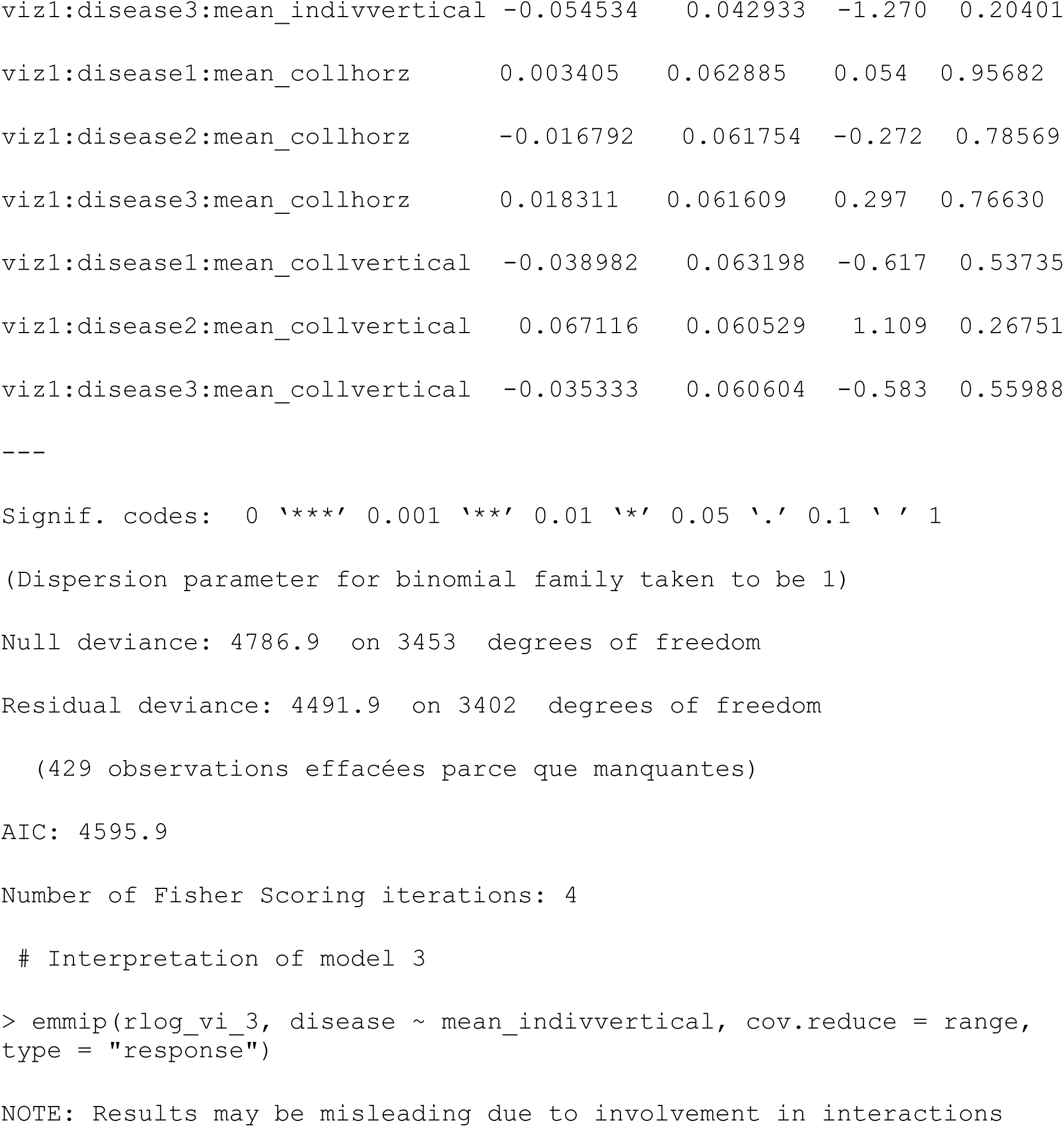

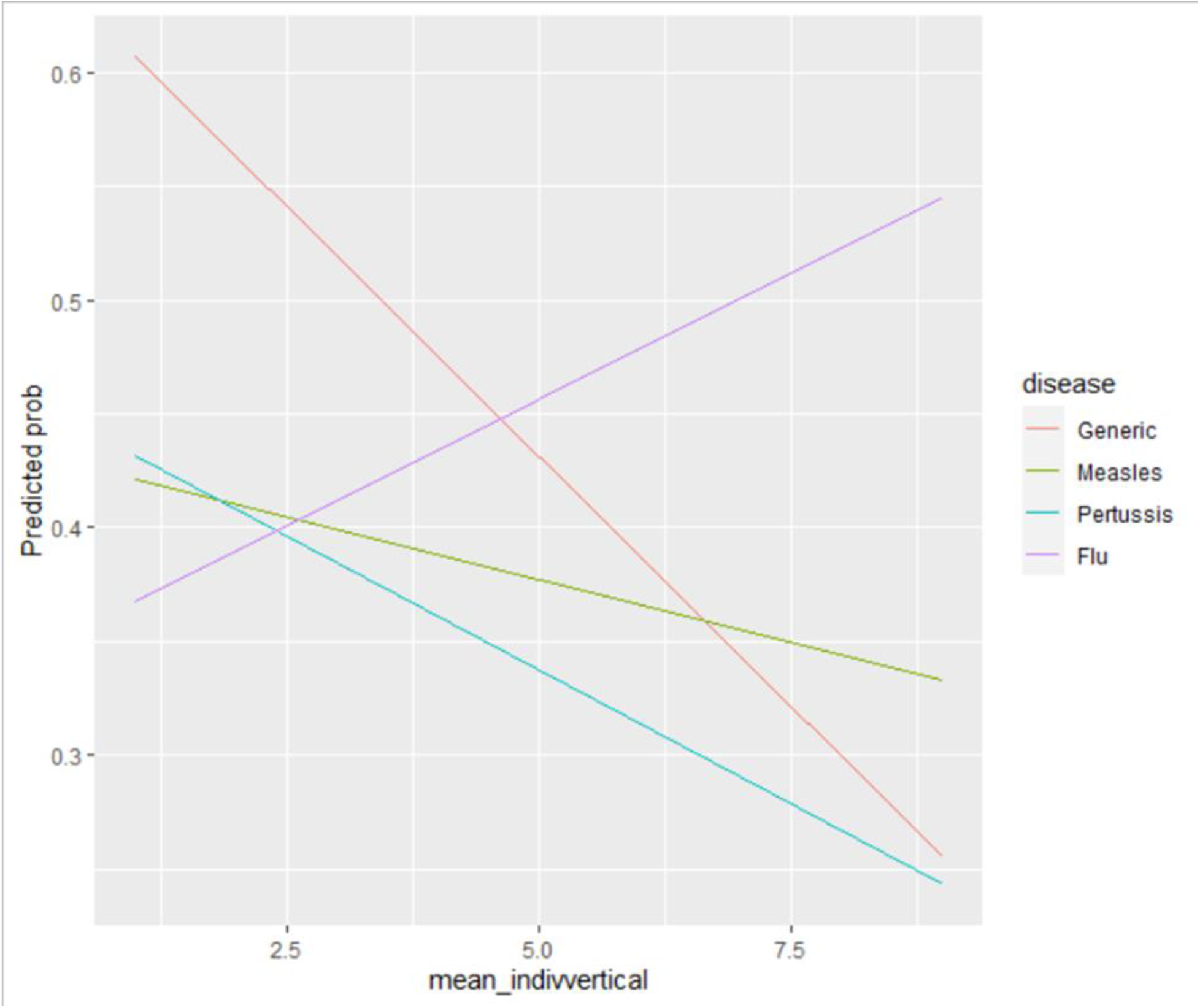

## Appendix 6: Outcomes versus Individualism and Collectivism subscales

### Objective risk perception (risk percep_1)

**S1:**
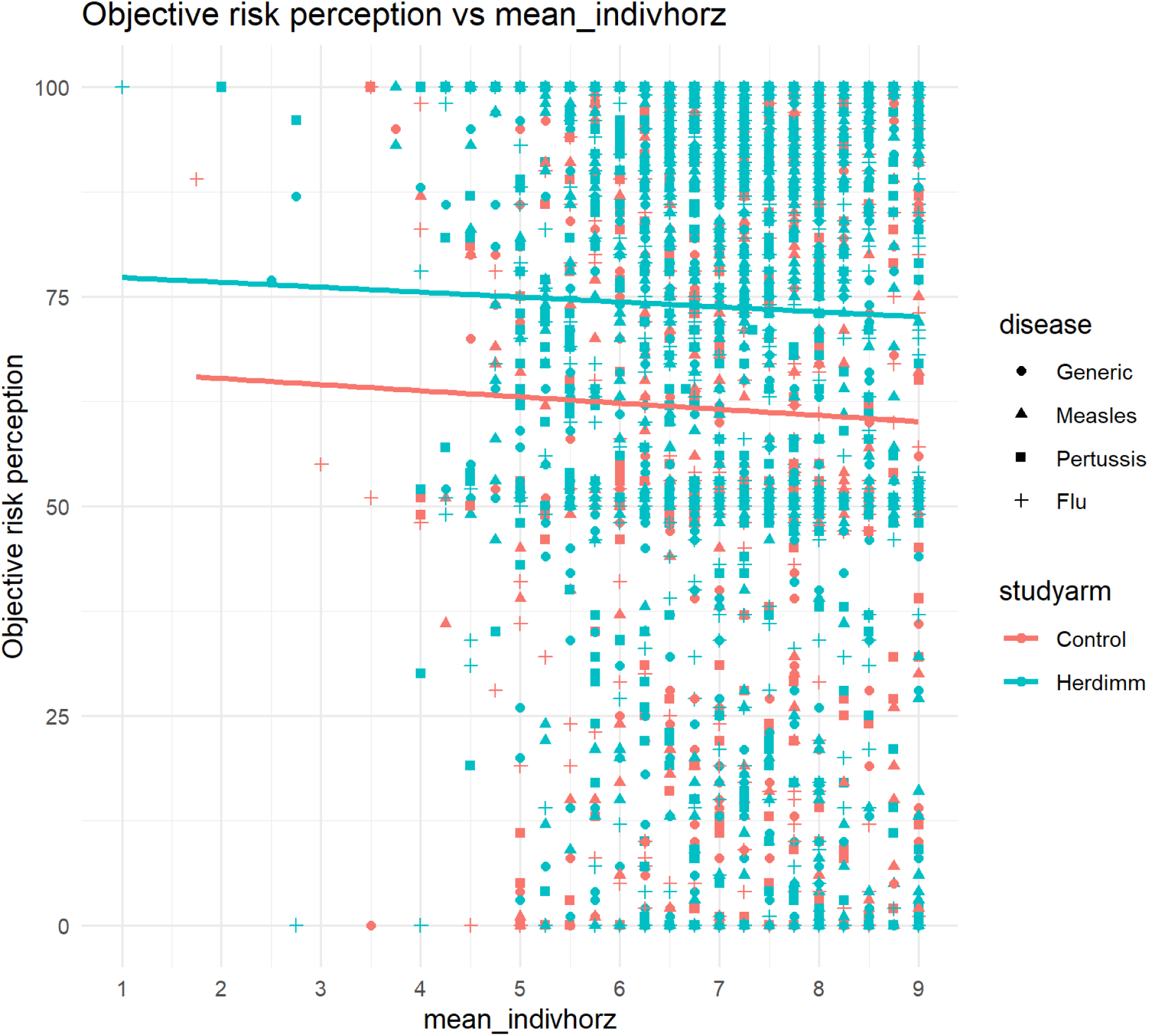
objective risk perception vs mean horizontal individualism.

**S2:**
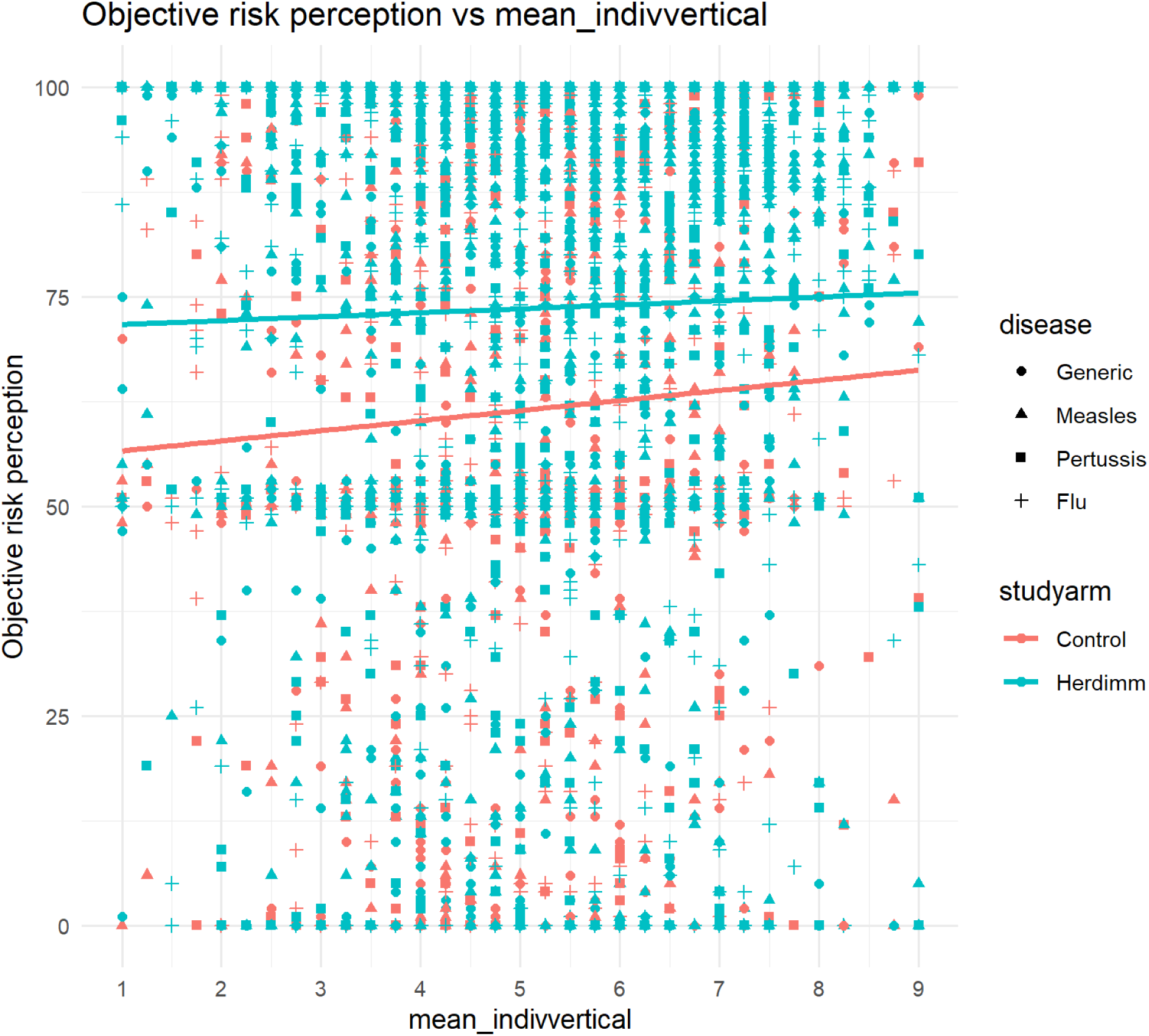
objective risk perception vs mean vertical individualism.

**S3:**
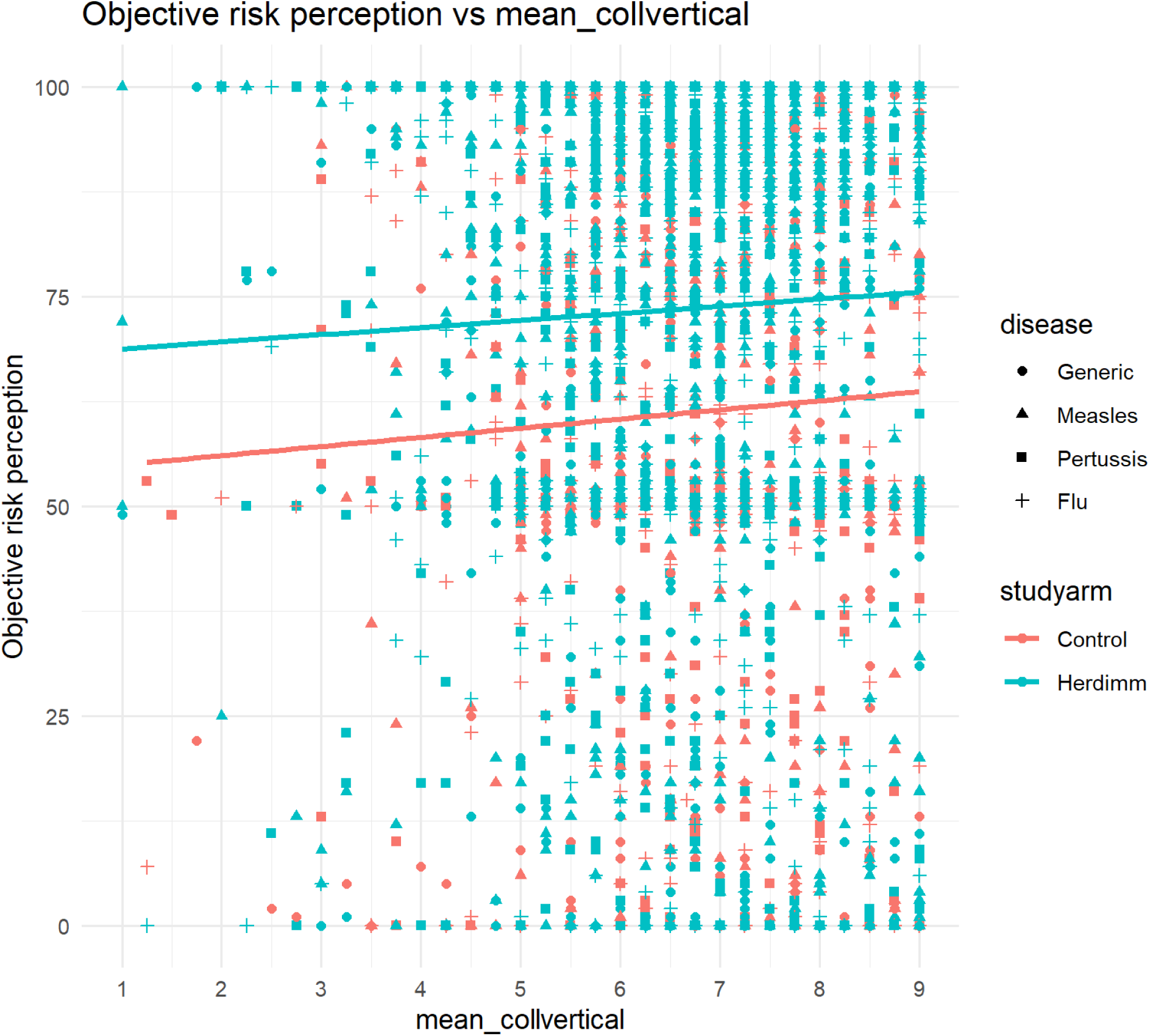
objective risk perception vs mean vertical collectivism.

**S4:**
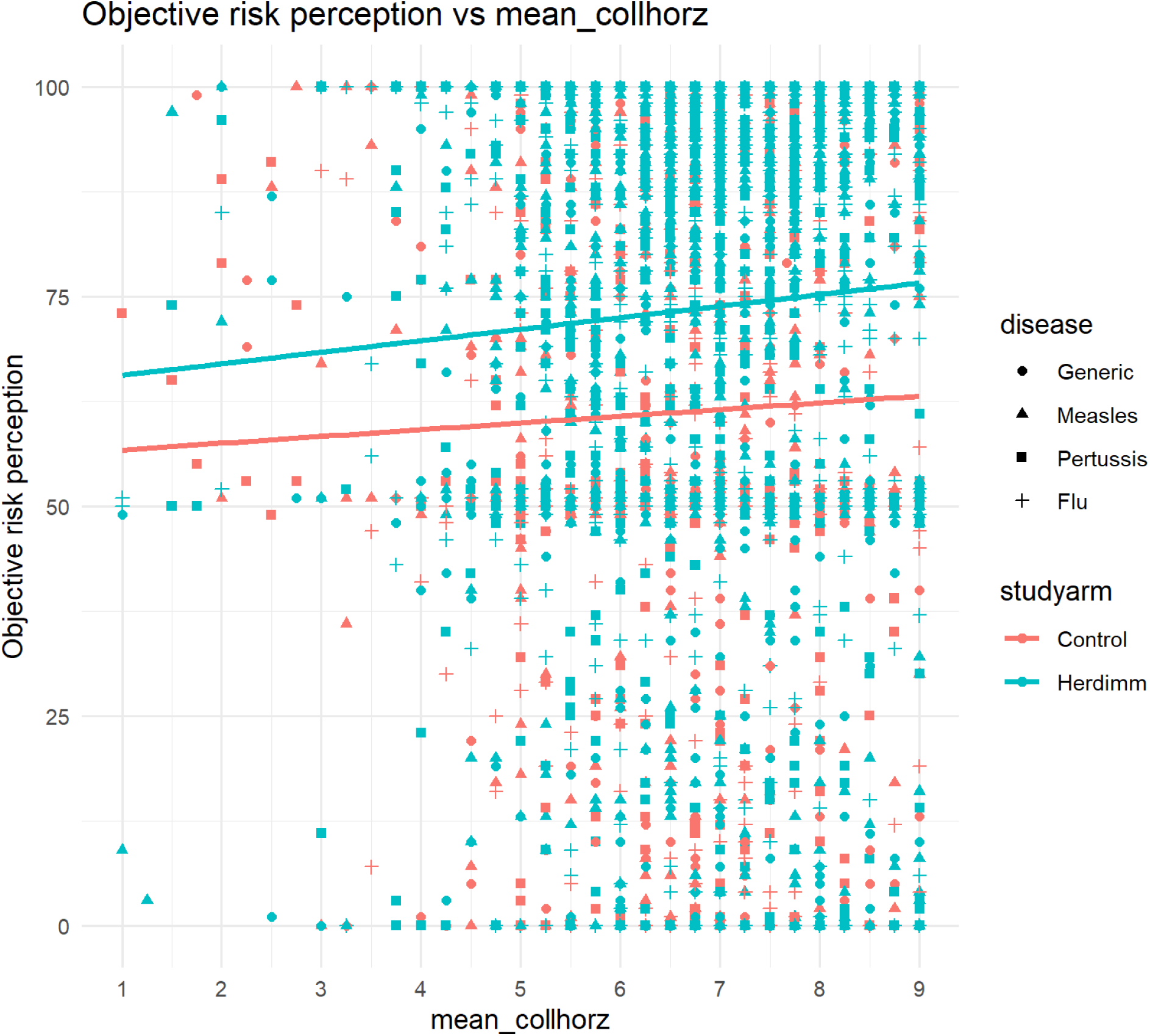
objective risk perception vs mean horizontal collectivism.

### Subjective risk perception (risk percept_2 to 6)

**S5:**
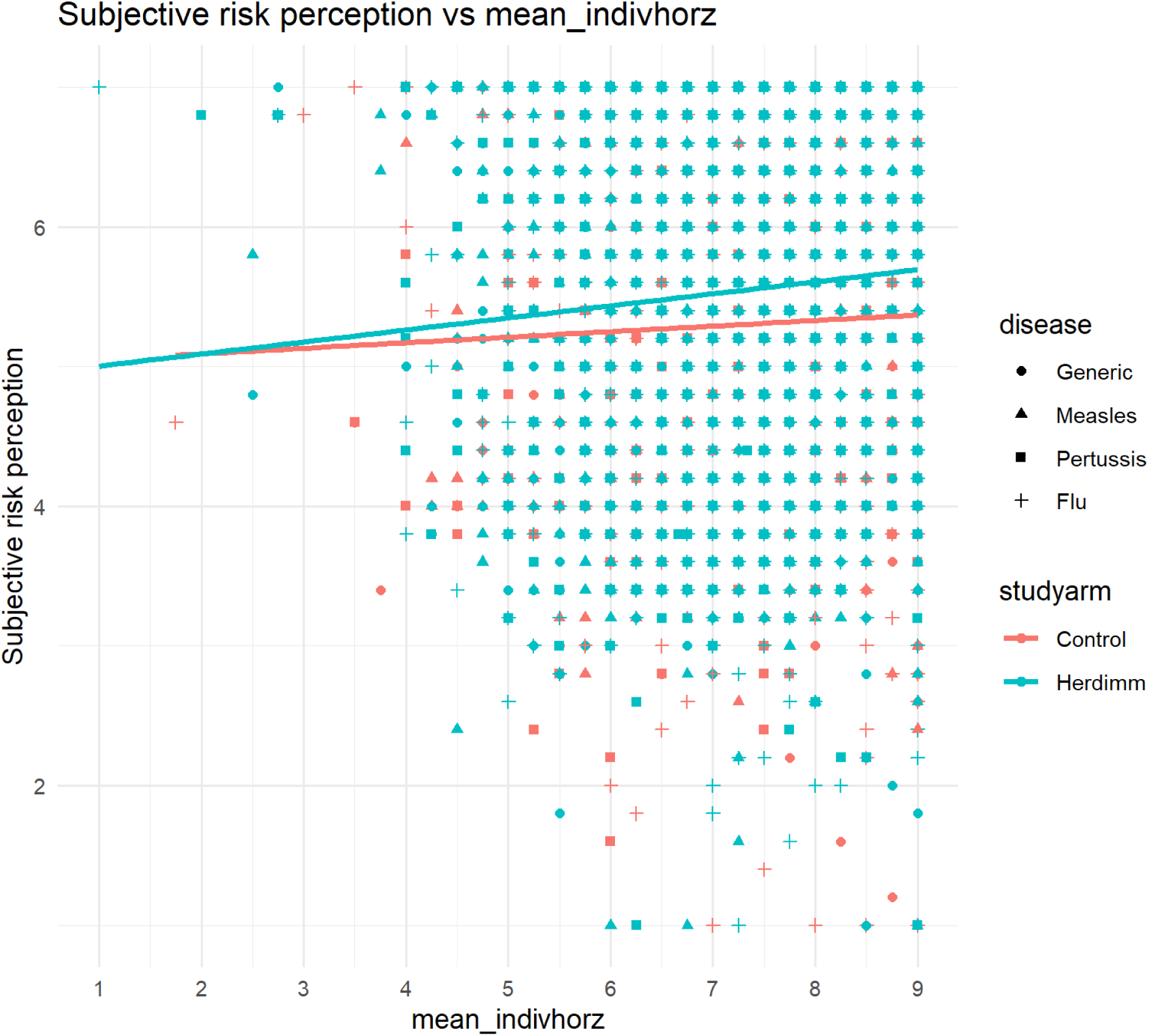
subjective risk perception vs mean horizontal individualism.

**S6:**
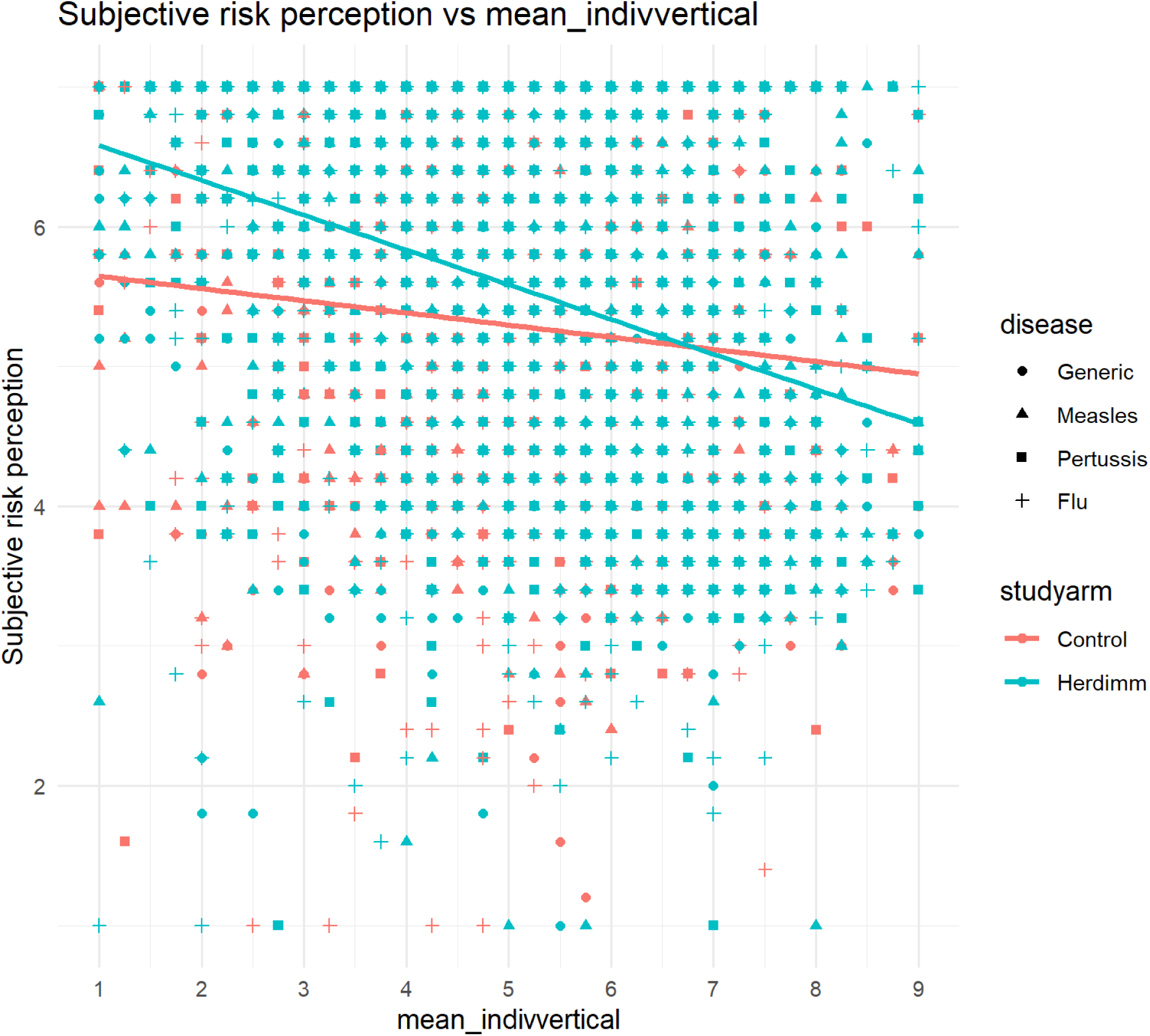
subjective risk perception vs mean vertical individualism.

**S7:**
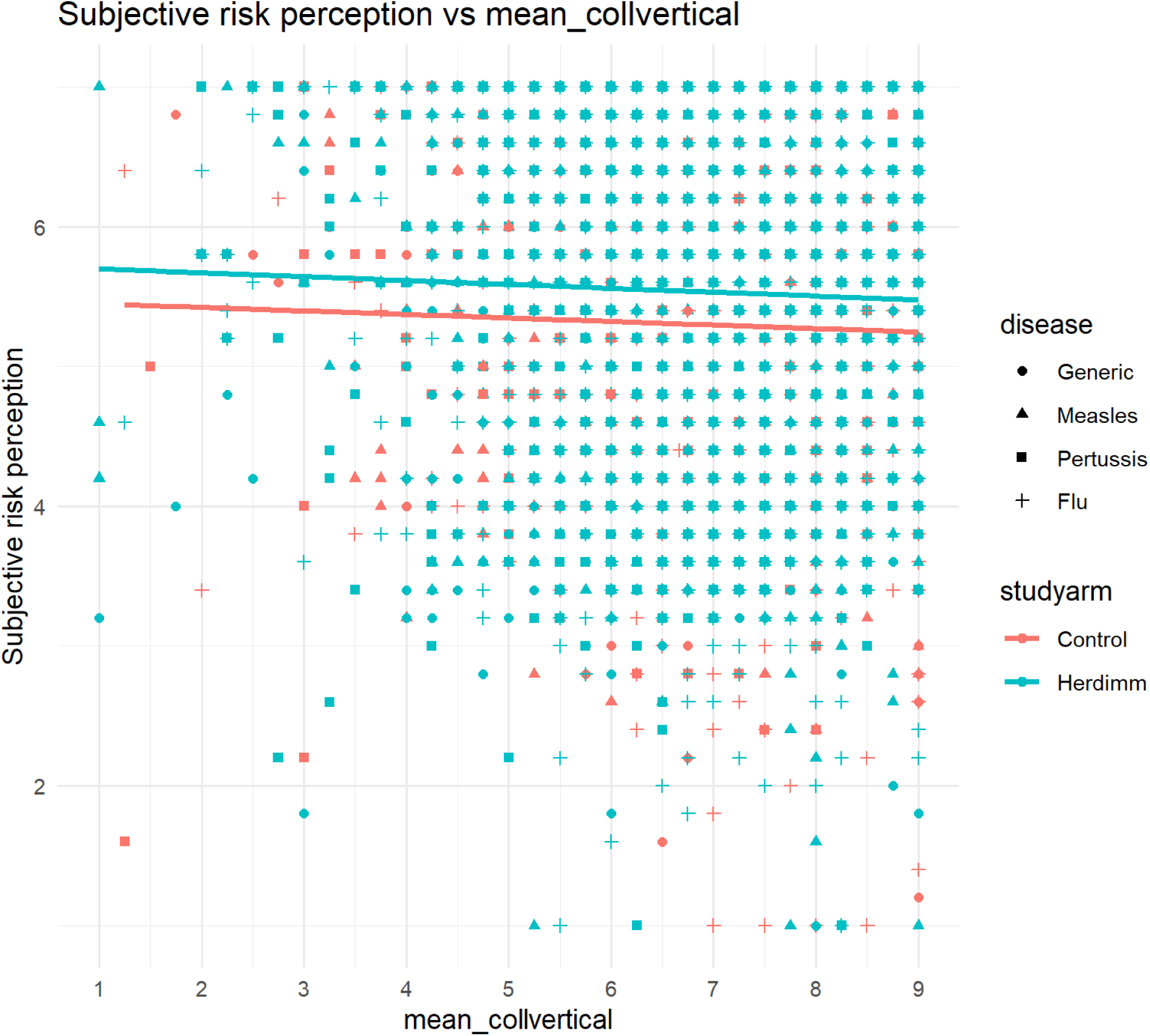
subjective risk perception vs mean vertical collectivism.

**S8:**
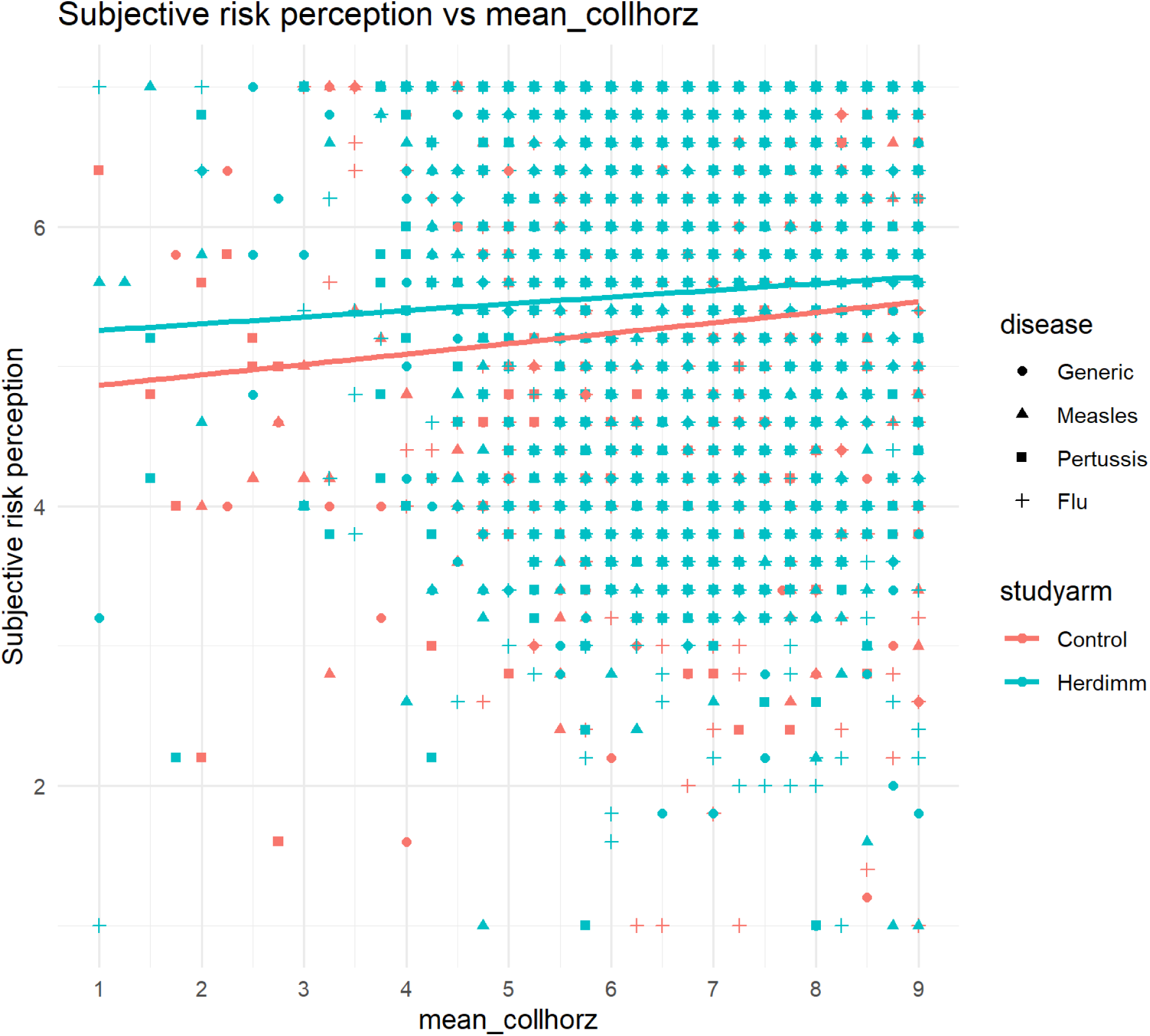
subjective risk perception vs mean horizontal collectivism.

### Emotions

**S9:**
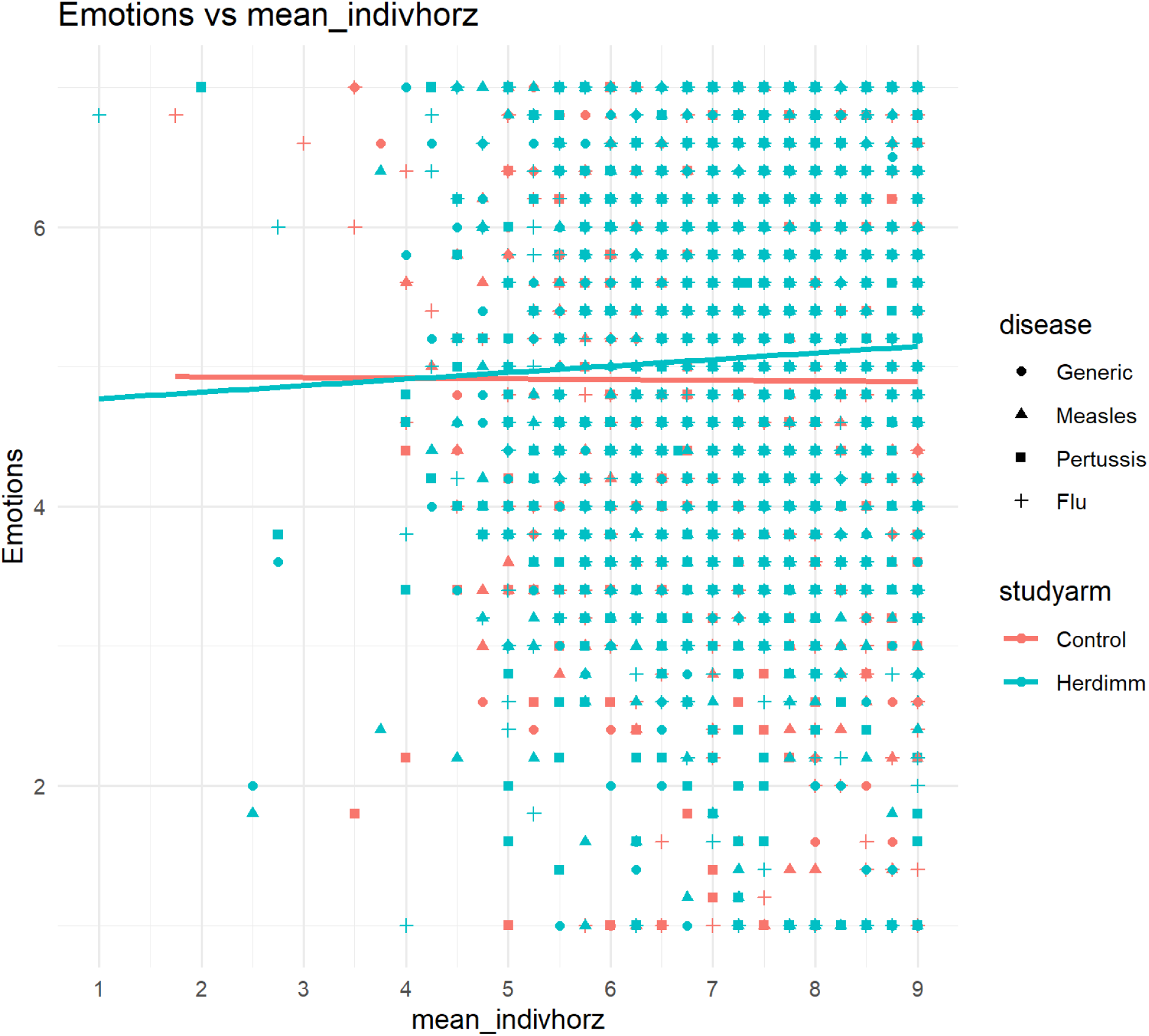
emotions vs mean horizontal individualism.

**S10:**
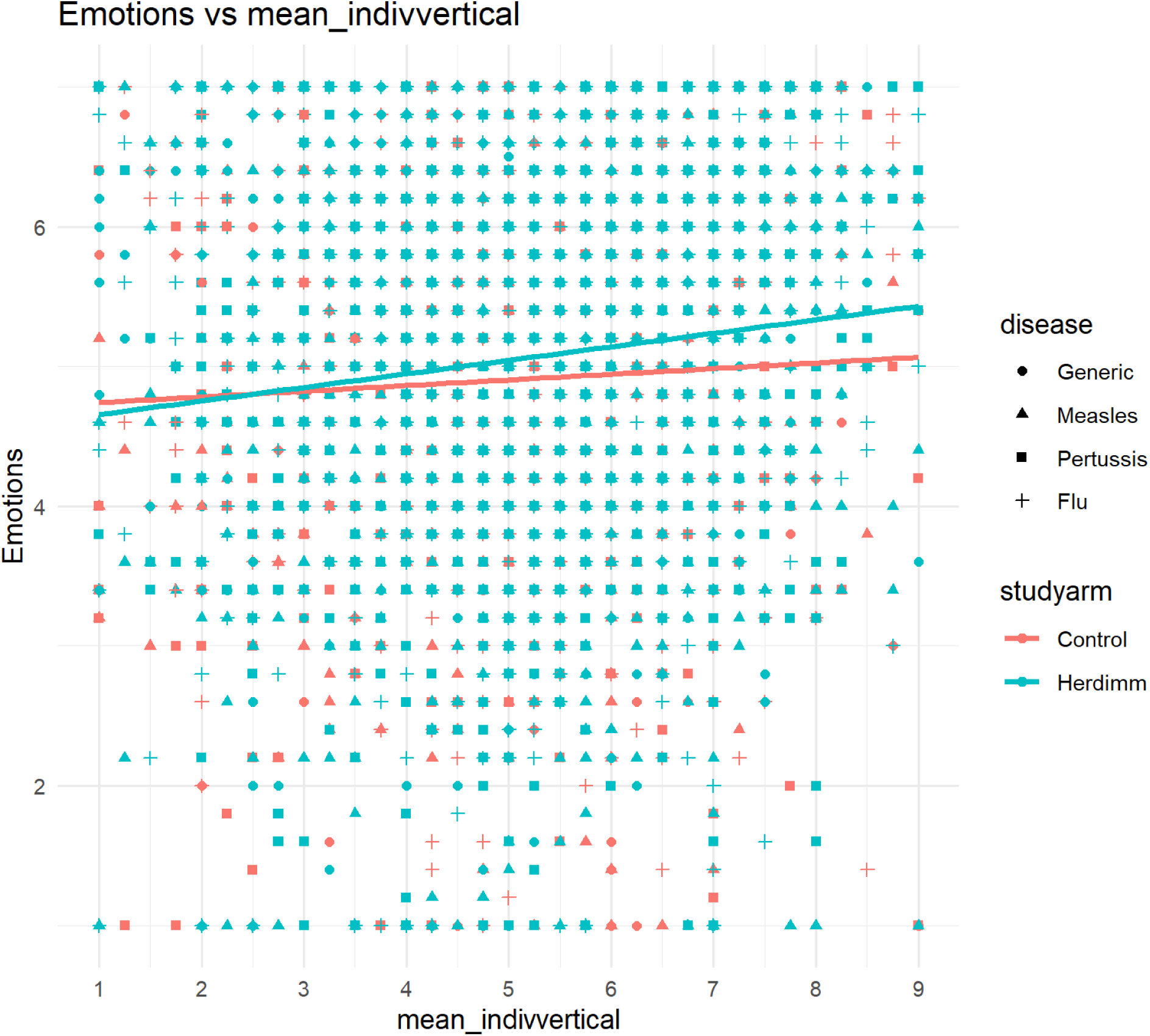
emotions vs mean vertical individualism.

**S11:**
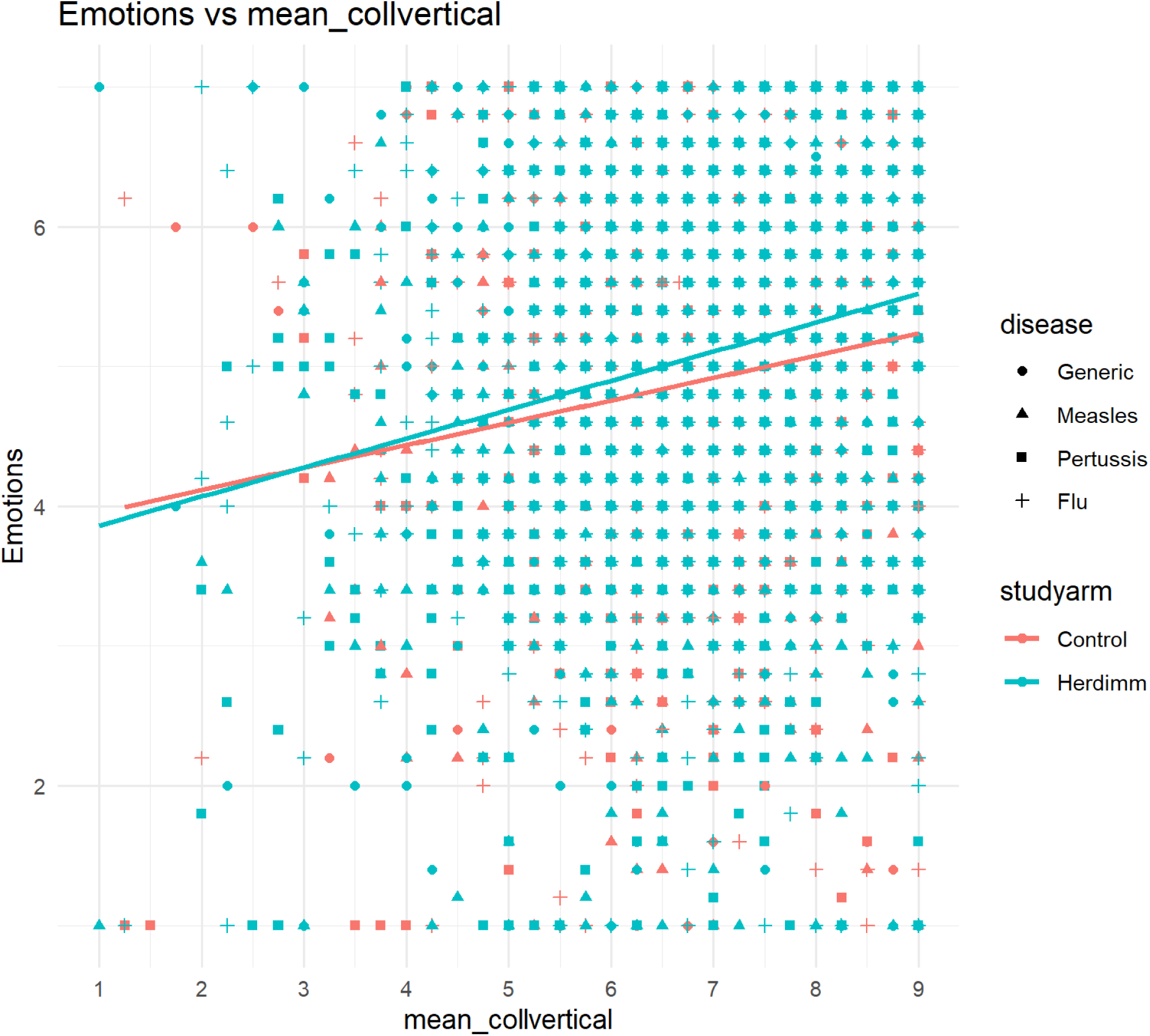
emotions vs mean vertical collectivism.

**S12:**
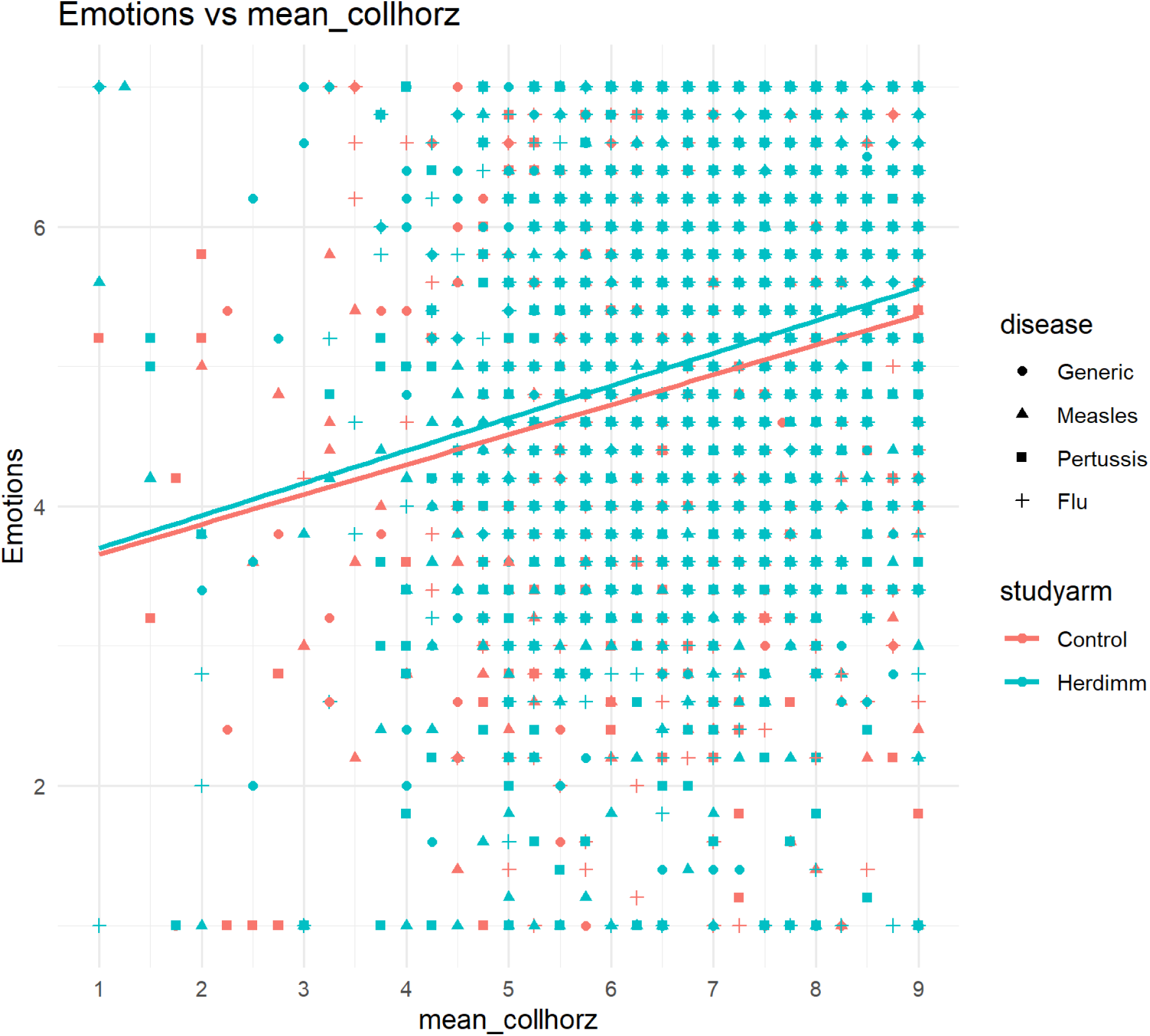
emotions vs mean horizontal collectivism.

### Knowledge

**S13:**
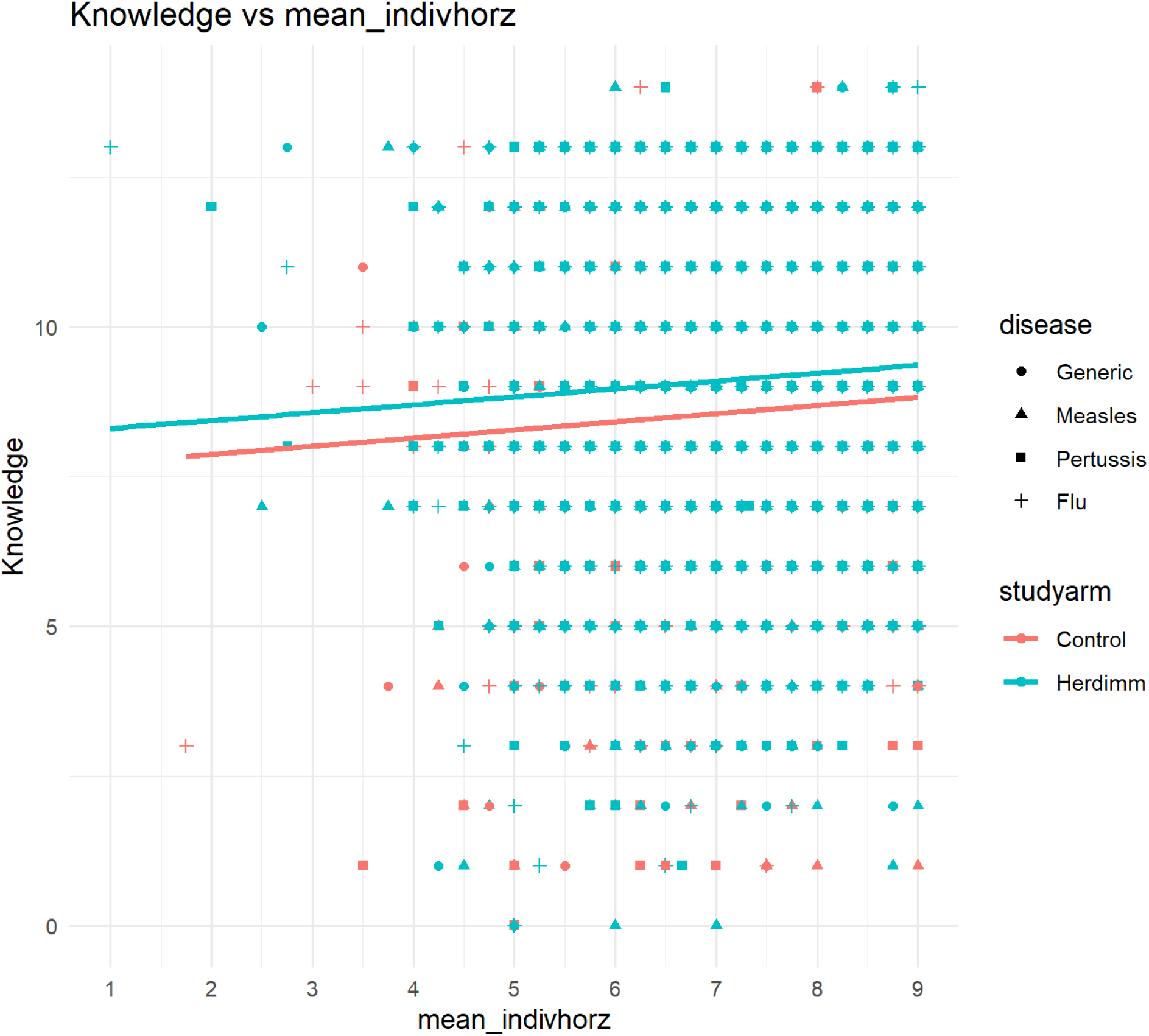
knowledge vs mean horizontal individualism.

**S14:**
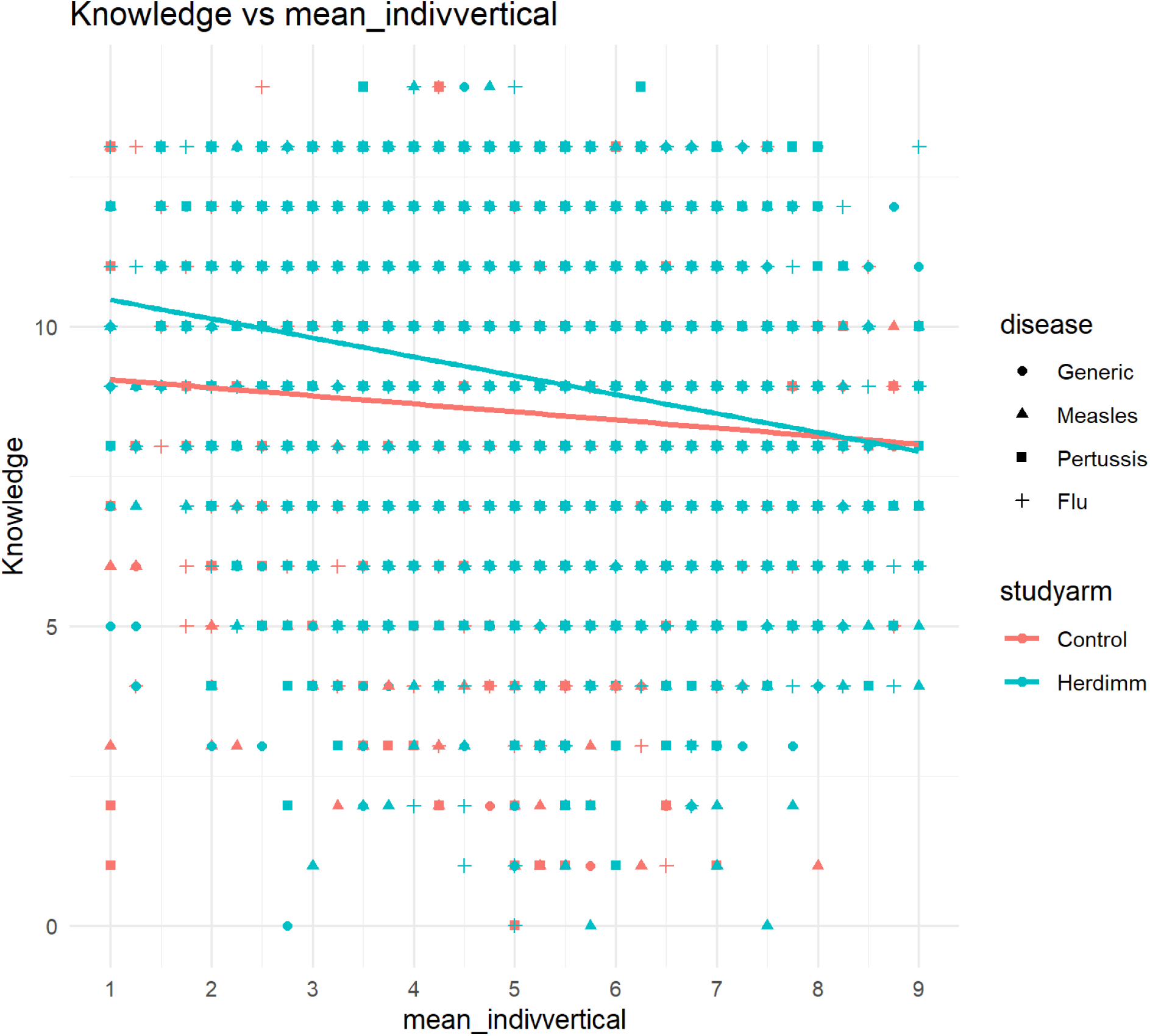
knowledge vs mean vertical individualism.

**S15:**
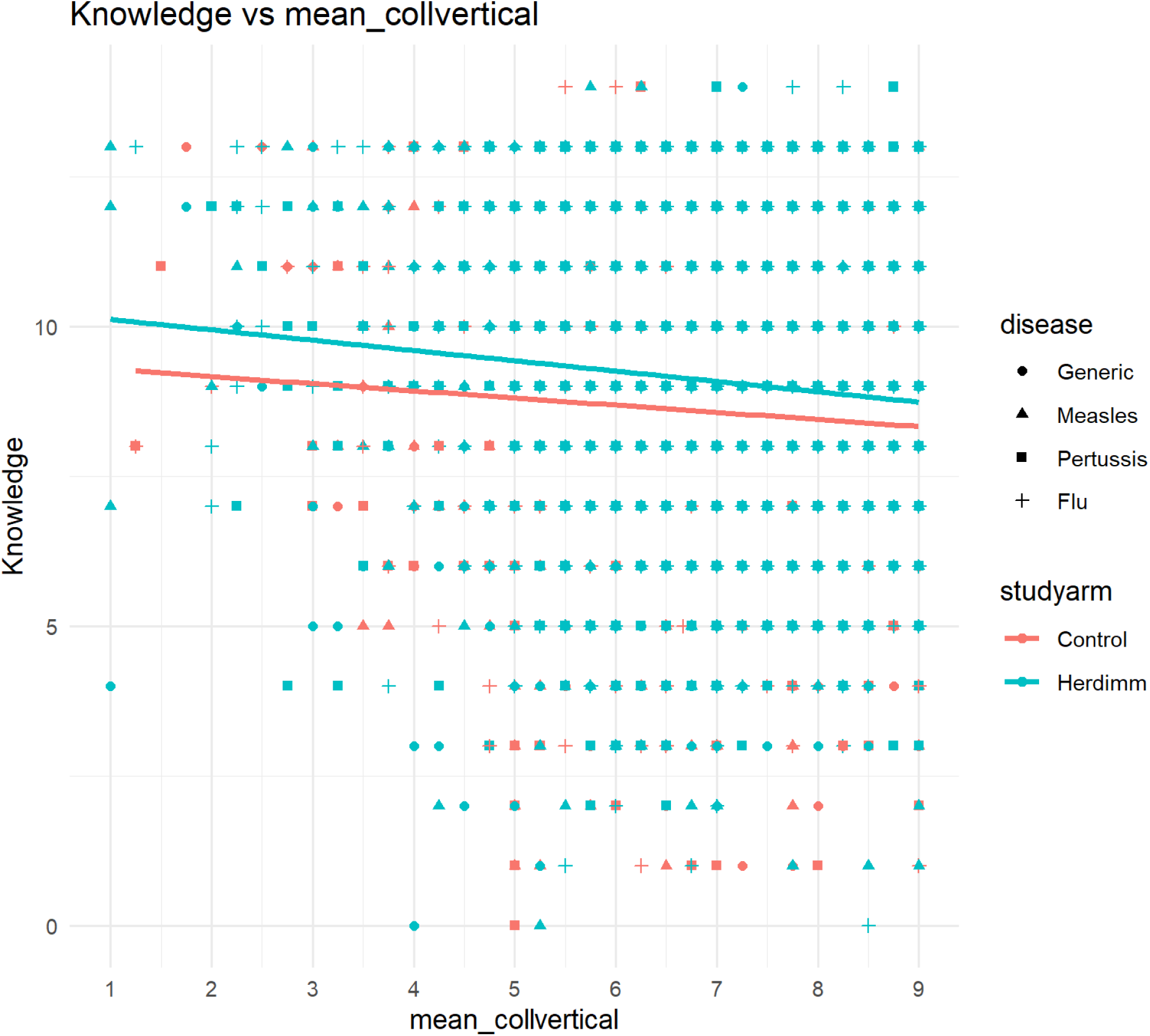
knowledge vs mean vertical collectivism.

**S16:**
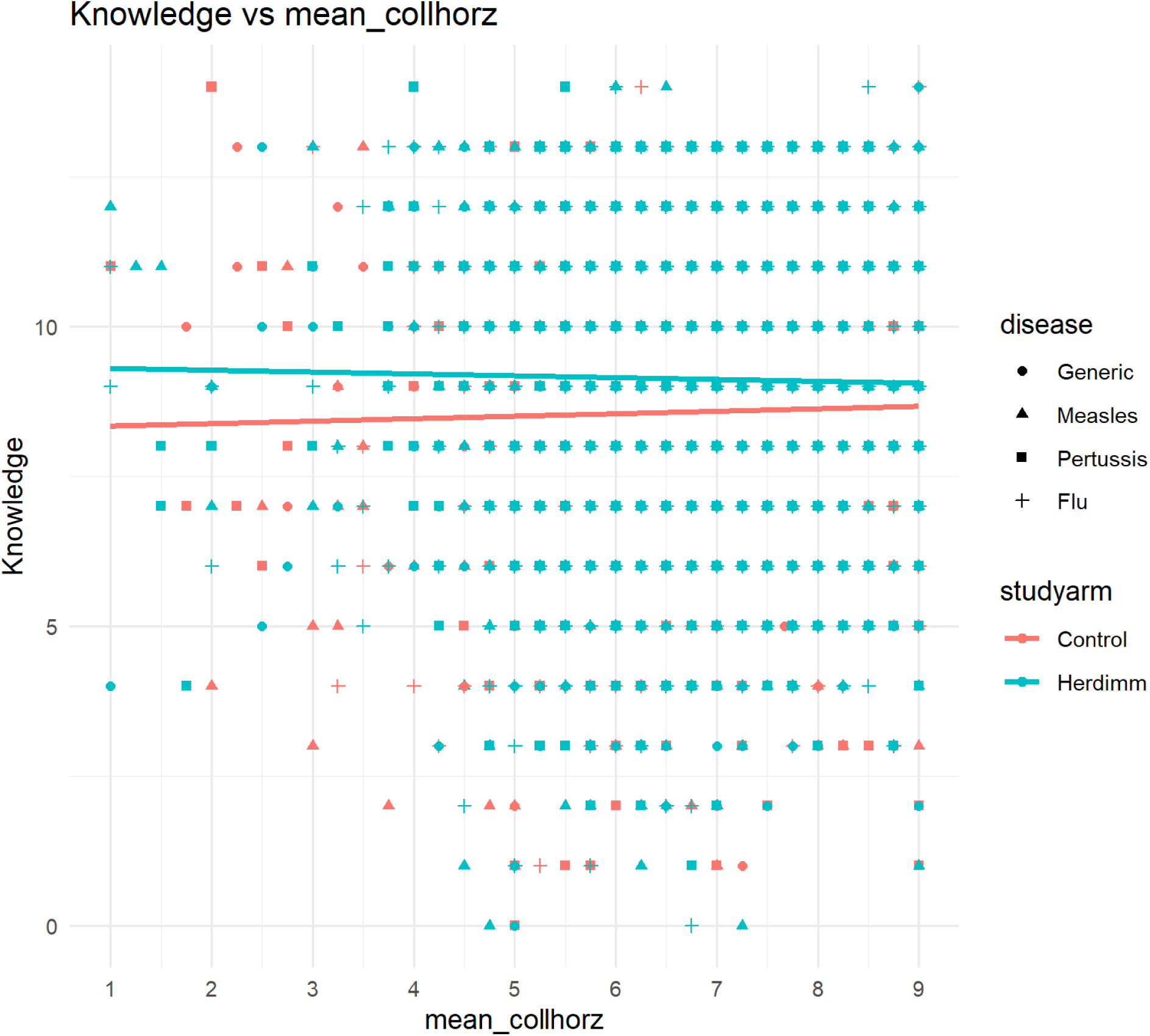
knowledge vs mean horizontal collectivism.

### Vaccine intentions

**S17:**
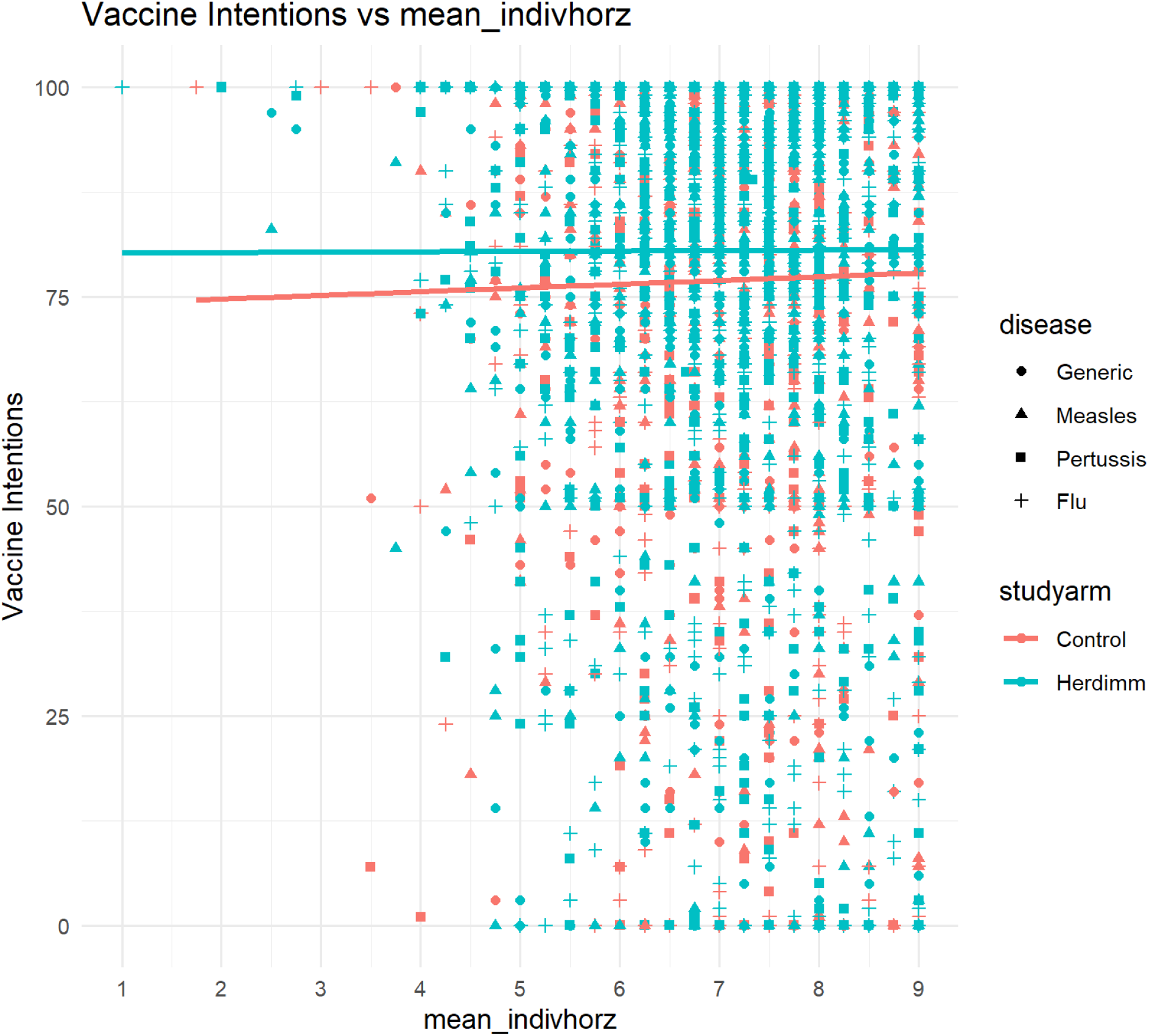
vaccine intentions vs mean horizontal individualism.

**S18:**
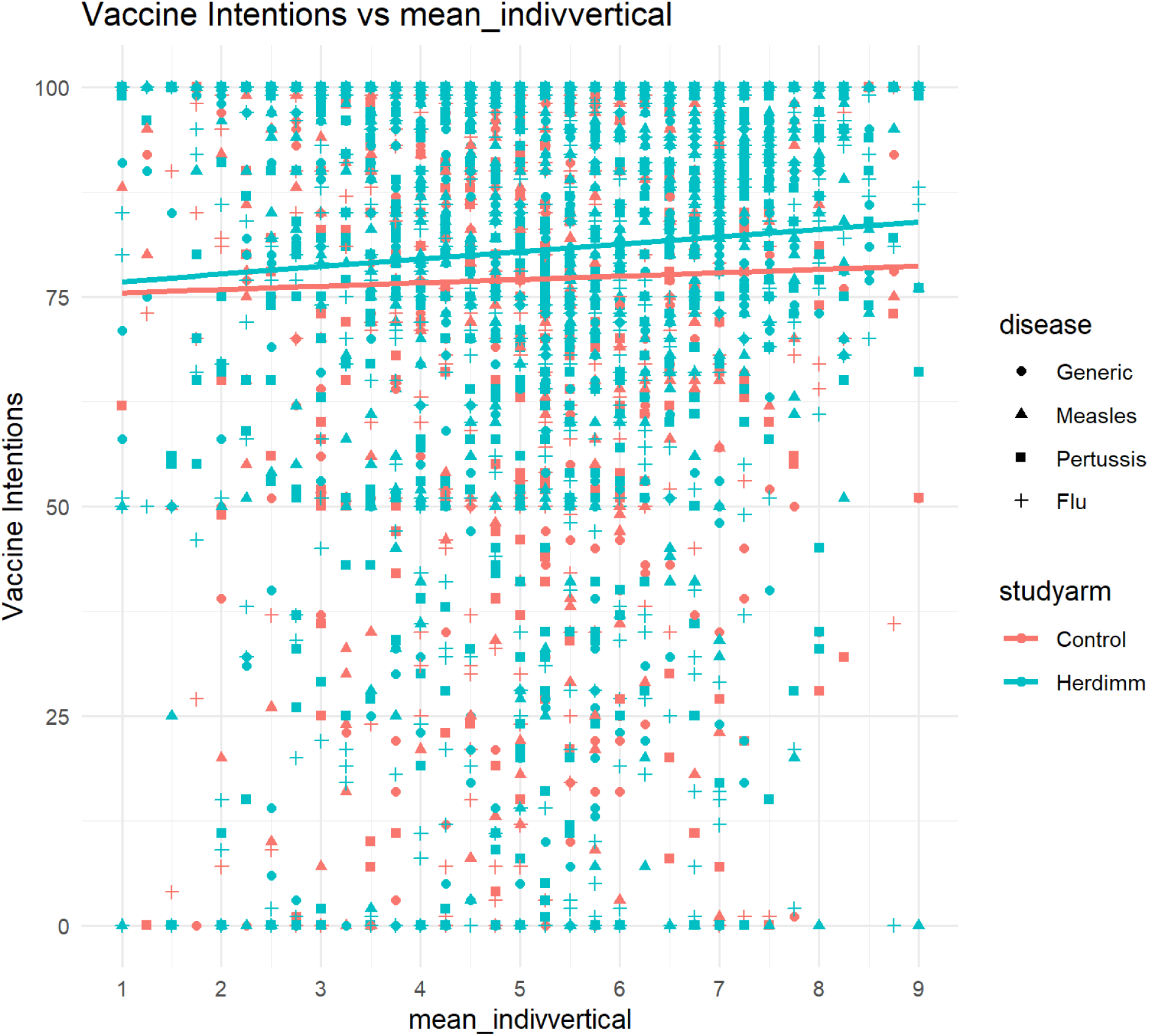
vaccine intentions vs mean vertical individualism.

**S19:**
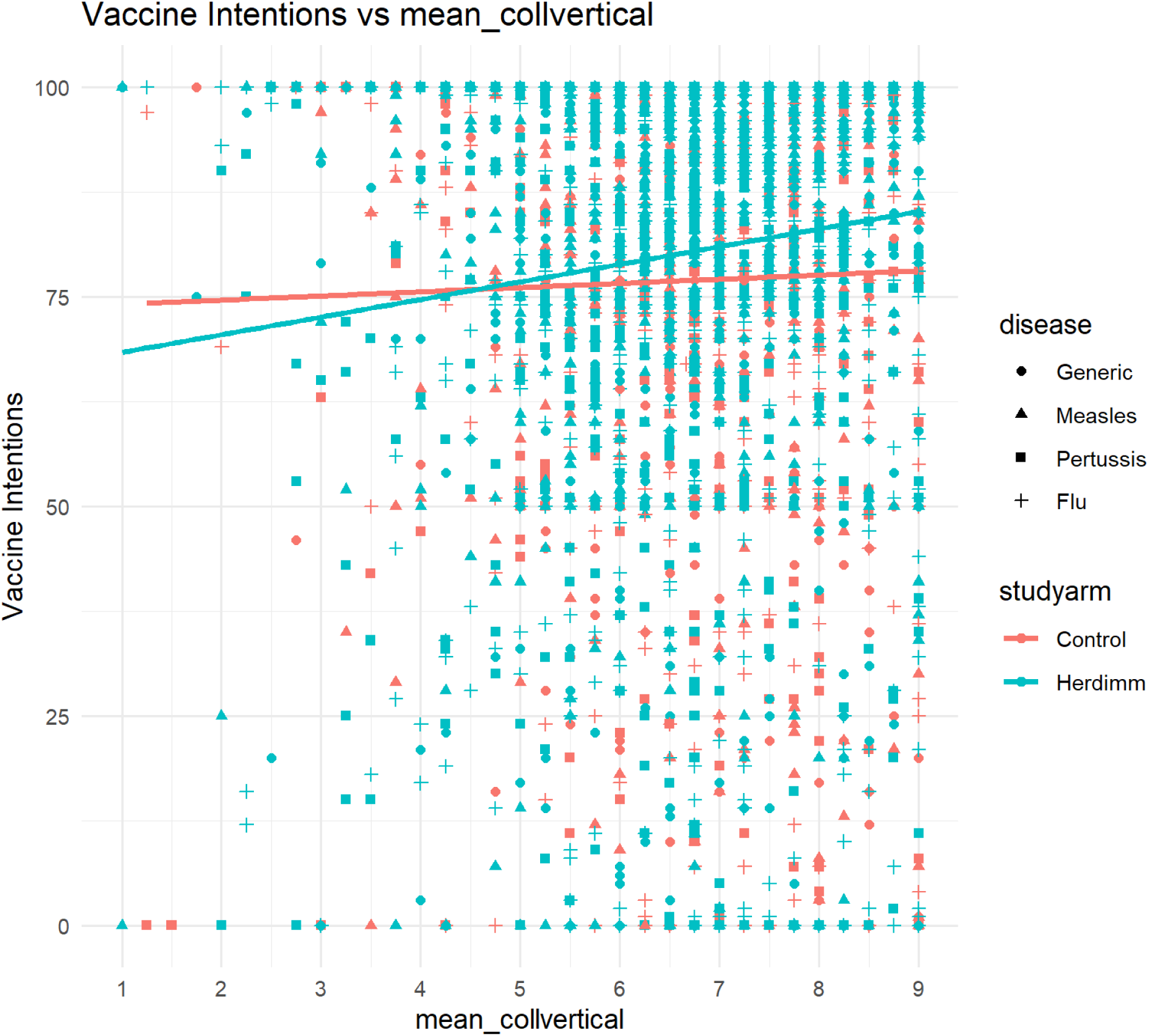
vaccine intentions vs mean vertical collectivism.

**S20:**
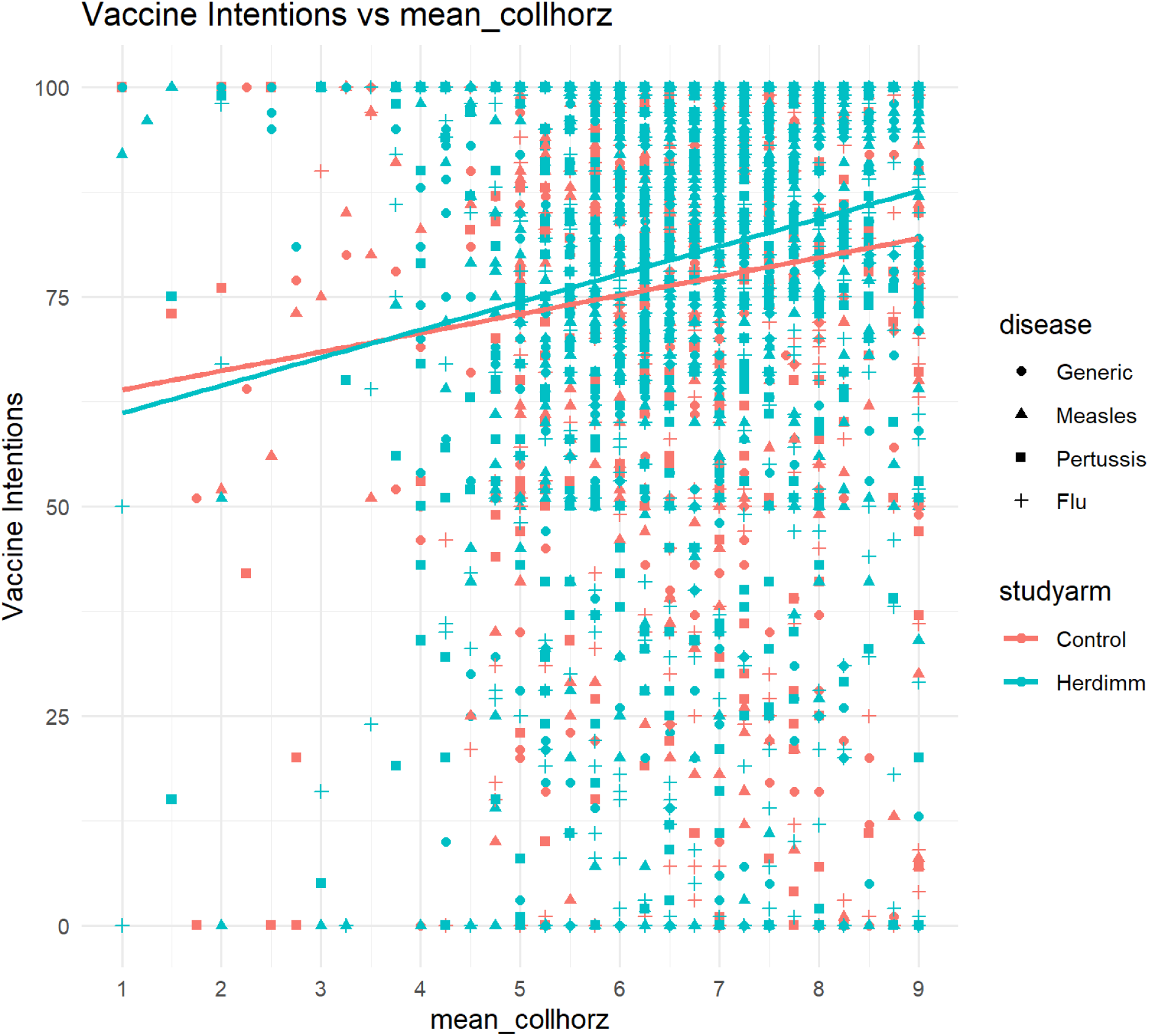
vaccine intentions vs mean horizontal collectivism.

### Timestamp for *herdimm* versus Individualism and Collectivism subscales (for participants assigned to *herdimm*)

Timestamp = seconds between reaching the page in which participants were directed to the *herdimm* intervention and returning to the page and clicking submit

**S21:**
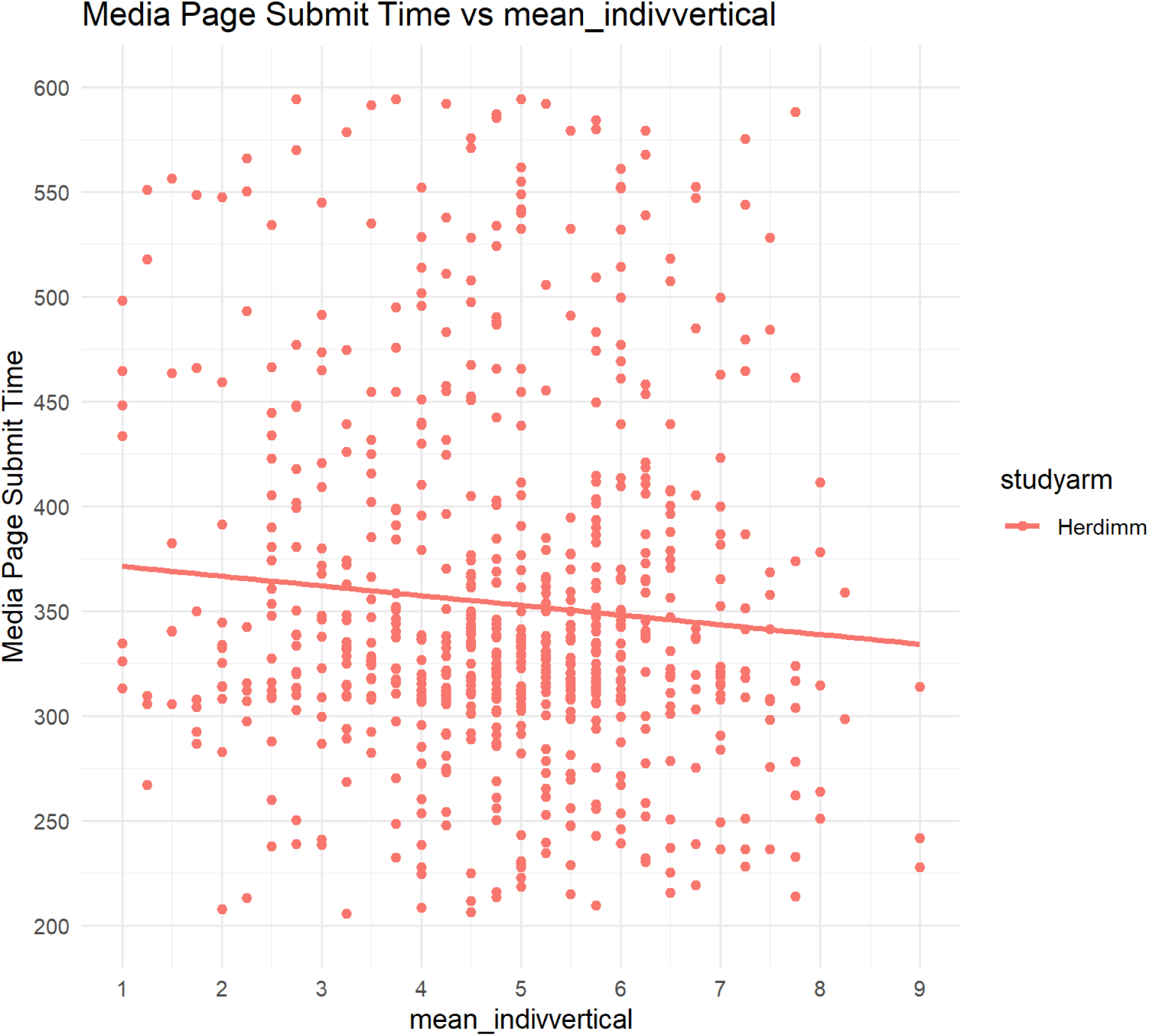
Timestamp submitted for *herdimm* versus mean vertical individualism.

**S22:**
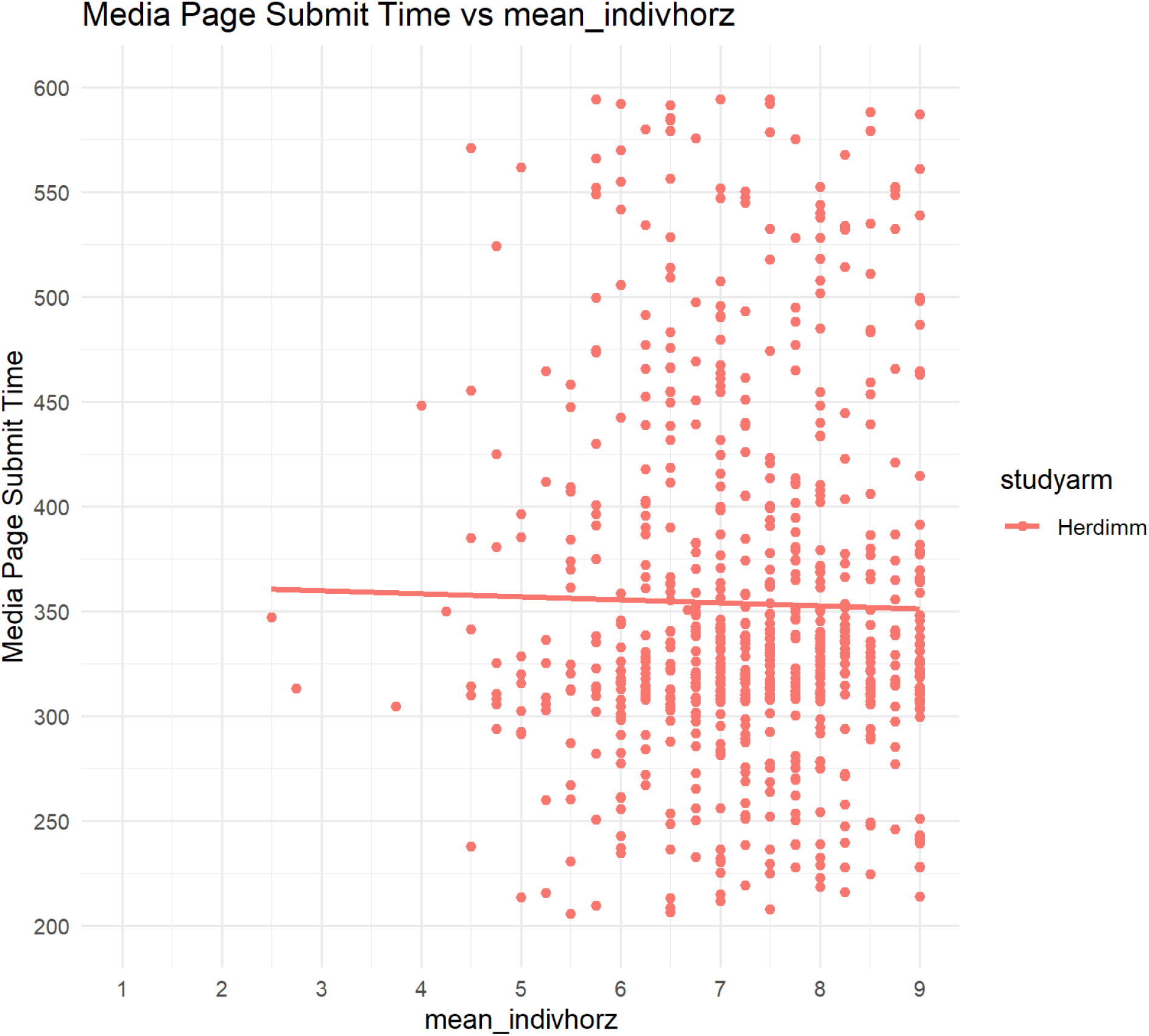
Timestamp submitted for *herdimm* versus mean horizontal individualism.

**S23:**
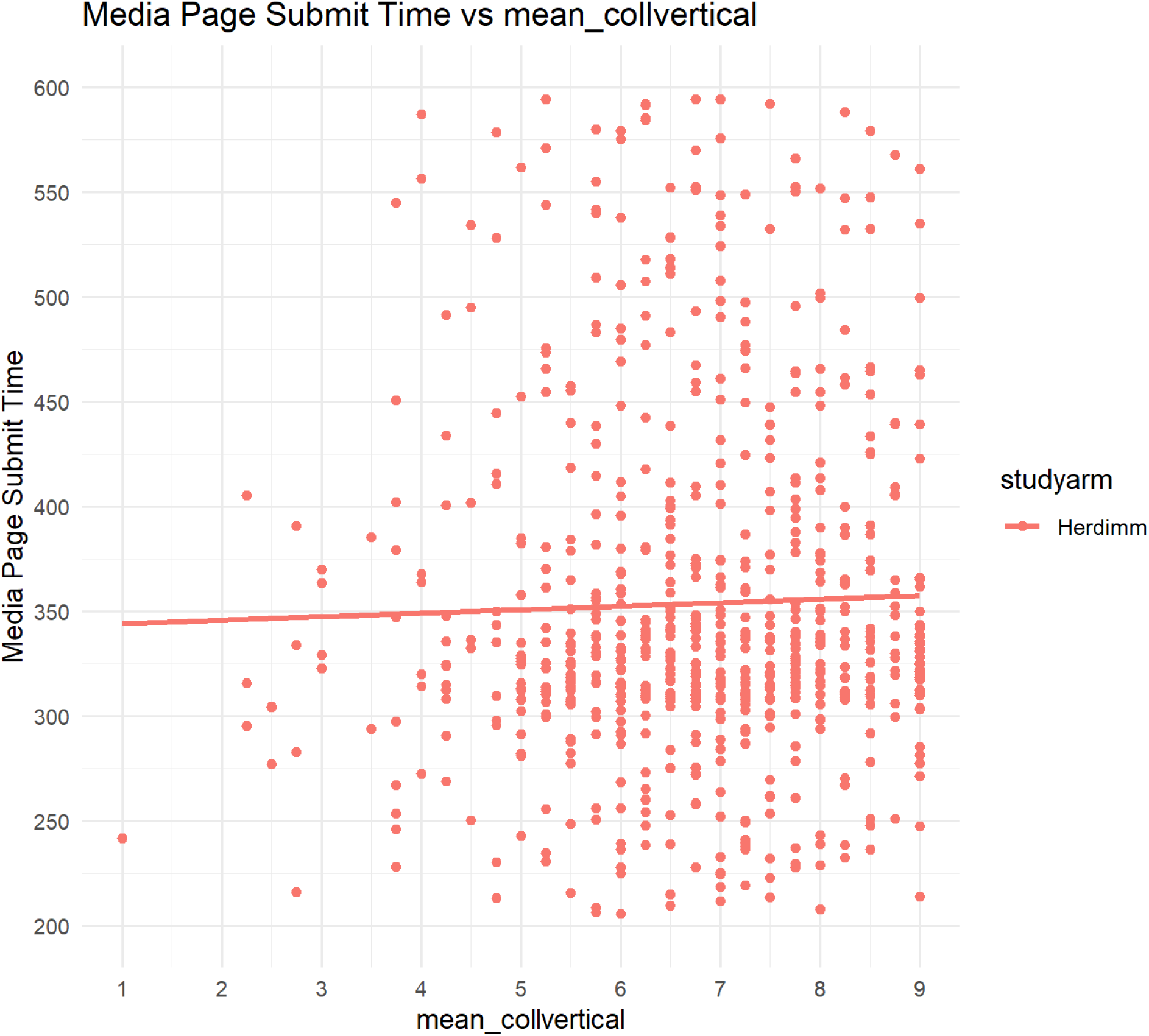
Timestamp submitted for *herdimm* versus mean vertical collectivism.

**S24:**
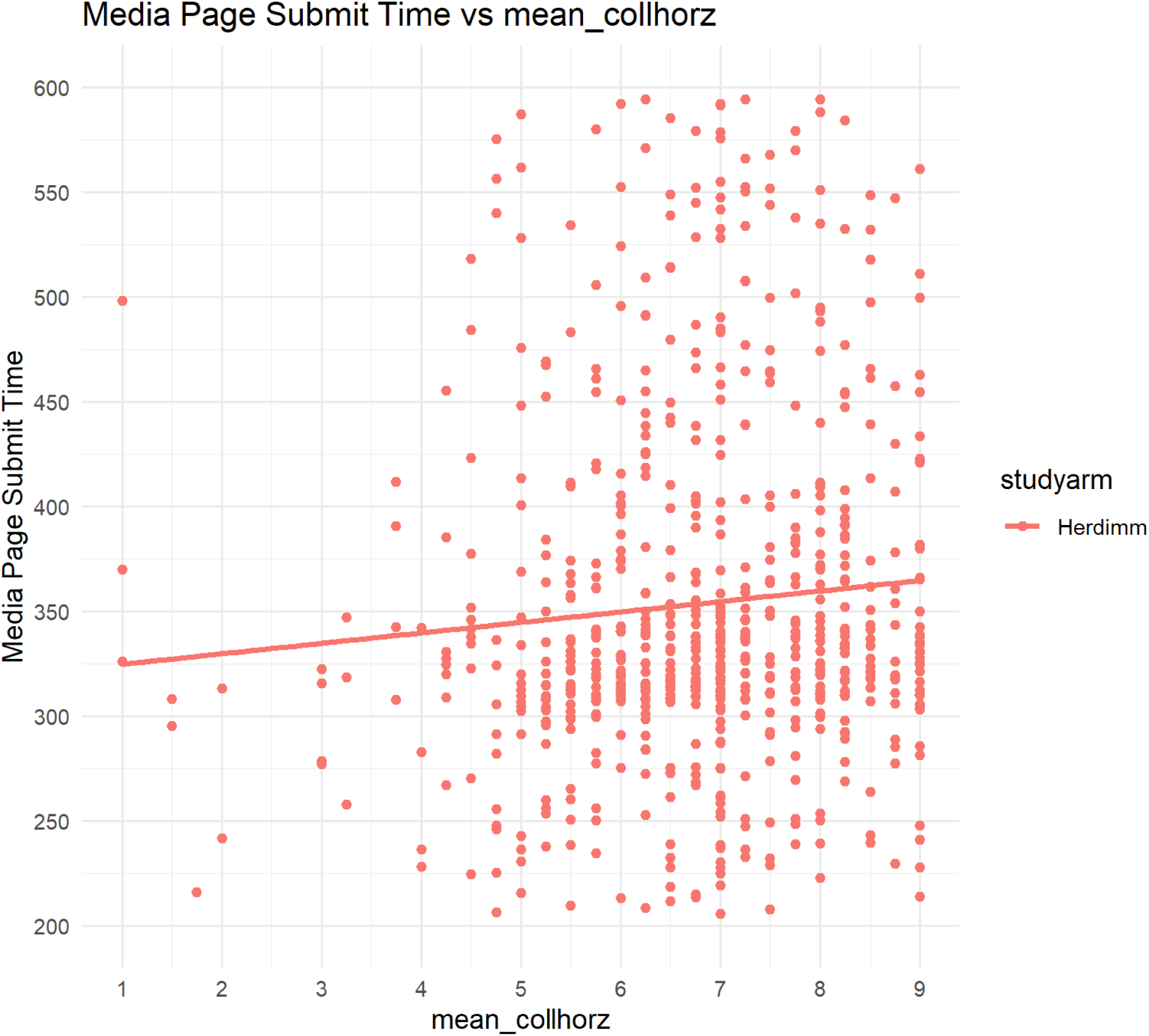
Timestamp submitted for *herdimm* versus mean horizontal collectivism.

## Notes

### Competing Interest Statement

The authors have declared no competing interest.

### Clinical Trial

NCT04787913

### Clinical Protocols

https://osf.io/hkysb/

### Author Declarations

This project is approved by the "Comité d'éthique de la recherche en sciences de la santé" ethics committee of Laval University (Approval No. 2017- 137 Phase II / 03-09-2019). All participants included in the study consented to participate in the study.

